# Mapping national and subnational zero-dose prevalence and multidimensional inequities across 77 countries

**DOI:** 10.64898/2026.09.03.26362103

**Authors:** Chuyao Huang, Ruixin Chi, Yanhong Gong, Chenyu Yan, Gatien De Broucker, Hai Fang, Xiaoxv Yin, Haijun Zhang, Bryan Patenaude

## Abstract

Zero-dose children remain a marker of populations unreached by routine immunisation, but national averages can obscure substantial subnational and multidimensional inequities. We analysed the most recent Demographic and Health Surveys and Multiple Indicator Cluster Surveys conducted between 2015 and 2024, including 170,553 children aged 12–23 months from 77 countries and 1,571 first-level administrative areas. Zero-dose prevalence ranged from 0.22% to 37.21% nationally, and 33 subnational areas had prevalence of at least 50%. Using the Vaccine Economics Research for Sustainability and Equity (VERSE) framework, we found that zero-dose children were consistently concentrated among multidimensionally disadvantaged groups, although the magnitude and drivers of inequity varied across countries. Maternal education, household wealth, geographic region, residence and health insurance were context-specific contributors. Combining burden, subnational distribution and multidimensional inequity can help identify where zero-dose children are concentrated and which barriers should be prioritised to advance equitable immunisation.

---

Vaccination has transformed child survival and remains one of the most effective public health interventions worldwide.^1^ Since the launch of the Expanded Programme on Immunization (EPI) in 1974, routine immunisation programmes have prevented substantial morbidity and mortality from infectious diseases and generated major social and economic returns.^2,3^ Vaccination is estimated to have averted 154 million deaths globally, including 146 million among children younger than 5 years, and to have accounted for approximately 40% of the observed decline in global infant mortality.^4^ In 2025, 90% of infants worldwide received at least one dose of a diphtheria – tetanus – pertussis (DTP)-containing vaccine and 85% completed the three-dose series, demonstrating the remarkable reach of routine immunisation while also highlighting persistent coverage gaps.^5^ The Global Vaccine Action Plan 2011–2020 set ambitious targets for national vaccination coverage and equitable access, and the Immunization Agenda 2030 further placed equity and “leaving no one behind” at the centre of global immunisation strategies.^5,6^ However, these achievements remain fragile. Expanding vaccine portfolios, constrained health-system resources and changing global immunisation financing have made it increasingly important not only to sustain high coverage, but also to identify the children and communities still missed by routine immunisation services.^7,8^

Zero-dose children represent one of the vital signals of this unfinished agenda. Commonly defined as children who have not received a DTP-containing vaccine, zero-dose status is not merely a delay in one vaccine dose, but a marker of being unreached by routine immunisation and often by primary health-care systems.^9^ Despite this high overall coverage, an estimated 13.5 million infants worldwide remained zero-dose in 2025, with most living in low- and middle-income countries.^5^ Previous studies suggest that zero-dose children are disproportionately concentrated among households facing multiple and overlapping disadvantages, including poverty, low maternal education, and rural residence.^10^ Evidence from India further shows that zero-dose status is a consistent marker of generalised vulnerability, with zero-dose children more likely to live in disadvantaged districts and to experience nutritional deficits.^11^ These findings indicate that zero-dose children should be understood not only as an immunisation coverage gap, but also as a sentinel population for broader social and health-system exclusion.

Despite growing attention to zero-dose children, important evidence gaps remain. Many existing analyses focus on national averages or examine single dimensions of inequality separately, such as wealth or education. Such approaches may obscure substantial within-country heterogeneity and the cumulative disadvantages that shape whether children are reached by routine immunisation.^12–14^ Subnational evidence is particularly important because children missed by vaccination programmes are often geographically clustered, and countries with similar national zero-dose prevalence may require different targeting strategies.^15–17^ Therefore, using the most recent nationally representative Demographic and Health Surveys (DHS) and Multiple Indicator Cluster Surveys (MICS) conducted since 2015, we estimated zero-dose prevalence among children aged 12-23 months across 77 countries, mapped subnational variation across 1,571 first-level administrative (ADM1) areas, and quantified multidimensional inequities and their contributing factors. By combining national estimates, subnational mapping and equity-oriented analysis, this study aims to provide actionable evidence for identifying which children are being left behind, where they are located and which social or geographic factors should be prioritised to accelerate progress towards Immunization Agenda 2030 (IA2030) globally.

## Results

### Study population and distribution of zero-dose vaccination status

The analytic sample included 170,553 children aged 12-23 months from 77 countries. Children were unevenly distributed across WHO regions, with the largest numbers from the African Region (66,745; 39.13%) and South-East Asia Region (53,673; 31.47%). Most children were from lower-middle-income countries (107,274; 62.90%) and lived in rural areas (115,972; 68.00%). In addition, 27.11% of children had mothers with no formal education (**Table 1**).

**Table 1.** Sociodemographic characteristics of children and zero-dose prevalence across 77 countries.

| **Variables** | **N (%)** | **Zero-dose prevalence (%)** |
| --- | --- | --- |
| **Region group by WHO** |  |  |
| African Region | 66745 (39.13) | 10.95 |
| Region of the Americas | 9691 (5.68) | 4.36 |
| Eastern Mediterranean Region | 27818 (16.31) | 2.84 |
| European Region | 3787 (2.22) | 8.20 |
| South-East Asia Region | 53673 (31.47) | 8.39 |
| Western Pacific Region | 8839 (5.18) | 9.88 |
| **World Bank income group** |  |  |
| High-income countries | 174 (0.10) | 3.12 |
| Upper-middle income countries | 43679 (25.61) | 9.85 |
| Lower-middle income countries | 107274 (62.90) | 10.44 |
| Low-income countries | 19426 (11.39) | 5.02 |
| **Sex** |  |  |
| Male | 87648 (51.39) | 8.69 |
| Female | 82905 (48.61) | 8.70 |
| **Maternal education** |  |  |
| No education | 46245 (27.11) | 16.18 |
| Primary | 43443 (25.47) | 8.39 |
| Secondary | 63210 (37.06) | 5.68 |
| Higher | 17614 (10.33) | 4.18 |
| **Wealth index** |  |  |
| Poorest | 43081 (25.26) | 12.36 |
| Poorer | 37491 (21.98) | 9.98 |
| Middle | 34134 (20.01) | 8.40 |
| Richer | 30611 (17.95) | 6.07 |
| Richest | 25226 (14.79) | 5.64 |
| **Place of residence** |  |  |
| Urban | 54378 (31.88) | 6.26 |
| Rural | 115972 (68.00) | 10.44 |
| **Health insurance** |  |  |
| With | 30049 (17.62) | 3.88 |
| Without | 121987 (71.52) | 10.52 |

Note: Due to varying amounts of missing data across variables, the sample size differs by variable.

Zero-dose prevalence varied substantially across geographic and socioeconomic groups. Across WHO regions, the highest prevalence was observed in the African Region (10.95%), followed by the Western Pacific Region (9.88%), whereas the lowest prevalence was observed in the Eastern Mediterranean Region (2.84%). By World Bank income group, zero-dose prevalence was highest in lower-middle-income countries (10.44%) and upper-middle-income countries (9.85%), and lowest in high-income countries (3.12%).

Differences in zero-dose prevalence were also observed across maternal education, household wealth, place of residence, and health insurance coverage. Prevalence was highest among children whose mothers had no formal education (16.18%) and lowest among those whose mothers had higher education (4.18%). Similarly, children from the poorest household wealth quintile had higher zero-dose prevalence than those from the richest quintile (12.36% vs 5.64%). Rural children had higher prevalence than urban children (10.44% vs 6.26%), and children without health insurance coverage had higher prevalence than those with insurance coverage (10.52% vs 3.88%). Zero-dose prevalence was similar between boys and girls (8.69% vs 8.70%).

### Zero-dose prevalence varies substantially across and within countries

At the national level, zero-dose prevalence showed substantial heterogeneity across the 77 included countries (**Supplementary Table 4**). National prevalence exceeded 10.0% in 22 countries and exceeded 30.0% in six countries, including Guinea (37.21%), Nigeria (36.79%), Azerbaijan (36.23%), Papua New Guinea (34.42%), Democratic Republic of the Congo (33.10%) and Angola (30.10%). In contrast, the lowest national prevalence estimates were observed in Zimbabwe (0.22%), São Tomé and Príncipe (0.25%), and Tunisia (0.36%).

Subnational analyses revealed considerable within-country variation that was not captured by national averages (**Fig. 1, Supplementary Table 5**). Across the 1,571 mapped ADM1 areas, zero-dose prevalence ranged from areas with no observed zero-dose children to areas with extremely high prevalence. Although approximately three-quarters of ADM1 areas had prevalence below 10.0%, 105 areas (7.0%) had prevalence of at least 30.0%, including 33 areas (2.2%) with prevalence of 50.0% or higher. High-prevalence areas were geographically concentrated in several countries. Zero-dose prevalence reached at least 50.0% in 9 of 26 mapped ADM1 areas in the Democratic Republic of the Congo (34.6%), 7 of 37 in Nigeria (18.9%), 5 of 81 in the Philippines (6.2%), and 4 of 18 in Angola (22.2%). Additional high-prevalence ADM1 areas were identified in Ethiopia (2/11), Mali (2/9), Guinea (1/8), and Cambodia (1/25).

**Figure 1.**
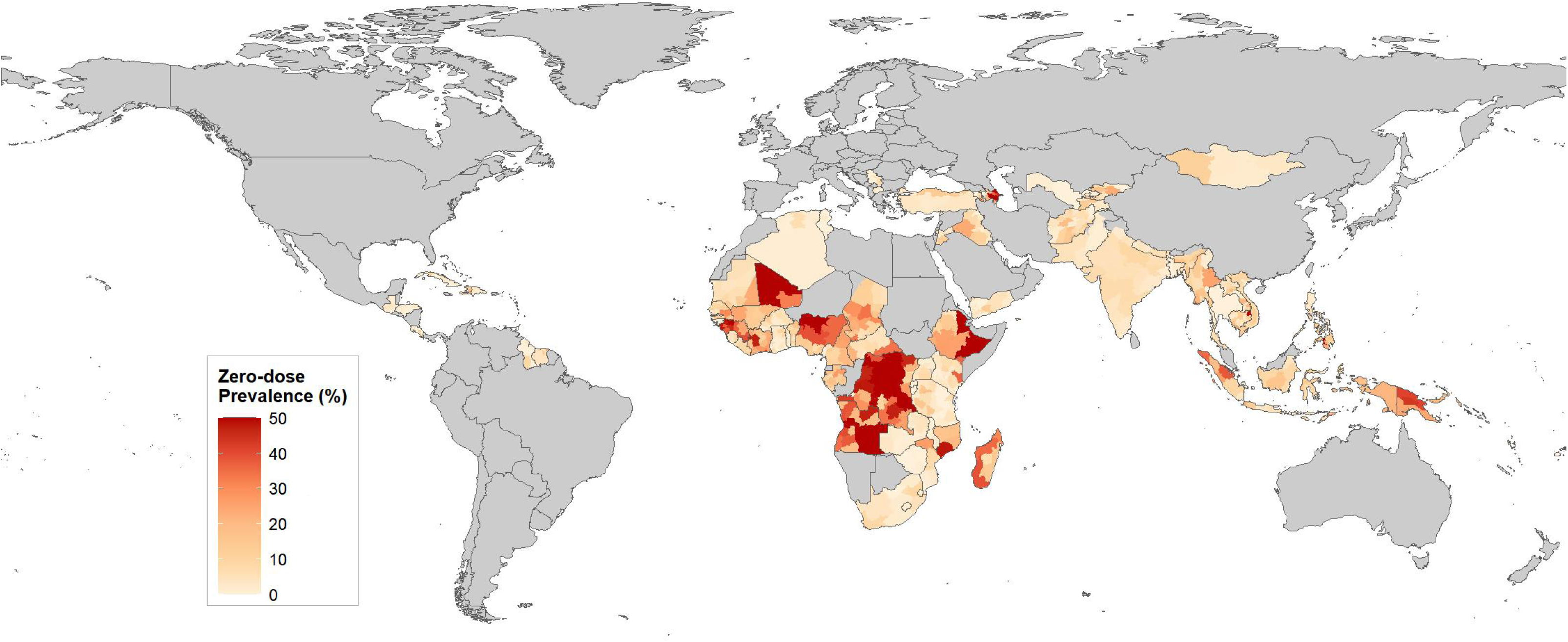
Subnational distribution of zero-dose prevalence across 77 countries, 2015 - 2024

### Zero-dose children are concentrated among multidimensionally disadvantaged populations

At the national level, zero-dose children were consistently concentrated among multidimensionally disadvantaged populations, with all included countries showing positive concentration indices (**Table 2**). The magnitude of relative inequity varied substantially across countries. The lowest CI values were observed in Sierra Leone (0.215), Guinea (0.254), Côte d’Ivoire (0.260), and Chad (0.266), whereas the highest values were observed in São Tomé and Príncipe (0.997), Tunisia (0.996), and Zimbabwe (0.979). Absolute equity gaps (AEGs) showed a different pattern, reflecting the magnitude of prevalence differences between the most and least disadvantaged groups. The largest AEGs were observed in Azerbaijan (0.878), Democratic Republic of the Congo (0.651), and Nigeria (0.649), indicating substantial absolute differences in zero-dose prevalence across disadvantage gradients. Conversely, countries with very high CI values but low zero-dose prevalence had minimal AEGs, including São Tomé and Príncipe (0.002), Tunisia (0.004), and Zimbabwe (0.010).

**Table 2.** Measures of multidimensional inequity in zero-dose prevalence across 77 countries, 2015 - 2024.

| Country | Year | Absolute Equity Gap (AEG) | Concentration Index (CI) |
| --- | --- | --- | --- |
| **African Region** |  |  |  |
| Angola | 2015 | 0.640 (0.597, 0.683) | 0.378 (0.353, 0.404) |
| Benin | 2021 | 0.184 (0.147, 0.221) | 0.467 (0.409, 0.524) |
| Burkina Faso | 2021 | 0.076 (0.047, 0.105) | 0.368 (0.257, 0.479) |
| Burundi | 2016 | 0.026 (0.012, 0.040) | 0.705 (0.567, 0.842) |
| Cameroon | 2018 | 0.271 (0.220, 0.322) | 0.348 (0.295, 0.400) |
| Central African Republic | 2019 | 0.246 (0.173, 0.319) | 0.456 (0.341, 0.572) |
| Chad | 2019 | 0.282 (0.215, 0.349) | 0.266 (0.202, 0.330) |
| Comoros | 2022 | 0.162 (0.091, 0.233) | 0.337 (0.192, 0.482) |
| Côte d'Ivoire | 2021 | 0.354 (0.295, 0.413) | 0.260 (0.219, 0.301) |
| Democratic Republic of the Congo | 2023 | 0.651 (0.616, 0.686) | 0.332 (0.314, 0.350) |
| Eswatini | 2021 | 0.092 (0.027, 0.157) | 0.583 (0.332, 0.834) |
| Ethiopia | 2018 | 0.503 (0.429, 0.577) | 0.458 (0.412, 0.503) |
| Gabon | 2021 | 0.247 (0.186, 0.308) | 0.326 (0.263, 0.389) |
| The Gambia | 2020 | 0.050 (0.026, 0.074) | 0.507 (0.348, 0.665) |
| Ghana | 2022 | 0.106 (0.077, 0.135) | 0.625 (0.532, 0.718) |
| Guinea | 2018 | 0.432 (0.359, 0.505) | 0.254 (0.219, 0.289) |
| Guinea-Bissau | 2019 | 0.041 (0.017, 0.065) | 0.603 (0.407, 0.800) |
| Kenya | 2022 | 0.147 (0.122, 0.172) | 0.715 (0.669, 0.761) |
| Lesotho | 2024 | 0.044 (0.005, 0.083) | 0.822 (0.712, 0.931) |
| Liberia | 2019 | 0.141 (0.092, 0.190) | 0.320 (0.224, 0.416) |
| Madagascar | 2021 | 0.404 (0.355, 0.453) | 0.405 (0.369, 0.440) |
| Malawi | 2020 | 0.029 (0.015, 0.043) | 0.394 (0.250, 0.539) |
| Mali | 2024 | 0.360 (0.317, 0.403) | 0.427 (0.395, 0.460) |
| Mauritania | 2020 | 0.221 (0.178, 0.264) | 0.368 (0.299, 0.437) |
| Mozambique | 2022 | 0.445 (0.392, 0.498) | 0.532 (0.488, 0.576) |
| Nigeria | 2024 | 0.649 (0.618, 0.680) | 0.398 (0.380, 0.416) |
| Rwanda | 2020 | 0.014 (0.000, 0.028) | 0.762 (0.531, 0.994) |
| São Tomé and Príncipe | 2019 | 0.002 (-0.004, 0.008) | 0.997 (0.991, 1.003) |
| Senegal | 2023 | 0.206 (0.163, 0.249) | 0.382 (0.314, 0.449) |
| Sierra Leone | 2019 | 0.068 (0.037, 0.099) | 0.215 (0.102, 0.329) |
| South Africa | 2016 | 0.138 (0.079, 0.197) | 0.511 (0.376, 0.645) |
| Tanzania | 2022 | 0.140 (0.107, 0.173) | 0.579 (0.507, 0.652) |
| Togo | 2017 | 0.065 (0.030, 0.100) | 0.599 (0.439, 0.760) |
| Uganda | 2016 | 0.080 (0.053, 0.107) | 0.392 (0.301, 0.483) |
| Zambia | 2018 | 0.050 (0.026, 0.074) | 0.453 (0.277, 0.629) |
| Zimbabwe | 2019 | 0.010 (-0.004, 0.024) | 0.979 (0.963, 0.994) |
| **Region of the Americas** |  |  |  |
| Costa Rica | 2018 | 0.124 (0.069, 0.179) | 0.740 (0.619, 0.862) |
| Cuba | 2019 | 0.089 (0.052, 0.126) | 0.743 (0.539, 0.948) |
| Dominican Republic | 2019 | 0.051 (0.026, 0.076) | 0.397 (0.257, 0.536) |
| Guatemala | 2015 | 0.048 (0.028, 0.068) | 0.311 (0.186, 0.435) |
| Guyana | 2019 | 0.046 (0.005, 0.087) | 0.794 (0.688, 0.901) |
| Haiti | 2017 | 0.286 (0.221, 0.351) | 0.386 (0.322, 0.450) |
| Honduras | 2019 | 0.032 (0.012, 0.052) | 0.813 (0.724, 0.902) |
| Suriname | 2018 | 0.165 (0.096, 0.234) | 0.385 (0.266, 0.504) |
| Trinidad and Tobago | 2022 | 0.092 (-0.002, 0.186) | 0.403 (0.042, 0.764) |
| **Eastern Mediterranean Region** |  |  |  |
| Afghanistan | 2023 | 0.107 (0.082, 0.132) | 0.408 (0.350, 0.465) |
| Algeria | 2019 | 0.053 (0.031, 0.075) | 0.543 (0.418, 0.669) |
| Iraq | 2018 | 0.141 (0.110, 0.172) | 0.440 (0.376, 0.504) |
| Jordan | 2023 | 0.114 (0.079, 0.149) | 0.476 (0.342, 0.609) |
| Pakistan | 2019 | 0.044 (0.036, 0.052) | 0.423 (0.367, 0.480) |
| Palestine | 2019 | 0.156 (0.107, 0.205) | 0.799 (0.716, 0.882) |
| Tunisia | 2023 | 0.004 (-0.004, 0.012) | 0.996 (0.989, 1.004) |
| Yemen | 2023 | 0.092 (0.065, 0.119) | 0.527 (0.419, 0.636) |
| **European Region** |  |  |  |
| Armenia | 2016 | 0.105 (0.032, 0.178) | 0.811 (0.707, 0.915) |
| Azerbaijan | 2023 | 0.878 (0.694, 1.062) | 0.456 (0.285, 0.628) |
| Kyrgyzstan | 2023 | 0.174 (0.094, 0.254) | 0.542 (0.440, 0.643) |
| North Macedonia | 2018 | 0.157 (0.063, 0.251) | 0.714 (0.547, 0.881) |
| Serbia | 2019 | 0.095 (0.024, 0.166) | 0.808 (0.653, 0.963) |
| Tajikistan | 2023 | 0.147 (0.090, 0.204) | 0.292 (0.197, 0.387) |
| Turkey | 2018 | 0.110 (0.045, 0.175) | 0.438 (0.227, 0.650) |
| Uzbekistan | 2021 | 0.032 (0.008, 0.056) | 0.898 (0.831, 0.966) |
| **South-East Asia Region** |  |  |  |
| Bangladesh | 2017 | 0.048 (0.024, 0.072) | 0.616 (0.486, 0.746) |
| India | 2019 | 0.101 (0.093, 0.109) | 0.298 (0.278, 0.317) |
| Indonesia | 2017 | 0.273 (0.238, 0.308) | 0.454 (0.410, 0.498) |
| Myanmar | 2016 | 0.290 (0.225, 0.355) | 0.531 (0.446, 0.615) |
| Nepal | 2021 | 0.099 (0.056, 0.142) | 0.429 (0.292, 0.565) |
| Thailand | 2022 | 0.095 (0.066, 0.124) | 0.796 (0.730, 0.862) |
| Timor-Leste | 2016 | 0.364 (0.299, 0.429) | 0.279 (0.225, 0.332) |
| **Western Pacific Region** |  |  |  |
| Cambodia | 2021 | 0.230 (0.183, 0.277) | 0.495 (0.429, 0.562) |
| Fiji | 2021 | 0.036 (-0.005, 0.077) | 0.975 (0.952, 0.997) |
| Laos | 2023 | 0.198 (0.149, 0.247) | 0.493 (0.409, 0.576) |
| Mongolia | 2018 | 0.103 (0.052, 0.154) | 0.436 (0.285, 0.587) |
| Papua New Guinea | 2016 | 0.479 (0.420, 0.538) | 0.298 (0.260, 0.337) |
| Philippines | 2022 | 0.401 (0.342, 0.460) | 0.530 (0.477, 0.584) |
| Tonga | 2019 | 0.091 (-0.011, 0.193) | 0.974 (0.943, 1.005) |
| Vanuatu | 2023 | 0.314 (0.194, 0.434) | 0.525 (0.408, 0.643) |
| Vietnam | 2020 | 0.205 (0.132, 0.278) | 0.286 (0.145, 0.427) |

At the subnational level, multidimensional inequity showed greater heterogeneity than national estimates (**Fig. 2, Supplementary Table 5**). Several ADM1 areas exhibited very high positive CI values, including Upper East in Ghana (0.992), Mashonaland West in Zimbabwe (0.992), and Guantánamo in Cuba (0.989). Notably, all subnational areas with CI values of 0.90 or higher had zero-dose prevalence below 5.0%, suggesting that strong relative concentration could occur even in settings with low overall burden.

**Figure 2.**
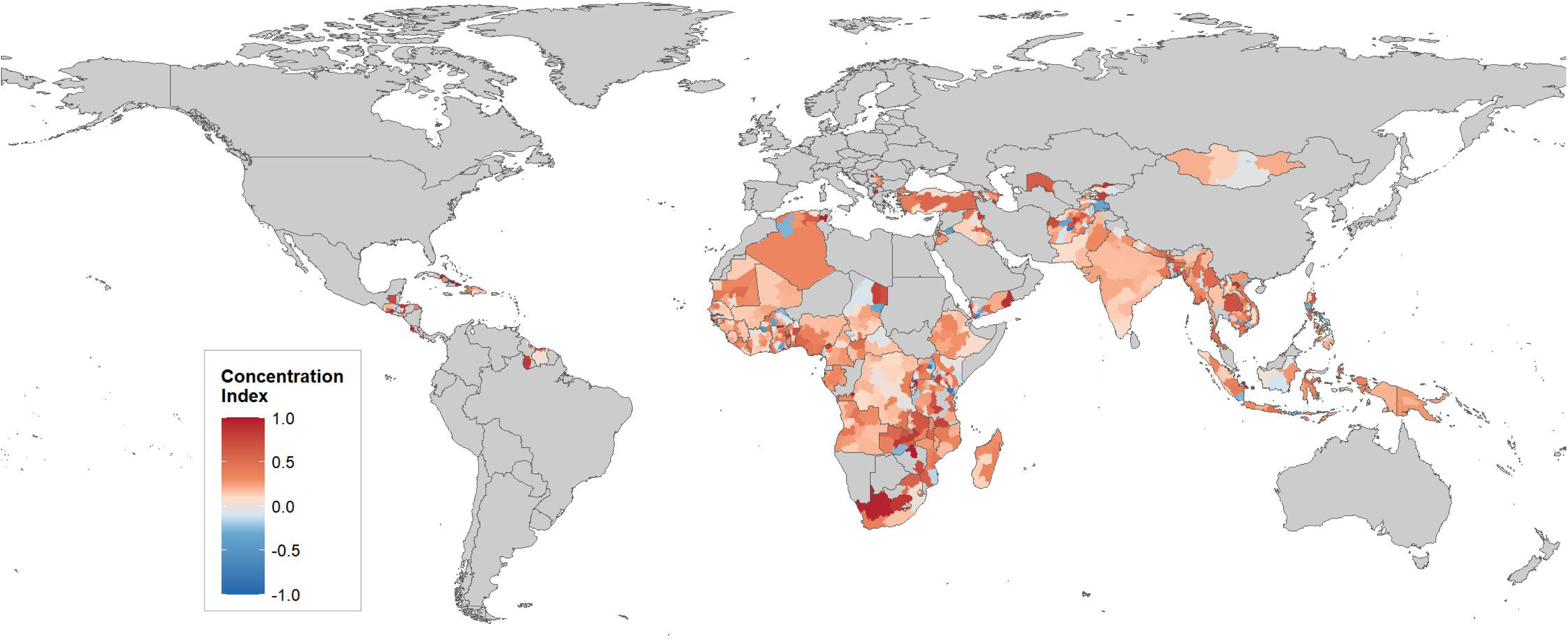
Subnational distribution of the concentration index for zero-dose prevalence across 77 countries, 2015 - 2024

Conversely, negative CI values were observed in some ADM1 areas, including Las Tunas in Cuba, several regions in Uganda, and Taita Taveta in Kenya, indicating that local patterns of disadvantage did not always align with national distributions. Subnational AEGs also varied substantially, with the largest gaps observed in Benguela, Angola (0.738), Gambela Peoples, Ethiopia (0.701), and Lunda Norte, Angola (0.670).

### Countries exhibit distinct profiles of zero-dose burden and inequity

The national equity–prevalence plane demonstrated that zero-dose burden and multidimensional inequity represented related but distinct dimensions of immunisation disadvantage (**Fig. 3**). Countries with high zero-dose prevalence were concentrated mainly in the African Region, with additional high-burden settings in the Western Pacific and European Regions. However, the distribution of countries across the equity–prevalence plane indicated that high burden did not necessarily correspond to greater multidimensional inequity.

**Figure 3.**
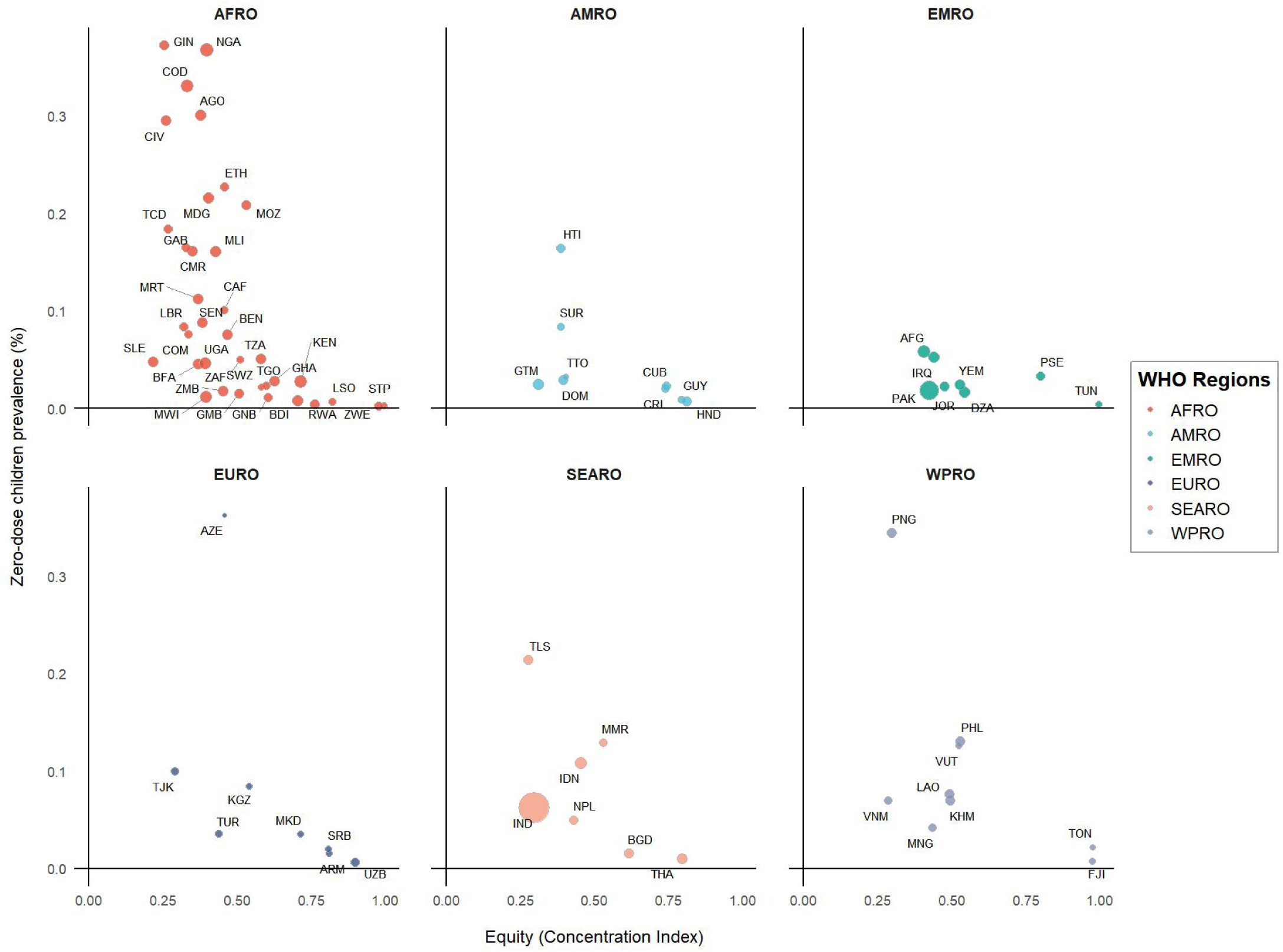
National equity–prevalence plane of zero-dose children by WHO region across 77 countries

Several countries experienced a dual challenge of high zero-dose prevalence and substantial concentration of zero-dose children among disadvantaged populations. These included Mozambique (20.82%; CI = 0.532), Ethiopia (22.70%; CI = 0.458), and Azerbaijan (36.23%; CI = 0.456). In contrast, some countries had very high zero-dose prevalence but comparatively lower CI values, indicating a more widespread distribution of zero-dose burden across disadvantage groups. For example, Guinea had the highest national zero-dose prevalence (37.21%) but a relatively low CI (0.254), while Papua New Guinea combined high prevalence (34.42%) with a relatively low CI (0.298). Conversely, several countries had low overall zero-dose prevalence but high multidimensional inequity, including São Tomé and Príncipe, Tunisia, Zimbabwe, Fiji, Tonga and Uzbekistan, highlighting settings where remaining zero-dose children were disproportionately concentrated among disadvantaged populations.

### Drivers of zero-dose inequity differ across countries

The decomposition analysis showed that the contributors of zero-dose inequity varied substantially across countries (**Table 3**). Among the measured determinants, maternal education and household wealth were the most frequent dominant contributors, although the relative importance of each dimension differed across settings. Maternal education contributed substantially to zero-dose inequity in Pakistan (71.01%), Nigeria (47.45%), and Jordan (46.27%), whereas household wealth was the dominant contributor in several high-burden countries, including Angola (62.50%), Papua New Guinea (49.45%), and Timor-Leste (48.79%).

**Table 3.** Decomposition of the concentration index for zero-dose prevalence by equity dimension across 77 countries.

| Country | Region | Place of residence | Maternal education | Wealth index | Sex | Health insurance | Residual |
| --- | --- | --- | --- | --- | --- | --- | --- |
| **African Region** |  |  |  |  |  |  |  |
| Angola | 1.04 (0.67, 1.42) | 10.55 (9.42, 11.69) | 24.05 (22.47, 25.63) | 62.50 (60.71, 64.29) | 0.06 (-0.03, 0.15) | 0.00 (-0.02, 0.03) | 1.78 (1.29, 2.27) |
| Benin | 4.59 (3.70, 5.47) | 3.38 (2.62, 4.15) | 38.75 (36.70, 40.81) | 14.88 (13.38, 16.38) | 6.57 (5.52, 7.61) | 1.35 (0.87, 1.84) | 30.48 (28.53, 32.42) |
| Burkina Faso | 2.01 (1.44, 2.59) | 4.90 (4.02, 5.79) | 2.96 (2.27, 3.65) | 19.57 (17.94, 21.19) | 1.39 (0.91, 1.87) | 0.17 (0.00, 0.34) | 69.00 (67.11, 70.89) |
| Burundi | 1.56 (1.08, 2.03) | 0.23 (0.05, 0.42) | 9.53 (8.40, 10.66) | 13.22 (11.91, 14.52) | 2.37 (1.79, 2.96) | 10.84 (9.64, 12.03) | 62.26 (60.39, 64.12) |
| Cameroon | 1.56 (0.99, 2.13) | 17.57 (15.82, 19.33) | 43.64 (41.35, 45.92) | 32.89 (30.73, 35.06) | 0.15 (-0.03, 0.32) | 2.64 (1.90, 3.38) | 1.56 (0.99, 2.13) |
| Central African Republic | 0.77 (0.13, 1.40) | 0.44 (-0.05, 0.92) | 2.05 (1.01, 3.08) | 3.93 (2.51, 5.35) | 0.04 (-0.10, 0.18) | 42.48 (38.87, 46.10) | 50.30 (46.65, 53.96) |
| Chad | 1.90 (1.15, 2.66) | 1.24 (0.63, 1.85) | 2.41 (1.56, 3.26) | 0.53 (0.13, 0.93) | 0.03 (-0.07, 0.13) | 0.25 (-0.02, 0.53) | 93.63 (92.28, 94.98) |
| Comoros | 14.87 (12.17, 17.57) | 2.66 (1.44, 3.88) | 3.22 (1.88, 4.56) | 20.58 (17.51, 23.65) | 55.23 (51.46, 59.00) | 1.34 (0.47, 2.22) | 2.09 (1.01, 3.18) |
| Côte d'Ivoire | 0.60 (0.25, 0.95) | 0.62 (0.26, 0.97) | 13.02 (11.51, 14.53) | 2.28 (1.61, 2.95) | 0.07 (-0.05, 0.19) | 0.00 (-0.02, 0.03) | 83.42 (81.75, 85.08) |
| Democratic Republic of the Congo | 0.04 (-0.02, 0.11) | 14.90 (13.82, 15.98) | 10.72 (9.78, 11.65) | 31.01 (29.60, 32.41) | 0.02 (-0.02, 0.06) | 1.73 (1.33, 2.12) | 41.58 (40.09, 43.08) |
| Eswatini | 10.26 (7.25, 13.27) | 46.93 (41.97, 51.88) | 0.08 (-0.20, 0.35) | 0.87 (-0.05, 1.79) | 0.03 (-0.15, 0.22) | NA | 41.83 (36.93, 46.72) |
| Ethiopia | 10.01 (8.14, 11.87) | 12.97 (10.88, 15.06) | 28.95 (26.13, 31.76) | 32.03 (29.13, 34.93) | 0.81 (0.25, 1.37) | NA | 15.24 (13.00, 17.47) |
| Gabon | 1.08 (0.50, 1.65) | 20.88 (18.63, 23.14) | 2.45 (1.59, 3.30) | 18.44 (16.29, 20.59) | 0.06 (-0.08, 0.19) | NA | 57.10 (54.35, 59.84) |
| The Gambia | 1.17 (0.64, 1.70) | 17.86 (15.97, 19.75) | 7.51 (6.21, 8.81) | 1.82 (1.16, 2.48) | 3.41 (2.52, 4.31) | 41.71 (39.28, 44.14) | 26.52 (24.34, 28.70) |
| Ghana | 0.47 (0.17, 0.77) | 0.74 (0.36, 1.11) | 29.00 (27.00, 31.01) | 17.68 (16.00, 19.37) | 1.09 (0.63, 1.55) | 4.40 (3.49, 5.30) | 46.63 (44.42, 48.83) |
| Guinea | 0.46 (0.10, 0.81) | 14.68 (12.82, 16.53) | 20.00 (17.90, 22.10) | 46.65 (44.03, 49.26) | 0.38 (0.06, 0.71) | 3.10 (2.19, 4.01) | 14.74 (12.88, 16.59) |
| Guinea-Bissau | 17.51 (15.45, 19.57) | 3.00 (2.08, 3.93) | 26.41 (24.01, 28.80) | 1.42 (0.78, 2.06) | 9.17 (7.61, 10.74) | 0.12 (-0.07, 0.30) | 42.37 (39.69, 45.05) |
| Kenya | 9.15 (8.22, 10.08) | 1.51 (1.12, 1.91) | 21.30 (19.97, 22.62) | 0.50 (0.27, 0.73) | 0.26 (0.09, 0.42) | NA | 67.28 (65.76, 68.80) |
| Lesotho | 1.34 (0.36, 2.33) | 1.82 (0.68, 2.97) | 11.56 (8.82, 14.30) | 35.37 (31.27, 39.46) | 4.16 (2.45, 5.87) | 17.50 (14.24, 20.75) | 28.25 (24.39, 32.10) |
| Liberia | 2.05 (1.20, 2.91) | 0.45 (0.05, 0.85) | 34.68 (31.81, 37.54) | 8.69 (7.00, 10.39) | 9.37 (7.62, 11.13) | 21.93 (19.44, 24.42) | 22.82 (20.30, 25.35) |
| Madagascar | 44.34 (42.33, 46.36) | 0.19 (0.01, 0.36) | 26.09 (24.31, 27.87) | 19.30 (17.70, 20.90) | 0.02 (-0.04, 0.07) | 0.62 (0.30, 0.94) | 9.44 (8.26, 10.63) |
| Malawi | 39.70 (37.94, 41.46) | 0.54 (0.28, 0.81) | 32.03 (30.35, 33.71) | 6.82 (5.91, 7.73) | 3.01 (2.40, 3.63) | 10.32 (9.22, 11.41) | 7.57 (6.62, 8.52) |
| Mali | 0.31 (0.11, 0.52) | 9.14 (8.08, 10.20) | 21.36 (19.85, 22.87) | 7.48 (6.51, 8.45) | 0.10 (-0.02, 0.22) | 5.42 (4.59, 6.25) | 56.18 (54.36, 58.00) |
| Mauritania | 7.65 (6.51, 8.78) | 3.83 (3.01, 4.65) | 25.24 (23.38, 27.10) | 5.26 (4.31, 6.22) | 0.46 (0.17, 0.75) | 36.90 (34.83, 38.96) | 20.66 (18.93, 22.39) |
| Mozambique | 10.79 (9.31, 12.28) | 10.89 (9.40, 12.38) | 6.09 (4.95, 7.24) | 45.54 (43.15, 47.92) | 0.08 (-0.06, 0.21) | 11.98 (10.43, 13.54) | 14.63 (12.94, 16.32) |
| Nigeria | 16.74 (15.70, 17.78) | 0.04 (-0.02, 0.09) | 47.45 (46.06, 48.85) | 17.34 (16.28, 18.39) | 0.20 (0.08, 0.33) | 2.05 (1.65, 2.44) | 16.19 (15.16, 17.22) |
| Rwanda | 3.29 (2.41, 4.18) | 13.44 (11.75, 15.13) | 1.50 (0.90, 2.10) | 22.66 (20.59, 24.73) | 0.18 (-0.03, 0.38) | 2.30 (1.56, 3.04) | 56.63 (54.18, 59.08) |
| São Tomé and Príncipe | 5.66 (3.15, 8.16) | 9.68 (6.48, 12.89) | 4.73 (2.43, 7.03) | 23.33 (18.75, 27.91) | 11.51 (8.05, 14.97) | 0.55 (-0.25, 1.36) | 44.54 (39.15, 49.93) |
| Senegal | 1.41 (0.90, 1.92) | 0.50 (0.19, 0.81) | 12.23 (10.81, 13.65) | 12.12 (10.71, 13.54) | 3.68 (2.87, 4.50) | 1.42 (0.90, 1.93) | 68.63 (66.62, 70.65) |
| Sierra Leone | 15.61 (13.95, 17.26) | 5.54 (4.50, 6.58) | 2.00 (1.36, 2.64) | 7.50 (6.30, 8.70) | 0.52 (0.19, 0.84) | 4.28 (3.35, 5.20) | 64.56 (62.39, 66.74) |
| South Africa | 0.00 (-0.03, 0.03) | 0.42 (-0.08, 0.92) | 1.75 (0.74, 2.77) | 1.05 (0.26, 1.84) | 17.46 (14.52, 20.39) | NA | 79.32 (76.18, 82.45) |
| Tanzania | 8.08 (6.92, 9.24) | 0.13 (-0.02, 0.29) | 13.12 (11.69, 14.56) | 0.66 (0.32, 1.01) | 0.14 (-0.02, 0.30) | 0.17 (-0.01, 0.34) | 77.70 (75.93, 79.47) |
| Togo | 7.87 (6.10, 9.65) | 3.77 (2.51, 5.02) | 26.85 (23.93, 29.78) | 25.68 (22.80, 28.56) | 0.08 (-0.11, 0.27) | 0.23 (-0.09, 0.54) | 35.51 (32.36, 38.67) |
| Uganda | 19.84 (18.39, 21.29) | 0.62 (0.33, 0.90) | 10.28 (9.18, 11.38) | 1.73 (1.26, 2.21) | 1.02 (0.65, 1.38) | 1.10 (0.72, 1.48) | 65.41 (63.69, 67.14) |
| Zambia | 4.29 (3.38, 5.19) | 0.78 (0.38, 1.17) | 36.93 (34.78, 39.09) | 7.27 (6.11, 8.43) | 3.34 (2.54, 4.14) | 18.64 (16.90, 20.38) | 28.76 (26.73, 30.78) |
| Zimbabwe | 7.62 (6.04, 9.21) | 10.38 (8.55, 12.20) | 0.29 (-0.03, 0.61) | 1.98 (1.14, 2.81) | 41.82 (38.86, 44.77) | 0.27 (-0.04, 0.57) | 37.65 (34.76, 40.55) |
| **Region of the Americas** |  |  |  |  |  |  |  |
| Costa Rica | 3.50 (2.11, 4.89) | 0.37 (-0.09, 0.83) | 0.94 (0.21, 1.67) | 6.10 (4.29, 7.91) | 2.14 (1.05, 3.24) | 2.47 (1.30, 3.65) | 84.48 (81.73, 87.22) |
| Cuba | 10.72 (8.89, 12.56) | 7.26 (5.72, 8.80) | 5.28 (3.95, 6.61) | 2.10 (1.25, 2.95) | 18.88 (16.56, 21.21) | NA | 55.76 (52.81, 58.70) |
| Dominican Republic | 0.78 (0.33, 1.24) | 1.70 (1.03, 2.37) | 0.52 (0.14, 0.89) | 3.92 (2.91, 4.93) | 0.22 (-0.02, 0.47) | 1.51 (0.88, 2.14) | 91.35 (89.90, 92.81) |
| Guatemala | 1.13 (0.71, 1.55) | 14.42 (13.01, 15.82) | 24.85 (23.12, 26.58) | 1.02 (0.62, 1.42) | 3.43 (2.70, 4.15) | 8.90 (7.76, 10.04) | 46.26 (44.27, 48.25) |
| Guyana | 3.67 (2.00, 5.34) | 9.56 (6.96, 12.17) | 0.35 (-0.17, 0.88) | 4.09 (2.33, 5.84) | 0.00 (-0.05, 0.05) | 55.49 (51.08, 59.89) | 26.84 (22.91, 30.76) |
| Haiti | 0.23 (-0.04, 0.50) | 2.79 (1.85, 3.72) | 16.92 (14.80, 19.05) | 30.87 (28.25, 33.49) | 8.81 (7.21, 10.42) | 27.49 (24.96, 30.02) | 12.89 (10.99, 14.79) |
| Honduras | 8.84 (7.45, 10.22) | 2.43 (1.68, 3.18) | 18.00 (16.13, 19.88) | 9.42 (8.00, 10.84) | 0.84 (0.39, 1.28) | NA | 60.47 (58.08, 62.85) |
| Suriname | 16.73 (13.75, 19.71) | 7.27 (5.19, 9.34) | 0.40 (-0.10, 0.91) | 0.28 (-0.14, 0.70) | 1.58 (0.59, 2.58) | 0.68 (0.03, 1.34) | 73.06 (69.51, 76.60) |
| Trinidad and Tobago | 28.82 (22.05, 35.59) | 8.10 (4.02, 12.17) | 4.23 (1.22, 7.24) | 0.02 (-0.20, 0.24) | 33.26 (26.22, 40.30) | 0.77 (-0.54, 2.07) | 24.81 (18.35, 31.26) |
| **Eastern Mediterranean Region** |  |  |  |  |  |  |  |
| Afghanistan | 0.18 (0.04, 0.31) | 0.05 (-0.02, 0.13) | 23.90 (22.53, 25.28) | 0.91 (0.60, 1.21) | 7.31 (6.47, 8.14) | NA | 67.65 (66.15, 69.16) |
| Algeria | 1.01 (0.63, 1.39) | 0.62 (0.32, 0.92) | 16.30 (14.89, 17.72) | 10.55 (9.37, 11.73) | 0.57 (0.28, 0.85) | 1.35 (0.91, 1.79) | 69.60 (67.84, 71.36) |
| Iraq | 3.05 (2.40, 3.70) | 7.98 (6.95, 9.01) | 33.55 (31.76, 35.34) | 24.44 (22.81, 26.07) | 1.45 (1.00, 1.90) | 16.64 (15.23, 18.05) | 12.89 (11.62, 14.16) |
| Jordan | 10.40 (8.92, 11.87) | 2.21 (1.50, 2.93) | 46.27 (43.86, 48.68) | 39.05 (36.69, 41.41) | 0.55 (0.19, 0.90) | 0.07 (-0.06, 0.19) | 1.46 (0.88, 2.04) |
| Pakistan | 14.52 (13.93, 15.12) | 0.32 (0.23, 0.42) | 71.01 (70.25, 71.78) | 1.80 (1.58, 2.03) | 0.13 (0.07, 0.19) | 0.21 (0.13, 0.29) | 11.99 (11.44, 12.54) |
| Palestine | 25.12 (22.52, 27.72) | 1.60 (0.84, 2.35) | 1.04 (0.43, 1.64) | 19.24 (16.88, 21.61) | 0.35 (-0.01, 0.70) | 1.89 (1.07, 2.70) | 50.77 (47.77, 53.77) |
| Tunisia | 11.70 (7.80, 15.61) | 16.91 (12.36, 21.46) | 0.92 (-0.24, 2.07) | 2.13 (0.38, 3.88) | 25.55 (20.26, 30.85) | 1.15 (-0.14, 2.44) | 41.64 (35.66, 47.62) |
| Yemen | 1.03 (0.60, 1.46) | 12.27 (10.87, 13.66) | 0.49 (0.19, 0.78) | 0.17 (0.00, 0.35) | 0.19 (0.00, 0.37) | 5.00 (4.08, 5.93) | 80.86 (79.18, 82.53) |
| **European Region** |  |  |  |  |  |  |  |
| Armenia | 10.03 (6.86, 13.21) | 1.81 (0.41, 3.22) | 0.64 (-0.20, 1.49) | 8.77 (5.78, 11.75) | 0.93 (-0.08, 1.95) | 49.28 (44.00, 54.55) | 28.53 (23.77, 33.30) |
| Azerbaijan | 0.60 (-1.29, 2.49) | 1.33 (-1.48, 4.14) | 0.37 (-1.12, 1.87) | 7.58 (1.10, 14.07) | 0.33 (-1.08, 1.75) | NA | 89.78 (82.35, 97.20) |
| Kyrgyzstan | 30.22 (25.52, 34.92) | 2.71 (1.05, 4.37) | 9.81 (6.77, 12.86) | 5.17 (2.90, 7.43) | 0.02 (-0.12, 0.16) | NA | 52.07 (46.96, 57.18) |
| North Macedonia | 30.87 (25.50, 36.23) | 9.99 (6.51, 13.48) | 18.96 (14.41, 23.51) | 9.31 (5.94, 12.69) | 0.15 (-0.30, 0.59) | 29.53 (24.24, 34.83) | 1.18 (-0.07, 2.44) |
| Serbia | 5.24 (2.83, 7.65) | 30.41 (25.43, 35.39) | 1.63 (0.26, 3.00) | 16.56 (12.53, 20.58) | 0.19 (-0.28, 0.67) | 1.86 (0.40, 3.33) | 44.10 (38.73, 49.48) |
| Tajikistan | 34.39 (31.36, 37.41) | 5.23 (3.81, 6.64) | 0.17 (-0.09, 0.43) | 0.25 (-0.07, 0.57) | 0.42 (0.01, 0.83) | NA | 59.55 (56.42, 62.68) |
| Turkey | 1.92 (0.65, 3.19) | 5.43 (3.33, 7.53) | 11.07 (8.16, 13.98) | 13.97 (10.76, 17.18) | 6.45 (4.17, 8.73) | 14.12 (10.89, 17.35) | 47.05 (42.42, 51.68) |
| Uzbekistan | 0.94 (0.34, 1.53) | 6.86 (5.30, 8.42) | 0.18 (-0.08, 0.44) | 10.70 (8.79, 12.61) | 0.12 (-0.09, 0.33) | NA | 81.21 (78.79, 83.62) |
| **South-East Asia Region** |  |  |  |  |  |  |  |
| Bangladesh | 0.07 (-0.06, 0.20) | 0.48 (0.14, 0.81) | 22.05 (20.06, 24.04) | 7.92 (6.62, 9.21) | 4.40 (3.42, 5.39) | 4.74 (3.72, 5.76) | 60.34 (57.99, 62.69) |
| India | 2.50 (2.35, 2.65) | 6.71 (6.47, 6.94) | 19.10 (18.73, 19.47) | 13.54 (13.22, 13.86) | 1.01 (0.92, 1.10) | 5.12 (4.91, 5.33) | 52.02 (51.55, 52.49) |
| Indonesia | 4.63 (3.94, 5.33) | 2.59 (2.06, 3.11) | 11.94 (10.87, 13.02) | 6.41 (5.60, 7.22) | 0.51 (0.27, 0.74) | 2.20 (1.72, 2.69) | 71.72 (70.23, 73.21) |
| Myanmar | 11.05 (9.02, 13.08) | 1.68 (0.85, 2.52) | 14.80 (12.50, 17.11) | 19.51 (16.94, 22.08) | 1.20 (0.50, 1.91) | 13.26 (11.06, 15.46) | 38.49 (35.33, 41.64) |
| Nepal | 12.22 (10.19, 14.26) | 0.66 (0.16, 1.16) | 9.22 (7.43, 11.02) | 21.13 (18.60, 23.67) | 0.94 (0.34, 1.53) | 0.28 (-0.05, 0.61) | 55.54 (52.46, 58.63) |
| Thailand | 43.41 (41.14, 45.68) | 1.27 (0.76, 1.78) | 1.39 (0.85, 1.92) | 6.10 (5.00, 7.20) | 0.02 (-0.04, 0.08) | 6.25 (5.14, 7.36) | 41.56 (39.31, 43.82) |
| Timor-Leste | 2.25 (1.48, 3.02) | 0.66 (0.24, 1.08) | 23.47 (21.27, 25.68) | 48.79 (46.19, 51.40) | 1.13 (0.58, 1.68) | NA | 23.70 (21.48, 25.91) |
| **Western Pacific Region** |  |  |  |  |  |  |  |
| Cambodia | 2.35 (1.63, 3.08) | 0.50 (0.16, 0.84) | 1.33 (0.78, 1.88) | 15.31 (13.58, 17.04) | 0.00 (-0.03, 0.04) | 6.26 (5.09, 7.42) | 74.24 (72.14, 76.34) |
| Fiji | 23.36 (19.11, 27.61) | 0.75 (-0.11, 1.62) | 0.83 (-0.08, 1.74) | 0.13 (-0.23, 0.48) | 0.40 (-0.23, 1.03) | 2.63 (1.03, 4.24) | 71.89 (67.38, 76.41) |
| Laos | 3.52 (2.55, 4.49) | 5.61 (4.40, 6.83) | 0.13 (-0.06, 0.31) | 19.57 (17.48, 21.67) | 0.48 (0.11, 0.84) | 0.14 (-0.06, 0.34) | 70.55 (68.14, 72.96) |
| Mongolia | 9.63 (7.72, 11.54) | 0.27 (-0.07, 0.61) | 8.85 (7.01, 10.68) | 7.35 (5.66, 9.04) | 1.71 (0.87, 2.55) | NA | 72.19 (69.29, 75.09) |
| Papua New Guinea | 0.37 (0.09, 0.66) | 2.05 (1.39, 2.72) | 33.46 (31.25, 35.66) | 49.45 (47.12, 51.78) | 0.24 (0.01, 0.46) | 0.84 (0.42, 1.27) | 13.59 (11.99, 15.18) |
| Philippines | 30.83 (28.54, 33.11) | 4.16 (3.17, 5.15) | 5.07 (3.98, 6.15) | 12.64 (11.00, 14.29) | 0.43 (0.10, 0.75) | NA | 46.88 (44.41, 49.35) |
| Tonga | 1.37 (-0.47, 3.20) | 31.28 (23.96, 38.60) | 0.59 (-0.62, 1.80) | 1.52 (-0.41, 3.46) | 20.16 (13.83, 26.50) | 0.76 (-0.61, 2.14) | 44.31 (36.47, 52.16) |
| Vanuatu | 5.76 (3.19, 8.34) | 13.94 (10.11, 17.78) | 0.79 (-0.19, 1.77) | 15.46 (11.46, 19.46) | 2.80 (0.97, 4.62) | NA | 61.24 (55.85, 66.63) |
| Vietnam | 30.30 (26.85, 33.75) | 1.48 (0.57, 2.39) | 2.26 (1.14, 3.37) | 28.40 (25.01, 31.78) | 21.77 (18.67, 24.87) | 0.13 (-0.14, 0.40) | 15.67 (12.94, 18.40) |

Geographic and contextual factors also contributed importantly in selected countries. Subnational region was a major contributor in Madagascar (44.34%), Thailand (43.41%), and Malawi (39.70%), while place of residence showed large contributions in Eswatini (46.93%), Tonga (31.28%), and Serbia (30.41%). Health insurance coverage explained a substantial proportion of inequity in a smaller number of countries, including Guyana (55.49%), Armenia (49.28%), and Central African Republic (42.48%). Child sex generally contributed little to zero-dose inequity, although it was an important contributor in several settings, including Comoros (55.23%), Zimbabwe (41.82%), and Trinidad and Tobago (33.26%).

The residual component remained substantial in many countries, indicating that the measured equity dimensions did not fully capture all factors contributing to zero-dose inequity. The largest residual contributions were observed in Chad (93.63%), Dominican Republic (91.35%), and Azerbaijan (89.78%).

## Discussion

In this multicountry analysis of 170,553 children aged 12–23 months across 77 countries, we found substantial heterogeneity in zero-dose prevalence and multidimensional inequity both between and within countries. Zero-dose children were consistently concentrated among multidimensionally disadvantaged populations, particularly those facing socioeconomic and geographic disadvantages, whereas differences by child sex were minimal. Our subnational analyses showed that national averages frequently concealed localised pockets of under-vaccination, highlighting the importance of identifying inequities below the national level. Furthermore, although multidimensional inequity was widespread, its magnitude and contributing dimensions varied considerably across countries, suggesting that the drivers of zero-dose vaccination gaps are context specific. Together, these findings demonstrate that measuring both burden and inequity is essential for understanding where zero-dose children are concentrated and for informing more targeted approaches to improve equitable immunisation coverage.

Our country-specific estimates were broadly consistent with recent zero-dose estimation studies, although differences across data sources were also apparent. For India, our zero-dose prevalence estimate of 6.17% was lower than the 10.1% (95% CI 9.8–10.4) reported by Johri and colleagues using the 2015–2016 National Family Health Survey, and was consistent with the longer-term decline in zero-dose prevalence documented in that study, from 33.4% in 1992 to 10.1% in 2016.^11^ In Nigeria, we estimated a zero-dose prevalence of 36.79% in 2024, compared with 30.1% (95% CI, 27.9–32.4%) reported from the 2021 MICS/National Immunization Coverage Survey, with both estimates indicating a persistently high burden of children unreached by routine immunisation services.^18^ By contrast, our estimate for the Democratic Republic of the Congo (33.10% in 2023) was substantially higher than the 19.1% reported from a provincial vaccination coverage survey conducted in 2021–2022. Notably, that study documented pronounced provincial variation, ranging from 2.4% to 62.4%, consistent with the substantial subnational heterogeneity observed in our analysis.^19^ At the global level, the Global Burden of Disease Study 2023 estimated that 15.7 million children remained zero-dose in 2023, with more than half residing in only eight countries, including Nigeria, India, the Democratic Republic of the Congo, and Ethiopia.^20^ Taken together, these comparisons situate our findings within the broader zero-dose estimation literature while also highlighting that direct numerical comparisons should be interpreted cautiously because estimates differ in survey year, sampling frame, population denominator, and estimation approach.

The marked socioeconomic inequalities by maternal education, household wealth, residence, and health insurance status indicate that zero-dose status is closely embedded in broader patterns of social disadvantage. They also align with multicounty evidence showing persistent wealth-related and education-related inequalities in childhood immunisation coverage across low-income and middle-income countries.^13^ Importantly, these inequalities should not be interpreted as isolated individual risk factors. Maternal education may reflect health literacy, autonomy, contact with services, and ability to navigate vaccination schedules, while household wealth and insurance coverage may capture broader access to transportation, opportunity costs, and health-system contact. Similar conclusions have been drawn from multidimensional vaccine equity analyses in China, where high overall coverage did not eliminate inequity and socioeconomic characteristics continued to explain substantial variation in vaccine uptake.^21^

The subnational findings show that national averages can conceal highly localised pockets of zero-dose children. Although some countries had moderate national prevalence, selected ADM1 areas had much higher levels of zero-dose status. This pattern is programmatically important because routine immunisation strategies designed around national averages may fail to identify the specific regions where children are most likely to be missed. Previous work on subnational inequality monitoring has emphasised that geographic disaggregation is essential for unmasking within-country differences and supporting targeted, equity-oriented interventions.^22^ Our results are consistent with this evidence, showing that the highest subnational prevalence was concentrated in specific areas of the Democratic Republic of the Congo, Angola, the Philippines, Nigeria, and several other countries, rather than being evenly distributed. However, the subnational estimates should be interpreted as signals for prioritisation rather than as precise rankings of all ADM1 units. Some extreme values, such as 100% prevalence in a very small ADM1 sample, are likely influenced by sparse data. For this reason, we treated national results as the primary basis for interpretation and used subnational estimates to identify geographic clustering and generate hypotheses for local programme review.

The joint distribution of national zero-dose prevalence and concentration index highlights the complementary information provided by burden and inequity. Countries with high prevalence and moderate-to-high CI, such as Mozambique, Ethiopia, Azerbaijan, and Nigeria, face both a substantial zero-dose burden and disproportionate concentration among disadvantaged children, suggesting the need for broad immunisation strengthening alongside targeted strategies. This interpretation is consistent with modelling evidence supporting geographically tailored immunisation planning under constrained resources.^7^ By contrast, Guinea and Papua New Guinea had high prevalence but comparatively lower CI values, suggesting a more widely distributed burden, whereas São Tomé and Príncipe, Tunisia, Zimbabwe, Fiji, Tonga, and Uzbekistan had low prevalence but very high CI values, indicating that the remaining zero-dose children were strongly concentrated among disadvantaged groups. This distinction is consistent with the IA2030 emphasis on reducing overall zero-dose burden while reaching underserved populations.^6^ At the subnational level, extreme CI values also occurred predominantly in low-prevalence settings, underscoring that relative CI values should be interpreted jointly with zero-dose prevalence and absolute equity measures.

The decomposition results suggest that no single equity dimension explains zero-dose inequity across all countries. Maternal education was the dominant contributor in countries such as Pakistan, Nigeria, Jordan, Cameroon, and Benin, whereas household wealth contributed most strongly in Angola, Papua New Guinea, Timor-Leste, Guinea, and Mozambique. Geographic region and place of residence were the main contributors in other countries, and health insurance explained a substantial share of inequity in Guyana, Armenia, Central African Republic, The Gambia, and Mauritania. These findings argue against one-size-fits-all explanations for zero-dose status. In countries where maternal education dominates, interventions may need to strengthen caregiver communication, community engagement, and links between maternal and child health services. Where wealth is the dominant contributor, reducing financial and indirect costs of vaccination, including transport and missed work, may be more relevant. Where geography contributes strongly, microplanning, outreach services, cold-chain reliability, and tailored delivery in remote or underserved areas may be more important. The diversity of decomposition patterns is also consistent with recent work highlighting that zero-dose children face multiple social, political, economic, and service-delivery barriers, and that strategies need to be tailored to local contexts.^23^

Several limitations should be considered when interpreting these findings. First, the analysis was based on cross-sectional household surveys, so the associations between social characteristics and zero-dose status should not be interpreted as causal. To reduce this concern, we focused on descriptive and equity-oriented interpretation rather than causal attribution, and used nationally representative DHS and MICS surveys with standardised sampling procedures wherever available. Second, vaccination information and covariates differed between DHS and MICS and across countries, including missing health insurance information in some surveys. We addressed this by harmonising variables using common DHS/MICS definitions, defining zero-dose status consistently based on non-receipt of any DTP-containing vaccine among surviving children aged 12-23 months, and using a modified specification when health insurance was unavailable rather than forcing non-comparable variables into the model. Third, subnational equity estimates should be interpreted cautiously because some ADM1 units had small samples or few zero-dose events. For binary outcomes, the conventional concentration index is mean-dependent and can be sensitive when prevalence is very low, as reflected by the concentration of extreme CI values in low-prevalence settings in our analysis.^24^ We therefore interpreted subnational CI values jointly with zero-dose prevalence and the absolute equity gap.

In conclusion, this study shows that zero-dose children remain unevenly distributed both across and within countries. High-burden countries require broad strengthening of routine immunisation systems, while low-prevalence but high-CI countries require strategies that identify and reach the small number of children still left behind. The large variation in decomposition patterns further indicates that equity-oriented immunisation policy must be country-specific: maternal education, household wealth, geography, health insurance, and other unmeasured barriers each dominate in different settings. Integrating national prevalence, subnational mapping, multidimensional equity metrics, and decomposition analysis can help translate the IA2030 goal of reaching zero-dose children into more targeted and locally actionable immunisation strategies.

## Methods

### Study design and participants

We conducted a cross-sectional, multicountry analysis using nationally representative household survey data from the Demographic and Health Surveys (DHS) and Multiple Indicator Cluster Surveys (MICS). Both survey programmes use standardised questionnaires and multistage stratified cluster sampling designs to collect comparable information on child health, immunisation, and household socioeconomic characteristics. We included the most recent DHS or MICS survey conducted in each eligible country from 2015 onwards. When multiple surveys were available during this period, we selected the latest survey containing the required vaccination and equity-related variables. Details of the included countries, survey platforms, survey years and sample sizes are provided in **Supplementary Table 1**.

We excluded countries without usable vaccination variables, one country without a standardised ISO3C country code and countries in which zero-dose prevalence was estimated to be 0.0%. At the individual level, we excluded children who had died before the survey, children outside the target age range of 12-23 months according to the definition of zero-dose and children with missing core variables for our equity analysis. The final analytic sample included 170,553 children from 77 countries across 1,571 first-level administrative areas. The specific data selection process is shown in **Supplementary Fig. 1**.

### Outcome definition

The primary outcome was zero-dose status, defined as surviving children aged 12-23 months who had not received any dose of a DTP-containing vaccine.^5^ Zero-dose prevalence was calculated as the weighted proportion of children classified as zero-dose. In settings where pentavalent vaccine was recorded instead of standalone DTP vaccine, receipt of the first dose of pentavalent vaccine was treated as receipt of a DTP-containing vaccine. Children who had received at least one dose of DTP or pentavalent vaccine were classified as non-zero-dose.

Vaccination status was obtained from DHS and MICS child immunisation modules. When available, vaccination histories were extracted from home-based vaccination cards or child health records; otherwise, caregiver-reported vaccination information was used according to standard DHS and MICS procedures. Vaccination variables were harmonised across survey platforms before analysis to ensure consistent identification of DTP-containing vaccine receipt across countries.

### Equity dimensions and covariates

We assessed multidimensional inequity in zero-dose vaccination using a set of equity-related dimensions capturing socioeconomic, demographic, and geographic barriers to immunisation access. These dimensions were selected based on their established associations with childhood vaccination coverage and their availability across DHS and MICS surveys. The equity dimensions included household wealth, maternal education, geographic region, place of residence, child sex, and health insurance coverage where available.

Household wealth was measured using the DHS and MICS wealth index, which is derived through principal component analysis of household assets and characteristics. Maternal education was categorised according to survey-specific definitions and harmonised across countries. Geographic region and place of residence were obtained directly from survey records, representing subnational location and urban–rural residence, respectively. Child sex was included as a demographic characteristic, while health insurance coverage was incorporated as an indicator of access to health-related resources when available.

All equity-related variables were harmonised across surveys before analysis. Because variable availability differed between DHS and MICS surveys, health insurance coverage was not available for all countries and was therefore included only in analyses where information was reported.

### VERSE multidimensional equity analysis

We applied the Vaccine Economics Research for Sustainability and Equity (VERSE) framework to quantify multidimensional inequity in zero-dose vaccination. The VERSE framework extends conventional equity analyses by integrating multiple dimensions of disadvantage into a composite ranking, allowing assessment of whether health outcomes are disproportionately concentrated among disadvantaged populations.^25^ This approach is consistent with broader theoretical perspectives on health inequity measurement, which emphasise that inequities may arise from multiple overlapping dimensions of disadvantage rather than from single socioeconomic gradients alone.^26–28^

For each country, children were ranked according to a multidimensional disadvantage index derived from the equity dimensions described above, including household wealth, maternal education, geographic region, place of residence, child sex, and health insurance coverage when available. The index represents the relative socioeconomic and demographic position of each child within the survey population, with higher values indicating greater disadvantage. Zero-dose prevalence was then evaluated along this disadvantage gradient.

We quantified the concentration of zero-dose children across the disadvantage ranking using the concentration index (CI), which measures the degree of socioeconomic-related inequality in a health variable.^25,29^ The CI was calculated as:

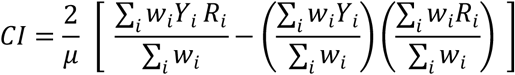

where Y*_i_* denotes zero-dose status, R_i_ denotes the weighted fractional rank of child *i* in the VERSE multidimensional disadvantage distribution, μ is the weighted mean zero-dose prevalence, and W_i_ represents the sampling weight. Because children were ranked from least to most disadvantaged and the outcome was adverse zero-dose status, positive concentration index values indicated that zero-dose children were disproportionately concentrated among more disadvantaged children. Values close to zero indicated a more equal distribution of zero-dose status across the multidimensional disadvantage ranking, whereas larger positive values indicated greater pro-disadvantaged concentration of zero-dose status.

Absolute inequality was measured using the Absolute Equity Gap (AEG), defined as the difference in zero-dose prevalence between the most disadvantaged and least disadvantaged quintiles of the VERSE ranking:

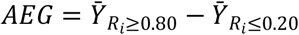

where *Y_Ri_*_≥_*_0.80_* denotes the weighted zero-dose prevalence among children in the highest quintile of multidimensional disadvantage, and *Y_Ri_*_≤_*_0.20_* denotes the weighted zero-dose prevalence among children in the lowest quintile. A positive AEG therefore indicated that zero-dose prevalence was higher among the most disadvantaged children than among the least disadvantaged children.

To identify the contribution of individual equity dimensions to observed multidimensional inequity, we decomposed the concentration index into the contributions attributable to each component of the disadvantage ranking. The decomposition approach follows the Blinder–Oaxaca framework, which has been widely used to partition differences in outcomes into contributions from observed characteristics and unexplained components.^30,31^ Within the VERSE framework, this approach allows quantification of how each equity dimension contributes to the overall concentration of zero-dose children among disadvantaged populations. The concentration index was decomposed as:

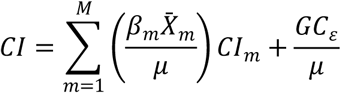

where *X_m_* denotes the *m* - th equity determinant, β_m_ is the corresponding regression coefficient or marginal effect, μ is the weighted mean zero-dose prevalence, *CI_m_* is the concentration index of *X_m_* over the VERSE multidimensional disadvantage ranking, and GC_ε_ is the generalised concentration index of the residual term. Percentage contributions were obtained by dividing each determinant-specific contribution by the overall concentration index. The residual component represents the share of inequity not explained by the included equity dimensions. Decomposition analyses were conducted separately for each country, allowing the dominant contributors to zero-dose inequity to vary across settings.

## Statistical analysis

We first summarised the characteristics of the study population using survey-weighted descriptive statistics. Zero-dose prevalence was estimated for each country and first-level administrative area (ADM1) using the sampling weights provided by DHS and MICS surveys. All estimates accounted for the complex survey design by incorporating survey weights, primary sampling units, and stratification variables where available.

Country-level and subnational estimates of zero-dose prevalence were calculated with corresponding 95% confidence intervals. We summarised multidimensional inequity using country-specific concentration index (CI) and absolute equity gap (AEG) estimates derived from the VERSE framework. Countries were classified according to the joint distribution of zero-dose prevalence and CI to identify settings with different combinations of disease burden and inequity profiles. Decomposition results were summarised by quantifying the relative contribution of each equity dimension to overall multidimensional inequity. All analyses were performed using R version 4.4.3.

## Ethics statement

This study used publicly available, deidentified secondary data from DHS and MICS. Ethical approval for primary data collection was obtained by the relevant national ethical review committees and survey implementing agencies. Permission to access and analyse the datasets was obtained from the DHS Program and UNICEF MICS.

## Data availability

DHS and MICS datasets are publicly available to registered users through the DHS Program (https://dhsprogram.com/) and UNICEF MICS (https://mics.unicef.org/) platforms. Administrative boundary data are available from the Global Administrative Areas database. The harmonised analytic dataset generated for this study contains recoded survey variables and will be made available by the corresponding author upon reasonable request, subject to the data-use terms of DHS and MICS.

## Code availability

The R code used to harmonise variables, implement the modified VERSE workflow, calculate inequality metrics and generate figures will be made available in a public repository upon publication.

## Acknowledgements

We thank the DHS Program, UNICEF MICS and the national survey teams and participants who made these analyses possible.

## Author contributions

Conceptualization: H.Z. and B.P. Data curation: C.H. and H.Z. Formal analysis: C.H. and H.Z. Funding acquisition: H.Z. and B.P. Methodology: H.Z. and B.P. Software: C.H. Supervision: H.Z., X.Y., H.F. and B.P. Writing—original draft preparation: C.H. and H.Z. Writing—review and editing: R.C., Y.G., C.Y., G.D.B., H.F., X.Y., H.Z. and B.P.

## Additional information

Supplementary information is available for this paper. Correspondence and requests for materials should be addressed to the corresponding authors.

