## Supplementary Information for "Mapping national and subnational zero-dose prevalence and multidimensional inequities across 77 countries"

**Supplemental Online Content**

**Authors:** Chuyao Huang, PhD[1]; Ruixin Chi, PhD[2]; Yanhong Gong, PhD[1]; Chenyu Yan, PhD[3]; Gatien De Broucker, PhD[4,5]; Hai Fang, PhD[6,7†]; Xiaoxv Yin, PhD[1†]; Haijun Zhang, PhD[1†]; Bryan Patenaude, ScD[4,5]

**Affiliations:**

1 School of Public Health, Tongji Medical College, Huazhong University of Science and Technology, Wuhan, Hubei, 430030, China

2 Lee Kong Chian School of Medicine, Nanyang Technological University, Singapore, 308232, Singapore

3 Vanke School of Public Health, Tsinghua University, Beijing, 100084, China

4 International Vaccine Access Center, Johns Hopkins Bloomberg School of Public Health, Baltimore, MD， 21231, USA

5 Department of International Health, Johns Hopkins Bloomberg School of Public Health, Baltimore, MD, 21231, USA

6 China Center for Health Development Studies, Peking University, Beijing, 100191, China

7 School of Public Health, Peking University, Beijing, 100191, China

### Appendix: Mapping national and subnational zero-dose prevalence and multidimensional inequities across 77 countries

#### A: Data description

##### (1) Demographic and Health Surveys (DHS)

DHS is a globally recognized survey initiative designed to provide nationally representative data on population health, nutrition, and demographic indicators in low- and middle-income countries. Funded primarily by the United States Agency for International Development (USAID) and implemented by ICF International, the DHS Program has conducted standardized household surveys since the mid-1980s across more than 90 countries. DHS surveys employ rigorous multistage stratified cluster sampling methodologies to ensure national representativeness and cross-country comparability. Standardized questionnaires are administered to women of reproductive age, men, and households, covering a broad range of topics including fertility, maternal and child health, immunization, nutrition, mortality, infectious diseases, health service utilization, and socioeconomic characteristics. Survey implementation follows harmonized protocols for questionnaire design, interviewer training, field supervision, and data quality assurance, thereby facilitating reliable temporal and international comparisons. All DHS datasets are publicly accessible upon formal request through the official DHS Program platform, and de-identified individual-level data are made available for research purposes.

##### (2) Multiple Indicator Cluster Surveys (MICS)

MICS is an international household survey programme developed and supported by United Nations Children's Fund to assist countries in generating internationally comparable estimates on the health, wellbeing, and living conditions of women and children. Since its inception in the mid-1990s, MICS has been implemented in more than 100 countries and territories, with repeated survey rounds enabling trend analyses over time. MICS surveys utilize standardized multistage probability sampling procedures to produce nationally representative estimates at national and subnational levels. The surveys collect extensive information on child health, immunization, nutrition, early childhood development, education, water and sanitation, maternal health, household characteristics, and other indicators relevant to monitoring global development agendas, including the Sustainable Development Goals (SDGs). To ensure comparability and methodological rigor, MICS adopts unified survey instruments, standardized interviewer training protocols, and comprehensive quality control procedures across participating countries. De-identified datasets, accompanying documentation, and survey reports are publicly available through the official UNICEF MICS repository upon registration and approval for research use.

**Supplementary Table 1. Included DHS and MICS surveys by country and year**

| Country | Questionnaire | Year | Sample size |
|---|---|---|---|
| Afghanistan | MICS6 | 2023 | 32909 |
| Albania | DHS7 | 2017 | 2762 |
| Algeria | MICS6 | 2019 | 14736 |
| Angola | DHS7 | 2015 | 14322 |
| Argentina | MICS6 | 2020 | 6343 |
| Armenia | DHS7 | 2016 | 1724 |
| Azerbaijan | MICS6 | 2023 | 2548 |
| Bangladesh | DHS7 | 2017 | 8759 |
| Belarus | MICS6 | 2019 | 3544 |
| Benin | MICS6 | 2021 | 13062 |
| Burkina Faso | DHS8 | 2021 | 12343 |
| Burundi | DHS7 | 2016 | 13192 |
| Cambodia | DHS8 | 2021 | 8153 |
| Cameroon | DHS7 | 2018 | 9733 |
| Central African Republic | MICS6 | 2019 | 8829 |
| Chad | MICS6 | 2019 | 21776 |
| Colombia | DHS7 | 2015 | 11759 |
| Comoros | MICS6 | 2022 | 4429 |
| Costa Rica | MICS6 | 2018 | 3591 |
| Côte d’Ivoire | DHS8 | 2021 | 10645 |
| Cuba | MICS6 | 2019 | 5239 |
| Democratic Republic of the Congo | DHS8 | 2023 | 24203 |
| Dominican Republic | MICS6 | 2019 | 8348 |
| Eswatini | MICS6 | 2021 | 2239 |
| Ethiopia | DHS7 | 2018 | 5753 |
| Fiji | MICS6 | 2021 | 2110 |
| Gabon | DHS8 | 2021 | 6376 |
| Ghana | DHS8 | 2022 | 9353 |

| Country | Questionnaire | Year | Sample size |
| --- | --- | --- | --- |
| Georgia | MICS6 | 2018 | 2824 |
| Guatemala | DHS7 | 2015 | 12440 |
| Guinea | DHS7 | 2018 | 7951 |
| Guinea-Bissau | MICS6 | 2019 | 7414 |
| Guyana | MICS6 | 2019 | 2776 |
| Haiti | DHS7 | 2017 | 6530 |
| Honduras | MICS6 | 2019 | 8400 |
| India | DHS7 | 2019 | 232920 |
| Indonesia | DHS7 | 2017 | 17848 |
| Iraq | MICS6 | 2018 | 16580 |
| Jamaica | MICS6 | 2022 | 1423 |
| Jordan | DHS8 | 2023 | 9106 |
| Kenya | DHS8 | 2022 | 19530 |
| Kiribati | MICS6 | 2018 | 2157 |
| Kosovo | MICS6 | 2020 | 1781 |
| Kyrgyzstan | MICS6 | 2023 | 3140 |
| Laos | MICS6 | 2023 | 9243 |
| Lesotho | DHS8 | 2024 | 2537 |
| Liberia | DHS7 | 2019 | 5704 |
| Madagascar | DHS8 | 2021 | 12499 |
| Malawi | MICS6 | 2020 | 15409 |
| Mali | DHS8 | 2024 | 15631 |
| Mauritania | DHS7 | 2020 | 11628 |
| Mongolia | MICS6 | 2018 | 6065 |
| Montenegro | MICS6 | 2018 | 1329 |
| Mozambique | DHS8 | 2022 | 9289 |
| Myanmar | DHS7 | 2016 | 4815 |
| Nauru | MICS6 | 2023 | 349 |
| Nepal | DHS8 | 2021 | 5372 |
| Nigeria | DHS8 | 2024 | 27783 |
| North Macedonia | MICS6 | 2018 | 1496 |
| Pakistan | MICS6 | 2019 | 106883 |
| Palestine | MICS6 | 2019 | 6281 |
| Papua New Guinea | DHS7 | 2016 | 9514 |
| Philippines | DHS8 | 2022 | 8478 |
| Qatar | MICS6 | 2023 | 2985 |
| Rwanda | DHS7 | 2020 | 8092 |

| Country | Questionnaire | Year | Sample size |
|---|---|---|---|
| São Tomé and Príncipe | MICS6 | 2019 | 1822 |
| Senegal | DHS8 | 2023 | 10498 |
| Serbia | MICS6 | 2019 | 1821 |
| Sierra Leone | DHS7 | 2019 | 9899 |
| South Africa | DHS7 | 2016 | 3548 |
| Suriname | MICS6 | 2018 | 4201 |
| Tajikistan | DHS8 | 2023 | 5068 |
| Tanzania | DHS8 | 2022 | 10783 |
| Thailand | MICS6 | 2022 | 10418 |
| The Gambia | DHS7 | 2020 | 8362 |
| Timor-Leste | DHS7 | 2016 | 7221 |
| Togo | MICS6 | 2017 | 4915 |
| Tonga | MICS6 | 2019 | 1340 |
| Trinidad and Tobago | MICS6 | 2022 | 1638 |
| Tunisia | MICS6 | 2023 | 1912 |
| Turkey | DHS7 | 2018 | 2755 |
| Turkmenistan | MICS6 | 2019 | 3729 |
| Turks and Caicos Islands | MICS6 | 2019 | 306 |
| Tuvalu | MICS6 | 2019 | 499 |
| Uganda | DHS7 | 2016 | 15522 |
| Uzbekistan | MICS6 | 2021 | 7849 |
| Vanuatu | MICS6 | 2023 | 2032 |
| Vietnam | MICS6 | 2020 | 4287 |
| Yemen | MICS6 | 2023 | 19257 |
| Zambia | DHS7 | 2018 | 9959 |
| Zimbabwe | MICS6 | 2019 | 6047 |

**Supplementary Fig.1 Flowchart of the process for constructing the final analysis sample**

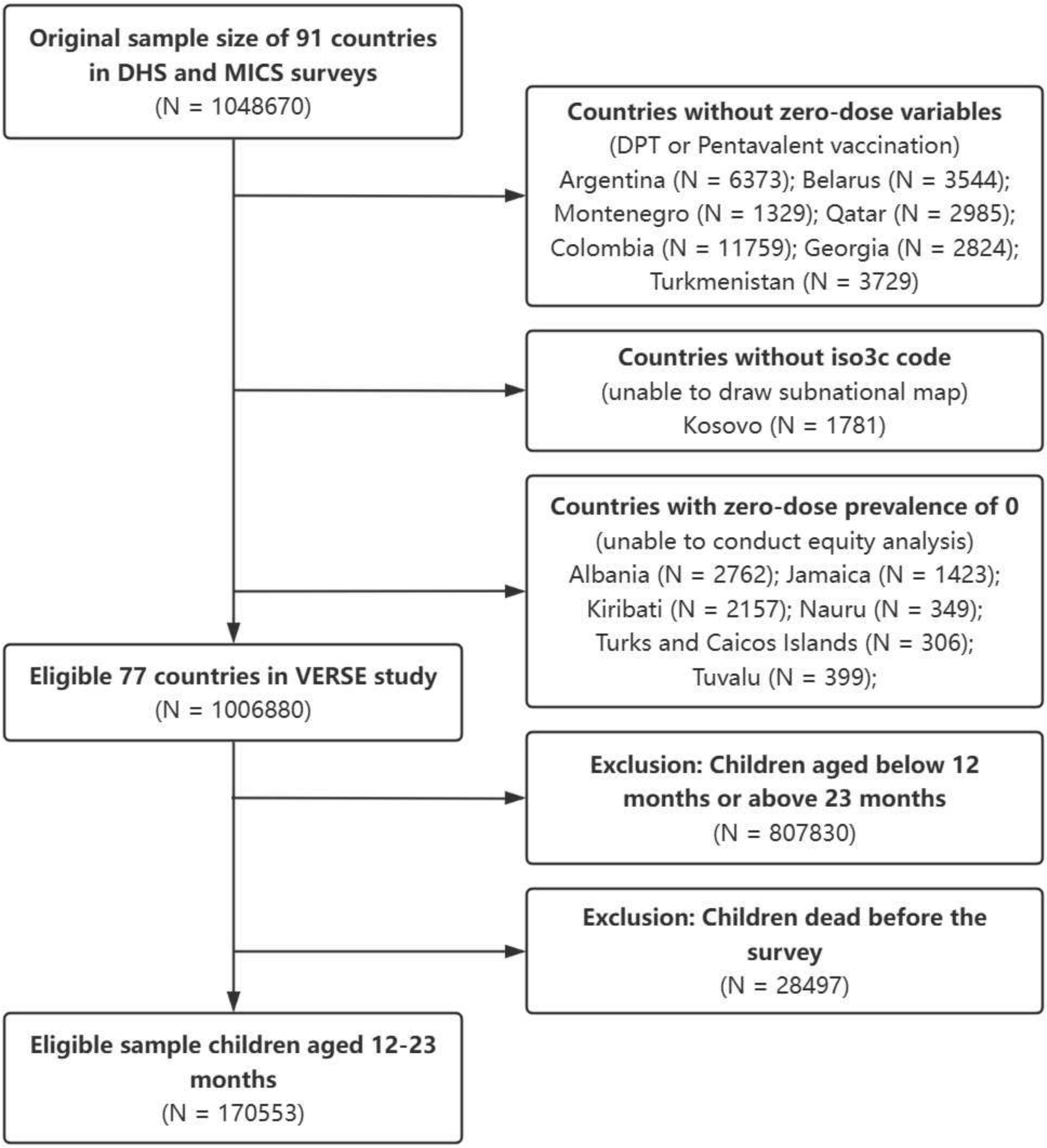

* The insurance variable was missing in some countries, where samples were analyzed with a corrected VERSE model.

**Supplementary Table 2. Harmonization of variables across DHS and MICS surveys**

| Variable | Definition | DHS | MICS |
|---|---|---|---|
| Sample weight | Sample weight of the child | V005 | chweight |
| Survival status | Whether child was alive or dead at the time of interview | B5 | NA |
| DTP1 | Received DTP 1 vaccination | H3 | IMDTP1* |
| DTP2 | Received DTP 2 vaccination | H5 | IMDTP2 |
| DTP3 | Received DTP 3 vaccination | H7 | IMDTP3 |
| PENTA1 | Received Pentavalent 1 vaccination | H51 | IM6PENTA1 |
| PENTA2 | Received Pentavalent 2 vaccination | H52 | IM6PENTA2 |
| PENTA3 | Received Pentavalent 3 vaccination | H53 | IM6PENTA3 |
| Age month | Current age of child in months | B19 | NA** |
| Sex | Sex of child | B4 | HL4 |
| Wealth index | Wealth index in quintiles | V190 | windex5 |
| Maternal education | Highest level of education the mother of child attended | V106 | melevel |
| Region | Region in which the respondent was interviewed | V101 | HH7 |
| Urban / Rural | Type of place of residence where the respondent was interviewed as either urban or rural | V025 | HH6 |
| Insurance | Whether child was covered by health insurance | V481 | UB9 |

* DPT vaccination data were only collected in a few countries of MICS, and the variable names were not uniform.

** The age of children in months in MICS is calculated by subtracting the time of birth from the survey period.

**Supplementary Table 3. National distribution of demographic characteristics across 77 countries, 2015 - 2024**

| Country | Sex | | Wealth Index | | | | |
|---|---|---|---|---|---|---|---|
| | Male | Female | Poorest | Poorer | Middle | Richer | Richest |
| Afghanistan | 1913 (51.5%) | 1799 (48.5%) | 625 (16.8%) | 760 (20.5%) | 894 (24.1%) | 795 (21.4%) | 637 (17.2%) |
| Algeria | 1325 (50.4%) | 1303 (49.6%) | 660 (25.1%) | 620 (23.6%) | 554 (21.1%) | 466 (17.7%) | 328 (12.5%) |
| Angola | 1417 (50.4%) | 1395 (49.6%) | 667 (23.7%) | 801 (28.5%) | 719 (25.6%) | 370 (13.2%) | 255 (9.1%) |
| Armenia | 182 (52.8%) | 163 (47.2%) | 70 (20.3%) | 69 (20%) | 77 (22.3%) | 64 (18.6%) | 65 (18.8%) |
| Azerbaijan | 37 (57.8%) | 27 (42.2%) | 16 (25%) | 15 (23.4%) | 15 (23.4%) | 13 (20.3%) | 5 (7.8%) |
| Bangladesh | 844 (50.7%) | 822 (49.3%) | 354 (21.2%) | 344 (20.6%) | 295 (17.7%) | 340 (20.4%) | 333 (20%) |
| Benin | 1152 (53.5%) | 1002 (46.5%) | 411 (19.1%) | 459 (21.3%) | 453 (21%) | 451 (20.9%) | 380 (17.6%) |
| Burkina Faso | 1175 (51.1%) | 1125 (48.9%) | 431 (18.7%) | 469 (20.4%) | 488 (21.2%) | 506 (22%) | 406 (17.7%) |
| Burundi | 1292 (49.8%) | 1303 (50.2%) | 519 (20%) | 553 (21.3%) | 549 (21.2%) | 470 (18.1%) | 504 (19.4%) |
| Cambodia | 859 (51.7%) | 804 (48.3%) | 488 (29.3%) | 308 (18.5%) | 294 (17.7%) | 325 (19.5%) | 248 (14.9%) |
| Cameroon | 933 (51.6%) | 876 (48.4%) | 317 (17.5%) | 443 (24.5%) | 448 (24.8%) | 338 (18.7%) | 263 (14.5%) |
| Central African Republic | 340 (47.4%) | 378 (52.6%) | 76 (10.6%) | 101 (14.1%) | 132 (18.4%) | 186 (25.9%) | 223 (31.1%) |
| Chad | 605 (48.2%) | 650 (51.8%) | 201 (16%) | 246 (19.6%) | 272 (21.7%) | 260 (20.7%) | 276 (22%) |
| Comoros | 331 (49.6%) | 337 (50.4%) | 136 (20.4%) | 138 (20.7%) | 141 (21.1%) | 134 (20.1%) | 119 (17.8%) |
| Costa Rica | 334 (49.9%) | 336 (50.1%) | 218 (32.5%) | 157 (23.4%) | 128 (19.1%) | 102 (15.2%) | 65 (9.7%) |
| Cuba | 554 (50.8%) | 536 (49.2%) | 202 (18.5%) | 192 (17.6%) | 219 (20.1%) | 241 (22.1%) | 236 (21.7%) |
| Côte d'Ivoire | 970 (50.8%) | 939 (49.2%) | 524 (27.4%) | 472 (24.7%) | 420 (22%) | 287 (15%) | 206 (10.8%) |
| Democratic Republic of the Congo | 2104 (50.3%) | 2080 (49.7%) | 1222 (29.2%) | 1077 (25.7%) | 907 (21.7%) | 652 (15.6%) | 326 (7.8%) |
| Dominican Republic | 732 (51%) | 704 (49%) | 466 (32.5%) | 309 (21.5%) | 273 (19%) | 225 (15.7%) | 163 (11.4%) |
| Eswatini | 195 (49.9%) | 196 (50.1%) | 101 (25.8%) | 93 (23.8%) | 76 (19.4%) | 64 (16.4%) | 57 (14.6%) |
| Ethiopia | 502 (50.5%) | 492 (49.5%) | 297 (29.9%) | 163 (16.4%) | 133 (13.4%) | 133 (13.4%) | 268 (27%) |

| Country | Sex | | Wealth Index | | | | |
|---|---|---|---|---|---|---|---|
| | Male | Female | Poorest | Poorer | Middle | Richer | Richest |
| Fiji | 190 (49.9%) | 191 (50.1%) | 103 (27%) | 92 (24.1%) | 65 (17.1%) | 65 (17.1%) | 56 (14.7%) |
| Gabon | 632 (50.6%) | 617 (49.4%) | 526 (42.1%) | 270 (21.6%) | 191 (15.3%) | 145 (11.6%) | 117 (9.4%) |
| The Gambia | 811 (51.3%) | 769 (48.7%) | 549 (34.7%) | 335 (21.2%) | 323 (20.4%) | 212 (13.4%) | 161 (10.2%) |
| Ghana | 1007 (51.1%) | 963 (48.9%) | 641 (32.5%) | 487 (24.7%) | 354 (18%) | 266 (13.5%) | 222 (11.3%) |
| Guatemala | 1202 (49.9%) | 1205 (50.1%) | 631 (26.2%) | 566 (23.5%) | 504 (20.9%) | 422 (17.5%) | 284 (11.8%) |
| Guinea | 729 (52.2%) | 668 (47.8%) | 330 (23.6%) | 317 (22.7%) | 260 (18.6%) | 260 (18.6%) | 230 (16.5%) |
| Guinea-Bissau | 634 (48.5%) | 674 (51.5%) | 399 (30.5%) | 303 (23.2%) | 267 (20.4%) | 220 (16.8%) | 119 (9.1%) |
| Guyana | 245 (49.9%) | 246 (50.1%) | 222 (45.2%) | 92 (18.7%) | 61 (12.4%) | 62 (12.6%) | 54 (11%) |
| Haiti | 604 (50.5%) | 591 (49.5%) | 392 (32.8%) | 259 (21.7%) | 233 (19.5%) | 175 (14.6%) | 136 (11.4%) |
| Honduras | 820 (50.8%) | 795 (49.2%) | 516 (32%) | 344 (21.3%) | 306 (18.9%) | 291 (18%) | 158 (9.8%) |
| India | 22492 (51.9%) | 20844 (48.1%) | 11350 (26.2%) | 10047 (23.2%) | 8505 (19.6%) | 7432 (17.1%) | 6002 (13.8%) |
| Indonesia | 1812 (51.6%) | 1699 (48.4%) | 956 (27.2%) | 683 (19.5%) | 635 (18.1%) | 646 (18.4%) | 591 (16.8%) |
| Iraq | 1381 (51.6%) | 1296 (48.4%) | 681 (25.4%) | 623 (23.3%) | 519 (19.4%) | 456 (17%) | 398 (14.9%) |
| Jordan | 887 (54%) | 757 (46%) | 634 (38.6%) | 391 (23.8%) | 329 (20%) | 221 (13.4%) | 69 (4.2%) |
| Kenya | 1874 (51%) | 1798 (49%) | 1215 (33.1%) | 661 (18%) | 631 (17.2%) | 664 (18.1%) | 501 (13.6%) |
| Kyrgyzstan | 197 (53.7%) | 170 (46.3%) | 86 (23.4%) | 61 (16.6%) | 76 (20.7%) | 84 (22.9%) | 60 (16.3%) |
| Laos | 711 (51.6%) | 668 (48.4%) | 372 (27%) | 326 (23.6%) | 246 (17.8%) | 242 (17.5%) | 193 (14%) |
| Lesotho | 254 (48.6%) | 269 (51.4%) | 179 (34.2%) | 108 (20.7%) | 95 (18.2%) | 83 (15.9%) | 58 (11.1%) |
| Liberia | 518 (48.9%) | 542 (51.1%) | 379 (35.8%) | 291 (27.5%) | 198 (18.7%) | 117 (11%) | 75 (7.1%) |
| Madagascar | 1210 (51.7%) | 1130 (48.3%) | 666 (28.5%) | 518 (22.1%) | 443 (18.9%) | 359 (15.3%) | 354 (15.1%) |
| Malawi | 1464 (49.4%) | 1497 (50.6%) | 744 (25.1%) | 607 (20.5%) | 570 (19.3%) | 566 (19.1%) | 474 (16%) |
| Mali | 1417 (49.8%) | 1427 (50.2%) | 518 (18.2%) | 577 (20.3%) | 684 (24.1%) | 588 (20.7%) | 477 (16.8%) |
| Mauritania | 1095 (52.2%) | 1002 (47.8%) | 417 (19.9%) | 439 (20.9%) | 518 (24.7%) | 385 (18.4%) | 338 (16.1%) |

| Country | Sex | | Wealth Index | | | | |
|---|---|---|---|---|---|---|---|
| | Male | Female | Poorest | Poorer | Middle | Richer | Richest |
| Mongolia | 466 (50.9%) | 450 (49.1%) | 272 (29.7%) | 234 (25.5%) | 175 (19.1%) | 134 (14.6%) | 101 (11%) |
| Mozambique | 837 (49.9%) | 841 (50.1%) | 360 (21.5%) | 309 (18.4%) | 342 (20.4%) | 380 (22.6%) | 287 (17.1%) |
| Myanmar | 503 (55%) | 411 (45%) | 262 (28.7%) | 206 (22.5%) | 160 (17.5%) | 166 (18.2%) | 120 (13.1%) |
| Nepal | 515 (51.7%) | 481 (48.3%) | 344 (34.5%) | 223 (22.4%) | 173 (17.4%) | 167 (16.8%) | 89 (8.9%) |
| Nigeria | 2509 (51.1%) | 2405 (48.9%) | 1248 (25.4%) | 976 (19.9%) | 903 (18.4%) | 959 (19.5%) | 828 (16.8%) |
| North Macedonia | 144 (50.5%) | 141 (49.5%) | 67 (23.5%) | 59 (20.7%) | 56 (19.6%) | 53 (18.6%) | 50 (17.5%) |
| Pakistan | 7044 (52.2%) | 6451 (47.8%) | 2661 (19.7%) | 2886 (21.4%) | 2910 (21.6%) | 2752 (20.4%) | 2277 (16.9%) |
| Papua New Guinea | 938 (52.6%) | 845 (47.4%) | 273 (15.3%) | 286 (16%) | 323 (18.1%) | 452 (25.4%) | 449 (25.2%) |
| Philippines | 827 (52.7%) | 741 (47.3%) | 601 (38.3%) | 328 (20.9%) | 265 (16.9%) | 187 (11.9%) | 187 (11.9%) |
| Rwanda | 810 (51.5%) | 762 (48.5%) | 362 (23%) | 338 (21.5%) | 306 (19.5%) | 310 (19.7%) | 256 (16.3%) |
| São Tomé and Príncipe | 182 (55.2%) | 148 (44.8%) | 78 (23.6%) | 72 (21.8%) | 52 (15.8%) | 69 (20.9%) | 59 (17.9%) |
| Senegal | 1047 (51.2%) | 997 (48.8%) | 646 (31.6%) | 530 (25.9%) | 385 (18.8%) | 282 (13.8%) | 201 (9.8%) |
| Serbia | 170 (51.8%) | 158 (48.2%) | 37 (11.3%) | 45 (13.7%) | 66 (20.1%) | 82 (25%) | 98 (29.9%) |
| Sierra Leone | 932 (50.3%) | 920 (49.7%) | 479 (25.9%) | 423 (22.8%) | 410 (22.1%) | 311 (16.8%) | 229 (12.4%) |
| South Africa | 344 (53.5%) | 299 (46.5%) | 179 (27.8%) | 161 (25%) | 134 (20.8%) | 106 (16.5%) | 63 (9.8%) |
| Palestine | 664 (52.2%) | 608 (47.8%) | 239 (18.8%) | 190 (14.9%) | 295 (23.2%) | 321 (25.2%) | 227 (17.8%) |
| Suriname | 344 (56.1%) | 269 (43.9%) | 159 (25.9%) | 151 (24.6%) | 123 (20.1%) | 113 (18.4%) | 67 (10.9%) |
| Tajikistan | 494 (52.2%) | 452 (47.8%) | 161 (17%) | 156 (16.5%) | 169 (17.9%) | 178 (18.8%) | 282 (29.8%) |
| Tanzania | 1104 (51.8%) | 1029 (48.2%) | 471 (22.1%) | 433 (20.3%) | 414 (19.4%) | 415 (19.5%) | 400 (18.8%) |
| Thailand | 956 (52.2%) | 876 (47.8%) | 412 (22.5%) | 388 (21.2%) | 430 (23.5%) | 344 (18.8%) | 258 (14.1%) |
| Timor-Leste | 725 (51.1%) | 693 (48.9%) | 257 (18.1%) | 305 (21.5%) | 284 (20%) | 334 (23.6%) | 238 (16.8%) |
| Togo | 455 (51.5%) | 428 (48.5%) | 183 (20.7%) | 208 (23.6%) | 179 (20.3%) | 162 (18.3%) | 151 (17.1%) |
| Tonga | 74 (48.1%) | 80 (51.9%) | 58 (37.7%) | 28 (18.2%) | 25 (16.2%) | 21 (13.6%) | 22 (14.3%) |

| Country | Sex | | Wealth Index | | | | |
|---|---|---|---|---|---|---|---|
| | Male | Female | Poorest | Poorer | Middle | Richer | Richest |
| Trinidad and Tobago | 81 (46.6%) | 93 (53.4%) | 41 (23.6%) | 41 (23.6%) | 39 (22.4%) | 31 (17.8%) | 22 (12.6%) |
| Tunisia | 123 (47.1%) | 138 (52.9%) | 71 (27.2%) | 76 (29.1%) | 61 (23.4%) | 36 (13.8%) | 17 (6.5%) |
| Turkey | 217 (48.5%) | 230 (51.5%) | 116 (26%) | 98 (21.9%) | 82 (18.3%) | 73 (16.3%) | 78 (17.4%) |
| Uganda | 1490 (51.2%) | 1420 (48.8%) | 757 (26%) | 649 (22.3%) | 526 (18.1%) | 484 (16.6%) | 494 (17%) |
| Uzbekistan | 524 (52.1%) | 481 (47.9%) | 201 (20%) | 210 (20.9%) | 200 (19.9%) | 216 (21.5%) | 178 (17.7%) |
| Vanuatu | 179 (57%) | 135 (43%) | 74 (23.6%) | 77 (24.5%) | 63 (20.1%) | 62 (19.7%) | 38 (12.1%) |
| Vietnam | 363 (53.3%) | 318 (46.7%) | 231 (33.9%) | 127 (18.6%) | 108 (15.9%) | 103 (15.1%) | 112 (16.4%) |
| Yemen | 1101 (51.7%) | 1028 (48.3%) | 458 (21.5%) | 413 (19.4%) | 410 (19.3%) | 433 (20.3%) | 415 (19.5%) |
| Zambia | 958 (49.8%) | 965 (50.2%) | 549 (28.5%) | 463 (24.1%) | 375 (19.5%) | 277 (14.4%) | 259 (13.5%) |
| Zimbabwe | 546 (50.9%) | 527 (49.1%) | 267 (24.9%) | 216 (20.1%) | 191 (17.8%) | 215 (20%) | 184 (17.1%) |

| Country | Maternal Education | | | | Residence | | Insurance | |
|---|---|---|---|---|---|---|---|---|
| | No education | Primary | Secondary | Higher | Urban | Rural | With | Without |
| Afghanistan | 2773 (74.7%) | 421 (11.3%) | 387 (10.4%) | 131 (3.5%) | 684 (18.4%) | 3028 (81.6%) | | |
| Algeria | 360 (13.7%) | 405 (15.4%) | 1445 (55%) | 418 (15.9%) | 1587 (60.4%) | 1041 (39.6%) | 1205 (45.9%) | 1419 (54%) |
| Angola | 944 (33.6%) | 1045 (37.2%) | 777 (27.6%) | 46 (1.6%) | 1572 (55.9%) | 1240 (44.1%) | 130 (4.6%) | 2682 (95.4%) |
| Armenia | 24 (7%) | 145 (42%) | 176 (51%) | | 195 (56.5%) | 150 (43.5%) | 26 (7.5%) | 319 (92.5%) |
| Azerbaijan | 2 (3.1%) | 50 (78.1%) | 12 (18.8%) | | 17 (26.6%) | 47 (73.4%) | | 64 (100%) |
| Bangladesh | 100 (6%) | 470 (28.2%) | 787 (47.2%) | 309 (18.5%) | 569 (34.2%) | 1097 (65.8%) | 5 (0.3%) | 1661 (99.7%) |
| Benin | 1030 (47.8%) | 506 (23.5%) | 453 (21%) | 165 (7.7%) | 949 (44.1%) | 1205 (55.9%) | 28 (1.3%) | 2125 (98.7%) |
| Burkina Faso | 1558 (67.7%) | 302 (13.1%) | 412 (17.9%) | 28 (1.2%) | 665 (28.9%) | 1635 (71.1%) | 11 (0.5%) | 2289 (99.5%) |
| Burundi | 1055 (40.7%) | 1198 (46.2%) | 323 (12.4%) | 19 (0.7%) | 392 (15.1%) | 2203 (84.9%) | 632 (24.4%) | 1963 (75.6%) |

| Country | Maternal Education | | | | Residence | | Insurance | |
|---|---|---|---|---|---|---|---|---|
| | No education | Primary | Secondary | Higher | Urban | Rural | With | Without |
| Cambodia | 214 (12.9%) | 681 (41%) | 678 (40.8%) | 90 (5.4%) | 583 (35.1%) | 1080 (64.9%) | 365 (21.9%) | 1298 (78.1%) |
| Cameroon | 399 (22.1%) | 592 (32.7%) | 713 (39.4%) | 105 (5.8%) | 784 (43.3%) | 1025 (56.7%) | 33 (1.8%) | 1776 (98.2%) |
| Central African Republic | 192 (26.7%) | 307 (42.8%) | 142 (19.8%) | 77 (10.7%) | 374 (52.1%) | 344 (47.9%) | 11 (1.5%) | 707 (98.5%) |
| Chad | 723 (57.6%) | 301 (24%) | 210 (16.7%) | 21 (1.7%) | 279 (22.2%) | 976 (77.8%) | 6 (0.5%) | 1247 (99.4%) |
| Comoros | 127 (19%) | 154 (23.1%) | 241 (36.1%) | 146 (21.9%) | 252 (37.7%) | 416 (62.3%) | 43 (6.4%) | 625 (93.6%) |
| Costa Rica | 8 (1.2%) | 146 (21.8%) | 342 (51%) | 174 (26%) | 384 (57.3%) | 286 (42.7%) | 613 (91.5%) | 57 (8.5%) |
| Cuba | 12 (1.1%) | 793 (72.8%) | 285 (26.1%) | | 785 (72%) | 305 (28%) | | |
| Côte d'Ivoire | 1231 (64.5%) | 343 (18%) | 270 (14.1%) | 65 (3.4%) | 766 (40.1%) | 1143 (59.9%) | 94 (4.9%) | 1815 (95.1%) |
| Democratic Republic of the Congo | 870 (20.8%) | 1326 (31.7%) | 1905 (45.5%) | 83 (2%) | 1076 (25.7%) | 3108 (74.3%) | 84 (2%) | 4100 (98%) |
| Dominican Republic | 44 (3.1%) | 300 (20.9%) | 577 (40.2%) | 511 (35.6%) | 979 (68.2%) | 457 (31.8%) | 689 (48%) | 746 (51.9%) |
| Eswatini | 26 (6.6%) | 90 (23%) | 234 (59.8%) | 40 (10.2%) | 53 (13.6%) | 338 (86.4%) | | |
| Ethiopia | 483 (48.6%) | 339 (34.1%) | 98 (9.9%) | 74 (7.4%) | 266 (26.8%) | 728 (73.2%) | | |
| Fiji | 30 (7.9%) | 198 (52%) | 153 (40.2%) | | 193 (50.7%) | 188 (49.3%) | 24 (6.3%) | 357 (93.7%) |
| Gabon | 82 (6.6%) | 272 (21.8%) | 799 (64%) | 96 (7.7%) | 867 (69.4%) | 382 (30.6%) | 614 (49.2%) | 202 (16.2%) |
| The Gambia | 788 (49.9%) | 322 (20.4%) | 425 (26.9%) | 45 (2.8%) | 707 (44.7%) | 873 (55.3%) | 28 (1.8%) | 1552 (98.2%) |
| Ghana | 567 (28.8%) | 317 (16.1%) | 944 (47.9%) | 142 (7.2%) | 826 (41.9%) | 1144 (58.1%) | 1848 (93.8%) | 122 (6.2%) |
| Guatemala | 402 (16.7%) | 1282 (53.3%) | 638 (26.5%) | 85 (3.5%) | 807 (33.5%) | 1600 (66.5%) | 247 (10.3%) | 2159 (89.7%) |
| Guinea | 1028 (73.6%) | 163 (11.7%) | 172 (12.3%) | 34 (2.4%) | 407 (29.1%) | 990 (70.9%) | 13 (0.9%) | 1384 (99.1%) |
| Guinea-Bissau | 647 (49.5%) | 572 (43.7%) | 79 (6%) | 8 (0.6%) | 278 (21.3%) | 1030 (78.7%) | 12 (0.9%) | 1296 (99.1%) |
| Guyana | 15 (3.1%) | 40 (8.1%) | 368 (74.9%) | 66 (13.4%) | 125 (25.5%) | 366 (74.5%) | 25 (5.1%) | 466 (94.9%) |
| Haiti | 233 (19.5%) | 484 (40.5%) | 447 (37.4%) | 31 (2.6%) | 352 (29.5%) | 843 (70.5%) | 16 (1.3%) | 1179 (98.7%) |

| Country | Maternal Education | | | | Residence | | Insurance | |
|---|---|---|---|---|---|---|---|---|
| | No education | Primary | Secondary | Higher | Urban | Rural | With | Without |
| Honduras | 61 (3.8%) | 1099 (68%) | 348 (21.5%) | 107 (6.6%) | 575 (35.6%) | 1040 (64.4%) | | |
| India | 8447 (19.5%) | 5158 (11.9%) | 23158 (53.4%) | 6573 (15.2%) | 8858 (20.4%) | 34478 (79.6%) | 11294 (26.1%) | 32042 (73.9%) |
| Indonesia | 38 (1.1%) | 801 (22.8%) | 1988 (56.6%) | 684 (19.5%) | 1738 (49.5%) | 1773 (50.5%) | 2218 (63.2%) | 1293 (36.8%) |
| Iraq | 513 (19.2%) | 1168 (43.6%) | 463 (17.3%) | 533 (19.9%) | 1738 (64.9%) | 939 (35.1%) | 17 (0.6%) | 2657 (99.3%) |
| Jordan | 44 (2.7%) | 132 (8%) | 958 (58.3%) | 510 (31%) | 1316 (80%) | 328 (20%) | 1325 (80.6%) | 319 (19.4%) |
| Kenya | 796 (21.7%) | 1274 (34.7%) | 1135 (30.9%) | 467 (12.7%) | 1229 (33.5%) | 2443 (66.5%) | | |
| Kyrgyzstan | 2 (0.5%) | 259 (70.6%) | 106 (28.9%) | | 133 (36.2%) | 234 (63.8%) | | |
| Laos | 150 (10.9%) | 400 (29%) | 656 (47.6%) | 171 (12.4%) | 361 (26.2%) | 1018 (73.8%) | 445 (32.3%) | 934 (67.7%) |
| Lesotho | 4 (0.8%) | 178 (34%) | 302 (57.7%) | 39 (7.5%) | 154 (29.4%) | 369 (70.6%) | 9 (1.7%) | 514 (98.3%) |
| Liberia | 427 (40.3%) | 344 (32.5%) | 276 (26%) | 13 (1.2%) | 345 (32.5%) | 715 (67.5%) | 30 (2.8%) | 1030 (97.2%) |
| Madagascar | 551 (23.5%) | 1026 (43.8%) | 694 (29.7%) | 69 (2.9%) | 473 (20.2%) | 1867 (79.8%) | 52 (2.2%) | 2288 (97.8%) |
| Malawi | 251 (8.5%) | 2010 (67.9%) | 660 (22.3%) | 38 (1.3%) | 378 (12.8%) | 2583 (87.2%) | 11 (0.4%) | 2948 (99.6%) |
| Mali | 1715 (60.3%) | 475 (16.7%) | 593 (20.9%) | 61 (2.1%) | 705 (24.8%) | 2139 (75.2%) | 156 (5.5%) | 2688 (94.5%) |
| Mauritania | 768 (36.6%) | 888 (42.3%) | 402 (19.2%) | 39 (1.9%) | 889 (42.4%) | 1208 (57.6%) | 175 (8.3%) | 1922 (91.7%) |
| Mongolia | 41 (4.5%) | 54 (5.9%) | 463 (50.5%) | 358 (39.1%) | 478 (52.2%) | 438 (47.8%) | | |
| Mozambique | 455 (27.1%) | 779 (46.4%) | 412 (24.6%) | 32 (1.9%) | 518 (30.9%) | 1160 (69.1%) | 16 (1%) | 1662 (99%) |
| Myanmar | 130 (14.2%) | 405 (44.3%) | 310 (33.9%) | 69 (7.5%) | 209 (22.9%) | 705 (77.1%) | 6 (0.7%) | 908 (99.3%) |
| Nepal | 197 (19.8%) | 360 (36.1%) | 408 (41%) | 31 (3.1%) | 510 (51.2%) | 486 (48.8%) | 79 (7.9%) | 917 (92.1%) |
| Nigeria | 2067 (42.1%) | 541 (11%) | 1740 (35.4%) | 566 (11.5%) | 1950 (39.7%) | 2964 (60.3%) | 129 (2.6%) | 4785 (97.4%) |
| North Macedonia | 60 (21.1%) | 112 (39.3%) | 113 (39.6%) | | 147 (51.6%) | 138 (48.4%) | 276 (96.8%) | 9 (3.2%) |
| Pakistan | 6993 (51.8%) | 2188 (16.2%) | 2718 (20.1%) | 1586 (11.8%) | 3692 (27.4%) | 9803 (72.6%) | 259 (1.9%) | 13210 (97.9%) |
| Papua New Guinea | 342 (19.2%) | 900 (50.5%) | 467 (26.2%) | 74 (4.2%) | 410 (23%) | 1373 (77%) | 50 (2.8%) | 1716 (96.2%) |

| Country | Maternal Education | | | | Residence | | Insurance | |
|---|---|---|---|---|---|---|---|---|
| | No education | Primary | Secondary | Higher | Urban | Rural | With | Without |
| Philippines | 27 (1.7%) | 219 (14%) | 764 (48.7%) | 558 (35.6%) | 531 (33.9%) | 1037 (66.1%) | | |
| Rwanda | 151 (9.6%) | 1027 (65.3%) | 319 (20.3%) | 75 (4.8%) | 304 (19.3%) | 1268 (80.7%) | 1247 (79.3%) | 325 (20.7%) |
| São Tomé and Príncipe | 7 (2.1%) | 149 (45.2%) | 163 (49.4%) | 10 (3%) | 182 (55.2%) | 148 (44.8%) | 4 (1.2%) | 324 (98.2%) |
| Senegal | 1121 (54.8%) | 392 (19.2%) | 480 (23.5%) | 51 (2.5%) | 742 (36.3%) | 1302 (63.7%) | 163 (8%) | 1881 (92%) |
| Serbia | 18 (5.5%) | 140 (42.7%) | 170 (51.8%) | | 210 (64%) | 118 (36%) | 326 (99.4%) | 2 (0.6%) |
| Sierra Leone | 1027 (55.5%) | 276 (14.9%) | 491 (26.5%) | 58 (3.1%) | 549 (29.6%) | 1303 (70.4%) | 68 (3.7%) | 1784 (96.3%) |
| South Africa | 5 (0.8%) | 58 (9%) | 509 (79.2%) | 71 (11%) | 330 (51.3%) | 313 (48.7%) | 26 (4%) | 289 (44.9%) |
| Palestine | 240 (18.9%) | 438 (34.4%) | 594 (46.7%) | | 743 (58.4%) | 326 (25.6%) | 860 (67.6%) | 412 (32.4%) |
| Suriname | 22 (3.6%) | 101 (16.5%) | 405 (66.1%) | 74 (12.1%) | 339 (55.3%) | 274 (44.7%) | 563 (91.8%) | 50 (8.2%) |
| Tajikistan | 13 (1.4%) | 31 (3.3%) | 640 (67.7%) | 262 (27.7%) | 387 (40.9%) | 559 (59.1%) | | |
| Tanzania | 453 (21.2%) | 1077 (50.5%) | 583 (27.3%) | 20 (0.9%) | 586 (27.5%) | 1547 (72.5%) | 85 (4%) | 2048 (96%) |
| Thailand | 58 (3.2%) | 402 (21.9%) | 841 (45.9%) | 530 (28.9%) | 792 (43.2%) | 1040 (56.8%) | 1785 (97.4%) | 47 (2.6%) |
| Timor-Leste | 331 (23.3%) | 255 (18%) | 711 (50.1%) | 121 (8.5%) | 410 (28.9%) | 1008 (71.1%) | | |
| Togo | 285 (32.3%) | 319 (36.1%) | 279 (31.6%) | | 299 (33.9%) | 584 (66.1%) | 39 (4.4%) | 843 (95.5%) |
| Tonga | 4 (2.6%) | 150 (97.4%) | | | 33 (21.4%) | 121 (78.6%) | 131 (85.1%) | 23 (14.9%) |
| Trinidad and Tobago | 4 (2.3%) | 129 (74.1%) | 39 (22.4%) | | 90 (51.7%) | 84 (48.3%) | 22 (12.6%) | 152 (87.4%) |
| Tunisia | 11 (4.2%) | 48 (18.4%) | 142 (54.4%) | 60 (23%) | 127 (48.7%) | 134 (51.3%) | 196 (75.1%) | 65 (24.9%) |
| Turkey | 49 (11%) | 114 (25.5%) | 200 (44.7%) | 84 (18.8%) | 316 (70.7%) | 131 (29.3%) | 392 (87.7%) | 55 (12.3%) |
| Uganda | 334 (11.5%) | 1812 (62.3%) | 578 (19.9%) | 186 (6.4%) | 572 (19.7%) | 2338 (80.3%) | 37 (1.3%) | 2873 (98.7%) |
| Uzbekistan | 918 (91.3%) | 87 (8.7%) | | | 476 (47.4%) | 529 (52.6%) | | |
| Vanuatu | 123 (39.2%) | 179 (57%) | 12 (3.8%) | | 58 (18.5%) | 256 (81.5%) | | 314 (100%) |
| Vietnam | 60 (8.8%) | 83 (12.2%) | 370 (54.3%) | 168 (24.7%) | 201 (29.5%) | 480 (70.5%) | 662 (97.2%) | 18 (2.6%) |

| Country | Maternal Education | | | | Residence | | Insurance | |
|---|---|---|---|---|---|---|---|---|
| | No education | Primary | Secondary | Higher | Urban | Rural | With | Without |
| Yemen | 751 (35.3%) | 540 (25.4%) | 739 (34.7%) | 96 (4.5%) | 603 (28.3%) | 1526 (71.7%) | 5 (0.2%) | 2123 (99.7%) |
| Zambia | 199 (10.3%) | 1009 (52.5%) | 633 (32.9%) | 82 (4.3%) | 587 (30.5%) | 1336 (69.5%) | 36 (1.9%) | 1887 (98.1%) |
| Zimbabwe | 15 (1.4%) | 293 (27.3%) | 686 (63.9%) | 79 (7.4%) | 332 (30.9%) | 741 (69.1%) | 53 (4.9%) | 1020 (95.1%) |

#### B: Model details of VERSE Framework

##### Conceptual framework

The Vaccine Economics Research for Sustainability and Equity (VERSE) framework was applied to assess multidimensional inequities in zero-dose prevalence across countries. The VERSE framework provides a standardised approach for measuring multivariate equity in vaccination and health outcomes by integrating multiple social, demographic and geographic determinants into a unified ranking of unfair disadvantage. Unlike conventional inequality analyses that rank individuals according to a single socioeconomic indicator, such as household wealth or maternal education alone, the VERSE framework estimates each child's position in a multidimensional distribution of disadvantage and then evaluates how the outcome is distributed along that ranking.

In this study, the outcome was zero-dose status, defined as non-receipt of any DTP-containing vaccine among surviving children aged 12–23 months. Because zero-dose status represents an adverse immunisation outcome, higher predicted probabilities of zero-dose status were interpreted as indicating greater multidimensional disadvantage. The resulting VERSE ranking was used to estimate relative and absolute inequities in zero-dose prevalence within each country.

##### Selection and harmonisation of equity stratifiers

Equity stratifiers were selected a priori based on previous applications of the VERSE methodology, the literature on unfair inequalities in health-care access, and the availability of comparable variables across DHS and MICS surveys. The core equity dimensions included in this study were: subnational region, place of residence, maternal education, household wealth quintile, child sex and health insurance coverage.

Maternal age and birth order were not included in the final VERSE specification because they were not part of the harmonised core equity dimensions used in the main analysis. Health insurance was included when comparable information was available. Because health insurance was structurally unavailable in some MICS surveys, countries without a comparable insurance variable were analysed using a modified VERSE specification that omitted health insurance while retaining the remaining equity dimensions.

Variables derived from DHS and MICS datasets were harmonised before model estimation. Maternal education was recoded into common educational attainment categories, household wealth was harmonised using survey-specific wealth quintiles, place of residence was classified as urban or rural, and subnational regions were harmonised according to survey-specific administrative classifications and the ADM1 mapping framework used for the main analysis.

**Estimation of multidimensional disadvantage**

For each country, we fitted a survey-weighted logistic regression model with zero-dose status as the binary dependent variable. The general direct unfairness score can be conceptualised as:

$$d_i = E(Y_i | N_i = N^{ref}, P_i = P^{ref}, Z_i, X_i = X^{ref})$$

where $N_i$ denotes legitimate need-related factors, $P_i$ denotes preference-related factors, $Z_i$ denotes unfair social, demographic and geographic determinants, and $X_i$ denotes other covariates. In this analysis, all children were restricted to the standard assessment age of 12–23 months, and the outcome was zero-dose status rather than receipt of a vaccine scheduled at different ages. Therefore, legitimate need was largely standardised by design, and the operational model focused on the selected unfair determinants:

By deriving disadvantage rankings from the joint distribution of multiple social determinants, the VERSE framework captures the cumulative effects of intersecting vulnerabilities that may not be adequately identified using unidimensional socioeconomic indicators alone.

$$d_i = E(Y_i | Z_i)$$

For each country, the survey-weighted logistic regression model was specified as:

$$\text{logit}\{Pr(Y_i = 1)\} = \alpha + \sum_{k=1}^{K} \beta_k Z_{ik}$$

where $Pr(Y_i = 1)$ is the probability that child iii was zero-dose, $Z_{ik}$ represents the *k*-th equity stratifier, and $\beta_k$ represents the corresponding regression coefficient.

The predicted probability from the fitted model was calculated as:

$$\widehat{d}_i = \hat{p}_i = \frac{\exp\left(\widehat{\alpha} + \sum_{k=1}^{K} \widehat{\beta}_k Z_{ik}\right)}{1 + \exp\left(\widehat{\alpha} + \sum_{k=1}^{K} \widehat{\beta}_k Z_{ik}\right)}$$

where $\widehat{d}_i$ represents the individual-level multidimensional disadvantage score. Children were then ranked within each country from the lowest to the highest predicted probability of zero-dose status. The weighted fractional rank was denoted as:

$$R_i = F(\widehat{d}_i)$$

A higher value of $R_i$ indicated greater estimated multidimensional disadvantage. This ranking captures the joint distribution of multiple social and geographic disadvantages and differs from conventional equity analyses based on a single stratifier, such as wealth quintile alone.

**Measurement of multidimensional inequity**

Relative multidimensional inequity in zero-dose prevalence was measured using a Wagstaff concentration index calculated over the VERSE disadvantage ranking. For each country, the concentration index was defined as:

$$CI = \frac{2}{\mu} Cov_w(Y_i, R_i)$$

where $Y_i$ denotes zero-dose status, $R_i$ denotes the weighted fractional rank of child *i* in the multidimensional disadvantage distribution, $\mu$ is the weighted mean zero-dose prevalence, and $Cov_w$ denotes the survey-weighted covariance.

Equivalently, the weighted concentration index can be written as:

$$CI = \frac{2}{\mu}\left[\frac{\sum_i w_i Y_i R_i}{\sum_i w_i} - \left(\frac{\sum_i w_i Y_i}{\sum_i w_i}\right)\left(\frac{\sum_i w_i R_i}{\sum_i w_i}\right)\right]$$

where $w_i$ represents the survey sampling weight. Because children were ranked from the least to the most disadvantaged and the outcome was adverse zero-dose status, positive concentration index values indicate that zero-dose children were disproportionately concentrated among more disadvantaged children. Values close to zero indicate a more equal distribution of zero-dose status across the multidimensional disadvantage ranking. Negative values, when observed, indicate that zero-dose status was more concentrated among less disadvantaged children or that the local distribution deviated from the national disadvantage gradient. At subnational levels, negative or extreme CI values should be interpreted cautiously because they can be influenced by small samples and low numbers of zero-dose children.

Absolute multidimensional inequity was measured using the Absolute Equity Gap (AEG), defined as the difference in zero-dose prevalence between the most disadvantaged and least disadvantaged quintiles of the VERSE ranking:

**Decomposition analysis**

To identify the main contributors to multidimensional inequity in zero-dose prevalence, we decomposed the concentration index using a regression-based concentration-index decomposition approach. This approach partitions the observed concentration index into the contributions attributable to each included equity dimension and a residual component.

For each country, the concentration index of zero-dose status was expressed as:

$$CI = \sum_{m=1}^{M} \left( \frac{\beta_m \bar{X}_m}{\mu} \right) CI_m + \frac{GC_\varepsilon}{\mu}$$

where $\bar{X}_m$ denotes the mmm-th equity determinant, $\beta_m$ is the corresponding regression coefficient or marginal effect, $\bar{X}_m$ is the weighted mean of $X_m$, μ is the weighted mean zero-dose prevalence, $CI_m$ is the concentration index of $X_m$ over the VERSE disadvantage ranking, and $GC_\varepsilon$ is the generalised concentration index of the residual term.

The contribution of each determinant depends on two components: the strength of association between the determinant and zero-dose status, and the extent to which the determinant itself is unequally distributed across the multidimensional disadvantage ranking. Percentage contributions were obtained by dividing each determinant-specific contribution by the overall concentration index. The residual component represents the share of multidimensional inequity not explained by the included equity dimensions. Decomposition analyses were conducted separately for each country, allowing the dominant contributors to zero-dose inequity to vary across settings.

**Survey design and weighting**

All analyses accounted for the complex sampling design of DHS and MICS surveys, including sampling weights, stratification and clustering. Survey weights provided within each dataset were applied to generate nationally representative estimates. Country-specific models were estimated independently to avoid imposing a common coefficient structure across heterogeneous national contexts. Estimates were reported as country-level concentration indices, absolute equity gaps and percentage contributions of each equity dimension. Subnational estimates were used to describe geographic heterogeneity and were interpreted cautiously when based on small local samples.

**Interpretation of the VERSE framework**

In this study, the VERSE framework should be interpreted as a multidimensional equity assessment tool rather than a causal model. The predicted probability from the VERSE model was used to rank children according to estimated multidimensional disadvantage, not to infer that any single determinant caused zero-dose status. This ranking allowed zero-dose prevalence to be evaluated across the joint distribution of multiple disadvantages, including socioeconomic, demographic and geographic dimensions.

Positive national concentration indices indicated that zero-dose children were concentrated among children with higher estimated multidimensional disadvantage. A large AEG indicated a large absolute difference in zero-dose prevalence between the most and least disadvantaged quintiles. The decomposition results identified which measured equity dimensions contributed most to the observed concentration of zero-dose status, while the residual component captured the portion of inequity not explained by the included variables.

Together, these metrics provide complementary information for interpreting zero-dose inequity. National zero-dose prevalence reflects the overall burden, the concentration index reflects relative concentration among disadvantaged children, the AEG reflects the absolute gap between the extremes of the disadvantage distribution, and the decomposition identifies context-specific contributors to inequity.

#### C: Analyses results of VERSE model across 77 countries

#### Supplementary Table 4. Measures of zero-dose prevalence across 77 countries

| Country | Year | Zero-dose prevalence (%) |
|---|---|---|
| **African Region** | | |
| Angola | 2015 | 30.10 |
| Benin | 2021 | 7.55 |
| Burkina Faso | 2021 | 4.52 |
| Burundi | 2016 | 0.76 |
| Cameroon | 2018 | 16.15 |
| Central African Republic | 2019 | 10.08 |
| Chad | 2019 | 18.42 |
| Comoros | 2022 | 7.57 |
| Côte d'Ivoire | 2021 | 29.52 |
| Democratic Republic of the Congo | 2023 | 33.10 |
| Eswatini | 2021 | 2.11 |
| Ethiopia | 2018 | 22.70 |
| Gabon | 2021 | 16.51 |
| The Gambia | 2020 | 1.48 |
| Ghana | 2022 | 2.77 |
| Guinea | 2018 | 37.21 |
| Guinea-Bissau | 2019 | 1.12 |
| Kenya | 2022 | 2.69 |
| Lesotho | 2024 | 0.66 |
| Liberia | 2019 | 8.37 |
| Madagascar | 2021 | 21.53 |
| Malawi | 2020 | 1.15 |
| Mali | 2024 | 16.06 |
| Mauritania | 2020 | 11.26 |
| Mozambique | 2022 | 20.82 |
| Nigeria | 2024 | 36.79 |
| Rwanda | 2020 | 0.39 |
| São Tomé and Príncipe | 2019 | 0.25 |
| Senegal | 2023 | 8.80 |
| Sierra Leone | 2019 | 4.79 |
| South Africa | 2016 | 4.97 |
| Tanzania | 2022 | 5.09 |
| Togo | 2017 | 2.27 |
| Uganda | 2016 | 4.65 |

| Country | Year | Zero-dose prevalence (%) |
|---|---|---|
| Zambia | 2018 | 1.80 |
| Zimbabwe | 2019 | 0.22 |
| **Region of the Americas** | | |
| Costa Rica | 2018 | 1.99 |
| Cuba | 2019 | 2.31 |
| Dominican Republic | 2019 | 2.89 |
| Guatemala | 2015 | 2.44 |
| Guyana | 2019 | 0.89 |
| Haiti | 2017 | 16.41 |
| Honduras | 2019 | 0.65 |
| Suriname | 2018 | 8.39 |
| Trinidad and Tobago | 2022 | 3.20 |
| **Eastern Mediterranean Region** | | |
| Afghanistan | 2023 | 5.82 |
| Algeria | 2019 | 1.60 |
| Iraq | 2018 | 5.22 |
| Jordan | 2023 | 2.18 |
| Pakistan | 2019 | 1.87 |
| Palestine | 2019 | 3.33 |
| Tunisia | 2023 | 0.36 |
| Yemen | 2023 | 2.40 |
| **European Region** | | |
| Armenia | 2016 | 1.54 |
| Azerbaijan | 2023 | 36.23 |
| Kyrgyzstan | 2023 | 8.39 |
| North Macedonia | 2018 | 3.45 |
| Serbia | 2019 | 1.94 |
| Tajikistan | 2023 | 9.95 |
| Turkey | 2018 | 3.54 |
| Uzbekistan | 2021 | 0.59 |
| **South-East Asia Region** | | |
| Bangladesh | 2017 | 1.51 |
| India | 2019 | 6.17 |
| Indonesia | 2017 | 10.80 |
| Myanmar | 2016 | 12.89 |
| Nepal | 2021 | 4.98 |
| Thailand | 2022 | 0.97 |

| Country | Year | Zero-dose prevalence (%) |
|---|---|---|
| Timor-Leste | 2016 | 21.38 |
| **Western Pacific Region** | | |
| Cambodia | 2021 | 6.93 |
| Fiji | 2021 | 0.67 |
| Laos | 2023 | 7.57 |
| Mongolia | 2018 | 4.17 |
| Papua New Guinea | 2016 | 34.42 |
| Philippines | 2022 | 13.04 |
| Tonga | 2019 | 2.13 |
| Vanuatu | 2023 | 12.57 |
| Vietnam | 2020 | 6.90 |

**Supplementary Table 5. Subnational distribution of zero-dose children prevalence and equity metrics across 77 countries in DHS & MICS survey, 2015 - 2024**

| Country | adm1_gid | adm1_name | N | Zero-dose Prevalence | CI | AEG |
|---|---|---|---|---|---|---|
| Afghanistan | AFG.10_1 | Ghor | 23 | 19.22 | -0.348 | -0.09 |
| Afghanistan | AFG.11_1 | Hilmand | 110 | 10.12 | -0.097 | -0.05 |
| Afghanistan | AFG.12_1 | Hirat | 158 | 3.92 | 0.728 | 0.136 |
| Afghanistan | AFG.13_1 | Jawzjan | 145 | 8.83 | 0.141 | 0.067 |
| Afghanistan | AFG.14_1 | Kabul | 149 | 4.43 | 0.119 | 0.018 |
| Afghanistan | AFG.15_1 | Kandahar | 123 | 14.65 | 0.146 | 0.08 |
| Afghanistan | AFG.16_1 | Kapisa | 86 | 8.98 | 0.048 | -0.022 |
| Afghanistan | AFG.17_1 | Khost | 225 | 13.76 | -0.015 | 0.06 |
| Afghanistan | AFG.18_1 | Kunar | 121 | 5.66 | -0.109 | -0.066 |
| Afghanistan | AFG.19_1 | Kunduz | 93 | 7.91 | 0.237 | 0.105 |
| Afghanistan | AFG.1_1 | Badakhshan | 95 | 0 | | 0 |
| Afghanistan | AFG.20_1 | Laghman | 131 | 2.82 | 0.286 | 0.034 |
| Afghanistan | AFG.21_1 | Logar | 150 | 5.05 | -0.031 | -0.037 |
| Afghanistan | AFG.22_1 | Nangarhar | 134 | 6.22 | 0.46 | 0.087 |
| Afghanistan | AFG.23_1 | Nimroz | 142 | 1.28 | 0.246 | 0 |
| Afghanistan | AFG.24_1 | Nuristan | 46 | 1.53 | 0.37 | 0 |
| Afghanistan | AFG.25_1 | Paktika | 79 | 8.58 | 0.435 | 0.296 |
| Afghanistan | AFG.26_1 | Paktya | 134 | 12.67 | -0.091 | -0.062 |
| Afghanistan | AFG.27_1 | Panjshir | 80 | 8.06 | 0.354 | 0.08 |
| Afghanistan | AFG.28_1 | Parwan | 135 | 4.65 | 0.41 | 0.069 |
| Afghanistan | AFG.29_1 | Samangan | 42 | 5.87 | 0.381 | 0.191 |
| Afghanistan | AFG.2_1 | Badghis | 48 | 8.46 | 0.021 | 0.119 |
| Afghanistan | AFG.30_1 | Sari Pul | 141 | 7.34 | 0.429 | 0.174 |
| Afghanistan | AFG.31_1 | Takhar | 134 | 4.59 | -0.072 | -0.041 |
| Afghanistan | AFG.32_1 | Uruzgan | 6 | 17.61 | -0.833 | -0.59 |
| Afghanistan | AFG.33_1 | Wardak | 116 | 5.41 | 0.494 | 0.169 |
| Afghanistan | AFG.34_1 | Zabul | 83 | 7.45 | 0.313 | 0.053 |
| Afghanistan | AFG.3_1 | Baghlan | 100 | 6.79 | 0.621 | 0.139 |
| Afghanistan | AFG.4_1 | Balkh | 119 | 2.75 | 0.179 | 0.042 |
| Afghanistan | AFG.5_1 | Bamyan | 123 | 0.58 | 0.862 | 0.029 |
| Afghanistan | AFG.6_1 | Daykundi | 134 | 1.04 | 0.664 | 0.049 |
| Afghanistan | AFG.7_1 | Farah | 88 | 1.62 | 0.193 | 0 |
| Afghanistan | AFG.8_1 | Faryab | 128 | 5.43 | 0.123 | 0.022 |
| Afghanistan | AFG.9_1 | Ghazni | 91 | 5.19 | 0.185 | 0.007 |

| Country | adm1_gid | adm1_name | N | Zero-dose Prevalence | CI | AEG |
|---|---|---|---|---|---|---|
| Albania | ALB.10_1 | Shkodër | 29 | 0 | | |
| Albania | ALB.11_1 | Tiranë | 43 | 0 | | |
| Albania | ALB.12_1 | Vlorë | 28 | 0 | | |
| Albania | ALB.1_1 | Berat | 44 | 0 | | |
| Albania | ALB.2_1 | Dibër | 56 | 0 | | |
| Albania | ALB.3_1 | Durrës | 47 | 0 | | |
| Albania | ALB.4_1 | Elbasan | 33 | 0 | | |
| Albania | ALB.5_1 | Fier | 37 | 0 | | |
| Albania | ALB.6_1 | Gjirokastër | 32 | 0 | | |
| Albania | ALB.7_1 | Korçë | 50 | 0 | | |
| Albania | ALB.8_1 | Kukës | 55 | 0 | | |
| Albania | ALB.9_1 | Lezhë | 41 | 0 | | |
| Algeria | DZA.2_1 | Aïn Defla | 439 | 6.85 | 0.273 | 0.136 |
| Algeria | DZA.12_1 | Bouira | 439 | 6.85 | 0.273 | 0.136 |
| Algeria | DZA.16_1 | Djelfa | 439 | 6.85 | 0.273 | 0.136 |
| Algeria | DZA.25_1 | Laghouat | 439 | 6.85 | 0.273 | 0.136 |
| Algeria | DZA.26_1 | M'Sila | 439 | 6.85 | 0.273 | 0.136 |
| Algeria | DZA.28_1 | Médéa | 439 | 6.85 | 0.273 | 0.136 |
| Algeria | DZA.6_1 | Batna | 373 | 2.18 | 0.609 | 0.067 |
| Algeria | DZA.9_1 | Biskra | 373 | 2.18 | 0.609 | 0.067 |
| Algeria | DZA.11_1 | Bordj Bou Arréridj | 373 | 2.18 | 0.609 | 0.067 |
| Algeria | DZA.24_1 | Khenchela | 373 | 2.18 | 0.609 | 0.067 |
| Algeria | DZA.37_1 | Sétif | 373 | 2.18 | 0.609 | 0.067 |
| Algeria | DZA.42_1 | Tébessa | 373 | 2.18 | 0.609 | 0.067 |
| Algeria | DZA.17_1 | El Bayadh | 350 | 0.72 | -0.263 | -0.021 |
| Algeria | DZA.31_1 | Naâma | 350 | 0.72 | -0.263 | -0.021 |
| Algeria | DZA.36_1 | Saïda | 350 | 0.72 | -0.263 | -0.021 |
| Algeria | DZA.43_1 | Tiaret | 350 | 0.72 | -0.263 | -0.021 |
| Algeria | DZA.46_1 | Tissemsilt | 350 | 0.72 | -0.263 | -0.021 |
| Algeria | DZA.4_1 | Alger | 313 | 0.9 | 0.077 | 0.01 |
| Algeria | DZA.10_1 | Blida | 313 | 0.9 | 0.077 | 0.01 |
| Algeria | DZA.13_1 | Boumerdès | 313 | 0.9 | 0.077 | 0.01 |
| Algeria | DZA.45_1 | Tipaza | 313 | 0.9 | 0.077 | 0.01 |
| Algeria | DZA.47_1 | Tizi Ouzou | 313 | 0.9 | 0.077 | 0.01 |
| Algeria | DZA.5_1 | Annaba | 338 | 0.59 | 0.059 | 0 |
| Algeria | DZA.8_1 | Béjaïa | 338 | 0.59 | 0.059 | 0 |
| Algeria | DZA.15_1 | Constantine | 338 | 0.59 | 0.059 | 0 |

| Country | adm1_gid | adm1_name | N | Zero-dose Prevalence | CI | AEG |
|---|---|---|---|---|---|---|
| Algeria | DZA.19_1 | El Tarf | 338 | 0.59 | 0.059 | 0 |
| Algeria | DZA.21_1 | Guelma | 338 | 0.59 | 0.059 | 0 |
| Algeria | DZA.23_1 | Jijel | 338 | 0.59 | 0.059 | 0 |
| Algeria | DZA.29_1 | Mila | 338 | 0.59 | 0.059 | 0 |
| Algeria | DZA.34_1 | Oum el Bouaghi | 338 | 0.59 | 0.059 | 0 |
| Algeria | DZA.39_1 | Skikda | 338 | 0.59 | 0.059 | 0 |
| Algeria | DZA.40_1 | Souk Ahras | 338 | 0.59 | 0.059 | 0 |
| Algeria | DZA.27_1 | Mascara | 299 | 1.46 | 0.396 | 0.012 |
| Algeria | DZA.3_1 | Aïn Témouchent | 299 | 1.46 | 0.396 | 0.012 |
| Algeria | DZA.14_1 | Chlef | 299 | 1.46 | 0.396 | 0.012 |
| Algeria | DZA.30_1 | Mostaganem | 299 | 1.46 | 0.396 | 0.012 |
| Algeria | DZA.32_1 | Oran | 299 | 1.46 | 0.396 | 0.012 |
| Algeria | DZA.35_1 | Relizane | 299 | 1.46 | 0.396 | 0.012 |
| Algeria | DZA.38_1 | Sidi Bel Abbès | 299 | 1.46 | 0.396 | 0.012 |
| Algeria | DZA.48_1 | Tlemcen | 299 | 1.46 | 0.396 | 0.012 |
| Algeria | DZA.1_1 | Adrar | 516 | 1.17 | 0.328 | 0.022 |
| Algeria | DZA.7_1 | Béchar | 516 | 1.17 | 0.328 | 0.022 |
| Algeria | DZA.20_1 | Ghardaïa | 516 | 1.17 | 0.328 | 0.022 |
| Algeria | DZA.22_1 | Illizi | 516 | 1.17 | 0.328 | 0.022 |
| Algeria | DZA.33_1 | Ouargla | 516 | 1.17 | 0.328 | 0.022 |
| Algeria | DZA.41_1 | Tamanghasset | 516 | 1.17 | 0.328 | 0.022 |
| Algeria | DZA.44_1 | Tindouf | 516 | 1.17 | 0.328 | 0.022 |
| Algeria | DZA.18_1 | El Oued | 516 | 1.17 | 0.328 | 0.022 |
| Angola | AGO.10_1 | Huíla | 163 | 37.84 | 0.258 | 0.526 |
| Angola | AGO.11_1 | Luanda | 224 | 9.87 | 0.305 | 0.142 |
| Angola | AGO.12_1 | Lunda Norte | 166 | 47.18 | 0.307 | 0.67 |
| Angola | AGO.13_1 | Lunda Sul | 161 | 18.13 | 0.292 | 0.311 |
| Angola | AGO.14_1 | Malanje | 152 | 20.49 | 0.445 | 0.4 |
| Angola | AGO.15_1 | Moxico | 130 | 67.15 | 0.168 | 0.569 |
| Angola | AGO.16_1 | Namibe | 146 | 34.66 | 0.276 | 0.41 |
| Angola | AGO.17_1 | Uíge | 155 | 38.87 | 0.277 | 0.586 |
| Angola | AGO.18_1 | Zaire | 162 | 17.89 | 0.159 | 0.037 |
| Angola | AGO.1_1 | Bengo | 122 | 39.08 | 0.18 | 0.486 |
| Angola | AGO.2_1 | Benguela | 175 | 40.26 | 0.438 | 0.738 |
| Angola | AGO.3_1 | Bié | 171 | 50.68 | 0.232 | 0.519 |
| Angola | AGO.4_1 | Cabinda | 118 | 13.89 | 0.355 | 0.24 |
| Angola | AGO.5_1 | Cuando Cubango | 146 | 54.36 | 0.191 | 0.463 |

| Country | adm1_gid | adm1_name | N | Zero-dose Prevalence | CI | AEG |
|---|---|---|---|---|---|---|
| Angola | AGO.6_1 | Cuanza Norte | 123 | 37.79 | 0.279 | 0.294 |
| Angola | AGO.7_1 | Cuanza Sul | 147 | 57.24 | 0.113 | 0.41 |
| Angola | AGO.8_1 | Cunene | 171 | 19.04 | 0.138 | 0.175 |
| Angola | AGO.9_1 | Huambo | 180 | 18.26 | 0.443 | 0.385 |
| Armenia | ARM.10_1 | Tavush | 40 | 1.85 | 0.925 | 0.093 |
| Armenia | ARM.11_1 | Vayots Dzor | 25 | 4.51 | -0.08 | 0 |
| Armenia | ARM.1_1 | Aragatsotn | 12 | 0 | | 0 |
| Armenia | ARM.2_1 | Ararat | 47 | 4.75 | 0.362 | 0.136 |
| Armenia | ARM.3_1 | Armavir | 39 | 0 | | 0 |
| Armenia | ARM.4_1 | Erevan | 51 | 0 | | 0 |
| Armenia | ARM.5_1 | Gegharkunik | 14 | 10.46 | 0.429 | 0.341 |
| Armenia | ARM.6_1 | Kotayk | 52 | 0 | | 0 |
| Armenia | ARM.7_1 | Lori | 15 | 0 | | 0 |
| Armenia | ARM.8_1 | Shirak | 30 | 0 | | 0 |
| Armenia | ARM.9_1 | Syunik | 20 | 10.58 | 0.4 | 0 |
| Azerbaijan | AZE.10_1 | Yukhari-Karabakh | 10 | 21.54 | 0 | -0.285 |
| Azerbaijan | AZE.1_1 | Absheron | 11 | 46.56 | 0.545 | 0.882 |
| Azerbaijan | AZE.2_1 | Aran | 10 | 54.51 | 0.26 | 0.633 |
| Azerbaijan | AZE.3_1 | Daglig-Shirvan | 4 | 35.3 | 0.5 | 1 |
| Azerbaijan | AZE.4_1 | Ganja-Qazakh | 6 | 0 | | 0 |
| Azerbaijan | AZE.6_1 | Lankaran | 5 | 22.77 | 0.8 | 1 |
| Azerbaijan | AZE.7_1 | Nakhchivan | 12 | 29.46 | 0 | -0.237 |
| Azerbaijan | AZE.8_1 | Quba-Khachmaz | 1 | 100 | 0 | 0 |
| Azerbaijan | AZE.9_1 | Shaki-Zaqatala | 5 | 17.82 | 0.4 | 0 |
| Bangladesh | BGD.1_1 | Barisal | 176 | 2.79 | -0.261 | -0.024 |
| Bangladesh | BGD.2_1 | Chittagong | 274 | 1.18 | 0.515 | 0.011 |
| Bangladesh | BGD.3_1 | Dhaka | 243 | 2.08 | 0.683 | 0.048 |
| Bangladesh | BGD.4_1 | Khulna | 160 | 0 | | 0 |
| Bangladesh | BGD.5_1 | Rajshahi | 173 | 0 | | 0 |
| Bangladesh | BGD.6_1 | Rangpur | 192 | 0.61 | 0.693 | 0.028 |
| Bangladesh | BGD.7_1 | Sylhet | 248 | 4.23 | 0.449 | 0.046 |
| Bangladesh | BGD.8_1 | Mymensingh | 200 | 1.81 | 0.445 | 0.067 |
| Benin | BEN.10_1 | Ouémé | 138 | 3.21 | 0.489 | 0.083 |
| Benin | BEN.11_1 | Plateau | 119 | 5.66 | 0.445 | 0.126 |
| Benin | BEN.12_1 | Zou | 192 | 9.93 | 0.344 | 0.164 |
| Benin | BEN.1_1 | Alibori | 197 | 18.16 | -0.038 | -0.154 |
| Benin | BEN.2_1 | Atakora | 247 | 8.4 | 0.297 | 0.151 |

| Country | adm1_gid | adm1_name | N | Zero-dose Prevalence | CI | AEG |
|---|---|---|---|---|---|---|
| Benin | BEN.3_1 | Atlantique | 153 | 3.63 | 0.583 | 0.096 |
| Benin | BEN.4_1 | Borgou | 182 | 9.47 | 0.736 | 0.228 |
| Benin | BEN.5_1 | Collines | 177 | 4.5 | 0.117 | -0.008 |
| Benin | BEN.6_1 | Donga | 258 | 12.2 | 0.133 | 0.117 |
| Benin | BEN.7_1 | Kouffo | 194 | 15.76 | 0.27 | 0.155 |
| Benin | BEN.8_1 | Littoral | 100 | 3.41 | 0.53 | 0.089 |
| Benin | BEN.9_1 | Mono | 197 | 0 | | 0 |
| Burkina Faso | BFA.10_1 | Nord | 201 | 3.59 | 0.304 | 0.03 |
| Burkina Faso | BFA.11_1 | Plateau-Central | 224 | 2.39 | -0.167 | -0.013 |
| Burkina Faso | BFA.12_1 | Sahel | 85 | 4.67 | -0.247 | -0.013 |
| Burkina Faso | BFA.13_1 | Sud-Ouest | 173 | 3.43 | 0.447 | 0.082 |
| Burkina Faso | BFA.1_1 | Boucle du Mouhoun | 189 | 11.79 | 0.458 | 0.261 |
| Burkina Faso | BFA.2_1 | Cascades | 99 | 0.95 | -0.465 | 0 |
| Burkina Faso | BFA.3_1 | Centre-Est | 223 | 2.83 | 0.217 | 0.029 |
| Burkina Faso | BFA.4_1 | Centre-Nord | 181 | 6.68 | 0.096 | 0.09 |
| Burkina Faso | BFA.5_1 | Centre-Ouest | 188 | 1.65 | -0.424 | 0 |
| Burkina Faso | BFA.6_1 | Centre-Sud | 129 | 1.65 | 0.31 | 0.042 |
| Burkina Faso | BFA.7_1 | Centre | 204 | 2.76 | 0.299 | 0.069 |
| Burkina Faso | BFA.8_1 | Est | 157 | 5.92 | -0.058 | 0.019 |
| Burkina Faso | BFA.9_1 | Haut-Bassins | 247 | 5.6 | 0.076 | 0.026 |
| Burundi | BDI.10_1 | Kirundo | 174 | 2.67 | 0.796 | 0.125 |
| Burundi | BDI.11_1 | Makamba | 149 | 0 | | 0 |
| Burundi | BDI.12_1 | Muramvya | 131 | 0.79 | 0.214 | 0 |
| Burundi | BDI.13_1 | Muyinga | 173 | 0 | | 0 |
| Burundi | BDI.14_1 | Mwaro | 106 | 0 | | 0 |
| Burundi | BDI.15_1 | Ngozi | 167 | 1.19 | 0.784 | 0.055 |
| Burundi | BDI.16_1 | Rutana | 138 | 0 | | 0 |
| Burundi | BDI.17_1 | Ruyigi | 174 | 0.62 | 0.73 | 0.03 |
| Burundi | BDI.1_1 | Bubanza | 173 | 2.63 | 0.107 | 0.06 |
| Burundi | BDI.2_1 | Bujumbura Mairie | 104 | 0.8 | 0.952 | 0.046 |
| Burundi | BDI.3_1 | Bujumbura Rural | 150 | 0 | | 0 |
| Burundi | BDI.4_1 | Bururi | 230 | 0.48 | 0.509 | 0 |
| Burundi | BDI.5_1 | Cankuzo | 142 | 0 | | 0 |
| Burundi | BDI.6_1 | Cibitoke | 156 | 1.29 | 0.244 | 0.029 |
| Burundi | BDI.7_1 | Gitega | 145 | 0 | | 0 |
| Burundi | BDI.8_1 | Karuzi | 140 | 0.86 | 0.021 | 0 |
| Burundi | BDI.9_1 | Kayanza | 143 | 0.21 | -0.713 | -0.011 |

| Country | adm1_gid | adm1_name | N | Zero-dose Prevalence | CI | AEG |
|---|---|---|---|---|---|---|
| Cambodia | KHM.10_1 | Kep | 64 | 3.22 | 0.63 | 0.105 |
| Cambodia | KHM.11_1 | Krâchéh | 59 | 2.93 | 0.288 | 0.038 |
| Cambodia | KHM.12_1 | Krong Pailin | 60 | 18.66 | -0.042 | -0.205 |
| Cambodia | KHM.13_1 | Krong Preah Sihanouk | 58 | 10.96 | -0.106 | -0.005 |
| Cambodia | KHM.14_1 | Môndól Kiri | 75 | 10.86 | 0.543 | 0.186 |
| Cambodia | KHM.15_1 | Otdar Mean Chey | 69 | 15.11 | 0.047 | 0.106 |
| Cambodia | KHM.16_1 | Phnom Penh | 88 | 6.18 | -0.008 | 0 |
| Cambodia | KHM.17_1 | Pouthisat | 39 | 11.95 | 0.154 | 0.116 |
| Cambodia | KHM.18_1 | Preah Vihéar | 64 | 11.58 | 0.203 | 0.155 |
| Cambodia | KHM.19_1 | Prey Vêng | 82 | 1.25 | 0.427 | 0 |
| Cambodia | KHM.1_1 | Bântéay Méanchey | 52 | 7.21 | 0.353 | 0 |
| Cambodia | KHM.20_1 | Rôtânôkiri | 59 | 51.98 | 0.089 | 0.265 |
| Cambodia | KHM.21_1 | Siemréab | 76 | 2.02 | 0.285 | 0.022 |
| Cambodia | KHM.22_1 | Stœng Trêng | 90 | 8.92 | 0.411 | 0.165 |
| Cambodia | KHM.23_1 | Svay Rieng | 69 | 2.9 | 0.507 | 0.077 |
| Cambodia | KHM.24_1 | Takêv | 62 | 6.49 | 0.218 | 0.003 |
| Cambodia | KHM.25_1 | Tbong Khmum | 59 | 7.31 | 0.432 | 0.073 |
| Cambodia | KHM.2_1 | Batdâmbâng | 63 | 0 | | 0 |
| Cambodia | KHM.3_1 | Kâmpóng Cham | 60 | 4.24 | -0.25 | -0.077 |
| Cambodia | KHM.4_1 | Kâmpóng Chhnang | 64 | 0.95 | -0.422 | 0 |
| Cambodia | KHM.5_1 | Kâmpóng Spœ | 64 | 4.16 | 0.536 | 0.141 |
| Cambodia | KHM.6_1 | Kâmpóng Thum | 79 | 13.71 | 0.289 | 0.249 |
| Cambodia | KHM.7_1 | Kâmpôt | 65 | 24.65 | 0.017 | -0.075 |
| Cambodia | KHM.8_1 | Kândal | 71 | 4.89 | 0.667 | 0.149 |
| Cambodia | KHM.9_1 | Kaôh Kong | 72 | 18.28 | 0.33 | 0.217 |
| Cameroon | CMR.10_1 | Sud | 162 | 11.23 | 0.238 | 0.131 |
| Cameroon | CMR.1_1 | Adamaoua | 119 | 24.83 | 0.078 | 0.119 |
| Cameroon | CMR.2_1 | Centre | 338 | 9.4 | 0.22 | 0.153 |
| Cameroon | CMR.3_1 | Est | 166 | 20.05 | 0.275 | 0.334 |
| Cameroon | CMR.4_1 | Extrême-Nord | 241 | 23.2 | 0.184 | 0.182 |
| Cameroon | CMR.5_1 | Littoral | 203 | 9.18 | 0.126 | 0.075 |
| Cameroon | CMR.6_1 | Nord-Ouest | 99 | 5.13 | 0.862 | 0.166 |
| Cameroon | CMR.7_1 | Nord | 245 | 25.73 | 0.223 | 0.22 |
| Cameroon | CMR.8_1 | Ouest | 213 | 12.82 | 0.348 | 0.221 |
| Cameroon | CMR.9_1 | Sud-Ouest | 23 | 0 | | 0 |
| Central African Republic | CAF.7_1 | Lobaye | 131 | 4.57 | 0.321 | 0.098 |

| Country | adm1_gid | adm1_name | N | Zero-dose Prevalence | CI | AEG |
|---|---|---|---|---|---|---|
| Central African Republic | CAF.12_1 | Ombella-M'Poko | 131 | 4.57 | 0.321 | 0.098 |
| Central African Republic | CAF.8_1 | Mambéré-Kadéï | 101 | 21.52 | 0.161 | 0.219 |
| Central African Republic | CAF.16_1 | Sangha-Mbaéré | 101 | 21.52 | 0.161 | 0.219 |
| Central African Republic | CAF.11_1 | Nana-Mambéré | 101 | 21.52 | 0.161 | 0.219 |
| Central African Republic | CAF.15_1 | Ouham | 94 | 6.49 | 0.5 | 0.179 |
| Central African Republic | CAF.14_1 | Ouham-Pendé | 94 | 6.49 | 0.5 | 0.179 |
| Central African Republic | CAF.6_1 | Kémo | 97 | 7.64 | 0.174 | 0.085 |
| Central African Republic | CAF.10_1 | Nana-Grébizi | 97 | 7.64 | 0.174 | 0.085 |
| Central African Republic | CAF.13_1 | Ouaka | 97 | 7.64 | 0.174 | 0.085 |
| Central African Republic | CAF.1_1 | Bamingui-Bangoran | 78 | 4.52 | -0.038 | 0 |
| Central African Republic | CAF.5_1 | Haute-Kotto | 78 | 4.52 | -0.038 | 0 |
| Central African Republic | CAF.17_1 | Vakaga | 78 | 4.52 | -0.038 | 0 |
| Central African Republic | CAF.3_1 | Basse-Kotto | 34 | 33.9 | 0.167 | 0.41 |
| Central African Republic | CAF.9_1 | Mbomou | 34 | 33.9 | 0.167 | 0.41 |
| Central African Republic | CAF.4_1 | Haut-Mbomou | 34 | 33.9 | 0.167 | 0.41 |
| Central African Republic | | Région 7 | 183 | 6.18 | 0.242 | 0.094 |
| Chad | TCD.10_1 | Lac | 38 | 13.73 | 0.07 | 0.116 |
| Chad | TCD.11_1 | Logone Occidental | 81 | 9.84 | -0.187 | -0.143 |
| Chad | TCD.12_1 | Logone Oriental | 59 | 33.39 | 0.028 | 0.151 |
| Chad | TCD.13_1 | Mandoul | 129 | 4.84 | 0.463 | 0.12 |
| Chad | TCD.14_1 | Mayo-Kebbi Est | 71 | 23.11 | 0.13 | -0.019 |

| Country | adm1_gid | adm1_name | N | Zero-dose Prevalence | CI | AEG |
|---|---|---|---|---|---|---|
| Chad | TCD.15_1 | Mayo-Kebbi Ouest | 126 | 12.56 | -0.145 | -0.137 |
| Chad | TCD.16_1 | Moyen-Chari | 86 | 6.61 | 0.135 | 0.055 |
| Chad | TCD.17_1 | Ouaddaï | 33 | 16.74 | 0.26 | 0.197 |
| Chad | TCD.18_1 | Salamat | 50 | 9.57 | 0.117 | -0.142 |
| Chad | TCD.19_1 | Sila | 27 | 30.53 | 0.159 | 0.175 |
| Chad | TCD.1_1 | Barh el Ghazel | 49 | 28.67 | 0.079 | 0.077 |
| Chad | TCD.20_1 | Tandjilé | 60 | 15.06 | 0.008 | -0.035 |
| Chad | TCD.22_1 | Ville de N'Djamena | 74 | 33.42 | 0.255 | 0.433 |
| Chad | TCD.23_1 | Wadi Fira | 40 | 10.89 | -0.317 | -0.262 |
| Chad | TCD.2_1 | Batha | 36 | 33.99 | 0.241 | 0.249 |
| Chad | TCD.3_1 | Borkou | 19 | 12.07 | -0.07 | -0.199 |
| Chad | TCD.4_1 | Chari-Baguirmi | 38 | 22.18 | 0.176 | 0.006 |
| Chad | TCD.5_1 | Ennedi Est | 17 | 5.27 | 0.706 | 0.2 |
| Chad | TCD.6_1 | Ennedi Ouest | 14 | 6.85 | 0.786 | 0.354 |
| Chad | TCD.7_1 | Guéra | 77 | 25.01 | 0.209 | 0.079 |
| Chad | TCD.8_1 | Hadjer-Lamis | 90 | 24.43 | 0.171 | 0.353 |
| Chad | TCD.9_1 | Kanem | 41 | 28.71 | -0.07 | -0.019 |
| Comoros | COM.1_1 | Nzwani | 192 | 10.27 | 0.375 | 0.227 |
| Comoros | COM.2_1 | Njazídja | 308 | 5.16 | 0.403 | 0.135 |
| Comoros | COM.3_1 | Mwali | 168 | 8.33 | 0.004 | -0.001 |
| Costa Rica | CRI.1_1 | Alajuela | 130 | 0 |  | 0 |
| Costa Rica | CRI.2_1 | Cartago | 61 | 14.17 | 0.574 | 0.41 |
| Costa Rica | CRI.3_1 | Guanacaste | 72 | 1.12 | 0.986 | 0.051 |
| Costa Rica | CRI.4_1 | Heredia | 36 | 0.2 | 0.75 | 0.009 |
| Costa Rica | CRI.5_1 | Limón | 104 | 0 |  | 0 |
| Costa Rica | CRI.6_1 | Puntarenas | 115 | 6.9 | 0.243 | 0.063 |
| Costa Rica | CRI.7_1 | San José | 152 | 0.12 | -0.257 | 0 |
| Côte d'Ivoire | CIV.10_1 | Savanes | 138 | 17.72 | 0.056 | 0.045 |
| Côte d'Ivoire | CIV.11_1 | Vallée du Bandama | 116 | 10.32 | 0.247 | 0.028 |
| Côte d'Ivoire | CIV.12_1 | Woroba | 149 | 47.61 | 0.115 | 0.267 |
| Côte d'Ivoire | CIV.13_1 | Yamoussoukro | 77 | 11.29 | 0.199 | 0.14 |
| Côte d'Ivoire | CIV.14_1 | Zanzan | 122 | 23.66 | 0.189 | 0.222 |
| Côte d'Ivoire | CIV.1_1 | Abidjan | 132 | 41.3 | 0.183 | 0.317 |
| Côte d'Ivoire | CIV.2_1 | Bas-Sassandra | 144 | 22.99 | 0.126 | 0.122 |
| Côte d'Ivoire | CIV.3_1 | Comoé | 98 | 25.64 | -0.018 | -0.072 |
| Côte d'Ivoire | CIV.4_1 | Denguélé | 196 | 21.93 | -0.101 | -0.15 |
| Côte d'Ivoire | CIV.5_1 | Gôh-Djiboua | 137 | 27.8 | 0.15 | 0.193 |

| Country | adm1_gid | adm1_name | N | Zero-dose Prevalence | CI | AEG |
|---|---|---|---|---|---|---|
| Côte d'Ivoire | CIV.6_1 | Lacs | 132 | 19.91 | 0.301 | 0.388 |
| Côte d'Ivoire | CIV.7_1 | Lagunes | 111 | 27.49 | 0.091 | 0.244 |
| Côte d'Ivoire | CIV.8_1 | Montagnes | 179 | 16.45 | 0.326 | 0.132 |
| Côte d'Ivoire | CIV.9_1 | Sassandra-Marahoué | 178 | 46.66 | 0.031 | 0.092 |
| Cuba | CUB.10_1 | Las Tunas | 72 | 0.54 | -0.958 | -0.022 |
| Cuba | CUB.11_1 | Matanzas | 80 | 9.01 | -0.013 | 0.415 |
| Cuba | CUB.12_1 | Mayabeque | 67 | 0 | | 0 |
| Cuba | CUB.13_1 | Pinar del Río | 85 | 0 | | 0 |
| Cuba | CUB.14_1 | Sancti Spíritus | 70 | 0 | | 0 |
| Cuba | CUB.15_1 | Santiago de Cuba | 57 | 0 | | 0 |
| Cuba | CUB.16_1 | Villa Clara | 68 | 0 | | 0 |
| Cuba | CUB.1_1 | Camagüey | 69 | 1.88 | 0.29 | 0 |
| Cuba | CUB.2_1 | Ciego de Ávila | 68 | 4.52 | 0.985 | 0.233 |
| Cuba | CUB.3_1 | Cienfuegos | 59 | 0 | | 0 |
| Cuba | CUB.4_1 | Ciudad de la Habana | 70 | 0 | | 0 |
| Cuba | CUB.5_1 | Granma | 66 | 0 | | 0 |
| Cuba | CUB.6_1 | Guantánamo | 93 | 0.59 | 0.989 | 0.025 |
| Cuba | CUB.7_1 | Holguín | 60 | 1.09 | 0.983 | 0.03 |
| Cuba | CUB.8_1 | Isla de la Juventud | 15 | 0 | | 0 |
| Cuba | CUB.9_1 | La Habana | 91 | 9.44 | 0.542 | 0.339 |
| Democratic Republic of the Congo | COD.10_1 | Kinshasa | 116 | 0.75 | 0.957 | 0.028 |
| Democratic Republic of the Congo | COD.11_1 | Kongo-Central | 99 | 42.53 | 0.325 | 0.543 |
| Democratic Republic of the Congo | COD.12_1 | Kwango | 150 | 29.01 | 0.092 | 0.221 |
| Democratic Republic of the Congo | COD.13_1 | Kwilu | 175 | 21.3 | 0.095 | 0.084 |
| Democratic Republic of the Congo | COD.14_1 | Lomami | 166 | 34.65 | 0.334 | 0.594 |
| Democratic Republic of the | COD.15_1 | Lualaba | 215 | 33.94 | 0.148 | 0.403 |

| Country | adm1_gid | adm1_name | N | Zero-dose Prevalence | CI | AEG |
|---|---|---|---|---|---|---|
| Congo | | | | | | |
| Democratic Republic of the Congo | COD.16_1 | Mai-Ndombe | 97 | 47.8 | 0.196 | 0.338 |
| Democratic Republic of the Congo | COD.17_1 | Maniema | 207 | 78.35 | 0.042 | 0.188 |
| Democratic Republic of the Congo | COD.18_1 | Mongala | 135 | 76.09 | 0.096 | 0.496 |
| Democratic Republic of the Congo | COD.19_1 | Nord-Kivu | 196 | 9.42 | 0.354 | 0.174 |
| Democratic Republic of the Congo | COD.1_1 | Bas-Uele | 115 | 62.16 | 0.081 | 0.407 |
| Democratic Republic of the Congo | COD.20_1 | Nord-Ubangi | 150 | 38.27 | 0.208 | 0.347 |
| Democratic Republic of the Congo | COD.21_1 | Sankuru | 121 | 63.98 | 0.027 | 0.226 |
| Democratic Republic of the Congo | COD.22_1 | Sud-Kivu | 203 | 26.88 | 0.38 | 0.509 |
| Democratic Republic of the Congo | COD.23_1 | Sud-Ubangi | 199 | 59.76 | 0.151 | 0.38 |
| Democratic Republic of the Congo | COD.24_1 | Tanganyika | 168 | 51.65 | -0.015 | 0.275 |
| Democratic Republic of the Congo | COD.25_1 | Tshopo | 162 | 56.1 | 0.116 | 0.543 |
| Democratic Republic of the Congo | COD.26_1 | Tshuapa | 117 | 65.79 | 0.004 | -0.089 |

| Country | adm1_gid | adm1_name | N | Zero-dose Prevalence | CI | AEG |
|---|---|---|---|---|---|---|
| Democratic Republic of the Congo | COD.2_1 | Équateur | 150 | 46.45 | 0.154 | 0.423 |
| Democratic Republic of the Congo | COD.3_1 | Haut-Katanga | 202 | 25.73 | 0.449 | 0.404 |
| Democratic Republic of the Congo | COD.4_1 | Haut-Lomami | 220 | 46.93 | 0.182 | 0.271 |
| Democratic Republic of the Congo | COD.5_1 | Haut-Uele | 93 | 45.71 | 0.169 | 0.137 |
| Democratic Republic of the Congo | COD.6_1 | Ituri | 176 | 21.22 | 0.161 | 0.339 |
| Democratic Republic of the Congo | COD.7_1 | Kasaï-Central | 222 | 9.72 | 0.147 | 0.024 |
| Democratic Republic of the Congo | COD.8_1 | Kasaï-Oriental | 148 | 18.13 | 0.121 | 0.299 |
| Democratic Republic of the Congo | COD.9_1 | Kasaï | 182 | 51.83 | 0.123 | 0.517 |
| Dominican Republic | DOM.6_1 | Duarte | 134 | 2.74 | 0.366 | 0.064 |
| Dominican Republic | DOM.15_1 | María Trinidad Sánchez | 134 | 2.74 | 0.366 | 0.064 |
| Dominican Republic | DOM.22_1 | Salcedo | 134 | 2.74 | 0.366 | 0.064 |
| Dominican Republic | DOM.23_1 | Samaná | 134 | 2.74 | 0.366 | 0.064 |
| Dominican Republic | DOM.4_1 | Dajabón | 128 | 3.46 | 0.317 | 0.076 |
| Dominican Republic | DOM.17_1 | Monte Cristi | 128 | 3.46 | 0.317 | 0.076 |
| Dominican | DOM.32_1 | Valverde | 128 | 3.46 | 0.317 | 0.076 |

| Country | adm1_gid | adm1_name | N | Zero-dose Prevalence | CI | AEG |
|---|---|---|---|---|---|---|
| Republic | | | | | | |
| Dominican Republic | DOM.29_1 | Santiago Rodríguez | 128 | 3.46 | 0.317 | 0.076 |
| Dominican Republic | DOM.30_1 | Santiago | 153 | 6.42 | 0.198 | 0.039 |
| Dominican Republic | DOM.8_1 | Espaillat | 153 | 6.42 | 0.198 | 0.039 |
| Dominican Republic | DOM.21_1 | Puerto Plata | 153 | 6.42 | 0.198 | 0.039 |
| Dominican Republic | DOM.14_1 | La Vega | 122 | 0 | | 0 |
| Dominican Republic | DOM.16_1 | Monseñor Nouel | 122 | 0 | | 0 |
| Dominican Republic | DOM.28_1 | Sánchez Ramírez | 122 | 0 | | 0 |
| Dominican Republic | DOM.12_1 | La Estrelleta | 89 | 4.78 | 0.066 | 0.03 |
| Dominican Republic | DOM.26_1 | San Juan | 89 | 4.78 | 0.066 | 0.03 |
| Dominican Republic | DOM.2_1 | Bahoruco | 180 | 1.04 | 0.022 | -0.007 |
| Dominican Republic | DOM.3_1 | Barahona | 180 | 1.04 | 0.022 | -0.007 |
| Dominican Republic | DOM.10_1 | Independencia | 180 | 1.04 | 0.022 | -0.007 |
| Dominican Republic | DOM.19_1 | Pedernales | 180 | 1.04 | 0.022 | -0.007 |
| Dominican Republic | DOM.27_1 | San Pedro de Macorís | 107 | 2.02 | 0.047 | -0.002 |
| Dominican Republic | DOM.9_1 | Hato Mayor | 107 | 2.02 | 0.047 | -0.002 |
| Dominican Republic | DOM.18_1 | Monte Plata | 107 | 2.02 | 0.047 | -0.002 |
| Dominican Republic | DOM.5_1 | Distrito Nacional | 175 | 2.26 | 0.603 | 0.077 |
| Dominican Republic | DOM.31_1 | Santo Domingo | 175 | 2.26 | 0.603 | 0.077 |

| Country | adm1_gid | adm1_name | N | Zero-dose Prevalence | CI | AEG |
|---|---|---|---|---|---|---|
| Dominican Republic | DOM.1_1 | Azua | 223 | 1.74 | 0.11 | 0.024 |
| Dominican Republic | DOM.24_1 | San Cristóbal | 223 | 1.74 | 0.11 | 0.024 |
| Dominican Republic | DOM.20_1 | Peravia | 223 | 1.74 | 0.11 | 0.024 |
| Dominican Republic | DOM.25_1 | San José de Ocoa | 223 | 1.74 | 0.11 | 0.024 |
| Dominican Republic | DOM.13_1 | La Romana | 125 | 3.98 | 0.088 | -0.021 |
| Dominican Republic | DOM.7_1 | El Seybo | 125 | 3.98 | 0.088 | -0.021 |
| Dominican Republic | DOM.11_1 | La Altagracia | 125 | 3.98 | 0.088 | -0.021 |
| Eswatini | SWZ.1_1 | Hhohho | 94 | 0 | | 0 |
| Eswatini | SWZ.2_1 | Lubombo | 106 | 3.93 | 0.375 | 0.041 |
| Eswatini | SWZ.3_1 | Manzini | 90 | 0.95 | -0.056 | 0 |
| Eswatini | SWZ.4_1 | Shiselweni | 101 | 4.48 | 0.49 | 0.144 |
| Ethiopia | ETH.10_1 | Southern Nations, Nationalities | 114 | 26.14 | 0.221 | 0.382 |
| Ethiopia | ETH.11_1 | Tigray | 93 | 4.57 | 0.129 | 0.104 |
| Ethiopia | ETH.1_1 | Addis Abeba | 64 | 3.74 | 0.094 | -0.018 |
| Ethiopia | ETH.2_1 | Afar | 109 | 53 | 0.172 | 0.443 |
| Ethiopia | ETH.3_1 | Amhara | 97 | 14.15 | 0.325 | 0.203 |
| Ethiopia | ETH.4_1 | Benshangul-Gumaz | 82 | 9.81 | 0.183 | 0.154 |
| Ethiopia | ETH.5_1 | Dire Dawa | 80 | 4.78 | 0.332 | 0.112 |
| Ethiopia | ETH.6_1 | Gambela Peoples | 75 | 22.42 | 0.293 | 0.701 |
| Ethiopia | ETH.7_1 | Harari People | 70 | 33.34 | 0.379 | 0.525 |
| Ethiopia | ETH.8_1 | Oromia | 125 | 25.5 | 0.285 | 0.355 |
| Ethiopia | ETH.9_1 | Somali | 85 | 55.99 | 0.071 | -0.003 |
| Fiji | FJI.1_1 | Central | 142 | 0 | | 0 |
| Fiji | FJI.2_1 | Eastern | 36 | 0 | | 0 |
| Fiji | FJI.3_1 | Northern | 67 | 4.93 | 0.856 | 0.13 |
| Fiji | FJI.5_1 | Western | 136 | 0 | | 0 |
| Fiji | FJI.4_1 | Rotuma | 136 | 0 | | 0 |
| Gabon | GAB.1_1 | Estuaire | 246 | 18.25 | 0.223 | 0.137 |
| Gabon | GAB.2_1 | Haut-Ogooué | 102 | 14.47 | 0.249 | 0.219 |

| Country | adm1_gid | adm1_name | N | Zero-dose Prevalence | CI | AEG |
|---|---|---|---|---|---|---|
| Gabon | GAB.3_1 | Moyen-Ogooué | 99 | 7.53 | 0.311 | 0.122 |
| Gabon | GAB.4_1 | Ngounié | 114 | 18.39 | 0.338 | 0.424 |
| Gabon | GAB.5_1 | Nyanga | 78 | 9.88 | 0.252 | 0.043 |
| Gabon | GAB.6_1 | Ogooué-Ivindo | 119 | 26.9 | 0.35 | 0.452 |
| Gabon | GAB.7_1 | Ogooué-Lolo | 137 | 12.5 | 0.31 | 0.166 |
| Gabon | GAB.8_1 | Ogooué-Maritime | 228 | 11.52 | 0.292 | 0.326 |
| Gabon | GAB.9_1 | Wouleu-Ntem | 126 | 14.32 | 0.312 | 0.26 |
| The Gambia | GMB.1_1 | Banjul | 212 | 3.92 | 0.42 | 0.132 |
| The Gambia | GMB.2_1 | Lower River | 158 | 0.91 | 0.576 | 0.036 |
| The Gambia | GMB.3_1 | Maccarthy Island | 397 | 0.7 | 0.902 | 0.024 |
| The Gambia | GMB.4_1 | North Bank | 221 | 0 | | 0 |
| The Gambia | GMB.5_1 | Upper River | 316 | 2.67 | -0.264 | -0.031 |
| The Gambia | GMB.6_1 | Western | 276 | 0.88 | 0.478 | 0 |
| Ghana | GHA10_2 | Oti | 124 | 1.83 | 0.352 | 0.029 |
| Ghana | GHA11_2 | Savannah | 155 | 3.65 | 0.171 | 0.039 |
| Ghana | GHA12_2 | Upper East | 125 | 1.16 | 0.992 | 0.05 |
| Ghana | GHA13_2 | Upper West | 135 | 2.71 | 0 | -0.064 |
| Ghana | GHA14_2 | Volta | 89 | 0 | | 0 |
| Ghana | GHA15_2 | Western | 94 | 6.16 | 0.418 | 0.195 |
| Ghana | GHA16_2 | Western North | 98 | 0.93 | 0.5 | 0 |
| Ghana | GHA1_2 | Ahafo | 97 | 8.34 | 0.509 | 0.244 |
| Ghana | GHA2_2 | Ashanti | 140 | 0.62 | -0.036 | 0 |
| Ghana | GHA3_2 | Bono | 94 | 0 | | 0 |
| Ghana | GHA4_2 | Bono East | 154 | 1.13 | 0.435 | 0.042 |
| Ghana | GHA5_2 | Central | 111 | 0 | | 0 |
| Ghana | GHA6_2 | Eastern | 79 | 3.65 | -0.397 | -0.053 |
| Ghana | GHA7_2 | Greater Accra | 88 | 1.2 | 0.807 | 0.051 |
| Ghana | GHA8_2 | North East | 179 | 4.65 | 0.256 | 0.055 |
| Ghana | GHA9_2 | Northern | 208 | 10.94 | 0.518 | 0.259 |
| Guatemala | GTM.12_1 | Petén | 110 | 0.92 | 0.755 | 0.039 |
| Guatemala | GTM.7_1 | Guatemala | 168 | 1.94 | 0.688 | 0.078 |
| Guatemala | GTM.3_1 | Chimaltenango | 272 | 2.32 | 0.365 | 0.07 |
| Guatemala | GTM.16_1 | Sacatepéquez | 272 | 2.32 | 0.365 | 0.07 |
| Guatemala | GTM.6_1 | Escuintla | 272 | 2.32 | 0.365 | 0.07 |
| Guatemala | GTM.8_1 | Huehuetenango | 288 | 3.07 | 0.18 | 0.033 |
| Guatemala | GTM.14_1 | Quiché | 288 | 3.07 | 0.18 | 0.033 |
| Guatemala | GTM.4_1 | Chiquimula | 402 | 2.17 | -0.16 | -0.001 |

| Country | adm1_gid | adm1_name | N | Zero-dose Prevalence | CI | AEG |
|---|---|---|---|---|---|---|
| Guatemala | GTM.5_1 | El Progreso | 402 | 2.17 | -0.16 | -0.001 |
| Guatemala | GTM.9_1 | Izabal | 402 | 2.17 | -0.16 | -0.001 |
| Guatemala | GTM.22_1 | Zacapa | 402 | 2.17 | -0.16 | -0.001 |
| Guatemala | GTM.1_1 | Alta Verapaz | 227 | 3.43 | 0.184 | 0.055 |
| Guatemala | GTM.2_1 | Baja Verapaz | 227 | 3.43 | 0.184 | 0.055 |
| Guatemala | GTM.13_1 | Quezaltenango | 630 | 3.16 | 0.104 | 0.041 |
| Guatemala | GTM.15_1 | Retalhuleu | 630 | 3.16 | 0.104 | 0.041 |
| Guatemala | GTM.17_1 | San Marcos | 630 | 3.16 | 0.104 | 0.041 |
| Guatemala | GTM.19_1 | Sololá | 630 | 3.16 | 0.104 | 0.041 |
| Guatemala | GTM.20_1 | Suchitepéquez | 630 | 3.16 | 0.104 | 0.041 |
| Guatemala | GTM.21_1 | Totonicapán | 630 | 3.16 | 0.104 | 0.041 |
| Guatemala | GTM.10_1 | Jalapa | 310 | 0.18 | 0.939 | 0.008 |
| Guatemala | GTM.11_1 | Jutiapa | 310 | 0.18 | 0.939 | 0.008 |
| Guatemala | GTM.18_1 | Santa Rosa | 310 | 0.18 | 0.939 | 0.008 |
| Guinea | GIN.1_1 | Boké | 207 | 48.04 | 0.187 | 0.471 |
| Guinea | GIN.2_1 | Conakry | 126 | 19.59 | 0.204 | 0.136 |
| Guinea | GIN.3_1 | Faranah | 172 | 37.22 | 0.315 | 0.486 |
| Guinea | GIN.4_1 | Kankan | 241 | 24.62 | 0.15 | 0.149 |
| Guinea | GIN.5_1 | Kindia | 198 | 37.18 | 0.216 | 0.432 |
| Guinea | GIN.6_1 | Labé | 168 | 58.67 | 0.079 | 0.244 |
| Guinea | GIN.7_1 | Mamou | 117 | 45.55 | 0.194 | 0.504 |
| Guinea | GIN.8_1 | Nzérékoré | 168 | 39.51 | 0.16 | 0.4 |
| Guinea-Bissau | GNB.1_1 | Bafatá | 205 | 0 | | 0 |
| Guinea-Bissau | GNB.2_1 | Biombo | 139 | 0.78 | 0.277 | 0 |
| Guinea-Bissau | GNB.3_1 | Bissau | 105 | 0 | | 0 |
| Guinea-Bissau | GNB.4_1 | Bolama | 92 | 0 | | 0 |
| Guinea-Bissau | GNB.5_1 | Cacheu | 128 | 0.88 | 0.961 | 0.04 |
| Guinea-Bissau | GNB.6_1 | Gabú | 175 | 2.88 | 0.288 | 0.054 |
| Guinea-Bissau | GNB.7_1 | Oio | 199 | 2.37 | 0.571 | 0.076 |
| Guinea-Bissau | GNB.8_1 | Quinara | 147 | 1.26 | -0.156 | -0.021 |
| Guinea-Bissau | GNB.9_1 | Tombali | 118 | 1.01 | -0.161 | 0 |
| Guyana | GUY.10_1 | Upper Takutu-Upper Essequibo | 41 | 8.52 | 0.854 | 0.199 |
| Guyana | GUY.1_1 | Barima-Waini | 64 | 0 | | 0 |
| Guyana | GUY.2_1 | Cuyuni-Mazaruni | 29 | 0 | | 0 |
| Guyana | GUY.3_1 | Demerara-Mahaica | 72 | 0.54 | 0.847 | 0.024 |
| Guyana | GUY.4_1 | East Berbice-Corentyne | 68 | 0 | | 0 |

| Country | adm1_gid | adm1_name | N | Zero-dose Prevalence | CI | AEG |
|---|---|---|---|---|---|---|
| Guyana | GUY.5_1 | Essequibo Islands-West Demerara | 62 | 0 | | 0 |
| Guyana | GUY.6_1 | Mahaica-Berbice | 37 | 1.92 | 0.486 | 0 |
| Guyana | GUY.7_1 | Pomeroon-Supenaam | 52 | 4.13 | 0 | 0 |
| Guyana | GUY.8_1 | Potaro-Siparuni | 35 | 0 | | 0 |
| Guyana | GUY.9_1 | Upper Demerara-Berbice | 31 | 0 | | 0 |
| Haiti | HTI.10_1 | Sud | 90 | 25.01 | 0.437 | 0.567 |
| Haiti | HTI.1_1 | Centre | 121 | 18.12 | 0.332 | 0.278 |
| Haiti | HTI.2_1 | Grand'Anse | 107 | 18.33 | 0.433 | 0.405 |
| Haiti | HTI.3_1 | L'Artibonite | 154 | 15.04 | 0.449 | 0.242 |
| Haiti | HTI.4_1 | Nippes | 74 | 2.85 | 0.027 | 0 |
| Haiti | HTI.5_1 | Nord-Est | 96 | 8.54 | 0.374 | 0.212 |
| Haiti | HTI.6_1 | Nord-Ouest | 129 | 18.6 | 0.226 | 0.186 |
| Haiti | HTI.7_1 | Nord | 111 | 10.96 | 0.326 | 0.218 |
| Haiti | HTI.8_1 | Ouest | 233 | 18.35 | 0.352 | 0.203 |
| Haiti | HTI.9_1 | Sud-Est | 80 | 14.31 | 0.299 | 0.12 |
| Honduras | HND.10_1 | Intibucá | 108 | 0 | | 0 |
| Honduras | HND.11_1 | Islas de la Bahía | 51 | 0 | | 0 |
| Honduras | HND.12_1 | La Paz | 80 | 1.61 | 0.35 | 0.024 |
| Honduras | HND.13_1 | Lempira | 99 | 0 | | 0 |
| Honduras | HND.14_1 | Ocotepeque | 55 | 1.95 | -0.145 | 0 |
| Honduras | HND.15_1 | Olancho | 110 | 0 | | 0 |
| Honduras | HND.16_1 | Santa Bárbara | 73 | 0 | | 0 |
| Honduras | HND.17_1 | Valle | 75 | 0 | | 0 |
| Honduras | HND.18_1 | Yoro | 100 | 1.02 | 0.97 | 0.031 |
| Honduras | HND.1_1 | Atlántida | 69 | 1.88 | 0.609 | 0.065 |
| Honduras | HND.2_1 | Choluteca | 72 | 0 | | 0 |
| Honduras | HND.3_1 | Colón | 81 | 0 | | 0 |
| Honduras | HND.4_1 | Comayagua | 95 | 2.16 | 0.042 | 0 |
| Honduras | HND.5_1 | Copán | 84 | 0 | | 0 |
| Honduras | HND.6_1 | Cortés | 152 | 1.26 | 0.914 | 0.051 |
| Honduras | HND.7_1 | El Paraíso | 73 | 0 | | 0 |
| Honduras | HND.8_1 | Francisco Morazán | 147 | 0.15 | 0.789 | 0.007 |
| Honduras | HND.9_1 | Gracias a Dios | 91 | 2.63 | 0.337 | 0.059 |
| India | IND.10_1 | Goa | 68 | 2.11 | 0.279 | 0 |
| India | IND.11_1 | Gujarat | 186 | 7.35 | 0.242 | 0.084 |

| Country | adm1_gid | adm1_name | N | Zero-dose Prevalence | CI | AEG |
|---|---|---|---|---|---|---|
| | | | 3 | | | |
| India | IND.12_1 | Haryana | 1310 | 5.77 | 0.203 | 0.049 |
| India | IND.13_1 | Himachal Pradesh | 531 | 1.82 | -0.012 | -0.011 |
| India | IND.15_1 | Jharkhand | 1923 | 7.5 | 0.178 | 0.098 |
| India | IND.16_1 | Karnataka | 1584 | 4.06 | 0.129 | 0.008 |
| India | IND.17_1 | Kerala | 486 | 4.04 | 0.061 | 0.026 |
| India | IND.18_1 | Lakshadweep | 45 | 9.02 | 0.009 | 0.144 |
| India | IND.19_1 | Madhya Pradesh | 3102 | 5.72 | 0.173 | 0.058 |
| India | IND.1_1 | Andaman and Nicobar | 94 | 2.21 | -0.109 | 0.011 |
| India | IND.20_1 | Maharashtra | 1811 | 7.53 | 0.221 | 0.108 |
| India | IND.21_1 | Manipur | 566 | 6.51 | 0.221 | 0.068 |
| India | IND.22_1 | Meghalaya | 1125 | 16.31 | 0.347 | 0.203 |
| India | IND.23_1 | Mizoram | 484 | 14.29 | 0.305 | 0.133 |
| India | IND.24_1 | Nagaland | 556 | 15.23 | 0.145 | 0.062 |
| India | IND.25_1 | NCT of Delhi | 570 | 6.32 | 0.433 | 0.13 |
| India | IND.26_1 | Odisha | 1565 | 2.68 | 0.135 | 0.019 |
| India | IND.27_1 | Puducherry | 157 | 0.63 | 0.121 | 0.003 |
| India | IND.28_1 | Punjab | 1052 | 5.69 | 0.245 | 0.07 |
| India | IND.29_1 | Rajasthan | 2544 | 5.24 | 0.175 | 0.043 |
| India | IND.2_1 | Andhra Pradesh | 540 | 6.79 | 0.184 | 0.054 |
| India | IND.30_1 | Sikkim | 114 | 4.9 | 0.195 | 0.011 |
| India | IND.31_1 | Tamil Nadu | 1289 | 2.42 | 0.093 | 0.013 |
| India | IND.32_1 | Telangana | 1441 | 7.4 | 0.116 | 0.052 |
| India | IND.33_1 | Tripura | 390 | 5.28 | 0.11 | 0.031 |
| India | IND.34_1 | Uttar Pradesh | 6540 | 8.93 | 0.178 | 0.079 |

| Country | adm1_gid | adm1_name | N | Zero-dose Prevalence | CI | AEG |
|---|---|---|---|---|---|---|
| India | IND.35_1 | Uttarakhand | 712 | 4.61 | 0.28 | 0.025 |
| India | IND.36_1 | West Bengal | 1105 | 2.09 | 0.407 | 0.052 |
| India | IND.3_1 | Arunachal Pradesh | 965 | 13.06 | 0.242 | 0.158 |
| India | IND.4_1 | Assam | 1961 | 9.05 | 0.152 | 0.07 |
| India | IND.5_1 | Bihar | 3963 | 6.14 | 0.196 | 0.054 |
| India | IND.6_1 | Chandigarh | 28 | 6.58 | 0.571 | 0.1 |
| India | IND.7_1 | Chhattisgarh | 1599 | 4.61 | 0.193 | 0.045 |
| India | IND.8_1 | Dadra and Nagar Haveli | 151 | 2.15 | 0.795 | 0.071 |
| India | IND.9_1 | Daman and Diu | 151 | 2.15 | 0.795 | 0.071 |
| Indonesia | IDN.10_1 | Jawa Tengah | 218 | 2.95 | 0.197 | 0.021 |
| Indonesia | IDN.11_1 | Jawa Timur | 216 | 5.84 | 0.47 | 0.097 |
| Indonesia | IDN.12_1 | Kalimantan Barat | 66 | 8.39 | 0.025 | -0.022 |
| Indonesia | IDN.13_1 | Kalimantan Selatan | 48 | 7.27 | -0.083 | -0.02 |
| Indonesia | IDN.14_1 | Kalimantan Tengah | 39 | 15.69 | -0.088 | 0.024 |
| Indonesia | IDN.16_1 | Kepulauan Riau | 78 | 7.66 | 0.175 | 0.107 |
| Indonesia | IDN.17_1 | Lampung | 80 | 3.21 | -0.254 | -0.049 |
| Indonesia | IDN.18_1 | Maluku Utara | 81 | 11.53 | 0.388 | 0.209 |
| Indonesia | IDN.19_1 | Maluku | 176 | 17.74 | 0.106 | 0.098 |
| Indonesia | IDN.1_1 | Aceh | 191 | 35.42 | 0.104 | 0.194 |
| Indonesia | IDN.20_1 | Nusa Tenggara Barat | 103 | 2.14 | -0.272 | -0.041 |
| Indonesia | IDN.21_1 | Nusa Tenggara Timur | 215 | 5.37 | 0.278 | 0.07 |
| Indonesia | IDN.22_1 | Papua Barat | 47 | 17.4 | 0.355 | 0.28 |
| Indonesia | IDN.23_1 | Papua | 58 | 21.15 | 0.2 | 0.199 |
| Indonesia | IDN.24_1 | Riau | 73 | 37.98 | 0.129 | 0.137 |
| Indonesia | IDN.25_1 | Sulawesi Barat | 118 | 13.98 | 0.216 | 0.18 |
| Indonesia | IDN.26_1 | Sulawesi Selatan | 113 | 7.39 | 0.265 | 0.074 |
| Indonesia | IDN.27_1 | Sulawesi Tengah | 86 | 12.94 | 0.493 | 0.345 |
| Indonesia | IDN.28_1 | Sulawesi Tenggara | 122 | 7.57 | 0.342 | 0.132 |
| Indonesia | IDN.29_1 | Sulawesi Utara | 41 | 0 | | 0 |
| Indonesia | IDN.2_1 | Bali | 36 | 2.57 | 0.639 | 0.104 |
| Indonesia | IDN.30_1 | Sumatera Barat | 72 | 25.67 | 0.144 | 0.301 |
| Indonesia | IDN.31_1 | Sumatera Selatan | 82 | 9.15 | 0.409 | 0.2 |
| Indonesia | IDN.32_1 | Sumatera Utara | 178 | 19.29 | 0.329 | 0.337 |

| Country | adm1_gid | adm1_name | N | Zero-dose Prevalence | CI | AEG |
|---|---|---|---|---|---|---|
| Indonesia | IDN.33_1 | Yogyakarta | 38 | 4.05 | 0.474 | 0.081 |
| Indonesia | IDN.34_1 | Kalimantan Timur | 97 | 8.63 | 0.279 | 0.022 |
| Indonesia | IDN.35_1 | Kalimantan Utara | 49 | 4.23 | -0.061 | 0 |
| Indonesia | IDN.3_1 | Bangka Belitung | 62 | 1.62 | 0.919 | 0.074 |
| Indonesia | IDN.4_1 | Banten | 116 | 22.07 | 0.251 | 0.319 |
| Indonesia | IDN.5_1 | Bengkulu | 45 | 7.22 | 0.059 | -0.107 |
| Indonesia | IDN.6_1 | Gorontalo | 53 | 9.08 | 0.091 | 0.017 |
| Indonesia | IDN.7_1 | Jakarta Raya | 120 | 4.54 | 0.102 | 0.054 |
| Indonesia | IDN.8_1 | Jambi | 42 | 9.23 | 0.345 | 0.147 |
| Indonesia | IDN.9_1 | Jawa Barat | 352 | 12.25 | 0.31 | 0.213 |
| Iraq | IRQ.10_1 | Baghdad | 306 | 2.36 | 0.358 | 0.059 |
| Iraq | IRQ.11_1 | Dhi-Qar | 147 | 10.83 | 0.057 | 0.061 |
| Iraq | IRQ.12_1 | Dihok | 152 | 2.52 | 0.507 | 0.07 |
| Iraq | IRQ.13_1 | Diyala | 176 | 6.12 | 0.078 | 0.052 |
| Iraq | IRQ.14_1 | Karbala' | 176 | 4.22 | 0.376 | 0.088 |
| Iraq | IRQ.15_1 | Maysan | 211 | 3.87 | 0.33 | 0.087 |
| Iraq | IRQ.16_1 | Ninawa | 147 | 7.76 | 0.454 | 0.154 |
| Iraq | IRQ.17_1 | Sala ad-Din | 84 | 7.93 | -0.005 | -0.047 |
| Iraq | IRQ.18_1 | Wasit | 131 | 2.23 | 0.598 | 0.116 |
| Iraq | IRQ.1_1 | Al-Anbar | 98 | 23.53 | 0.12 | 0.081 |
| Iraq | IRQ.2_1 | Al-Basrah | 200 | 1.9 | 0.2 | 0.021 |
| Iraq | IRQ.3_1 | Al-Muthannia | 185 | 11.04 | 0.138 | 0.072 |
| Iraq | IRQ.4_1 | Al-Qadisiyah | 135 | 10.95 | 0.161 | 0.124 |
| Iraq | IRQ.5_1 | An-Najaf | 137 | 12.09 | 0.021 | 0.069 |
| Iraq | IRQ.6_1 | Arbil | 99 | 0.82 | 0.03 | -0.009 |
| Iraq | IRQ.7_1 | As-Sulaymaniyah | 82 | 1.55 | 0.841 | 0.05 |
| Iraq | IRQ.8_1 | At-Ta'mim | 63 | 8.41 | 0.048 | -0.046 |
| Iraq | IRQ.9_1 | Babil | 148 | 6.33 | 0.616 | 0.174 |
| Jamaica | JAM.10_1 | Saint James | 33 | 0 | | |
| Jamaica | JAM.11_1 | Saint Mary | 11 | 0 | | |
| Jamaica | JAM.12_1 | Saint Thomas | 9 | 0 | | |
| Jamaica | JAM.13_1 | Trelawny | 8 | 0 | | |
| Jamaica | JAM.14_1 | Westmoreland | 5 | 0 | | |
| Jamaica | JAM.1_1 | Clarendon | 20 | 0 | | |
| Jamaica | JAM.2_1 | Hanover | 4 | 0 | | |
| Jamaica | JAM.3_1 | Kingston | 17 | 0 | | |
| Jamaica | JAM.4_1 | Manchester | 16 | 0 | | |

| Country | adm1_gid | adm1_name | N | Zero-dose Prevalence | CI | AEG |
|---|---|---|---|---|---|---|
| Jamaica | JAM.5_1 | Portland | 7 | 0 | | |
| Jamaica | JAM.6_1 | Saint Andrew | 42 | 0 | | |
| Jamaica | JAM.7_1 | Saint Ann | 40 | 0 | | |
| Jamaica | JAM.8_1 | Saint Catherine | 47 | 0 | | |
| Jamaica | JAM.9_1 | Saint Elizabeth | 10 | 0 | | |
| Jordan | JOR.10_1 | Mafraq | 183 | 2.64 | -0.426 | 0 |
| Jordan | JOR.11_1 | Tafilah | 96 | 4.59 | 0.354 | 0.054 |
| Jordan | JOR.12_1 | Zarqa | 230 | 4.69 | 0.089 | 0.081 |
| Jordan | JOR.1_1 | Ajlun | 142 | 2.07 | 0.167 | 0.029 |
| Jordan | JOR.2_1 | Amman | 199 | 1.6 | 0.663 | 0.088 |
| Jordan | JOR.3_1 | Aqaba | 68 | 7.25 | 0.315 | 0.184 |
| Jordan | JOR.4_1 | Balqa | 89 | 0.6 | 0.921 | 0.033 |
| Jordan | JOR.5_1 | Irbid | 237 | 0.99 | 0.241 | 0 |
| Jordan | JOR.6_1 | Jarash | 148 | 1.71 | 0.166 | 0.002 |
| Jordan | JOR.7_1 | Karak | 85 | 1.29 | -0.776 | -0.065 |
| Jordan | JOR.8_1 | Ma`an | 81 | 12.17 | 0.268 | 0.177 |
| Jordan | JOR.9_1 | Madaba | 86 | 0.9 | 0.547 | 0 |
| Kenya | KEN.10_1 | Kajiado | 72 | 4.77 | 0.108 | 0.031 |
| Kenya | KEN.11_1 | Kakamega | 88 | 0 | | 0 |
| Kenya | KEN.12_1 | Kericho | 79 | 4.62 | 0.574 | 0.131 |
| Kenya | KEN.13_1 | Kiambu | 63 | 0 | | 0 |
| Kenya | KEN.14_1 | Kilifi | 66 | 0 | | 0 |
| Kenya | KEN.15_1 | Kirinyaga | 54 | 0 | | 0 |
| Kenya | KEN.16_1 | Kisii | 60 | 0.9 | -0.55 | 0 |
| Kenya | KEN.17_1 | Kisumu | 82 | 0 | | 0 |
| Kenya | KEN.18_1 | Kitui | 73 | 7.13 | 0.142 | 0.025 |
| Kenya | KEN.19_1 | Kwale | 84 | 3.25 | -0.337 | -0.027 |
| Kenya | KEN.1_1 | Baringo | 78 | 0 | | 0 |
| Kenya | KEN.20_1 | Laikipia | 61 | 2.93 | -0.098 | 0 |
| Kenya | KEN.21_1 | Lamu | 86 | 0 | | 0 |
| Kenya | KEN.22_1 | Machakos | 47 | 2.38 | 0.383 | 0 |
| Kenya | KEN.23_1 | Makueni | 69 | 2.25 | 0.812 | 0.08 |
| Kenya | KEN.24_1 | Mandera | 160 | 36.2 | 0.03 | 0.11 |
| Kenya | KEN.25_1 | Marsabit | 97 | 6.76 | 0.005 | -0.056 |
| Kenya | KEN.26_1 | Meru | 55 | 1.72 | 0.727 | 0.062 |
| Kenya | KEN.27_1 | Migori | 102 | 0.44 | 0.245 | 0 |
| Kenya | KEN.28_1 | Mombasa | 57 | 0 | | 0 |

| Country | adm1_gid | adm1_name | N | Zero-dose Prevalence | CI | AEG |
|---|---|---|---|---|---|---|
| Kenya | KEN.29_1 | Murang'a | 42 | 3.29 | 0.357 | 0 |
| Kenya | KEN.2_1 | Bomet | 81 | 1.47 | 0.963 | 0.047 |
| Kenya | KEN.30_1 | Nairobi | 94 | 2.65 | 0.613 | 0.042 |
| Kenya | KEN.31_1 | Nakuru | 88 | 0.92 | 0.807 | 0.049 |
| Kenya | KEN.32_1 | Nandi | 57 | 0 | | 0 |
| Kenya | KEN.33_1 | Narok | 97 | 2.52 | 0.763 | 0.051 |
| Kenya | KEN.34_1 | Nyamira | 44 | 0 | | 0 |
| Kenya | KEN.35_1 | Nyandarua | 65 | 0 | | 0 |
| Kenya | KEN.36_1 | Nyeri | 39 | 5.35 | 0.179 | 0 |
| Kenya | KEN.37_1 | Samburu | 108 | 9.15 | 0.383 | 0.07 |
| Kenya | KEN.38_1 | Siaya | 73 | 0.68 | -0.63 | -0.067 |
| Kenya | KEN.39_1 | Taita Taveta | 51 | 1.55 | -0.824 | -0.094 |
| Kenya | KEN.3_1 | Bungoma | 90 | 1.33 | -0.144 | 0 |
| Kenya | KEN.40_1 | Tana River | 109 | 6.46 | 0.409 | 0.099 |
| Kenya | KEN.41_1 | Tharaka-Nithi | 50 | 0 | | 0 |
| Kenya | KEN.42_1 | Trans Nzoia | 64 | 0 | | 0 |
| Kenya | KEN.43_1 | Turkana | 128 | 3.37 | 0.389 | 0.051 |
| Kenya | KEN.44_1 | Uasin Gishu | 77 | 1.38 | 0.312 | 0 |
| Kenya | KEN.45_1 | Vihiga | 59 | 0 | | 0 |
| Kenya | KEN.46_1 | Wajir | 115 | 5.44 | -0.02 | -0.022 |
| Kenya | KEN.47_1 | West Pokot | 147 | 1.93 | 0.15 | 0.022 |
| Kenya | KEN.4_1 | Busia | 73 | 0 | | 0 |
| Kenya | KEN.5_1 | Elgeyo-Marakwet | 81 | 1.23 | 0.272 | 0 |
| Kenya | KEN.6_1 | Embu | 47 | 0 | | 0 |
| Kenya | KEN.7_1 | Garissa | 95 | 36.74 | -0.117 | -0.214 |
| Kenya | KEN.8_1 | Homa Bay | 72 | 2.53 | -0.653 | -0.101 |
| Kenya | KEN.9_1 | Isiolo | 93 | 3.24 | 0.355 | 0.086 |
| Kiribati | | | 225 | 0 | | |
| Kyrgyzstan | KGZ.1_1 | Batken | 62 | 13.42 | 0.058 | -0.043 |
| Kyrgyzstan | KGZ.2_1 | Biškek | 21 | 11.37 | 0.413 | 0.133 |
| Kyrgyzstan | KGZ.3_1 | Chüy | 27 | 3.69 | 0.889 | 0.132 |
| Kyrgyzstan | KGZ.4_1 | Jalal-Abad | 26 | 16.93 | 0.077 | 0.022 |
| Kyrgyzstan | KGZ.5_1 | Naryn | 46 | 23.33 | -0.073 | -0.237 |
| Kyrgyzstan | KGZ.6_1 | Osh (city) | 52 | 0 | | 0 |
| Kyrgyzstan | KGZ.7_1 | Osh | 66 | 4.57 | 0.823 | 0.135 |
| Kyrgyzstan | KGZ.8_1 | Talas | 40 | 5.58 | 0.625 | 0.194 |
| Kyrgyzstan | KGZ.9_1 | Ysyk-Köl | 27 | 0 | | 0 |

| Country | adm1_gid | adm1_name | N | Zero-dose Prevalence | CI | AEG |
|---|---|---|---|---|---|---|
| Laos | LAO.10_1 | Phôngsali | 134 | 4.81 | -0.103 | -0.043 |
| Laos | LAO.11_1 | Saravan | 60 | 4.83 | 0.316 | 0.098 |
| Laos | LAO.12_1 | Savannakhét | 47 | 20.87 | 0.472 | 0.458 |
| Laos | LAO.13_1 | Vientiane [prefecture] | 68 | 10.97 | 0.426 | 0.209 |
| Laos | LAO.14_1 | Vientiane | 63 | 11.62 | 0.127 | 0.042 |
| Laos | LAO.15_1 | Xaignabouri | 45 | 0 | | 0 |
| Laos | LAO.16_1 | Xaisômboun | 75 | 19.77 | 0.274 | 0.3 |
| Laos | LAO.17_1 | Xékong | 70 | 21.88 | 0.097 | 0.115 |
| Laos | LAO.18_1 | Xiangkhoang | 109 | 10.78 | 0.176 | 0.164 |
| Laos | LAO.1_1 | Attapu | 54 | 11.98 | 0.105 | -0.003 |
| Laos | LAO.2_1 | Bokeo | 83 | 5.06 | -0.084 | -0.018 |
| Laos | LAO.3_1 | Bolikhamxai | 85 | 0 | | 0 |
| Laos | LAO.4_1 | Champasak | 76 | 3.03 | 0.25 | 0.078 |
| Laos | LAO.5_1 | Houaphan | 74 | 2.69 | 0.878 | 0.127 |
| Laos | LAO.6_1 | Khammouan | 68 | 1.2 | 0.221 | 0 |
| Laos | LAO.7_1 | Louang Namtha | 80 | 2.66 | 0.075 | 0 |
| Laos | LAO.8_1 | Louangphrabang | 94 | 7.11 | 0.047 | 0 |
| Laos | LAO.9_1 | Oudômxai | 94 | 5.18 | 0.606 | 0.137 |
| Lesotho | LSO.10_1 | Thaba-Tseka | 74 | 2.29 | 0.243 | 0.028 |
| Lesotho | LSO.1_1 | Berea | 57 | 1.51 | 0.912 | 0.066 |
| Lesotho | LSO.2_1 | Butha-Buthe | 59 | 1.18 | 0.508 | 0.041 |
| Lesotho | LSO.3_1 | Leribe | 54 | 0 | | 0 |
| Lesotho | LSO.4_1 | Mafeteng | 31 | 0 | | 0 |
| Lesotho | LSO.5_1 | Maseru | 62 | 0 | | 0 |
| Lesotho | LSO.6_1 | Mohale's Hoek | 50 | 1.5 | 0.98 | 0.051 |
| Lesotho | LSO.7_1 | Mokhotlong | 50 | 0 | | 0 |
| Lesotho | LSO.8_1 | Qacha's Nek | 46 | 2.44 | 0.804 | 0.099 |
| Lesotho | LSO.9_1 | Quthing | 40 | 0 | | 0 |
| Liberia | LBR.2_1 | Bong | 288 | 7.9 | 0.271 | 0.093 |
| Liberia | LBR.12_1 | Nimba | 288 | 7.9 | 0.271 | 0.093 |
| Liberia | LBR.1_1 | Bomi | 171 | 6 | 0.387 | 0.158 |
| Liberia | LBR.4_1 | Grand Cape Mount | 171 | 6 | 0.387 | 0.158 |
| Liberia | LBR.3_1 | Gbapolu | 171 | 6 | 0.387 | 0.158 |
| Liberia | LBR.8_1 | Lofa | 171 | 6 | 0.387 | 0.158 |
| Liberia | LBR.11_1 | Montserrado | 240 | 9.19 | 0.348 | 0.286 |
| Liberia | LBR.9_1 | Margibi | 240 | 9.19 | 0.348 | 0.286 |
| Liberia | LBR.5_1 | Grand Bassa | 240 | 9.19 | 0.348 | 0.286 |

| Country | adm1_gid | adm1_name | N | Zero-dose Prevalence | CI | AEG |
|---|---|---|---|---|---|---|
| Liberia | LBR.6_1 | Grand Gedeh | 183 | 8.28 | 0.385 | 0.154 |
| Liberia | LBR.14_1 | River Gee | 183 | 8.28 | 0.385 | 0.154 |
| Liberia | LBR.7_1 | Grand Kru | 178 | 9.72 | 0.096 | 0.059 |
| Liberia | LBR.10_1 | Maryland | 178 | 9.72 | 0.096 | 0.059 |
| Liberia | LBR.13_1 | Rivercess | 178 | 9.72 | 0.096 | 0.059 |
| Liberia | LBR.15_1 | Sinoe | 178 | 9.72 | 0.096 | 0.059 |
| Madagascar | MDG.1_1 | Antananarivo | 510 | 6.81 | 0.107 | -0.004 |
| Madagascar | MDG.2_1 | Antsiranana | 180 | 30.88 | 0.274 | 0.348 |
| Madagascar | MDG.3_1 | Fianarantsoa | 560 | 14.04 | 0.344 | 0.207 |
| Madagascar | MDG.4_1 | Mahajanga | 345 | 35.61 | 0.292 | 0.505 |
| Madagascar | MDG.5_1 | Toamasina | 255 | 14.56 | 0.353 | 0.255 |
| Madagascar | MDG.6_1 | Toliary | 490 | 39.11 | 0.126 | 0.174 |
| Malawi | MWI.6_1 | Dedza | 959 | 0.85 | 0.58 | 0.025 |
| Malawi | MWI.7_1 | Dowa | 959 | 0.85 | 0.58 | 0.025 |
| Malawi | MWI.9_1 | Kasungu | 959 | 0.85 | 0.58 | 0.025 |
| Malawi | MWI.11_1 | Lilongwe | 959 | 0.85 | 0.58 | 0.025 |
| Malawi | MWI.14_1 | Mchinji | 959 | 0.85 | 0.58 | 0.025 |
| Malawi | MWI.20_1 | Nkhotakota | 959 | 0.85 | 0.58 | 0.025 |
| Malawi | MWI.22_1 | Ntcheu | 959 | 0.85 | 0.58 | 0.025 |
| Malawi | MWI.23_1 | Ntchisi | 959 | 0.85 | 0.58 | 0.025 |
| Malawi | MWI.26_1 | Salima | 959 | 0.85 | 0.58 | 0.025 |
| Malawi | MWI.5_1 | Chitipa | 613 | 0.33 | 0.294 | 0.006 |
| Malawi | MWI.8_1 | Karonga | 613 | 0.33 | 0.294 | 0.006 |
| Malawi | MWI.17_1 | Mzimba | 613 | 0.33 | 0.294 | 0.006 |
| Malawi | MWI.19_1 | Nkhata Bay | 613 | 0.33 | 0.294 | 0.006 |
| Malawi | MWI.25_1 | Rumphi | 613 | 0.33 | 0.294 | 0.006 |
| Malawi | MWI.10_1 | Likoma | 613 | 0.33 | 0.294 | 0.006 |
| Malawi | MWI.1_1 | Balaka | 1389 | 1.63 | 0.227 | 0.025 |
| Malawi | MWI.2_1 | Blantyre | 1389 | 1.63 | 0.227 | 0.025 |
| Malawi | MWI.3_1 | Chikwawa | 1389 | 1.63 | 0.227 | 0.025 |
| Malawi | MWI.4_1 | Chiradzulu | 1389 | 1.63 | 0.227 | 0.025 |
| Malawi | MWI.12_1 | Machinga | 1389 | 1.63 | 0.227 | 0.025 |

| Country | adm1_gid | adm1_name | N | Zero-dose Prevalence | CI | AEG |
|---|---|---|---|---|---|---|
| Malawi | MWI.13_1 | Mangochi | 1389 | 1.63 | 0.227 | 0.025 |
| Malawi | MWI.15_1 | Mulanje | 1389 | 1.63 | 0.227 | 0.025 |
| Malawi | MWI.16_1 | Mwanza | 1389 | 1.63 | 0.227 | 0.025 |
| Malawi | MWI.18_1 | Neno | 1389 | 1.63 | 0.227 | 0.025 |
| Malawi | MWI.21_1 | Nsanje | 1389 | 1.63 | 0.227 | 0.025 |
| Malawi | MWI.24_1 | Phalombe | 1389 | 1.63 | 0.227 | 0.025 |
| Malawi | MWI.27_1 | Thyolo | 1389 | 1.63 | 0.227 | 0.025 |
| Malawi | MWI.28_1 | Zomba | 1389 | 1.63 | 0.227 | 0.025 |
| Maldives | | | 585 | 8.79 | | |
| Mali | MLI.1_1 | Bamako | 306 | 4.11 | 0.556 | 0.107 |
| Mali | MLI.2_1 | Gao | 196 | 32.37 | 0.195 | 0.366 |
| Mali | MLI.3_1 | Kayes | 458 | 24.68 | 0.134 | 0.164 |
| Mali | MLI.4_1 | Kidal | 43 | 51 | 0.209 | 0.358 |
| Mali | MLI.5_1 | Koulikoro | 469 | 14.09 | 0.139 | 0.102 |
| Mali | MLI.6_1 | Mopti | 339 | 20.21 | 0.24 | 0.222 |
| Mali | MLI.7_1 | Ségou | 358 | 12.46 | 0.19 | 0.157 |
| Mali | MLI.8_1 | Sikasso | 385 | 13.62 | 0.168 | 0.099 |
| Mali | MLI.9_1 | Timbuktu | 290 | 57.29 | 0.142 | 0.371 |
| Mauritania | MRT.10_1 | Nouakchott | 262 | 10.16 | 0.332 | 0.17 |
| Mauritania | MRT.11_1 | Tagant | 112 | 6.49 | 0.5 | 0.193 |
| Mauritania | MRT.13_1 | Trarza | 153 | 5.57 | 0.415 | 0.103 |
| Mauritania | MRT.1_1 | Adrar | 85 | 4.43 | 0.329 | 0.075 |
| Mauritania | MRT.2_1 | Assaba | 185 | 5.61 | 0.017 | 0.019 |
| Mauritania | MRT.3_1 | Brakna | 217 | 8.64 | 0.236 | 0.074 |
| Mauritania | MRT.4_1 | Dakhlet Nouadhibou | 87 | 1.57 | 0 | 0 |
| Mauritania | MRT.5_1 | Gorgol | 202 | 14.78 | 0.218 | 0.155 |
| Mauritania | MRT.6_1 | Guidimaka | 311 | 8.4 | 0.105 | 0.016 |
| Mauritania | MRT.7_1 | Hodh ech Chargui | 185 | 23.3 | 0.29 | 0.29 |
| Mauritania | MRT.8_1 | Hodh el Gharbi | 194 | 13.67 | 0.308 | 0.247 |

| Country | adm1_gid | adm1_name | N | Zero-dose Prevalence | CI | AEG |
|---|---|---|---|---|---|---|
| Mauritania | MRT.9_1 | Inchiri | 104 | 5.58 | 0.136 | 0.01 |
| Mauritania | MRT.12_1 | Tiris Zemmour | 104 | 5.58 | 0.136 | 0.01 |
| Mongolia | MNG.21_1 | Ulaanbaatar | 215 | 3.84 | -0.188 | 0.008 |
| Mongolia | MNG.5_1 | Darhan-Uul | 166 | 1.95 | -0.038 | 0 |
| Mongolia | MNG.7_1 | Dornogovi | 166 | 1.95 | -0.038 | 0 |
| Mongolia | MNG.8_1 | Dundgovi | 166 | 1.95 | -0.038 | 0 |
| Mongolia | MNG.11_1 | Govisümber | 166 | 1.95 | -0.038 | 0 |
| Mongolia | MNG.15_1 | Ömnögovi | 166 | 1.95 | -0.038 | 0 |
| Mongolia | MNG.18_1 | Selenge | 166 | 1.95 | -0.038 | 0 |
| Mongolia | MNG.20_1 | Töv | 166 | 1.95 | -0.038 | 0 |
| Mongolia | MNG.6_1 | Dornod | 127 | 3.33 | 0.217 | 0.046 |
| Mongolia | MNG.12_1 | Hentiy | 127 | 3.33 | 0.217 | 0.046 |
| Mongolia | MNG.19_1 | Sühbaatar | 127 | 3.33 | 0.217 | 0.046 |
| Mongolia | MNG.1_1 | Arhangay | 183 | 2.6 | 0.129 | 0.001 |
| Mongolia | MNG.3_1 | Bayanhongor | 183 | 2.6 | 0.129 | 0.001 |
| Mongolia | MNG.4_1 | Bulgan | 183 | 2.6 | 0.129 | 0.001 |
| Mongolia | MNG.14_1 | Hövsgöl | 183 | 2.6 | 0.129 | 0.001 |
| Mongolia | MNG.16_1 | Orhon | 183 | 2.6 | 0.129 | 0.001 |
| Mongolia | MNG.17_1 | Övörhangay | 183 | 2.6 | 0.129 | 0.001 |
| Mongolia | MNG.2_1 | Bayan-Ölgiy | 225 | 11.12 | 0.209 | 0.131 |
| Mongolia | MNG.10_1 | Govi-Altay | 225 | 11.12 | 0.209 | 0.131 |
| Mongolia | MNG.22_1 | Uvs | 225 | 11.12 | 0.209 | 0.131 |
| Mongolia | MNG.13_1 | Hovd | 225 | 11.12 | 0.209 | 0.131 |
| Mongolia | MNG.9_1 | Dzavhan | 225 | 11.12 | 0.209 | 0.131 |
| Mozambique | MOZ.10_1 | Tete | 162 | 26.57 | 0.266 | 0.322 |
| Mozambique | MOZ.11_1 | Zambezia | 132 | 48.17 | 0.181 | 0.365 |
| Mozambique | MOZ.1_1 | Cabo Delgado | 212 | 17.52 | 0.209 | 0.173 |
| Mozambique | MOZ.2_1 | Gaza | 135 | 1.15 | 0.6 | 0.021 |
| Mozambique | MOZ.3_1 | Inhambane | 109 | 2.71 | -0.147 | 0 |
| Mozambique | MOZ.4_1 | Manica | 172 | 8.27 | 0.339 | 0.094 |
| Mozambique | MOZ.5_1 | Maputo City | 82 | 0 |  | 0 |
| Mozambique | MOZ.6_1 | Maputo | 79 | 0 |  | 0 |
| Mozambique | MOZ.7_1 | Nampula | 213 | 20.18 | 0.342 | 0.323 |
| Mozambique | MOZ.8_1 | Nassa | 215 | 16.55 | 0.413 | 0.307 |
| Mozambique | MOZ.9_1 | Sofala | 167 | 7.39 | 0.453 | 0.168 |
| Myanmar | MMR.10_1 | Naypyitaw | 46 | 9.01 | 0.185 | 0 |
| Myanmar | MMR.11_1 | Rakhine | 79 | 9.45 | 0.282 | 0.09 |

| Country | adm1_gid | adm1_name | N | Zero-dose Prevalence | CI | AEG |
|---|---|---|---|---|---|---|
| Myanmar | MMR.12_1 | Sagaing | 58 | 13.53 | 0.478 | 0.446 |
| Myanmar | MMR.13_1 | Shan | 72 | 25.31 | 0.523 | 0.615 |
| Myanmar | MMR.14_1 | Tanintharyi | 56 | 1.86 | 0.232 | 0 |
| Myanmar | MMR.15_1 | Yangon | 55 | 3.58 | 0.745 | 0.172 |
| Myanmar | MMR.1_1 | Ayeyarwady | 68 | 21.73 | 0.421 | 0.274 |
| Myanmar | MMR.2_1 | Bago | 57 | 16.57 | 0.351 | 0.166 |
| Myanmar | MMR.3_1 | Chin | 83 | 8.5 | 0.231 | 0.101 |
| Myanmar | MMR.4_1 | Kachin | 60 | 1.61 | 0.183 | 0 |
| Myanmar | MMR.5_1 | Kayah | 66 | 0 |  | 0 |
| Myanmar | MMR.6_1 | Kayin | 66 | 13.13 | 0.227 | 0.224 |
| Myanmar | MMR.7_1 | Magway | 48 | 6.66 | 0.271 | 0.083 |
| Myanmar | MMR.8_1 | Mandalay | 56 | 6.58 | -0.116 | -0.041 |
| Myanmar | MMR.9_1 | Mon | 44 | 4.61 | 0.182 | 0.101 |
| Nauru | NRU.11_1 | Meneng | 2 | 0 |  |  |
| Nauru | NRU.12_1 | Nibok | 3 | 0 |  |  |
| Nauru | NRU.13_1 | Uaboe | 1 | 0 |  |  |
| Nauru | NRU.14_1 | Yaren | 3 | 0 |  |  |
| Nauru | NRU.1_1 | Aiwo | 2 | 0 |  |  |
| Nauru | NRU.2_1 | Anabar | 2 | 0 |  |  |
| Nauru | NRU.3_1 | Anetan | 2 | 0 |  |  |
| Nauru | NRU.4_1 | Anibare | 1 | 0 |  |  |
| Nauru | NRU.5_1 | Baiti | 2 | 0 |  |  |
| Nauru | NRU.6_1 | Boe | 2 | 0 |  |  |
| Nauru | NRU.7_1 | Buada | 3 | 0 |  |  |
| Nauru | NRU.9_1 | Ewa | 1 | 0 |  |  |
| Nepal | NPL.1_1 | Central | 327 | 6.76 | 0.338 | 0.107 |
| Nepal | NPL.2_1 | East | 145 | 5.73 | 0.481 | 0.142 |
| Nepal | NPL.3_1 | Far-Western | 140 | 2.25 | 0.155 | 0 |
| Nepal | NPL.4_1 | Mid-Western | 172 | 4.04 | 0.004 | 0.029 |
| Nepal | NPL.5_1 | West | 212 | 2.6 | 0.229 | 0.009 |
| Nigeria | NGA.7_1 | Benue | 866 | 39.4 | 0.366 | 0.664 |
| Nigeria | NGA.15_1 | Federal Capital Territory | 866 | 39.4 | 0.366 | 0.664 |
| Nigeria | NGA.23_1 | Kogi | 866 | 39.4 | 0.366 | 0.664 |
| Nigeria | NGA.24_1 | Kwara | 866 | 39.4 | 0.366 | 0.664 |
| Nigeria | NGA.26_1 | Nasarawa | 866 | 39.4 | 0.366 | 0.664 |
| Nigeria | NGA.27_1 | Niger | 866 | 39.4 | 0.366 | 0.664 |

| Country | adm1_gid | adm1_name | N | Zero-dose Prevalence | CI | AEG |
|---|---|---|---|---|---|---|
| Nigeria | NGA.32_1 | Plateau | 866 | 39.4 | 0.366 | 0.664 |
| Nigeria | NGA.2_1 | Adamawa | 1030 | 35.64 | 0.181 | 0.306 |
| Nigeria | NGA.5_1 | Bauchi | 1030 | 35.64 | 0.181 | 0.306 |
| Nigeria | NGA.8_1 | Borno | 1030 | 35.64 | 0.181 | 0.306 |
| Nigeria | NGA.16_1 | Gombe | 1030 | 35.64 | 0.181 | 0.306 |
| Nigeria | NGA.35_1 | Taraba | 1030 | 35.64 | 0.181 | 0.306 |
| Nigeria | NGA.36_1 | Yobe | 1030 | 35.64 | 0.181 | 0.306 |
| Nigeria | NGA.18_1 | Jigawa | 1476 | 53.74 | 0.169 | 0.425 |
| Nigeria | NGA.19_1 | Kaduna | 1476 | 53.74 | 0.169 | 0.425 |
| Nigeria | NGA.20_1 | Kano | 1476 | 53.74 | 0.169 | 0.425 |
| Nigeria | NGA.21_1 | Katsina | 1476 | 53.74 | 0.169 | 0.425 |
| Nigeria | NGA.22_1 | Kebbi | 1476 | 53.74 | 0.169 | 0.425 |
| Nigeria | NGA.34_1 | Sokoto | 1476 | 53.74 | 0.169 | 0.425 |
| Nigeria | NGA.37_1 | Zamfara | 1476 | 53.74 | 0.169 | 0.425 |
| Nigeria | NGA.1_1 | Abia | 544 | 6.93 | 0.383 | 0.096 |
| Nigeria | NGA.4_1 | Anambra | 544 | 6.93 | 0.383 | 0.096 |
| Nigeria | NGA.11_1 | Ebonyi | 544 | 6.93 | 0.383 | 0.096 |
| Nigeria | NGA.14_1 | Enugu | 544 | 6.93 | 0.383 | 0.096 |
| Nigeria | NGA.17_1 | Imo | 544 | 6.93 | 0.383 | 0.096 |
| Nigeria | NGA.3_1 | Akwa Ibom | 523 | 12.21 | 0.439 | 0.231 |
| Nigeria | NGA.6_1 | Bayelsa | 523 | 12.21 | 0.439 | 0.231 |
| Nigeria | NGA.9_1 | Cross River | 523 | 12.21 | 0.439 | 0.231 |
| Nigeria | NGA.10_1 | Delta | 523 | 12.21 | 0.439 | 0.231 |
| Nigeria | NGA.12_1 | Edo | 523 | 12.21 | 0.439 | 0.231 |

| Country | adm1_gid | adm1_name | N | Zero-dose Prevalence | CI | AEG |
|---|---|---|---|---|---|---|
| Nigeria | NGA.33_1 | Rivers | 523 | 12.21 | 0.439 | 0.231 |
| Nigeria | NGA.13_1 | Ekiti | 475 | 13.9 | 0.559 | 0.448 |
| Nigeria | NGA.25_1 | Lagos | 475 | 13.9 | 0.559 | 0.448 |
| Nigeria | NGA.28_1 | Ogun | 475 | 13.9 | 0.559 | 0.448 |
| Nigeria | NGA.29_1 | Ondo | 475 | 13.9 | 0.559 | 0.448 |
| Nigeria | NGA.30_1 | Osun | 475 | 13.9 | 0.559 | 0.448 |
| Nigeria | NGA.31_1 | Oyo | 475 | 13.9 | 0.559 | 0.448 |
| North Macedonia | MKD.3_1 | Berovo | 30 | 0 | | 0 |
| North Macedonia | MKD.14_1 | Češinovo-Obleševo | 30 | 0 | | 0 |
| North Macedonia | MKD.18_1 | Delčevo | 30 | 0 | | 0 |
| North Macedonia | MKD.30_1 | Karbinci | 30 | 0 | | 0 |
| North Macedonia | MKD.35_1 | Kočani | 30 | 0 | | 0 |
| North Macedonia | MKD.45_1 | Makedonska Kamenica | 30 | 0 | | 0 |
| North Macedonia | MKD.54_1 | Pehčevo | 30 | 0 | | 0 |
| North Macedonia | MKD.58_1 | Probištip | 30 | 0 | | 0 |
| North Macedonia | MKD.67_1 | Štip | 30 | 0 | | 0 |
| North Macedonia | MKD.79_1 | Vinitsa | 30 | 0 | | 0 |
| North Macedonia | MKD.85_1 | Zrnovci | 30 | 0 | | 0 |
| North Macedonia | MKD.37_1 | Kratovo | 42 | 0 | | 0 |
| North Macedonia | MKD.38_1 | Kriva Palanka | 42 | 0 | | 0 |
| North Macedonia | MKD.41_1 | Kumanovo | 42 | 0 | | 0 |
| North Macedonia | MKD.43_1 | Lipkovo | 42 | 0 | | 0 |
| North Macedonia | MKD.60_1 | Rankovce | 42 | 0 | | 0 |
| North Macedonia | MKD.66_1 | Staro Nagoričane | 42 | 0 | | 0 |
| North Macedonia | MKD.4_1 | Bitola | 32 | 1.51 | 0.969 | 0.101 |
| North Macedonia | MKD.19_1 | Demir Hisar | 32 | 1.51 | 0.969 | 0.101 |
| North Macedonia | MKD.21_1 | Dolneni | 32 | 1.51 | 0.969 | 0.101 |
| North Macedonia | MKD.39_1 | Krivogaštani | 32 | 1.51 | 0.969 | 0.101 |
| North Macedonia | MKD.40_1 | Kruševo | 32 | 1.51 | 0.969 | 0.101 |
| North Macedonia | MKD.48_1 | Mogila | 32 | 1.51 | 0.969 | 0.101 |
| North Macedonia | MKD.50_1 | Novatsi | 32 | 1.51 | 0.969 | 0.101 |
| North Macedonia | MKD.57_1 | Prilep | 32 | 1.51 | 0.969 | 0.101 |
| North Macedonia | MKD.61_1 | Resen | 32 | 1.51 | 0.969 | 0.101 |
| North Macedonia | MKD.6_1 | Bogovinje | 35 | 4.25 | 0.4 | 0.16 |
| North Macedonia | MKD.8_1 | Brvenica | 35 | 4.25 | 0.4 | 0.16 |
| North Macedonia | MKD.26_1 | Gostivar | 35 | 4.25 | 0.4 | 0.16 |
| North Macedonia | MKD.29_1 | Jegunovtse | 35 | 4.25 | 0.4 | 0.16 |

| Country | adm1_gid | adm1_name | N | Zero-dose Prevalence | CI | AEG |
|---|---|---|---|---|---|---|
| North Macedonia | MKD.47_1 | Mavrovo and Rostuša | 35 | 4.25 | 0.4 | 0.16 |
| North Macedonia | MKD.73_1 | Tearce | 35 | 4.25 | 0.4 | 0.16 |
| North Macedonia | MKD.74_1 | Tetovo | 35 | 4.25 | 0.4 | 0.16 |
| North Macedonia | MKD.81_1 | Vrapčište | 35 | 4.25 | 0.4 | 0.16 |
| North Macedonia | MKD.84_1 | Želino | 35 | 4.25 | 0.4 | 0.16 |
| North Macedonia | MKD.1_1 | Aerodrom | 63 | 7.4 | 0.516 | 0.228 |
| North Macedonia | MKD.2_1 | Aracinovo | 63 | 7.4 | 0.516 | 0.228 |
| North Macedonia | MKD.9_1 | Butel | 63 | 7.4 | 0.516 | 0.228 |
| North Macedonia | MKD.10_1 | Čair | 63 | 7.4 | 0.516 | 0.228 |
| North Macedonia | MKD.12_1 | Centar | 63 | 7.4 | 0.516 | 0.228 |
| North Macedonia | MKD.15_1 | Čučer Sandevo | 63 | 7.4 | 0.516 | 0.228 |
| North Macedonia | MKD.23_1 | Gazi Baba | 63 | 7.4 | 0.516 | 0.228 |
| North Macedonia | MKD.25_1 | Gjorče Petrov | 63 | 7.4 | 0.516 | 0.228 |
| North Macedonia | MKD.28_1 | Ilinden | 63 | 7.4 | 0.516 | 0.228 |
| North Macedonia | MKD.31_1 | Karpoš | 63 | 7.4 | 0.516 | 0.228 |
| North Macedonia | MKD.34_1 | Kisela Vod | 63 | 7.4 | 0.516 | 0.228 |
| North Macedonia | MKD.55_1 | Petrovec | 63 | 7.4 | 0.516 | 0.228 |
| North Macedonia | MKD.63_1 | Saraj | 63 | 7.4 | 0.516 | 0.228 |
| North Macedonia | MKD.64_1 | Sopište | 63 | 7.4 | 0.516 | 0.228 |
| North Macedonia | MKD.70_1 | Studeničani | 63 | 7.4 | 0.516 | 0.228 |
| North Macedonia | MKD.71_1 | Šuto Orizari | 63 | 7.4 | 0.516 | 0.228 |
| North Macedonia | MKD.83_1 | Zelenikovo | 63 | 7.4 | 0.516 | 0.228 |
| North Macedonia | MKD.5_1 | Bogdanci | 21 | 0 | | 0 |
| North Macedonia | MKD.7_1 | Bosilovo | 21 | 0 | | 0 |
| North Macedonia | MKD.24_1 | Gevgelija | 21 | 0 | | 0 |
| North Macedonia | MKD.65_1 | Star Dojran | 21 | 0 | | 0 |
| North Macedonia | MKD.36_1 | Konče | 21 | 0 | | 0 |
| North Macedonia | MKD.51_1 | Novo Selo | 21 | 0 | | 0 |
| North Macedonia | MKD.59_1 | Radoviš | 21 | 0 | | 0 |
| North Macedonia | MKD.69_1 | Strumitsa | 21 | 0 | | 0 |
| North Macedonia | MKD.75_1 | Valandovo | 21 | 0 | | 0 |
| North Macedonia | MKD.76_1 | Vasilevo | 21 | 0 | | 0 |
| North Macedonia | MKD.13_1 | Centar župa | 35 | 1.24 | -0.629 | -0.056 |
| North Macedonia | MKD.16_1 | Debar | 35 | 1.24 | -0.629 | -0.056 |
| North Macedonia | MKD.17_1 | Debarca | 35 | 1.24 | -0.629 | -0.056 |
| North Macedonia | MKD.33_1 | Kičevo | 35 | 1.24 | -0.629 | -0.056 |
| North Macedonia | MKD.42_1 | Lake Ohrid | 35 | 1.24 | -0.629 | -0.056 |

| Country | adm1_gid | adm1_name | N | Zero-dose Prevalence | CI | AEG |
|---|---|---|---|---|---|---|
| North Macedonia | MKD.46_1 | Makedonski Brod | 35 | 1.24 | -0.629 | -0.056 |
| North Macedonia | MKD.52_1 | Ohrid | 35 | 1.24 | -0.629 | -0.056 |
| North Macedonia | MKD.56_1 | Plasnica | 35 | 1.24 | -0.629 | -0.056 |
| North Macedonia | MKD.68_1 | Struga | 35 | 1.24 | -0.629 | -0.056 |
| North Macedonia | MKD.78_1 | Vevčani | 35 | 1.24 | -0.629 | -0.056 |
| North Macedonia | MKD.11_1 | Čaška | 27 | 0 | | 0 |
| North Macedonia | MKD.20_1 | Demir Kapija | 27 | 0 | | 0 |
| North Macedonia | MKD.27_1 | Gradsko | 27 | 0 | | 0 |
| North Macedonia | MKD.32_1 | Kavadartsi | 27 | 0 | | 0 |
| North Macedonia | MKD.49_1 | Negotino | 27 | 0 | | 0 |
| North Macedonia | MKD.62_1 | Rosoman | 27 | 0 | | 0 |
| North Macedonia | MKD.72_1 | Sveti Nikole | 27 | 0 | | 0 |
| North Macedonia | MKD.44_1 | Lozovo | 27 | 0 | | 0 |
| North Macedonia | MKD.77_1 | Veles | 27 | 0 | | 0 |
| Pakistan | PAK.2_1 | Balochistan | 1903 | 5.12 | 0.091 | 0.047 |
| Pakistan | PAK.5_1 | Khyber-Pakhtunkhwa | 2809 | 1.8 | 0.193 | 0.014 |
| Pakistan | PAK.8_1 | Sindh | 1784 | 2.86 | 0.172 | 0.026 |
| Pakistan | PAK.7_1 | Punjab | 6999 | 0.83 | 0.313 | 0.013 |
| Pakistan | PAK.4_1 | Islamabad | 6999 | 0.83 | 0.313 | 0.013 |
| Papua New Guinea | PNG.3_1 | Chimbu | 415 | 43.53 | 0.194 | 0.525 |
| Papua New Guinea | PNG.6_1 | Eastern Highlands | 415 | 43.53 | 0.194 | 0.525 |
| Papua New Guinea | PNG.7_1 | Enga | 415 | 43.53 | 0.194 | 0.525 |
| Papua New Guinea | PNG.9_1 | Hela | 415 | 43.53 | 0.194 | 0.525 |
| Papua New Guinea | PNG.10_1 | Jiwaka | 415 | 43.53 | 0.194 | 0.525 |
| Papua New Guinea | PNG.19_1 | Southern Highlands | 415 | 43.53 | 0.194 | 0.525 |
| Papua New | PNG.21_1 | Western Highlands | 415 | 43.53 | 0.194 | 0.525 |

| Country | adm1_gid | adm1_name | N | Zero-dose Prevalence | CI | AEG |
|---|---|---|---|---|---|---|
| Guinea | | | | | | |
| Papua New Guinea | PNG.1_1 | Bougainville | 457 | 19.95 | 0.236 | 0.285 |
| Papua New Guinea | PNG.4_1 | East New Britain | 457 | 19.95 | 0.236 | 0.285 |
| Papua New Guinea | PNG.12_1 | Manus | 457 | 19.95 | 0.236 | 0.285 |
| Papua New Guinea | PNG.16_1 | New Ireland | 457 | 19.95 | 0.236 | 0.285 |
| Papua New Guinea | PNG.20_1 | West New Britain | 457 | 19.95 | 0.236 | 0.285 |
| Papua New Guinea | PNG.5_1 | East Sepik | 375 | 41.35 | 0.262 | 0.485 |
| Papua New Guinea | PNG.11_1 | Madang | 375 | 41.35 | 0.262 | 0.485 |
| Papua New Guinea | PNG.14_1 | Morobe | 375 | 41.35 | 0.262 | 0.485 |
| Papua New Guinea | PNG.18_1 | Sandaun | 375 | 41.35 | 0.262 | 0.485 |
| Papua New Guinea | PNG.2_1 | Central | 536 | 23.88 | 0.269 | 0.301 |
| Papua New Guinea | PNG.8_1 | Gulf | 536 | 23.88 | 0.269 | 0.301 |
| Papua New Guinea | PNG.13_1 | Milne Bay | 536 | 23.88 | 0.269 | 0.301 |
| Papua New Guinea | PNG.15_1 | National Capital District | 536 | 23.88 | 0.269 | 0.301 |
| Papua New Guinea | PNG.17_1 | Oro | 536 | 23.88 | 0.269 | 0.301 |
| Papua New Guinea | PNG.22_1 | Western | 536 | 23.88 | 0.269 | 0.301 |
| Philippines | PHL.47_1 | Metropolitan Manila | 107 | 11.35 | 0.246 | 0.315 |
| Philippines | PHL.9_1 | Basilan | 178 | 66.89 | 0.059 | 0.206 |
| Philippines | PHL.42_1 | Lanao del Sur | 178 | 66.89 | 0.059 | 0.206 |
| Philippines | PHL.44_1 | Maguindanao | 178 | 66.89 | 0.059 | 0.206 |
| Philippines | PHL.73_1 | Sulu | 178 | 66.89 | 0.059 | 0.206 |
| Philippines | PHL.77_1 | Tawi-Tawi | 178 | 66.89 | 0.059 | 0.206 |

| Country | adm1_gid | adm1_name | N | Zero-dose Prevalence | CI | AEG |
|---|---|---|---|---|---|---|
| Philippines | PHL.1_1 | Abra | 93 | 1.99 | 0.108 | 0.025 |
| Philippines | PHL.7_1 | Apayao | 93 | 1.99 | 0.108 | 0.025 |
| Philippines | PHL.13_1 | Benguet | 93 | 1.99 | 0.108 | 0.025 |
| Philippines | PHL.33_1 | Ifugao | 93 | 1.99 | 0.108 | 0.025 |
| Philippines | PHL.38_1 | Kalinga | 93 | 1.99 | 0.108 | 0.025 |
| Philippines | PHL.50_1 | Mountain Province | 93 | 1.99 | 0.108 | 0.025 |
| Philippines | PHL.2_1 | Agusan del Norte | 90 | 14.17 | -0.081 | -0.079 |
| Philippines | PHL.3_1 | Agusan del Sur | 90 | 14.17 | -0.081 | -0.079 |
| Philippines | PHL.30_1 | Dinagat Islands | 90 | 14.17 | -0.081 | -0.079 |
| Philippines | PHL.74_1 | Surigao del Norte | 90 | 14.17 | -0.081 | -0.079 |
| Philippines | PHL.75_1 | Surigao del Sur | 90 | 14.17 | -0.081 | -0.079 |
| Philippines | PHL.45_1 | Marinduque | 83 | 9.89 | 0.44 | 0.167 |
| Philippines | PHL.57_1 | Occidental Mindoro | 83 | 9.89 | 0.44 | 0.167 |
| Philippines | PHL.58_1 | Oriental Mindoro | 83 | 9.89 | 0.44 | 0.167 |
| Philippines | PHL.59_1 | Palawan | 83 | 9.89 | 0.44 | 0.167 |
| Philippines | PHL.65_1 | Romblon | 83 | 9.89 | 0.44 | 0.167 |
| Philippines | PHL.34_1 | Ilocos Norte | 54 | 9.92 | 0.442 | 0.165 |
| Philippines | PHL.35_1 | Ilocos Sur | 54 | 9.92 | 0.442 | 0.165 |
| Philippines | PHL.39_1 | La Union | 54 | 9.92 | 0.442 | 0.165 |
| Philippines | PHL.61_1 | Pangasinan | 54 | 9.92 | 0.442 | 0.165 |
| Philippines | PHL.18_1 | Cagayan | 55 | 1.26 | 0.673 | 0.078 |
| Philippines | PHL.11_1 | Batanes | 55 | 1.26 | 0.673 | 0.078 |
| Philippines | PHL.37_1 | Isabela | 55 | 1.26 | 0.673 | 0.078 |
| Philippines | PHL.56_1 | Nueva Vizcaya | 55 | 1.26 | 0.673 | 0.078 |
| Philippines | PHL.63_1 | Quirino | 55 | 1.26 | 0.673 | 0.078 |
| Philippines | PHL.8_1 | Aurora | 128 | 3.09 | -0.289 | -0.021 |
| Philippines | PHL.10_1 | Bataan | 128 | 3.09 | -0.289 | -0.021 |
| Philippines | PHL.17_1 | Bulacan | 128 | 3.09 | -0.289 | -0.021 |
| Philippines | PHL.55_1 | Nueva Ecija | 128 | 3.09 | -0.289 | -0.021 |
| Philippines | PHL.60_1 | Pampanga | 128 | 3.09 | -0.289 | -0.021 |
| Philippines | PHL.76_1 | Tarlac | 128 | 3.09 | -0.289 | -0.021 |
| Philippines | PHL.78_1 | Zambales | 128 | 3.09 | -0.289 | -0.021 |
| Philippines | PHL.12_1 | Batangas | 65 | 5.88 | 0.718 | 0.162 |
| Philippines | PHL.24_1 | Cavite | 65 | 5.88 | 0.718 | 0.162 |
| Philippines | PHL.40_1 | Laguna | 65 | 5.88 | 0.718 | 0.162 |
| Philippines | PHL.62_1 | Quezon | 65 | 5.88 | 0.718 | 0.162 |
| Philippines | PHL.64_1 | Rizal | 65 | 5.88 | 0.718 | 0.162 |

| Country | adm1_gid | adm1_name | N | Zero-dose Prevalence | CI | AEG |
|---|---|---|---|---|---|---|
| Philippines | PHL.79_1 | Zamboanga del Norte | 74 | 17.17 | 0.345 | 0.223 |
| Philippines | PHL.80_1 | Zamboanga del Sur | 74 | 17.17 | 0.345 | 0.223 |
| Philippines | PHL.81_1 | Zamboanga Sibugay | 74 | 17.17 | 0.345 | 0.223 |
| Philippines | PHL.5_1 | Albay | 77 | 5.22 | -0.338 | -0.077 |
| Philippines | PHL.19_1 | Camarines Norte | 77 | 5.22 | -0.338 | -0.077 |
| Philippines | PHL.20_1 | Camarines Sur | 77 | 5.22 | -0.338 | -0.077 |
| Philippines | PHL.23_1 | Catanduanes | 77 | 5.22 | -0.338 | -0.077 |
| Philippines | PHL.46_1 | Masbate | 77 | 5.22 | -0.338 | -0.077 |
| Philippines | PHL.69_1 | Sorsogon | 77 | 5.22 | -0.338 | -0.077 |
| Philippines | PHL.4_1 | Aklan | 112 | 11.13 | 0.257 | 0.205 |
| Philippines | PHL.6_1 | Antique | 112 | 11.13 | 0.257 | 0.205 |
| Philippines | PHL.22_1 | Capiz | 112 | 11.13 | 0.257 | 0.205 |
| Philippines | PHL.32_1 | Guimaras | 112 | 11.13 | 0.257 | 0.205 |
| Philippines | PHL.36_1 | Iloilo | 112 | 11.13 | 0.257 | 0.205 |
| Philippines | PHL.51_1 | Negros Occidental | 112 | 11.13 | 0.257 | 0.205 |
| Philippines | PHL.15_1 | Bohol | 82 | 9.23 | 0.291 | 0.159 |
| Philippines | PHL.25_1 | Cebu | 82 | 9.23 | 0.291 | 0.159 |
| Philippines | PHL.52_1 | Negros Oriental | 82 | 9.23 | 0.291 | 0.159 |
| Philippines | PHL.68_1 | Siquijor | 82 | 9.23 | 0.291 | 0.159 |
| Philippines | PHL.14_1 | Biliran | 116 | 2.9 | -0.181 | 0.042 |
| Philippines | PHL.31_1 | Eastern Samar | 116 | 2.9 | -0.181 | 0.042 |
| Philippines | PHL.43_1 | Leyte | 116 | 2.9 | -0.181 | 0.042 |
| Philippines | PHL.54_1 | Northern Samar | 116 | 2.9 | -0.181 | 0.042 |
| Philippines | PHL.66_1 | Samar | 116 | 2.9 | -0.181 | 0.042 |
| Philippines | PHL.71_1 | Southern Leyte | 116 | 2.9 | -0.181 | 0.042 |
| Philippines | PHL.16_1 | Bukidnon | 106 | 12.25 | 0.138 | 0.02 |
| Philippines | PHL.21_1 | Camiguin | 106 | 12.25 | 0.138 | 0.02 |
| Philippines | PHL.41_1 | Lanao del Norte | 106 | 12.25 | 0.138 | 0.02 |
| Philippines | PHL.48_1 | Misamis Occidental | 106 | 12.25 | 0.138 | 0.02 |
| Philippines | PHL.49_1 | Misamis Oriental | 106 | 12.25 | 0.138 | 0.02 |
| Philippines | PHL.26_1 | Compostela Valley | 63 | 11 | 0.217 | -0.23 |
| Philippines | PHL.27_1 | Davao del Norte | 63 | 11 | 0.217 | -0.23 |
| Philippines | PHL.28_1 | Davao del Sur | 63 | 11 | 0.217 | -0.23 |
| Philippines | PHL.29_1 | Davao Oriental | 63 | 11 | 0.217 | -0.23 |
| Philippines | PHL.53_1 | North Cotabato | 85 | 22.91 | 0.221 | 0.235 |
| Philippines | PHL.67_1 | Sarangani | 85 | 22.91 | 0.221 | 0.235 |
| Philippines | PHL.70_1 | South Cotabato | 85 | 22.91 | 0.221 | 0.235 |

| Country | adm1_gid | adm1_name | N | Zero-dose Prevalence | CI | AEG |
|---|---|---|---|---|---|---|
| Philippines | PHL.72_1 | Sultan Kudarat | 85 | 22.91 | 0.221 | 0.235 |
| Rwanda | RWA.1_1 | Amajyaruguru | 260 | 0.7 | 0.869 | 0.027 |
| Rwanda | RWA.2_1 | Amajyepfo | 384 | 0 | | 0 |
| Rwanda | RWA.3_1 | Iburasirazuba | 368 | 0.35 | 0.97 | 0.019 |
| Rwanda | RWA.4_1 | Iburengerazuba | 399 | 0.27 | 0.085 | 0 |
| Rwanda | RWA.5_1 | Umujyi wa Kigali | 161 | 0.98 | 0.373 | 0.028 |
| Samoa | WSM.9_1 | Tuamasaga | 99 | 10.43 | | |
| Samoa | WSM.1_1 | A'ana | 98 | 17.77 | | |
| Samoa | WSM.2_1 | Aiga-i-le-Tai | 98 | 17.77 | | |
| Samoa | WSM.3_1 | Atua | 73 | 13.57 | | |
| Samoa | WSM.10_1 | Va'a-o-Fonoti | 73 | 13.57 | | |
| Samoa | WSM.11_1 | Vaisigano | 73 | 13.57 | | |
| Samoa | WSM.4_1 | Fa'asaleleaga | 93 | 3.27 | | |
| Samoa | WSM.5_1 | Gaga'emauga | 93 | 3.27 | | |
| Samoa | WSM.6_1 | Gagaifomauga | 93 | 3.27 | | |
| Samoa | WSM.7_1 | Palauli | 93 | 3.27 | | |
| Samoa | WSM.8_1 | Satupa'itea | 93 | 3.27 | | |
| São Tomé and Príncipe | STP.1_1 | Príncipe | 47 | 0 | | 0 |
| São Tomé and Príncipe | STP.2_1 | São Tomé | 283 | 0.25 | 0.954 | 0.013 |
| Senegal | SEN.10_1 | Saint-Louis | 119 | 5.95 | -0.061 | -0.012 |
| Senegal | SEN.11_1 | Sédhiou | 172 | 16.45 | 0.199 | 0.221 |
| Senegal | SEN.12_1 | Tambacounda | 173 | 30.5 | 0.134 | 0.161 |
| Senegal | SEN.13_1 | Thiès | 142 | 6.51 | 0.231 | 0.079 |
| Senegal | SEN.14_1 | Ziguinchor | 68 | 2.28 | 0.397 | 0 |
| Senegal | SEN.1_1 | Dakar | 137 | 6.48 | 0.409 | 0.176 |
| Senegal | SEN.2_1 | Diourbel | 133 | 12.32 | 0.269 | 0.1 |
| Senegal | SEN.3_1 | Fatick | 163 | 2.57 | 0.215 | 0.021 |
| Senegal | SEN.4_1 | Kaffrine | 195 | 4.77 | 0.102 | -0.008 |
| Senegal | SEN.5_1 | Kaolack | 137 | 6.12 | -0.125 | -0.103 |
| Senegal | SEN.6_1 | Kédougou | 134 | 14.76 | 0.108 | -0.009 |
| Senegal | SEN.7_1 | Kolda | 149 | 7.71 | -0.061 | -0.053 |
| Senegal | SEN.8_1 | Louga | 166 | 9.36 | 0.437 | 0.182 |
| Senegal | SEN.9_1 | Matam | 156 | 11.5 | 0.153 | 0.085 |
| Serbia | SRB.3_1 | Grad Beograd | 80 | 2.78 | 0.912 | 0.113 |
| Serbia | SRB.1_1 | Borski | 78 | 6.48 | 0.321 | 0.159 |

| Country | adm1_gid | adm1_name | N | Zero-dose Prevalence | CI | AEG |
|---|---|---|---|---|---|---|
| Serbia | SRB.2_1 | Braničevski | 78 | 6.48 | 0.321 | 0.159 |
| Serbia | SRB.22_1 | Toplički | 78 | 6.48 | 0.321 | 0.159 |
| Serbia | SRB.23_1 | Zaječarski | 78 | 6.48 | 0.321 | 0.159 |
| Serbia | SRB.10_1 | Nišavski | 78 | 6.48 | 0.321 | 0.159 |
| Serbia | SRB.11_1 | Pčinjski | 78 | 6.48 | 0.321 | 0.159 |
| Serbia | SRB.12_1 | Pirotski | 78 | 6.48 | 0.321 | 0.159 |
| Serbia | SRB.13_1 | Podunavski | 78 | 6.48 | 0.321 | 0.159 |
| Serbia | SRB.4_1 | Jablanički | 78 | 6.48 | 0.321 | 0.159 |
| Serbia | SRB.25_1 | Zlatiborski | 77 | 0 | | 0 |
| Serbia | SRB.21_1 | Šumadijski | 77 | 0 | | 0 |
| Serbia | SRB.14_1 | Pomoravski | 77 | 0 | | 0 |
| Serbia | SRB.15_1 | Rasinski | 77 | 0 | | 0 |
| Serbia | SRB.16_1 | Raški | 77 | 0 | | 0 |
| Serbia | SRB.7_1 | Kolubarski | 77 | 0 | | 0 |
| Serbia | SRB.8_1 | Mačvanski | 77 | 0 | | 0 |
| Serbia | SRB.9_1 | Moravički | 77 | 0 | | 0 |
| Serbia | SRB.5_1 | Južno-Bački | 93 | 0 | | 0 |
| Serbia | SRB.6_1 | Južno-Banatski | 93 | 0 | | 0 |
| Serbia | SRB.17_1 | Severno-Bački | 93 | 0 | | 0 |
| Serbia | SRB.18_1 | Severno-Banatski | 93 | 0 | | 0 |
| Serbia | SRB.19_1 | Srednje-Banatski | 93 | 0 | | 0 |
| Serbia | SRB.20_1 | Sremski | 93 | 0 | | 0 |
| Serbia | SRB.24_1 | Zapadno-Bački | 93 | 0 | | 0 |
| Sierra Leone | SLE.1_1 | Eastern | 360 | 6.95 | 0.15 | 0.056 |
| Sierra Leone | SLE.2_1 | Northern | 781 | 4.85 | 0.15 | 0.04 |
| Sierra Leone | SLE.3_1 | Southern | 478 | 3.55 | 0.234 | 0.04 |
| Sierra Leone | SLE.4_1 | Western | 233 | 3.7 | 0.073 | 0.057 |
| South Africa | ZAF.1_1 | Eastern Cape | 87 | 2.17 | 0.161 | 0.058 |
| South Africa | ZAF.2_1 | Free State | 49 | 2.21 | 0.857 | 0.087 |
| South Africa | ZAF.3_1 | Gauteng | 62 | 6.85 | 0.506 | 0.277 |
| South Africa | ZAF.4_1 | KwaZulu-Natal | 113 | 5.25 | 0.008 | 0.018 |
| South Africa | ZAF.5_1 | Limpopo | 91 | 1.81 | 0.582 | 0.052 |
| South Africa | ZAF.6_1 | Mpumalanga | 83 | 9.37 | 0.072 | 0.047 |
| South Africa | ZAF.7_1 | North West | 72 | 0 | | 0 |
| South Africa | ZAF.8_1 | Northern Cape | 51 | 4.22 | 0.961 | 0.198 |
| South Africa | ZAF.9_1 | Western Cape | 35 | 8.74 | 0.381 | 0.113 |
| Palestine | PSE.1_1 | Gaza | 375 | 0.3 | 0.89 | 0.014 |

| Country | adm1_gid | adm1_name | N | Zero-dose Prevalence | CI | AEG |
|---|---|---|---|---|---|---|
| Palestine | PSE.2_1 | West Bank | 897 | 5.35 | 0.309 | 0.088 |
| Suriname | SUR.10_1 | Wanica | 129 | 7.43 | 0.368 | 0.148 |
| Suriname | SUR.1_1 | Brokopondo | 29 | 13.42 | 0.06 | 0 |
| Suriname | SUR.2_1 | Commewijne | 59 | 4.66 | 0.716 | 0.317 |
| Suriname | SUR.3_1 | Coronie | 11 | 10.38 | 0.727 | 0.331 |
| Suriname | SUR.4_1 | Marowijne | 28 | 23.22 | -0.071 | -0.125 |
| Suriname | SUR.5_1 | Nickerie | 47 | 1.24 | 0.553 | 0.065 |
| Suriname | SUR.6_1 | Para | 67 | 2.86 | 0.328 | 0.133 |
| Suriname | SUR.7_1 | Paramaribo | 171 | 10.79 | 0.146 | 0.061 |
| Suriname | SUR.8_1 | Saramacca | 50 | 8.2 | 0.72 | 0.194 |
| Suriname | SUR.9_1 | Sipaliwini | 22 | 4.29 | 0.05 | 0 |
| Tajikistan | TJK.1_1 | Dushanbe | 179 | 10.64 | -0.179 | -0.07 |
| Tajikistan | TJK.2_1 | Gorno-Badakhshan | 66 | 2.9 | -0.333 | 0 |
| Tajikistan | TJK.3_1 | Khatlon | 276 | 12.3 | 0.245 | 0.23 |
| Tajikistan | TJK.4_1 | Sughd | 211 | 3.44 | 0.215 | 0.028 |
| Tajikistan | TJK.5_1 | Districts of Republican Subordin | 214 | 14.31 | 0.003 | 0.036 |
| Tanzania | TZA.10_1 | Lindi | 36 | 4.66 | 0.222 | 0 |
| Tanzania | TZA.11_1 | Manyara | 75 | 0 | | 0 |
| Tanzania | TZA.12_1 | Mara | 91 | 5.92 | 0.044 | 0.04 |
| Tanzania | TZA.13_1 | Mbeya | 56 | 14.62 | 0.451 | 0.326 |
| Tanzania | TZA.14_1 | Morogoro | 77 | 0 | | 0 |
| Tanzania | TZA.15_1 | Mtwara | 47 | 0 | | 0 |
| Tanzania | TZA.16_1 | Mwanza | 92 | 3.38 | 0.214 | 0.074 |
| Tanzania | TZA.17_1 | Njombe | 41 | 0 | | 0 |
| Tanzania | TZA.18_1 | Kaskazini Pemba | 69 | 0.75 | 0.058 | 0 |
| Tanzania | TZA.19_1 | Kusini Pemba | 78 | 0 | | 0 |
| Tanzania | TZA.1_1 | Arusha | 68 | 0.66 | 0.279 | 0 |
| Tanzania | TZA.20_1 | Pwani | 50 | 1.31 | 0.58 | 0 |
| Tanzania | TZA.21_1 | Rukwa | 66 | 5.69 | 0.389 | 0 |
| Tanzania | TZA.22_1 | Ruvuma | 62 | 1.88 | 0.79 | 0.091 |
| Tanzania | TZA.23_1 | Shinyanga | 88 | 18.74 | 0.016 | 0.034 |
| Tanzania | TZA.24_1 | Simiyu | 83 | 6.36 | 0.419 | 0.146 |
| Tanzania | TZA.25_1 | Singida | 52 | 2.73 | 0.692 | 0.098 |
| Tanzania | TZA.26_1 | Tabora | 115 | 6.15 | 0.256 | 0.072 |
| Tanzania | TZA.27_1 | Tanga | 68 | 6.9 | 0.456 | 0.168 |
| Tanzania | TZA.28_1 | Kaskazini Unguja | 51 | 4.37 | 0.02 | 0 |

| Country | adm1_gid | adm1_name | N | Zero-dose Prevalence | CI | AEG |
|---|---|---|---|---|---|---|
| Tanzania | TZA.29_1 | Kusini Unguja | 54 | 0 | | 0 |
| Tanzania | TZA.2_1 | Dar es Salaam | 78 | 3.92 | 0.064 | 0.081 |
| Tanzania | TZA.30_1 | Mjini Magharibi | 66 | 3.37 | -0.348 | 0 |
| Tanzania | TZA.31_1 | Songwe | 65 | 5.42 | 0.041 | 0.031 |
| Tanzania | TZA.3_1 | Dodoma | 57 | 0 | | 0 |
| Tanzania | TZA.4_1 | Geita | 94 | 15.13 | 0.32 | 0.248 |
| Tanzania | TZA.5_1 | Iringa | 62 | 2.68 | 0.806 | 0.12 |
| Tanzania | TZA.6_1 | Kagera | 87 | 4.24 | 0.382 | 0.05 |
| Tanzania | TZA.7_1 | Katavi | 81 | 15.24 | 0.233 | 0.112 |
| Tanzania | TZA.8_1 | Kigoma | 85 | 2 | -0.188 | 0 |
| Tanzania | TZA.9_1 | Kilimanjaro | 39 | 0 | | 0 |
| Thailand | THA.3_1 | Bangkok Metropolis | 84 | 0 | | 0 |
| Thailand | THA.2_1 | Ang Thong | 208 | 0 | | 0 |
| Thailand | THA.12_1 | Chon Buri | 208 | 0 | | 0 |
| Thailand | THA.6_1 | Chachoengsao | 208 | 0 | | 0 |
| Thailand | THA.46_1 | Phra Nakhon Si Ayutthaya | 208 | 0 | | 0 |
| Thailand | THA.49_1 | Prachin Buri | 208 | 0 | | 0 |
| Thailand | THA.53_1 | Rayong | 208 | 0 | | 0 |
| Thailand | THA.55_1 | Sa Kaeo | 208 | 0 | | 0 |
| Thailand | THA.71_1 | Trat | 208 | 0 | | 0 |
| Thailand | THA.7_1 | Chai Nat | 208 | 0 | | 0 |
| Thailand | THA.9_1 | Chanthaburi | 208 | 0 | | 0 |
| Thailand | THA.16_1 | Kanchanaburi | 208 | 0 | | 0 |
| Thailand | THA.22_1 | Lop Buri | 208 | 0 | | 0 |
| Thailand | THA.26_1 | Nakhon Nayok | 208 | 0 | | 0 |
| Thailand | THA.27_1 | Nakhon Pathom | 208 | 0 | | 0 |
| Thailand | THA.36_1 | Nonthaburi | 208 | 0 | | 0 |
| Thailand | THA.37_1 | Pathum Thani | 208 | 0 | | 0 |
| Thailand | THA.43_1 | Phetchaburi | 208 | 0 | | 0 |
| Thailand | THA.50_1 | Prachuap Khiri Khan | 208 | 0 | | 0 |
| Thailand | THA.52_1 | Ratchaburi | 208 | 0 | | 0 |
| Thailand | THA.57_1 | Samut Prakan | 208 | 0 | | 0 |
| Thailand | THA.58_1 | Samut Sakhon | 208 | 0 | | 0 |
| Thailand | THA.59_1 | Samut Songkhram | 208 | 0 | | 0 |
| Thailand | THA.60_1 | Saraburi | 208 | 0 | | 0 |
| Thailand | THA.63_1 | Sing Buri | 208 | 0 | | 0 |

| Country | adm1_gid | adm1_name | N | Zero-dose Prevalence | CI | AEG |
|---|---|---|---|---|---|---|
| Thailand | THA.66_1 | Suphan Buri | 208 | 0 | | 0 |
| Thailand | THA.10_1 | Chiang Mai | 331 | 0.07 | 0.157 | 0 |
| Thailand | THA.11_1 | Chiang Rai | 331 | 0.07 | 0.157 | 0 |
| Thailand | THA.19_1 | Lampang | 331 | 0.07 | 0.157 | 0 |
| Thailand | THA.20_1 | Lamphun | 331 | 0.07 | 0.157 | 0 |
| Thailand | THA.23_1 | Mae Hong Son | 331 | 0.07 | 0.157 | 0 |
| Thailand | THA.30_1 | Nakhon Sawan | 331 | 0.07 | 0.157 | 0 |
| Thailand | THA.32_1 | Nan | 331 | 0.07 | 0.157 | 0 |
| Thailand | THA.41_1 | Phayao | 331 | 0.07 | 0.157 | 0 |
| Thailand | THA.47_1 | Phrae | 331 | 0.07 | 0.157 | 0 |
| Thailand | THA.65_1 | Sukhothai | 331 | 0.07 | 0.157 | 0 |
| Thailand | THA.15_1 | Kamphaeng Phet | 331 | 0.07 | 0.157 | 0 |
| Thailand | THA.42_1 | Phetchabun | 331 | 0.07 | 0.157 | 0 |
| Thailand | THA.44_1 | Phichit | 331 | 0.07 | 0.157 | 0 |
| Thailand | THA.45_1 | Phitsanulok | 331 | 0.07 | 0.157 | 0 |
| Thailand | THA.74_1 | Uthai Thani | 331 | 0.07 | 0.157 | 0 |
| Thailand | THA.75_1 | Uttaradit | 331 | 0.07 | 0.157 | 0 |
| Thailand | THA.69_1 | Tak | 331 | 0.07 | 0.157 | 0 |
| Thailand | THA.1_1 | Amnat Charoen | 600 | 0.05 | 0.688 | 0.002 |
| Thailand | THA.4_1 | Bueng Kan | 600 | 0.05 | 0.688 | 0.002 |
| Thailand | THA.5_1 | Buri Ram | 600 | 0.05 | 0.688 | 0.002 |
| Thailand | THA.8_1 | Chaiyaphum | 600 | 0.05 | 0.688 | 0.002 |
| Thailand | THA.14_1 | Kalasin | 600 | 0.05 | 0.688 | 0.002 |
| Thailand | THA.17_1 | Khon Kaen | 600 | 0.05 | 0.688 | 0.002 |
| Thailand | THA.21_1 | Loei | 600 | 0.05 | 0.688 | 0.002 |
| Thailand | THA.24_1 | Maha Sarakham | 600 | 0.05 | 0.688 | 0.002 |
| Thailand | THA.25_1 | Mukdahan | 600 | 0.05 | 0.688 | 0.002 |
| Thailand | THA.28_1 | Nakhon Phanom | 600 | 0.05 | 0.688 | 0.002 |
| Thailand | THA.29_1 | Nakhon Ratchasima | 600 | 0.05 | 0.688 | 0.002 |
| Thailand | THA.34_1 | Nong Bua Lam Phu | 600 | 0.05 | 0.688 | 0.002 |
| Thailand | THA.35_1 | Nong Khai | 600 | 0.05 | 0.688 | 0.002 |
| Thailand | THA.54_1 | Roi Et | 600 | 0.05 | 0.688 | 0.002 |
| Thailand | THA.56_1 | Sakon Nakhon | 600 | 0.05 | 0.688 | 0.002 |
| Thailand | THA.62_1 | Si Sa Ket | 600 | 0.05 | 0.688 | 0.002 |
| Thailand | THA.68_1 | Surin | 600 | 0.05 | 0.688 | 0.002 |
| Thailand | THA.72_1 | Ubon Ratchathani | 600 | 0.05 | 0.688 | 0.002 |
| Thailand | THA.73_1 | Udon Thani | 600 | 0.05 | 0.688 | 0.002 |

| Country | adm1_gid | adm1_name | N | Zero-dose Prevalence | CI | AEG |
|---|---|---|---|---|---|---|
| Thailand | THA.77_1 | Yasothon | 600 | 0.05 | 0.688 | 0.002 |
| Thailand | THA.13_1 | Chumphon | 609 | 6.44 | 0.506 | 0.132 |
| Thailand | THA.18_1 | Krabi | 609 | 6.44 | 0.506 | 0.132 |
| Thailand | THA.31_1 | Nakhon Si Thammarat | 609 | 6.44 | 0.506 | 0.132 |
| Thailand | THA.33_1 | Narathiwat | 609 | 6.44 | 0.506 | 0.132 |
| Thailand | THA.38_1 | Pattani | 609 | 6.44 | 0.506 | 0.132 |
| Thailand | THA.39_1 | Phangnga | 609 | 6.44 | 0.506 | 0.132 |
| Thailand | THA.40_1 | Phatthalung | 609 | 6.44 | 0.506 | 0.132 |
| Thailand | THA.48_1 | Phuket | 609 | 6.44 | 0.506 | 0.132 |
| Thailand | THA.51_1 | Ranong | 609 | 6.44 | 0.506 | 0.132 |
| Thailand | THA.67_1 | Surat Thani | 609 | 6.44 | 0.506 | 0.132 |
| Thailand | THA.61_1 | Satun | 609 | 6.44 | 0.506 | 0.132 |
| Thailand | THA.64_1 | Songkhla | 609 | 6.44 | 0.506 | 0.132 |
| Thailand | THA.70_1 | Trang | 609 | 6.44 | 0.506 | 0.132 |
| Thailand | THA.76_1 | Yala | 609 | 6.44 | 0.506 | 0.132 |
| Timor-Leste | TLS.10_1 | Liquiçá | 112 | 22.96 | 0.085 | 0.131 |
| Timor-Leste | TLS.11_1 | Manatuto | 94 | 18.33 | 0.261 | 0.334 |
| Timor-Leste | TLS.12_1 | Manufahi | 126 | 30.27 | 0.31 | 0.367 |
| Timor-Leste | TLS.13_1 | Viqueque | 99 | 24.93 | 0.18 | 0.294 |
| Timor-Leste | TLS.1_1 | Aileu | 113 | 10.23 | 0.1 | -0.013 |
| Timor-Leste | TLS.2_1 | Ainaro | 102 | 30.78 | 0.19 | 0.161 |
| Timor-Leste | TLS.3_1 | Ambeno | 86 | 18.02 | 0.101 | 0.138 |
| Timor-Leste | TLS.4_1 | Baucau | 110 | 12.46 | 0.347 | 0.232 |
| Timor-Leste | TLS.5_1 | Bobonaro | 114 | 23.18 | 0.061 | -0.006 |
| Timor-Leste | TLS.6_1 | Covalima | 98 | 32.22 | 0.427 | 0.617 |
| Timor-Leste | TLS.7_1 | Dili | 162 | 11.13 | 0.221 | 0.089 |
| Timor-Leste | TLS.8_1 | Ermera | 104 | 41.38 | 0.089 | 0.279 |
| Timor-Leste | TLS.9_1 | Lautém | 98 | 24.39 | 0.173 | 0.204 |
| Togo | TGO.1_1 | Centre | 136 | 0.87 | 0.846 | 0.046 |
| Togo | TGO.2_1 | Kara | 139 | 5.32 | 0.476 | 0.144 |
| Togo | TGO.3_1 | Maritime | 306 | 1.81 | 0.62 | 0.05 |
| Togo | TGO.4_1 | Plateaux | 124 | 2.47 | 0.294 | 0.088 |
| Togo | TGO.5_1 | Savanes | 178 | 1.53 | 0.27 | 0 |
| Tonga | TON.1_1 | 'Eua | 17 | 5.31 | 0.706 | 0.181 |
| Tonga | TON.2_1 | Ha'apai | 29 | 0 | | 0 |
| Tonga | TON.3_1 | Niuas | 7 | 0 | | 0 |
| Tonga | TON.4_1 | Tongatapu | 65 | 2.84 | 0.985 | 0.1 |

| Country | adm1_gid | adm1_name | N | Zero-dose Prevalence | CI | AEG |
|---|---|---|---|---|---|---|
| Tonga | TON.5_1 | Vava'u | 36 | 0 | | 0 |
| Trinidad and Tobago | TTO.14_1 | Tobago | 31 | 0 | | 0 |
| Trinidad and Tobago | TTO.5_1 | Mayaro/Rio Claro | 31 | 1.06 | 0.1 | 0 |
| Trinidad and Tobago | TTO.12_1 | Sangre Grande | 31 | 1.06 | 0.1 | 0 |
| Trinidad and Tobago | TTO.1_1 | Arima | 41 | 7.96 | 0.813 | 0.592 |
| Trinidad and Tobago | TTO.11_1 | San Juan-Laventille | 41 | 7.96 | 0.813 | 0.592 |
| Trinidad and Tobago | TTO.15_1 | Tunapuna/Piarco | 41 | 7.96 | 0.813 | 0.592 |
| Trinidad and Tobago | TTO.4_1 | Diego Martin | 23 | 0 | | 0 |
| Trinidad and Tobago | TTO.8_1 | Port of Spain | 23 | 0 | | 0 |
| Trinidad and Tobago | TTO.2_1 | Chaguanas | 48 | 1.6 | -0.397 | -0.043 |
| Trinidad and Tobago | TTO.3_1 | Couva-Tabaquite-Talparo | 48 | 1.6 | -0.397 | -0.043 |
| Trinidad and Tobago | TTO.6_1 | Penal-Debe | 48 | 1.6 | -0.397 | -0.043 |
| Trinidad and Tobago | TTO.7_1 | Point Fortin | 48 | 1.6 | -0.397 | -0.043 |
| Trinidad and Tobago | TTO.9_1 | Princes Town | 48 | 1.6 | -0.397 | -0.043 |
| Trinidad and Tobago | TTO.10_1 | San Fernando | 48 | 1.6 | -0.397 | -0.043 |
| Trinidad and Tobago | TTO.13_1 | Siparia | 48 | 1.6 | -0.397 | -0.043 |
| Tunisia | TUN.12_1 | Mahdia | 44 | 0 | | 0 |
| Tunisia | TUN.15_1 | Monastir | 44 | 0 | | 0 |
| Tunisia | TUN.17_1 | Sfax | 44 | 0 | | 0 |
| Tunisia | TUN.20_1 | Sousse | 44 | 0 | | 0 |
| Tunisia | TUN.8_1 | Kairouan | 43 | 2.16 | 0.977 | 0.103 |
| Tunisia | TUN.9_1 | Kassérine | 43 | 2.16 | 0.977 | 0.103 |

| Country | adm1_gid | adm1_name | N | Zero-dose Prevalence | CI | AEG |
|---|---|---|---|---|---|---|
| Tunisia | TUN.18_1 | Sidi Bou Zid | 43 | 2.16 | 0.977 | 0.103 |
| Tunisia | TUN.23_1 | Tunis | 18 | 0 | | 0 |
| Tunisia | TUN.1_1 | Ariana | 18 | 0 | | 0 |
| Tunisia | TUN.3_1 | Ben Arous (Tunis Sud) | 18 | 0 | | 0 |
| Tunisia | TUN.13_1 | Manubah | 18 | 0 | | 0 |
| Tunisia | TUN.4_1 | Bizerte | 22 | 0 | | 0 |
| Tunisia | TUN.16_1 | Nabeul | 22 | 0 | | 0 |
| Tunisia | TUN.24_1 | Zaghouan | 22 | 0 | | 0 |
| Tunisia | TUN.2_1 | Béja | 47 | 0 | | 0 |
| Tunisia | TUN.7_1 | Jendouba | 47 | 0 | | 0 |
| Tunisia | TUN.11_1 | Le Kef | 47 | 0 | | 0 |
| Tunisia | TUN.19_1 | Siliana | 47 | 0 | | 0 |
| Tunisia | TUN.5_1 | Gabès | 40 | 0 | | 0 |
| Tunisia | TUN.14_1 | Médenine | 40 | 0 | | 0 |
| Tunisia | TUN.21_1 | Tataouine | 40 | 0 | | 0 |
| Tunisia | TUN.6_1 | Gafsa | 47 | 0 | | 0 |
| Tunisia | TUN.10_1 | Kebili | 47 | 0 | | 0 |
| Tunisia | TUN.22_1 | Tozeur | 47 | 0 | | 0 |
| Turkey | TUR.23_1 | Çankiri | 74 | 2.32 | 0.554 | 0.03 |
| Turkey | TUR.32_1 | Eskisehir | 74 | 2.32 | 0.554 | 0.03 |
| Turkey | TUR.44_1 | Karaman | 74 | 2.32 | 0.554 | 0.03 |
| Turkey | TUR.49_1 | Kinkkale | 74 | 2.32 | 0.554 | 0.03 |
| Turkey | TUR.5_1 | Aksaray | 74 | 2.32 | 0.554 | 0.03 |
| Turkey | TUR.51_1 | Kirsehir | 74 | 2.32 | 0.554 | 0.03 |
| Turkey | TUR.53_1 | Konya | 74 | 2.32 | 0.554 | 0.03 |
| Turkey | TUR.61_1 | Nevsehir | 74 | 2.32 | 0.554 | 0.03 |
| Turkey | TUR.62_1 | Nigde | 74 | 2.32 | 0.554 | 0.03 |
| Turkey | TUR.7_1 | Ankara | 74 | 2.32 | 0.554 | 0.03 |
| Turkey | TUR.47_1 | Kayseri | 74 | 2.32 | 0.554 | 0.03 |
| Turkey | TUR.72_1 | Sivas | 74 | 2.32 | 0.554 | 0.03 |
| Turkey | TUR.80_1 | Yozgat | 74 | 2.32 | 0.554 | 0.03 |
| Turkey | TUR.14_1 | Batman | 170 | 3.44 | 0.519 | 0.108 |
| Turkey | TUR.17_1 | Bingöl | 170 | 3.44 | 0.519 | 0.108 |
| Turkey | TUR.18_1 | Bitlis | 170 | 3.44 | 0.519 | 0.108 |
| Turkey | TUR.2_1 | Adiyaman | 170 | 3.44 | 0.519 | 0.108 |
| Turkey | TUR.26_1 | Diyarbakir | 170 | 3.44 | 0.519 | 0.108 |
| Turkey | TUR.29_1 | Elazığ | 170 | 3.44 | 0.519 | 0.108 |

| Country | adm1_gid | adm1_name | N | Zero-dose Prevalence | CI | AEG |
|---|---|---|---|---|---|---|
| Turkey | TUR.30_1 | Erzincan | 170 | 3.44 | 0.519 | 0.108 |
| Turkey | TUR.31_1 | Erzurum | 170 | 3.44 | 0.519 | 0.108 |
| Turkey | TUR.33_1 | Gaziantep | 170 | 3.44 | 0.519 | 0.108 |
| Turkey | TUR.36_1 | Hakkari | 170 | 3.44 | 0.519 | 0.108 |
| Turkey | TUR.38_1 | Iğdır | 170 | 3.44 | 0.519 | 0.108 |
| Turkey | TUR.4_1 | Agri | 170 | 3.44 | 0.519 | 0.108 |
| Turkey | TUR.45_1 | Kars | 170 | 3.44 | 0.519 | 0.108 |
| Turkey | TUR.48_1 | Kilis | 170 | 3.44 | 0.519 | 0.108 |
| Turkey | TUR.55_1 | Malatya | 170 | 3.44 | 0.519 | 0.108 |
| Turkey | TUR.57_1 | Mardin | 170 | 3.44 | 0.519 | 0.108 |
| Turkey | TUR.60_1 | Mus | 170 | 3.44 | 0.519 | 0.108 |
| Turkey | TUR.68_1 | Sanliurfa | 170 | 3.44 | 0.519 | 0.108 |
| Turkey | TUR.69_1 | Siirt | 170 | 3.44 | 0.519 | 0.108 |
| Turkey | TUR.71_1 | Sirnak | 170 | 3.44 | 0.519 | 0.108 |
| Turkey | TUR.76_1 | Tunceli | 170 | 3.44 | 0.519 | 0.108 |
| Turkey | TUR.78_1 | Van | 170 | 3.44 | 0.519 | 0.108 |
| Turkey | TUR.9_1 | Ardahan | 170 | 3.44 | 0.519 | 0.108 |
| Turkey | TUR.19_1 | Bolu | 40 | 13.26 | 0.045 | -0.022 |
| Turkey | TUR.27_1 | Düzce | 40 | 13.26 | 0.045 | -0.022 |
| Turkey | TUR.10_1 | Artvin | 40 | 13.26 | 0.045 | -0.022 |
| Turkey | TUR.13_1 | Bartın | 40 | 13.26 | 0.045 | -0.022 |
| Turkey | TUR.15_1 | Bayburt | 40 | 13.26 | 0.045 | -0.022 |
| Turkey | TUR.24_1 | Çorum | 40 | 13.26 | 0.045 | -0.022 |
| Turkey | TUR.34_1 | Giresun | 40 | 13.26 | 0.045 | -0.022 |
| Turkey | TUR.35_1 | Gümüshane | 40 | 13.26 | 0.045 | -0.022 |
| Turkey | TUR.43_1 | Karabük | 40 | 13.26 | 0.045 | -0.022 |
| Turkey | TUR.46_1 | Kastamonu | 40 | 13.26 | 0.045 | -0.022 |
| Turkey | TUR.6_1 | Amasya | 40 | 13.26 | 0.045 | -0.022 |
| Turkey | TUR.63_1 | Ordu | 40 | 13.26 | 0.045 | -0.022 |
| Turkey | TUR.65_1 | Rize | 40 | 13.26 | 0.045 | -0.022 |
| Turkey | TUR.67_1 | Samsun | 40 | 13.26 | 0.045 | -0.022 |
| Turkey | TUR.70_1 | Sinop | 40 | 13.26 | 0.045 | -0.022 |
| Turkey | TUR.74_1 | Tokat | 40 | 13.26 | 0.045 | -0.022 |
| Turkey | TUR.75_1 | Trabzon | 40 | 13.26 | 0.045 | -0.022 |
| Turkey | TUR.81_1 | Zinguldak | 40 | 13.26 | 0.045 | -0.022 |
| Turkey | TUR.1_1 | Adana | 62 | 3.12 | 0.097 | 0.073 |
| Turkey | TUR.20_1 | Burdur | 62 | 3.12 | 0.097 | 0.073 |

| Country | adm1_gid | adm1_name | N | Zero-dose Prevalence | CI | AEG |
|---|---|---|---|---|---|---|
| Turkey | TUR.37_1 | Hatay | 62 | 3.12 | 0.097 | 0.073 |
| Turkey | TUR.39_1 | Isparta | 62 | 3.12 | 0.097 | 0.073 |
| Turkey | TUR.42_1 | K. Maras | 62 | 3.12 | 0.097 | 0.073 |
| Turkey | TUR.58_1 | Mersin | 62 | 3.12 | 0.097 | 0.073 |
| Turkey | TUR.64_1 | Osmaniye | 62 | 3.12 | 0.097 | 0.073 |
| Turkey | TUR.8_1 | Antalya | 62 | 3.12 | 0.097 | 0.073 |
| Turkey | TUR.11_1 | Aydin | 101 | 3.37 | 0.389 | 0.09 |
| Turkey | TUR.12_1 | Balikesir | 101 | 3.37 | 0.389 | 0.09 |
| Turkey | TUR.16_1 | Bilecik | 101 | 3.37 | 0.389 | 0.09 |
| Turkey | TUR.21_1 | Bursa | 101 | 3.37 | 0.389 | 0.09 |
| Turkey | TUR.22_1 | Çanakkale | 101 | 3.37 | 0.389 | 0.09 |
| Turkey | TUR.25_1 | Denizli | 101 | 3.37 | 0.389 | 0.09 |
| Turkey | TUR.28_1 | Edirne | 101 | 3.37 | 0.389 | 0.09 |
| Turkey | TUR.40_1 | Istanbul | 101 | 3.37 | 0.389 | 0.09 |
| Turkey | TUR.41_1 | Izmir | 101 | 3.37 | 0.389 | 0.09 |
| Turkey | TUR.50_1 | Kirklareli | 101 | 3.37 | 0.389 | 0.09 |
| Turkey | TUR.54_1 | Kütahya | 101 | 3.37 | 0.389 | 0.09 |
| Turkey | TUR.56_1 | Manisa | 101 | 3.37 | 0.389 | 0.09 |
| Turkey | TUR.59_1 | Mugla | 101 | 3.37 | 0.389 | 0.09 |
| Turkey | TUR.73_1 | Tekirdag | 101 | 3.37 | 0.389 | 0.09 |
| Turkey | TUR.77_1 | Usak | 101 | 3.37 | 0.389 | 0.09 |
| Turkey | TUR.79_1 | Yalova | 101 | 3.37 | 0.389 | 0.09 |
| Turkey | TUR.3_1 | Afyon | 101 | 3.37 | 0.389 | 0.09 |
| Turkey | TUR.52_1 | Kocaeli | 101 | 3.37 | 0.389 | 0.09 |
| Turkey | TUR.66_1 | Sakarya | 101 | 3.37 | 0.389 | 0.09 |
| Turks and Caicos Islands | TCA.1_1 | Grand Turk | 10 | 0 | | |
| Turks and Caicos Islands | TCA.3_1 | North Caicos | 1 | 0 | | |
| Turks and Caicos Islands | TCA.4_1 | Providenciales and West Caicos | 16 | 0 | | |
| Turks and Caicos Islands | TCA.6_1 | South Caicos and East Caicos | 3 | 0 | | |
| Tuvalu | TUV.1_1 | Funafuti | 49 | 0 | | |
| Tuvalu | TUV.2_1 | Nanumanga | 4 | 0 | | |
| Tuvalu | TUV.3_1 | Nanumea | 10 | 0 | | |
| Tuvalu | TUV.5_1 | Niutao | 6 | 0 | | |

| Country | adm1_gid | adm1_name | N | Zero-dose Prevalence | CI | AEG |
|---|---|---|---|---|---|---|
| Tuvalu | TUV.6_1 | Nui | 5 | 0 | | |
| Tuvalu | TUV.7_1 | Nukufetau | 6 | 0 | | |
| Tuvalu | TUV.9_1 | Vaitupu | 9 | 0 | | |
| Uganda | UGA.16_1 | Kampala | 164 | 4.5 | 0.217 | 0.015 |
| Uganda | UGA.50_1 | Pallisa | 204 | 4.45 | | |
| Uganda | UGA.27_1 | Kitgum | 156 | 0.35 | 0.224 | 0 |
| Uganda | UGA.49_1 | Pader | 156 | 0.35 | 0.224 | 0 |
| Uganda | UGA.8_1 | Gulu | 156 | 0.35 | 0.224 | 0 |
| Uganda | UGA.39_1 | Mbarara | 178 | 3.11 | -0.062 | 0.018 |
| Uganda | UGA.48_1 | Ntungamo | 178 | 3.11 | -0.062 | 0.018 |
| Uganda | UGA.6_1 | Bushenyi | 178 | 3.11 | -0.062 | 0.018 |
| Uganda | UGA.20_1 | Kapchorwa | 155 | 2.1 | 0.106 | -0.016 |
| Uganda | UGA.38_1 | Mbale | 155 | 2.1 | 0.106 | -0.016 |
| Uganda | UGA.54_1 | Sironko | 155 | 2.1 | 0.106 | -0.016 |
| Uganda | UGA.24_1 | Kibale | 189 | 5.08 | 0.58 | 0.129 |
| Uganda | UGA.36_1 | Masindi | 189 | 5.08 | 0.58 | 0.129 |
| Uganda | UGA.9_1 | Hoima | 189 | 5.08 | 0.58 | 0.129 |
| Uganda | UGA.31_1 | Lake Albert | 189 | 5.08 | 0.58 | 0.129 |
| Uganda | UGA.10_1 | Iganga | 249 | 6.94 | 0.06 | -0.071 |
| Uganda | UGA.11_1 | Jinja | 249 | 6.94 | 0.06 | -0.071 |
| Uganda | UGA.17_1 | Kamuli | 249 | 6.94 | 0.06 | -0.071 |
| Uganda | UGA.37_1 | Mayuge | 249 | 6.94 | 0.06 | -0.071 |
| Uganda | UGA.4_1 | Bugiri | 249 | 6.94 | 0.06 | -0.071 |
| Uganda | UGA.28_1 | Kotido | 164 | 1.46 | -0.247 | -0.024 |
| Uganda | UGA.40_1 | Moroto | 164 | 1.46 | -0.247 | -0.024 |
| Uganda | UGA.45_1 | Nakapiripirit | 164 | 1.46 | -0.247 | -0.024 |
| Uganda | UGA.12_1 | Kabale | 119 | 0.82 | -0.739 | -0.039 |
| Uganda | UGA.26_1 | Kisoro | 119 | 0.82 | -0.739 | -0.039 |
| Uganda | UGA.19_1 | Kanungu | 119 | 0.82 | -0.739 | -0.039 |
| Uganda | UGA.52_1 | Rukungiri | 119 | 0.82 | -0.739 | -0.039 |
| Uganda | UGA.2_1 | Apac | 182 | 4 | 0.379 | 0.064 |
| Uganda | UGA.33_1 | Lira | 182 | 4 | 0.379 | 0.064 |
| Uganda | UGA.23_1 | Kayunga | 231 | 7.43 | 0.477 | 0.238 |
| Uganda | UGA.44_1 | Mukono | 231 | 7.43 | 0.477 | 0.238 |
| Uganda | UGA.25_1 | Kiboga | 231 | 7.43 | 0.477 | 0.238 |
| Uganda | UGA.34_1 | Luwero | 231 | 7.43 | 0.477 | 0.238 |
| Uganda | UGA.43_1 | Mubende | 231 | 7.43 | 0.477 | 0.238 |

| Country | adm1_gid | adm1_name | N | Zero-dose Prevalence | CI | AEG |
|---|---|---|---|---|---|---|
| Uganda | UGA.46_1 | Nakasongola | 231 | 7.43 | 0.477 | 0.238 |
| Uganda | UGA.15_1 | Kalangala | 242 | 8.45 | 0.357 | 0.104 |
| Uganda | UGA.35_1 | Masaka | 242 | 8.45 | 0.357 | 0.104 |
| Uganda | UGA.42_1 | Mpigi | 242 | 8.45 | 0.357 | 0.104 |
| Uganda | UGA.51_1 | Rakai | 242 | 8.45 | 0.357 | 0.104 |
| Uganda | UGA.53_1 | Sembabule | 242 | 8.45 | 0.357 | 0.104 |
| Uganda | UGA.57_1 | Wakiso | 242 | 8.45 | 0.357 | 0.104 |
| Uganda | UGA.32_1 | Lake Victoria | 242 | 8.45 | 0.357 | 0.104 |
| Uganda | UGA.14_1 | Kaberamaido | 232 | 1.72 | -0.853 | -0.071 |
| Uganda | UGA.22_1 | Katakwi | 232 | 1.72 | -0.853 | -0.071 |
| Uganda | UGA.29_1 | Kumi | 232 | 1.72 | -0.853 | -0.071 |
| Uganda | UGA.55_1 | Soroti | 232 | 1.72 | -0.853 | -0.071 |
| Uganda | UGA.13_1 | Kabarole | 227 | 5.05 | 0.15 | 0.033 |
| Uganda | UGA.18_1 | Kamwenge | 227 | 5.05 | 0.15 | 0.033 |
| Uganda | UGA.21_1 | Kasese | 227 | 5.05 | 0.15 | 0.033 |
| Uganda | UGA.30_1 | Kyenjojo | 227 | 5.05 | 0.15 | 0.033 |
| Uganda | UGA.5_1 | Bundibugyo | 227 | 5.05 | 0.15 | 0.033 |
| Uganda | UGA.1_1 | Adjumani | 218 | 2.38 | 0.151 | 0.017 |
| Uganda | UGA.3_1 | Arua | 218 | 2.38 | 0.151 | 0.017 |
| Uganda | UGA.41_1 | Moyo | 218 | 2.38 | 0.151 | 0.017 |
| Uganda | UGA.47_1 | Nebbi | 218 | 2.38 | 0.151 | 0.017 |
| Uganda | UGA.58_1 | Yumbe | 218 | 2.38 | 0.151 | 0.017 |
| Uzbekistan | UZB.10_1 | Samarqand' | 79 | 0 | | 0 |
| Uzbekistan | UZB.11_1 | Sirdaryo | 69 | 0 | | 0 |
| Uzbekistan | UZB.12_1 | Surxondaryo | 80 | 0 | | 0 |
| Uzbekistan | UZB.13_1 | Toshkent Shahri | 41 | 2.72 | 0.098 | -0.046 |
| Uzbekistan | UZB.14_1 | Toshkent | 52 | 0 | | 0 |
| Uzbekistan | UZB.1_1 | Andijon | 86 | 2.31 | 0.605 | 0.044 |
| Uzbekistan | UZB.2_1 | Buxoro | 48 | 0 | | 0 |
| Uzbekistan | UZB.3_1 | Farg'ona | 62 | 0 | | 0 |
| Uzbekistan | UZB.4_1 | Jizzax | 73 | 0 | | 0 |
| Uzbekistan | UZB.5_1 | Qaraqalpaqstan | 93 | 0.98 | 0.602 | 0.039 |
| Uzbekistan | UZB.6_1 | Qashqadaryo | 88 | 0 | | 0 |
| Uzbekistan | UZB.7_1 | Xorazm | 90 | 2.62 | 0.774 | 0.129 |
| Uzbekistan | UZB.8_1 | Namangan | 78 | 0 | | 0 |
| Uzbekistan | UZB.9_1 | Navoiy | 66 | 0 | | 0 |
| Vanuatu | VUT.1_1 | Malampa | 39 | 7.61 | 0.154 | 0.088 |

| Country | adm1_gid | adm1_name | N | Zero-dose Prevalence | CI | AEG |
|---|---|---|---|---|---|---|
| Vanuatu | VUT.2_1 | Penama | 39 | 12.6 | 0.287 | 0.08 |
| Vanuatu | VUT.3_1 | Sanma | 67 | 16.9 | 0.196 | 0.141 |
| Vanuatu | VUT.4_1 | Shefa | 75 | 1.28 | 0.053 | 0 |
| Vanuatu | VUT.5_1 | Tafea | 74 | 30.85 | 0.276 | 0.373 |
| Vanuatu | VUT.6_1 | Torba | 20 | 0 | | 0 |
| Vietnam | VNM.34_1 | Kon Tum | 103 | 12.01 | 0.039 | 0.039 |
| Vietnam | VNM.21_1 | Gia Lai | 103 | 12.01 | 0.039 | 0.039 |
| Vietnam | VNM.15_1 | Đắk Lắk | 103 | 12.01 | 0.039 | 0.039 |
| Vietnam | VNM.16_1 | Đắk Nông | 103 | 12.01 | 0.039 | 0.039 |
| Vietnam | VNM.37_1 | Lâm Đồng | 103 | 12.01 | 0.039 | 0.039 |
| Vietnam | VNM.12_1 | Cần Thơ | 109 | 11.83 | 0.405 | 0.4 |
| Vietnam | VNM.39_1 | Long An | 109 | 11.83 | 0.405 | 0.4 |
| Vietnam | VNM.58_1 | Tiền Giang | 109 | 11.83 | 0.405 | 0.4 |
| Vietnam | VNM.6_1 | Bến Tre | 109 | 11.83 | 0.405 | 0.4 |
| Vietnam | VNM.59_1 | Trà Vinh | 109 | 11.83 | 0.405 | 0.4 |
| Vietnam | VNM.61_1 | Vĩnh Long | 109 | 11.83 | 0.405 | 0.4 |
| Vietnam | VNM.18_1 | Đồng Tháp | 109 | 11.83 | 0.405 | 0.4 |
| Vietnam | VNM.1_1 | An Giang | 109 | 11.83 | 0.405 | 0.4 |
| Vietnam | VNM.33_1 | Kiên Giang | 109 | 11.83 | 0.405 | 0.4 |
| Vietnam | VNM.24_1 | Hậu Giang | 109 | 11.83 | 0.405 | 0.4 |
| Vietnam | VNM.51_1 | Sóc Trăng | 109 | 11.83 | 0.405 | 0.4 |
| Vietnam | VNM.2_1 | Bạc Liêu | 109 | 11.83 | 0.405 | 0.4 |
| Vietnam | VNM.13_1 | Cà Mau | 109 | 11.83 | 0.405 | 0.4 |
| Vietnam | VNM.57_1 | Thanh Hóa | 94 | 7.19 | 0.4 | 0.067 |
| Vietnam | VNM.41_1 | Nghệ An | 94 | 7.19 | 0.4 | 0.067 |
| Vietnam | VNM.29_1 | Hà Tĩnh | 94 | 7.19 | 0.4 | 0.067 |
| Vietnam | VNM.46_1 | Quảng Bình | 94 | 7.19 | 0.4 | 0.067 |
| Vietnam | VNM.50_1 | Quảng Trị | 94 | 7.19 | 0.4 | 0.067 |
| Vietnam | VNM.54_1 | Thừa Thiên Huế | 94 | 7.19 | 0.4 | 0.067 |
| Vietnam | VNM.19_1 | Đà Nẵng | 94 | 7.19 | 0.4 | 0.067 |
| Vietnam | VNM.47_1 | Quảng Nam | 94 | 7.19 | 0.4 | 0.067 |
| Vietnam | VNM.48_1 | Quảng Ngãi | 94 | 7.19 | 0.4 | 0.067 |
| Vietnam | VNM.8_1 | Bình Định | 94 | 7.19 | 0.4 | 0.067 |
| Vietnam | VNM.45_1 | Phú Yên | 94 | 7.19 | 0.4 | 0.067 |
| Vietnam | VNM.32_1 | Khánh Hòa | 94 | 7.19 | 0.4 | 0.067 |
| Vietnam | VNM.43_1 | Ninh Thuận | 94 | 7.19 | 0.4 | 0.067 |
| Vietnam | VNM.11_1 | Bình Thuận | 94 | 7.19 | 0.4 | 0.067 |

| Country | adm1_gid | adm1_name | N | Zero-dose Prevalence | CI | AEG |
|---|---|---|---|---|---|---|
| Vietnam | VNM.26_1 | Hà Giang | 139 | 9.11 | 0.276 | 0.164 |
| Vietnam | VNM.14_1 | Cao Bằng | 139 | 9.11 | 0.276 | 0.164 |
| Vietnam | VNM.4_1 | Bắc Kạn | 139 | 9.11 | 0.276 | 0.164 |
| Vietnam | VNM.60_1 | Tuyên Quang | 139 | 9.11 | 0.276 | 0.164 |
| Vietnam | VNM.38_1 | Lào Cai | 139 | 9.11 | 0.276 | 0.164 |
| Vietnam | VNM.63_1 | Yên Bái | 139 | 9.11 | 0.276 | 0.164 |
| Vietnam | VNM.56_1 | Thái Nguyên | 139 | 9.11 | 0.276 | 0.164 |
| Vietnam | VNM.35_1 | Lạng Sơn | 139 | 9.11 | 0.276 | 0.164 |
| Vietnam | VNM.3_1 | Bắc Giang | 139 | 9.11 | 0.276 | 0.164 |
| Vietnam | VNM.44_1 | Phú Thọ | 139 | 9.11 | 0.276 | 0.164 |
| Vietnam | VNM.20_1 | Điện Biên | 139 | 9.11 | 0.276 | 0.164 |
| Vietnam | VNM.36_1 | Lai Châu | 139 | 9.11 | 0.276 | 0.164 |
| Vietnam | VNM.52_1 | Sơn La | 139 | 9.11 | 0.276 | 0.164 |
| Vietnam | VNM.30_1 | Hoà Bình | 139 | 9.11 | 0.276 | 0.164 |
| Vietnam | VNM.27_1 | Hà Nội | 127 | 1.43 | -0.236 | -0.036 |
| Vietnam | VNM.23_1 | Hải Phòng | 127 | 1.43 | -0.236 | -0.036 |
| Vietnam | VNM.62_1 | Vĩnh Phúc | 127 | 1.43 | -0.236 | -0.036 |
| Vietnam | VNM.5_1 | Bắc Ninh | 127 | 1.43 | -0.236 | -0.036 |
| Vietnam | VNM.22_1 | Hải Dương | 127 | 1.43 | -0.236 | -0.036 |
| Vietnam | VNM.31_1 | Hưng Yên | 127 | 1.43 | -0.236 | -0.036 |
| Vietnam | VNM.28_1 | Hà Nam | 127 | 1.43 | -0.236 | -0.036 |
| Vietnam | VNM.40_1 | Nam Định | 127 | 1.43 | -0.236 | -0.036 |
| Vietnam | VNM.55_1 | Thái Bình | 127 | 1.43 | -0.236 | -0.036 |
| Vietnam | VNM.49_1 | Quảng Ninh | 127 | 1.43 | -0.236 | -0.036 |
| Vietnam | VNM.42_1 | Ninh Bình | 127 | 1.43 | -0.236 | -0.036 |
| Vietnam | VNM.25_1 | Hồ Chí Minh | 109 | 6.02 | -0.209 | -0.086 |
| Vietnam | VNM.7_1 | Bà Rịa - Vũng Tàu | 109 | 6.02 | -0.209 | -0.086 |
| Vietnam | VNM.9_1 | Bình Dương | 109 | 6.02 | -0.209 | -0.086 |
| Vietnam | VNM.10_1 | Bình Phước | 109 | 6.02 | -0.209 | -0.086 |
| Vietnam | VNM.17_1 | Đồng Nai | 109 | 6.02 | -0.209 | -0.086 |
| Vietnam | VNM.53_1 | Tây Ninh | 109 | 6.02 | -0.209 | -0.086 |
| Yemen | YEM.10_1 | Amran | 163 | 0 | | 0 |
| Yemen | YEM.11_1 | Dhamar | 146 | 0.79 | 0.103 | 0 |
| Yemen | YEM.12_1 | Hadramawt | 204 | 5.13 | 0.217 | 0.078 |
| Yemen | YEM.13_1 | Hajjah | 137 | 2.59 | 0.389 | 0.04 |
| Yemen | YEM.14_1 | Ibb | 151 | 3.2 | -0.129 | 0.002 |
| Yemen | YEM.15_1 | Lahij | 64 | 1.68 | -0.766 | -0.067 |

| Country | adm1_gid | adm1_name | N | Zero-dose Prevalence | CI | AEG |
|---|---|---|---|---|---|---|
| Yemen | YEM.16_1 | Ma'rib | 23 | 0 | | 0 |
| Yemen | YEM.17_1 | Raymah | 160 | 6.33 | -0.115 | 0.009 |
| Yemen | YEM.18_1 | Sa`dah | 50 | 4.72 | -0.06 | 0 |
| Yemen | YEM.19_1 | San`a' | 292 | 1.81 | 0.056 | -0.001 |
| Yemen | YEM.1_1 | `Adan | 68 | 7.23 | 0.103 | -0.006 |
| Yemen | YEM.20_1 | Shabwah | 53 | 13.23 | 0.528 | 0.434 |
| Yemen | YEM.21_1 | Ta`izz | 99 | 3.28 | 0.559 | 0.108 |
| Yemen | YEM.2_1 | Abyan | 54 | 0 | | 0 |
| Yemen | YEM.3_1 | Al Bayda' | 60 | 6.77 | -0.367 | -0.111 |
| Yemen | YEM.4_1 | Al Dali' | 45 | 4.95 | -0.207 | -0.071 |
| Yemen | YEM.5_1 | Al Hudaydah | 164 | 0.69 | 0.848 | 0.033 |
| Yemen | YEM.6_1 | Al Jawf | 13 | 0 | | 0 |
| Yemen | YEM.7_1 | Al Mahrah | 50 | 2.26 | 0.898 | 0.105 |
| Yemen | YEM.8_1 | Al Mahwit | 133 | 1.75 | 0.414 | 0.031 |
| Zambia | ZMB.10_1 | Western | 167 | 3.59 | 0.366 | 0.076 |
| Zambia | ZMB.1_1 | Central | 198 | 0.68 | 0.848 | 0.024 |
| Zambia | ZMB.2_1 | Copperbelt | 175 | 1.09 | 0.674 | 0.053 |
| Zambia | ZMB.3_1 | Eastern | 249 | 1.48 | 0.351 | 0.033 |
| Zambia | ZMB.4_1 | Luapula | 228 | 1.47 | 0.149 | -0.006 |
| Zambia | ZMB.5_1 | Lusaka | 190 | 0.99 | 0.598 | 0.034 |
| Zambia | ZMB.6_1 | Muchinga | 164 | 4.78 | 0.654 | 0.182 |
| Zambia | ZMB.7_1 | North-Western | 156 | 1.93 | 0.545 | 0.011 |
| Zambia | ZMB.8_1 | Northern | 188 | 3.82 | 0.652 | 0.113 |
| Zambia | ZMB.9_1 | Southern | 208 | 1.15 | -0.255 | -0.016 |
| Zimbabwe | ZWE.10_1 | Midlands | 121 | 0 | | 0 |
| Zimbabwe | ZWE.1_1 | Bulawayo | 86 | 0 | | 0 |
| Zimbabwe | ZWE.2_1 | Harare | 107 | 0 | | 0 |
| Zimbabwe | ZWE.3_1 | Manicaland | 128 | 0 | | 0 |
| Zimbabwe | ZWE.4_1 | Mashonaland Central | 100 | 0 | | 0 |
| Zimbabwe | ZWE.5_1 | Mashonaland East | 94 | 0 | | 0 |
| Zimbabwe | ZWE.6_1 | Mashonaland West | 122 | 0.64 | 0.992 | 0.031 |
| Zimbabwe | ZWE.7_1 | Masvingo | 109 | 1.15 | 0.734 | 0.059 |
| Zimbabwe | ZWE.8_1 | Matabeleland North | 118 | 0 | | 0 |
| Zimbabwe | ZWE.9_1 | Matabeleland South | 88 | 0 | | 0 |

**Supplementary Fig.2 Subnational distribution of Absolute Equity Gap (AEG) among zero-dose children across 77 countries, 2015 - 2024**

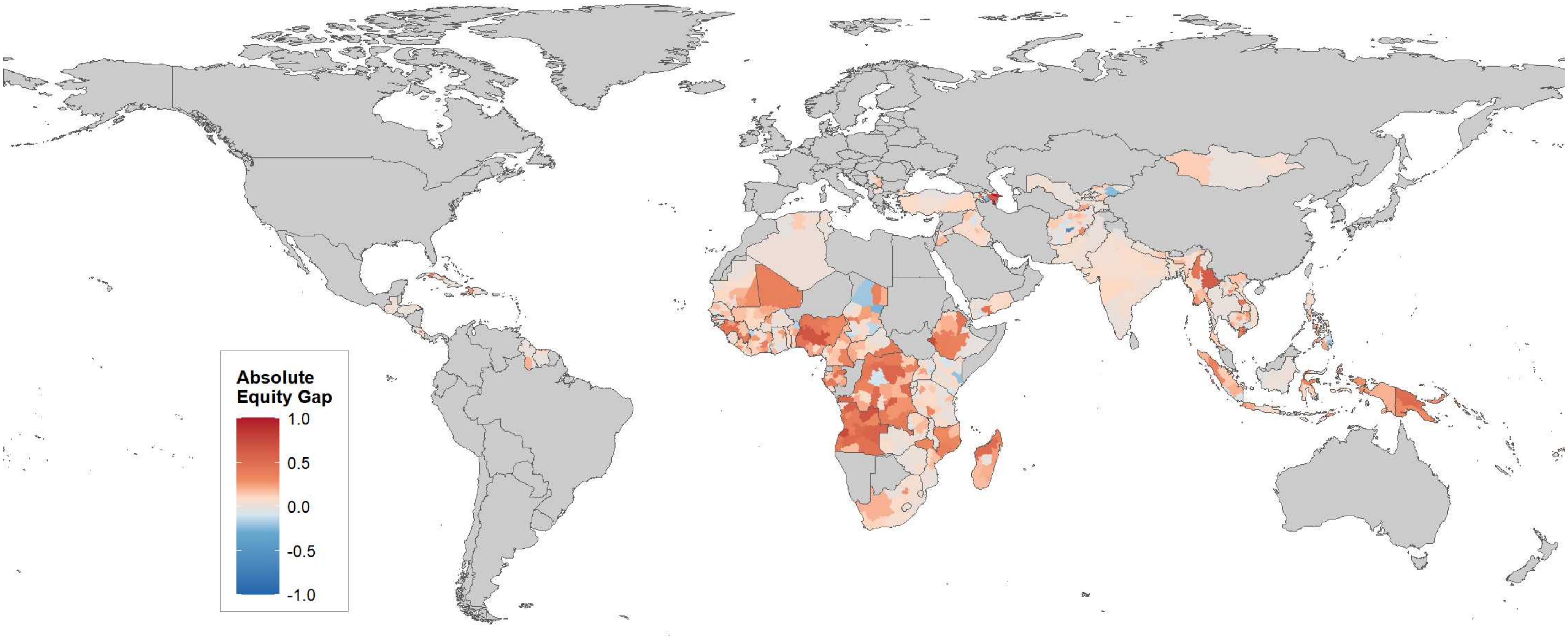

**Supplementary Fig.3 National Equity-Prevalence plane of zero-dose children by World Bank regions across 77 countries**

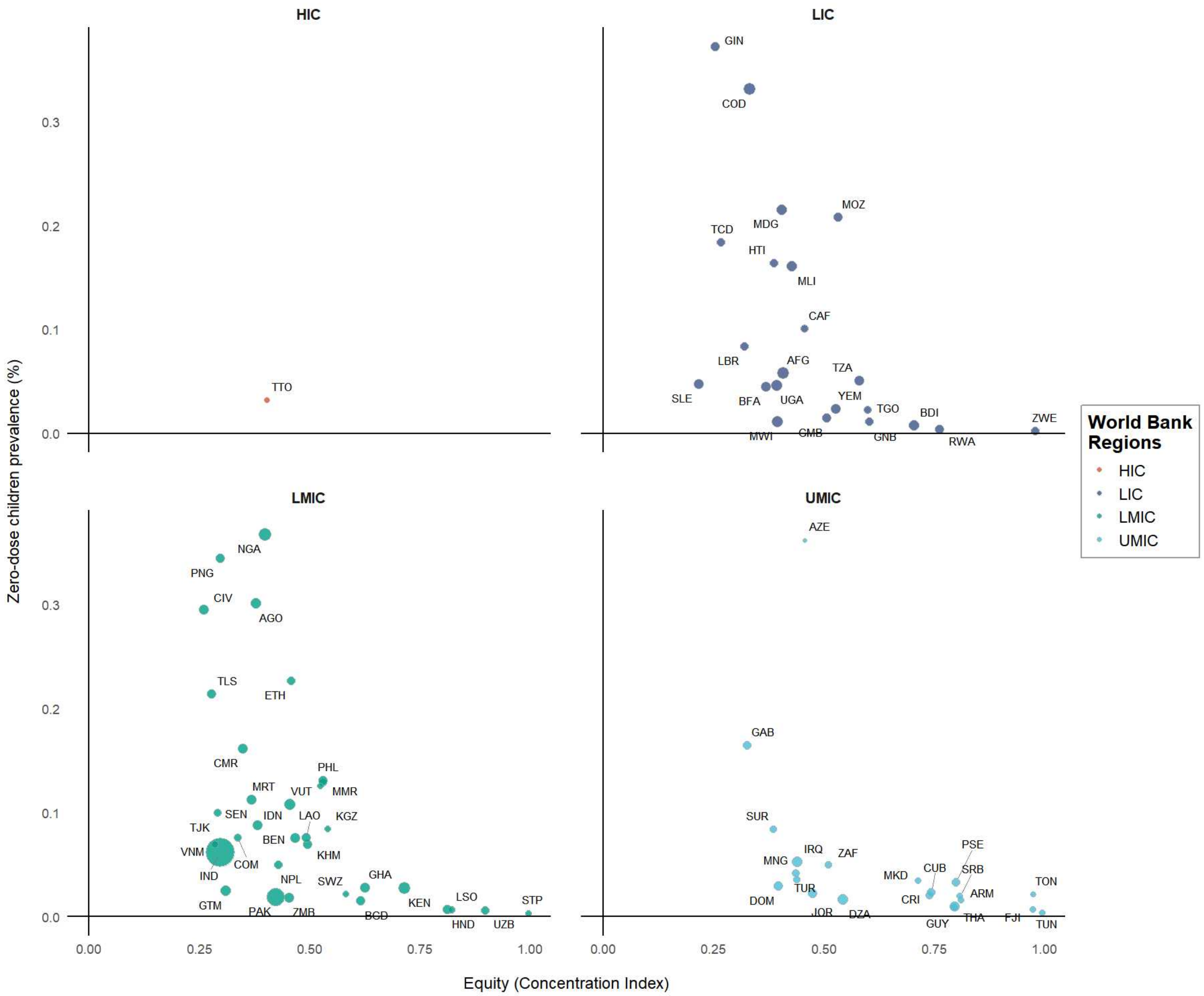

### Supplementary Fig.4 Subnational Equity-Prevalence plane of zero-dose children across 77 countries

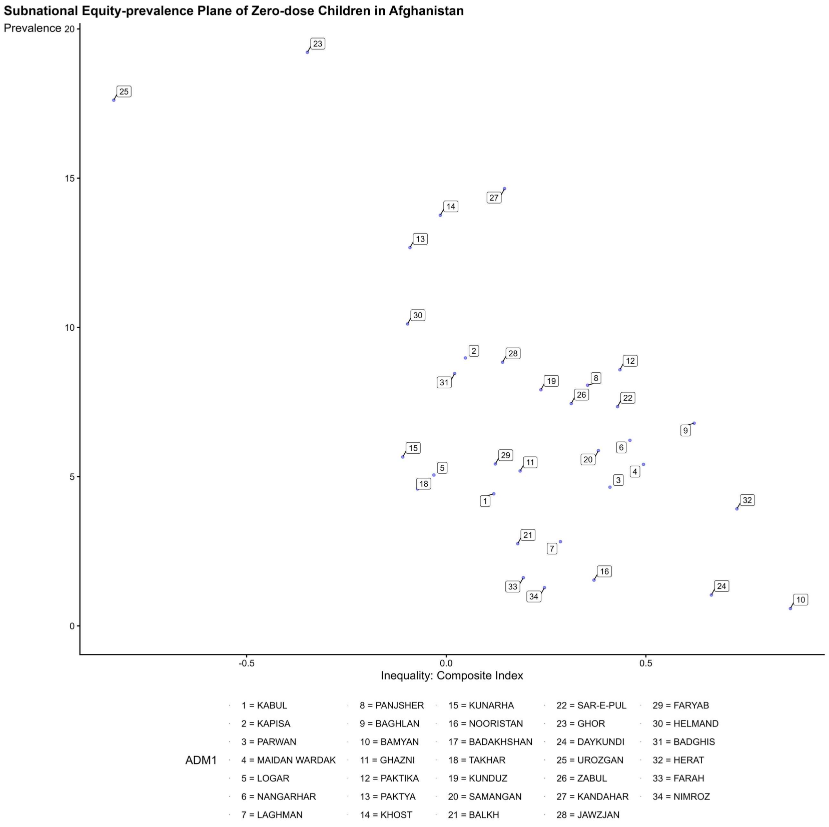

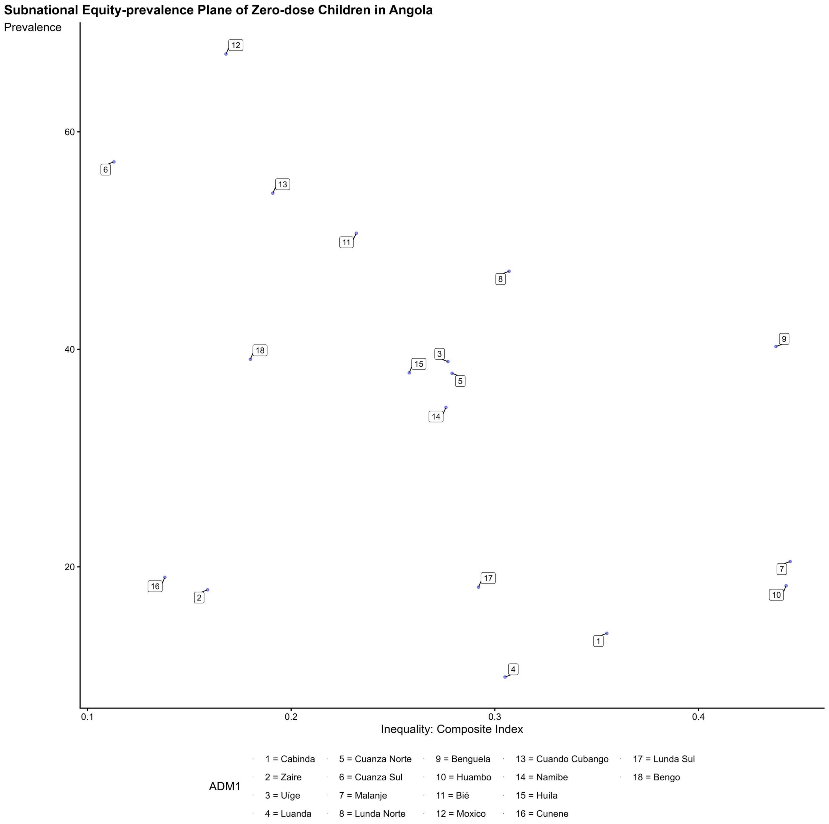

**Subnational Equity-prevalence Plane of Zero-dose Children in Armenia**

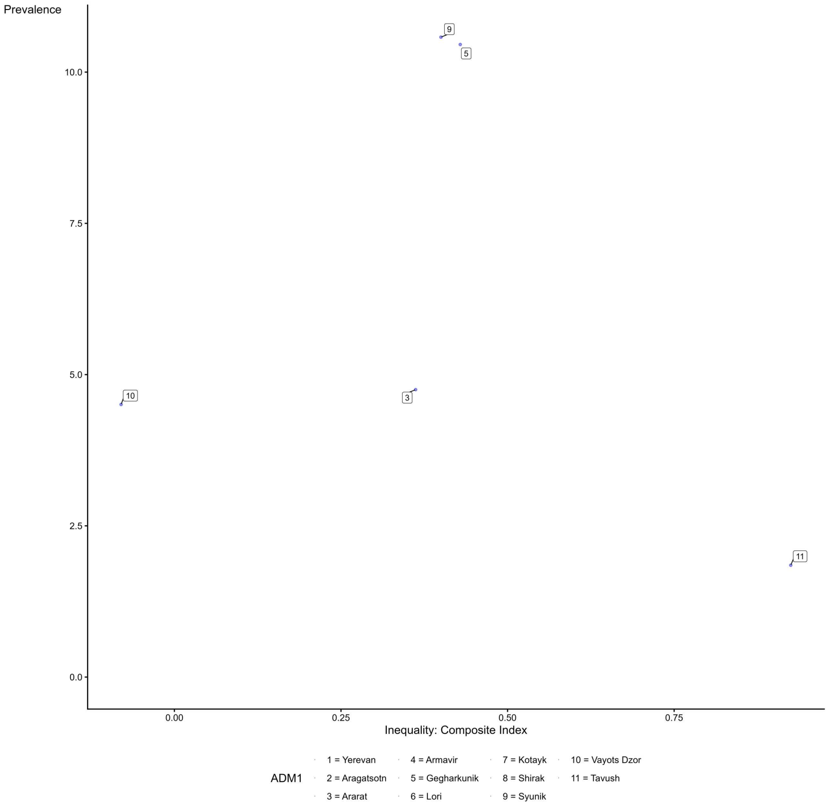

**Subnational Equity-prevalence Plane of Zero-dose Children in Azerbaijan**

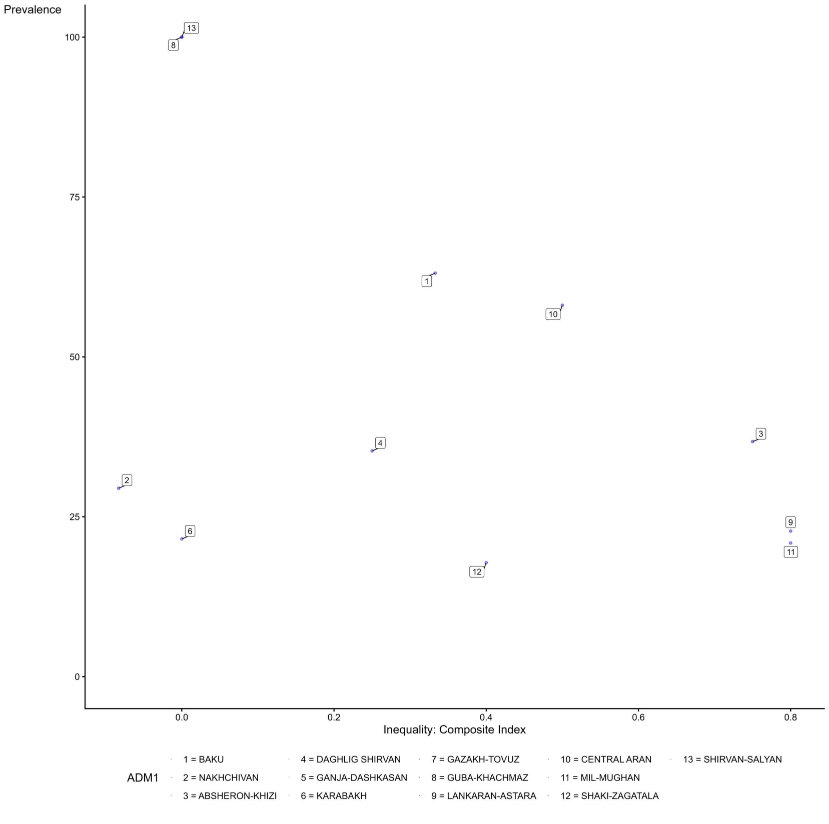

**Subnational Equity-prevalence Plane of Zero-dose Children in Burundi**

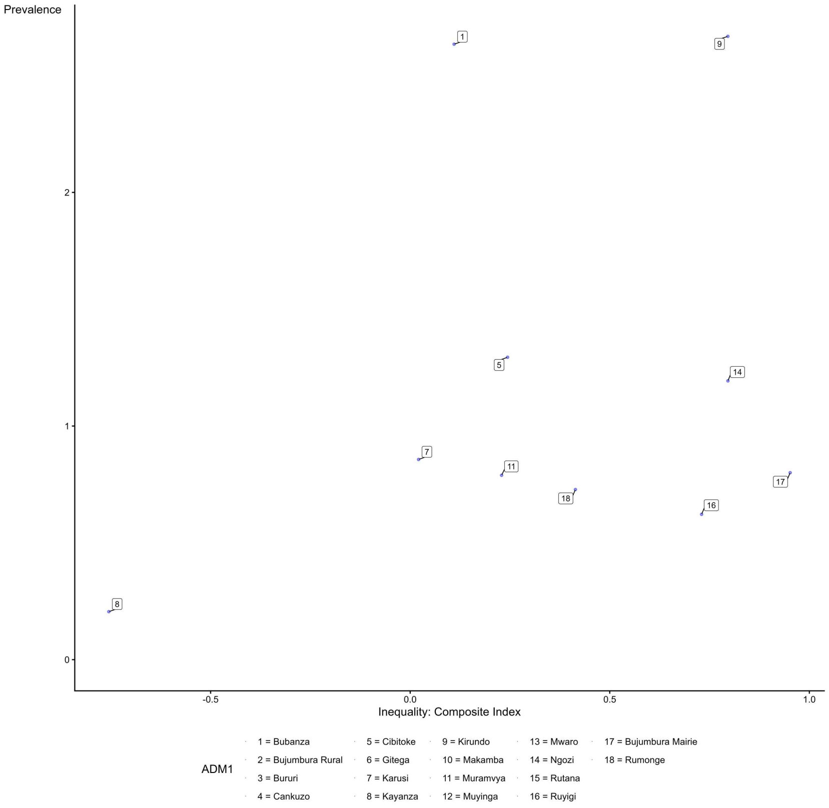

**Subnational Equity-prevalence Plane of Zero-dose Children in Benin**

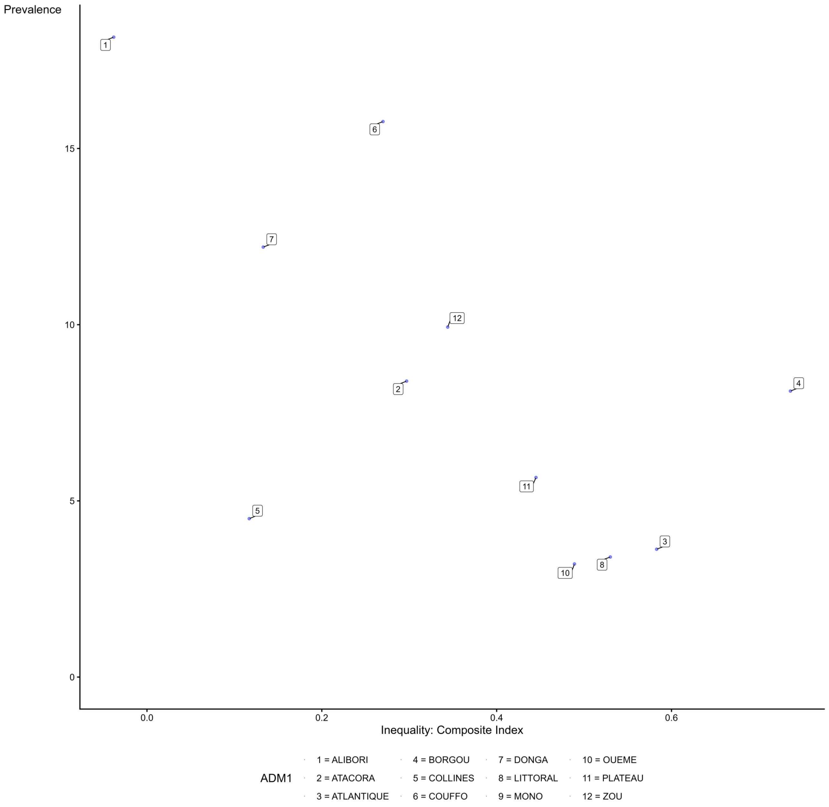

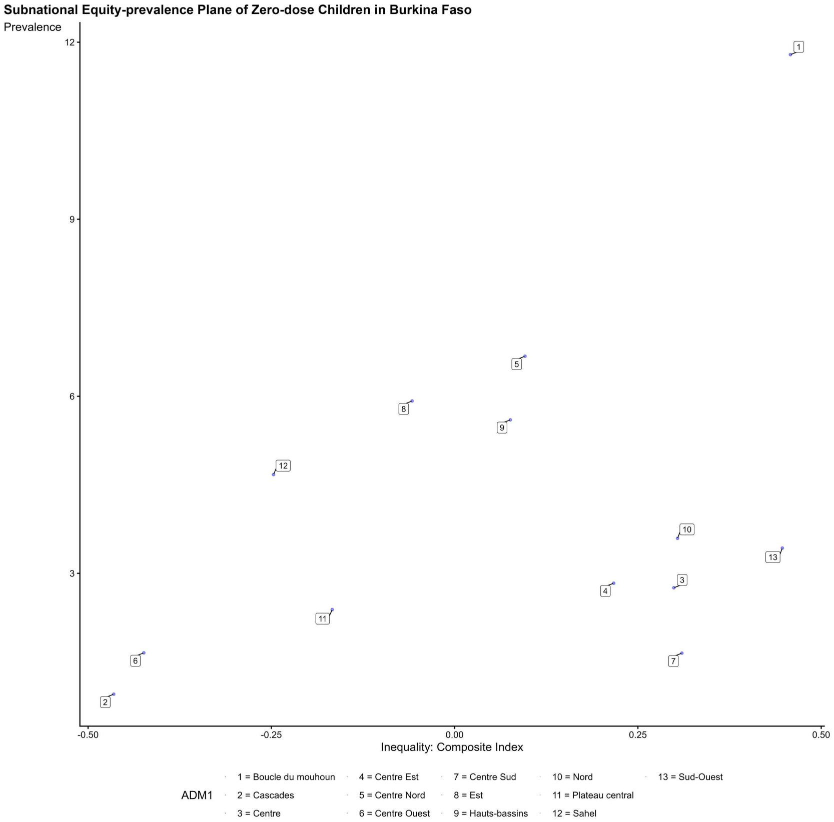
Subnational Equity-prevalence Plane of Zero-dose Children in Burkina Faso
Prevalence
Inequality: Composite Index
ADM1
1 = Boucle du mouhoun
2 = Cascades
3 = Centre
4 = Centre Est
5 = Centre Nord
6 = Centre Ouest
7 = Centre Sud
8 = Est
9 = Hauts-bassins
10 = Nord
11 = Plateau central
12 = Sahel
13 = Sud-Ouest

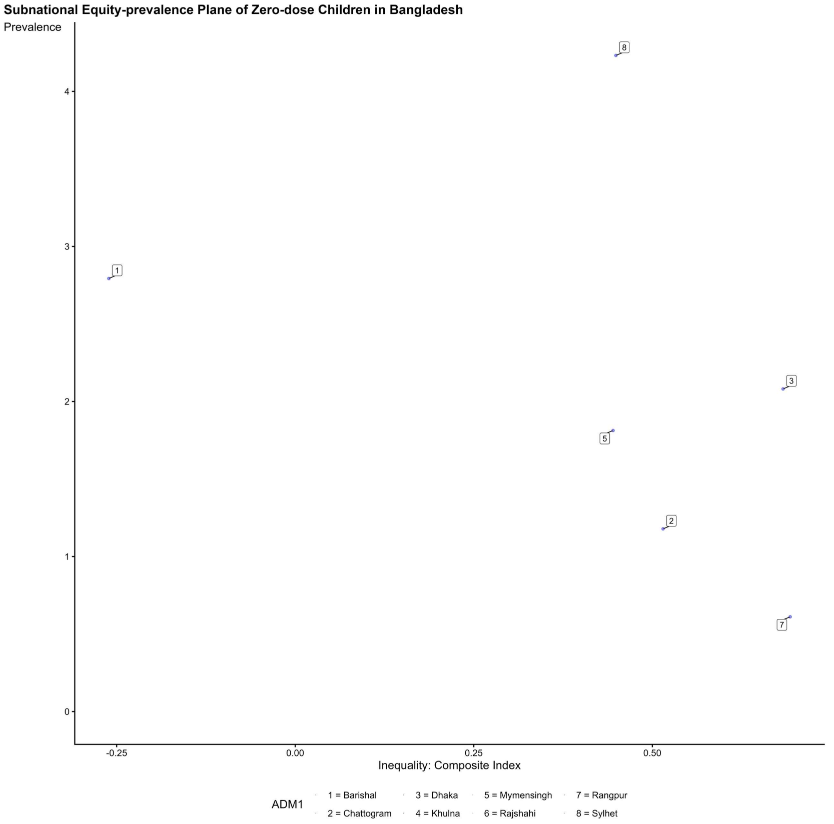
Subnational Equity-prevalence Plane of Zero-dose Children in Bangladesh
Prevalence
Inequality: Composite Index
ADM1
1 = Barishal
2 = Chattogram
3 = Dhaka
4 = Khulna
5 = Mymensingh
6 = Rajshahi
7 = Rangpur
8 = Sylhet

**Subnational Equity-prevalence Plane of Zero-dose Children in Central African Republic**

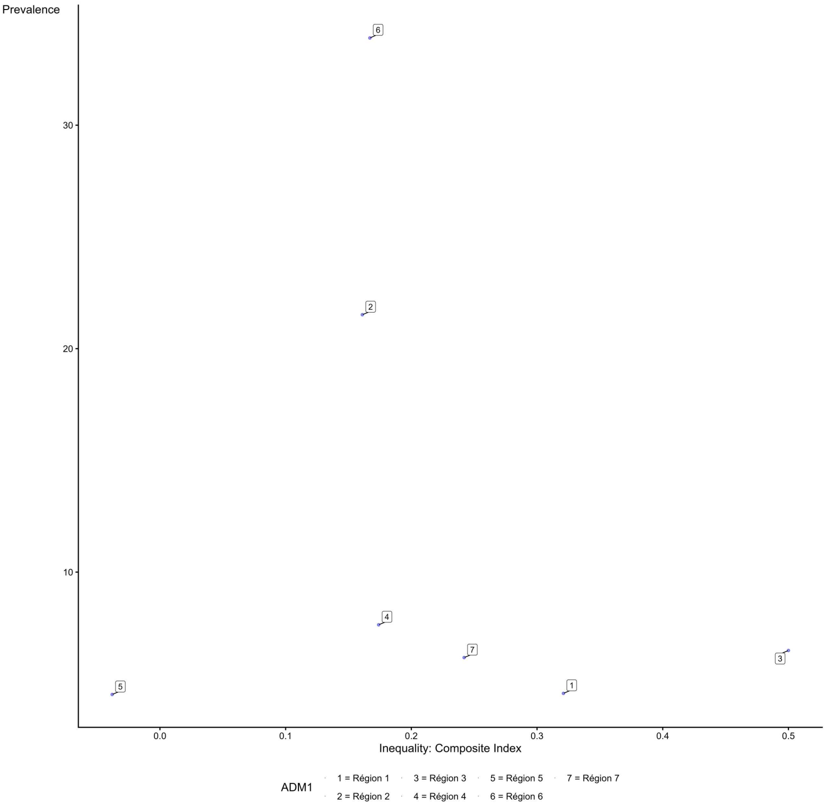

**Subnational Equity-prevalence Plane of Zero-dose Children in Côte d'Ivoire**

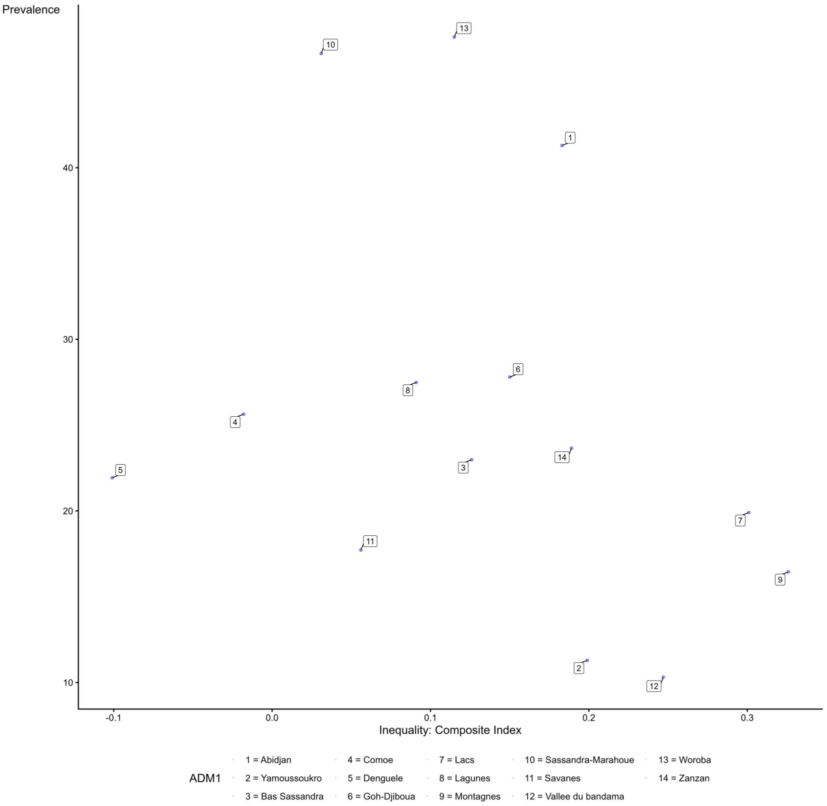

**Subnational Equity-prevalence Plane of Zero-dose Children in Cameroon**

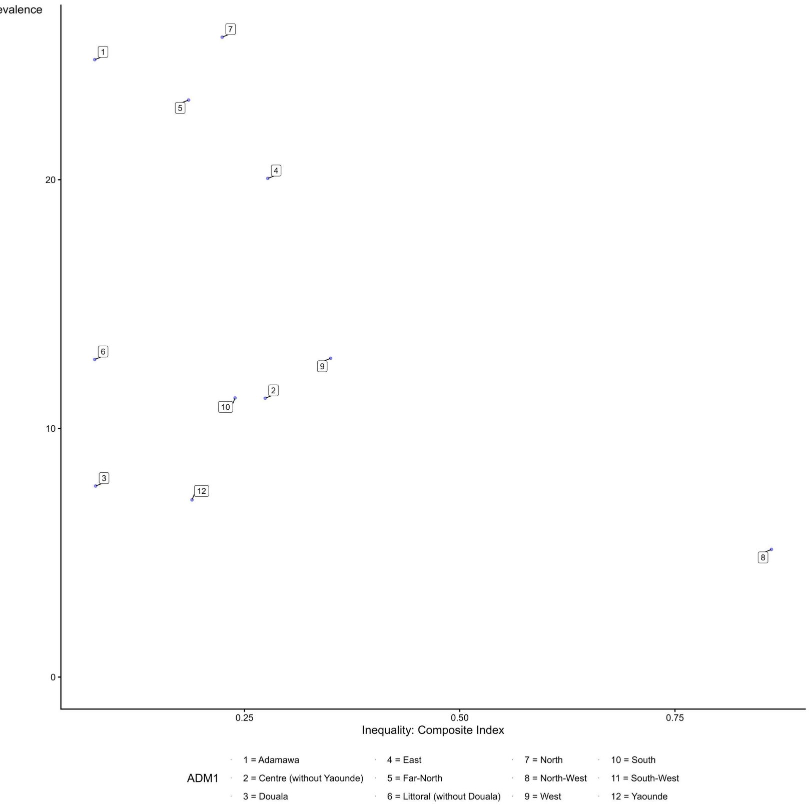

**Subnational Equity-prevalence Plane of Zero-dose Children in Democratic Republic of the Congo**

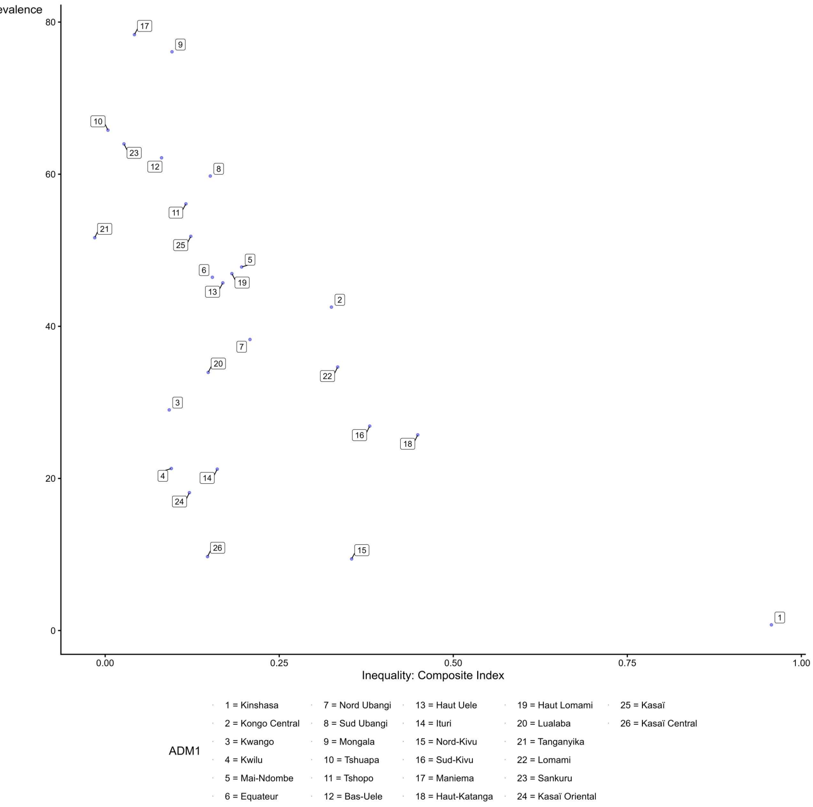

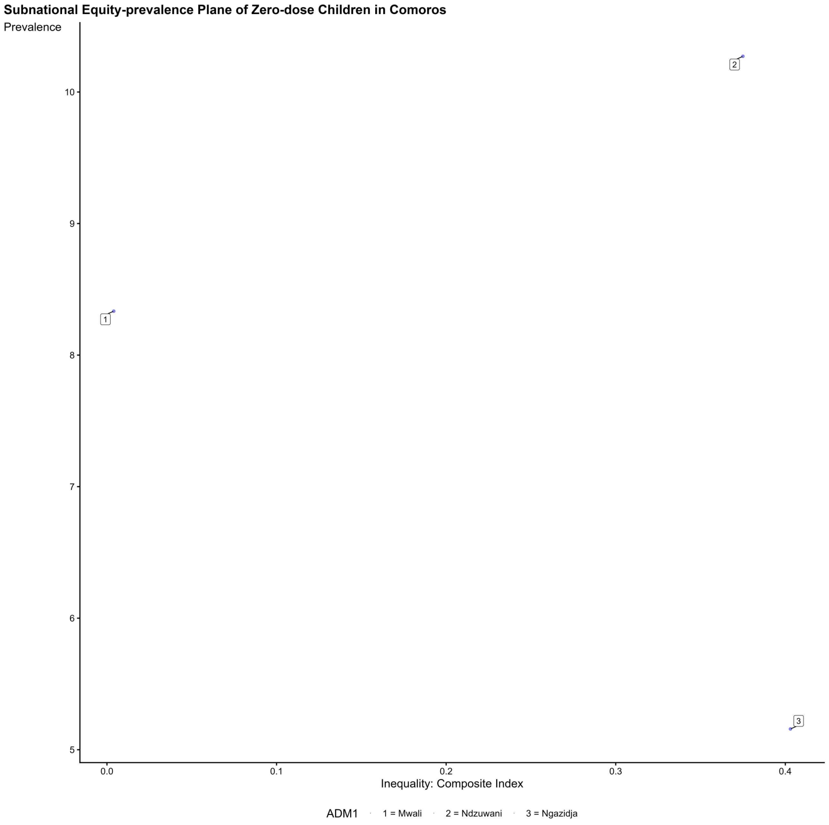
Subnational Equity-prevalence Plane of Zero-dose Children in Comoros
Prevalence
Inequality: Composite Index
ADM1
1 = Mwali
2 = Ndzuwani
3 = Ngazidja

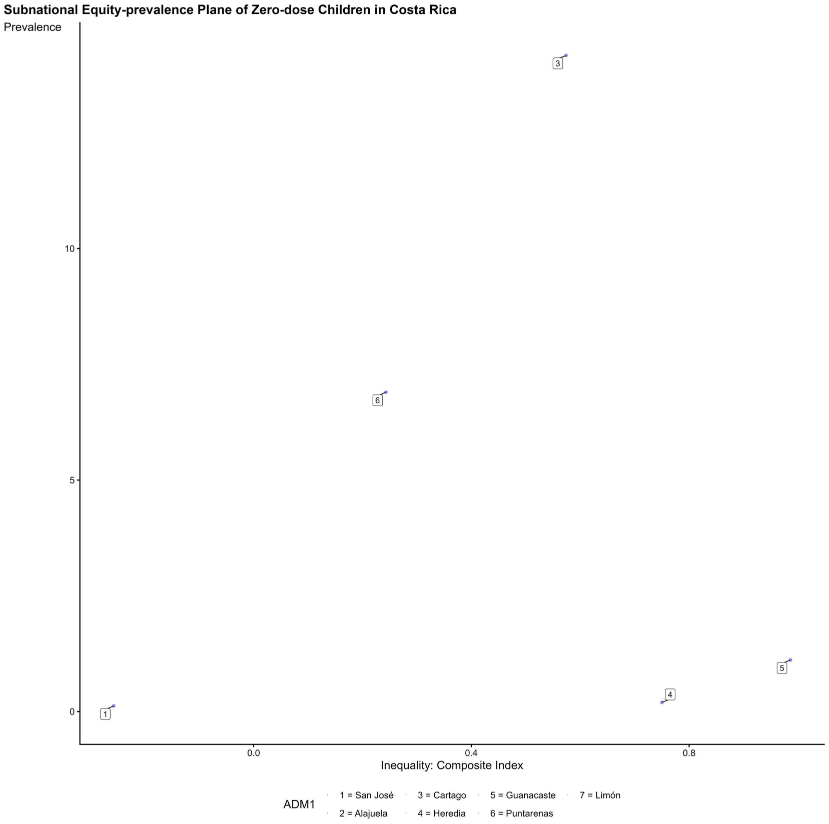
Subnational Equity-prevalence Plane of Zero-dose Children in Costa Rica
Prevalence
Inequality: Composite Index
ADM1
1 = San José
2 = Alajuela
3 = Cartago
4 = Heredia
5 = Guanacaste
6 = Puntarenas
7 = Limón

**Subnational Equity-prevalence Plane of Zero-dose Children in Cuba**

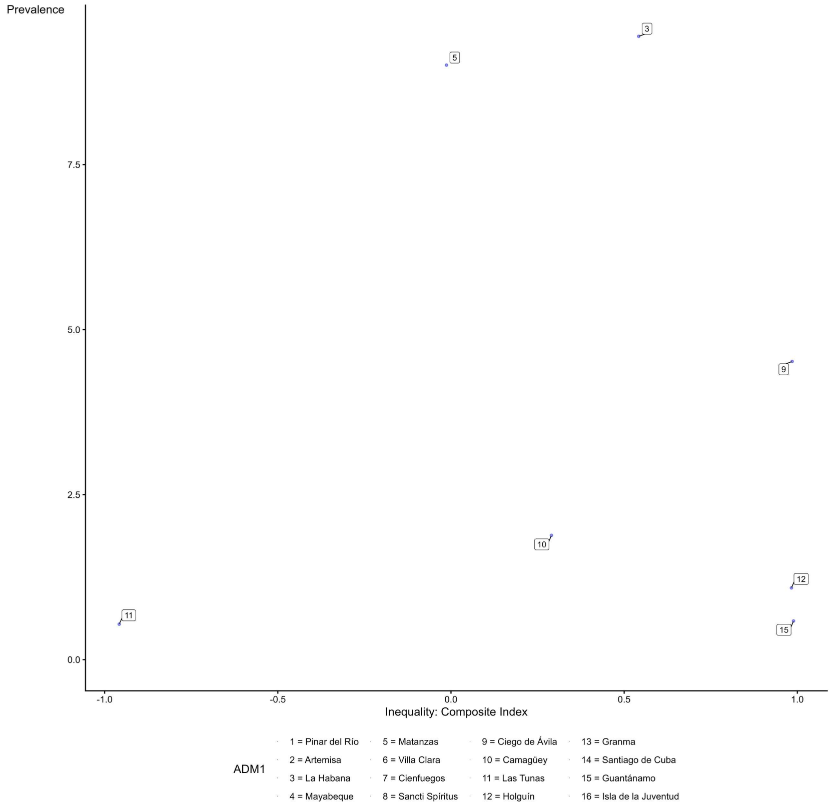

**Subnational Equity-prevalence Plane of Zero-dose Children in Dominican Republic**

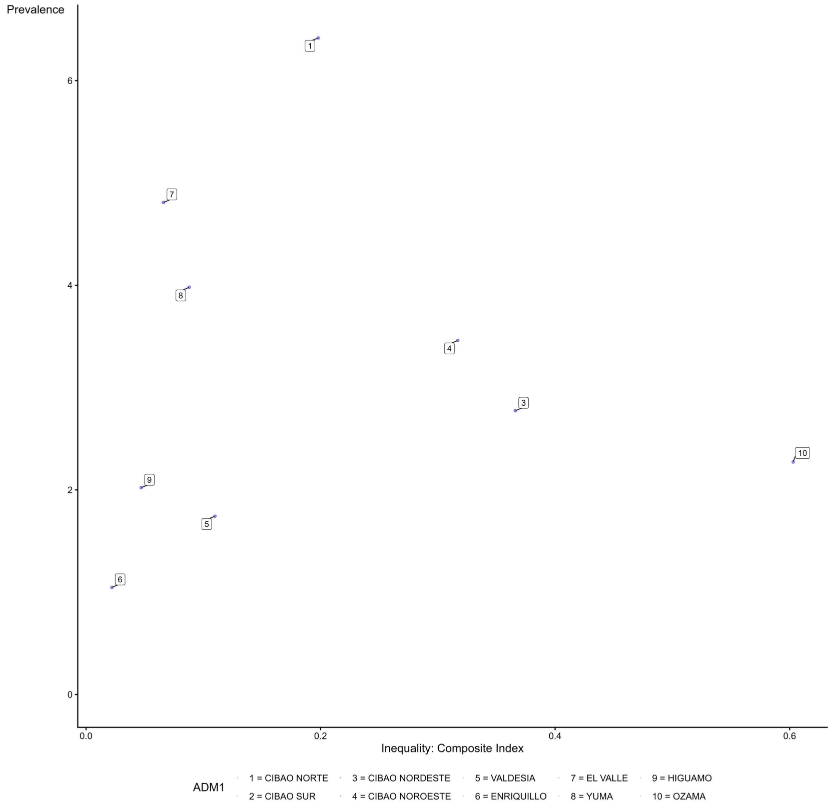

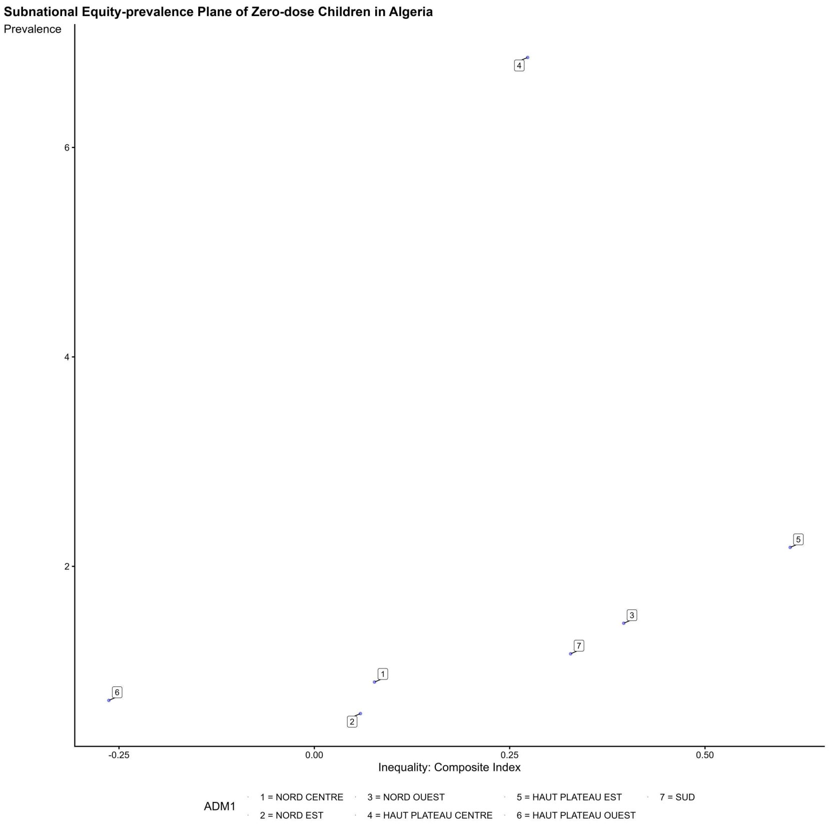
Subnational Equity-prevalence Plane of Zero-dose Children in Algeria
Prevalence
Inequality: Composite Index
-0.25
0.00
0.25
0.50
2
4
6
ADM1
1 = NORD CENTRE
2 = NORD EST
3 = NORD OUEST
4 = HAUT PLATEAU CENTRE
5 = HAUT PLATEAU EST
6 = HAUT PLATEAU OUEST
7 = SUD

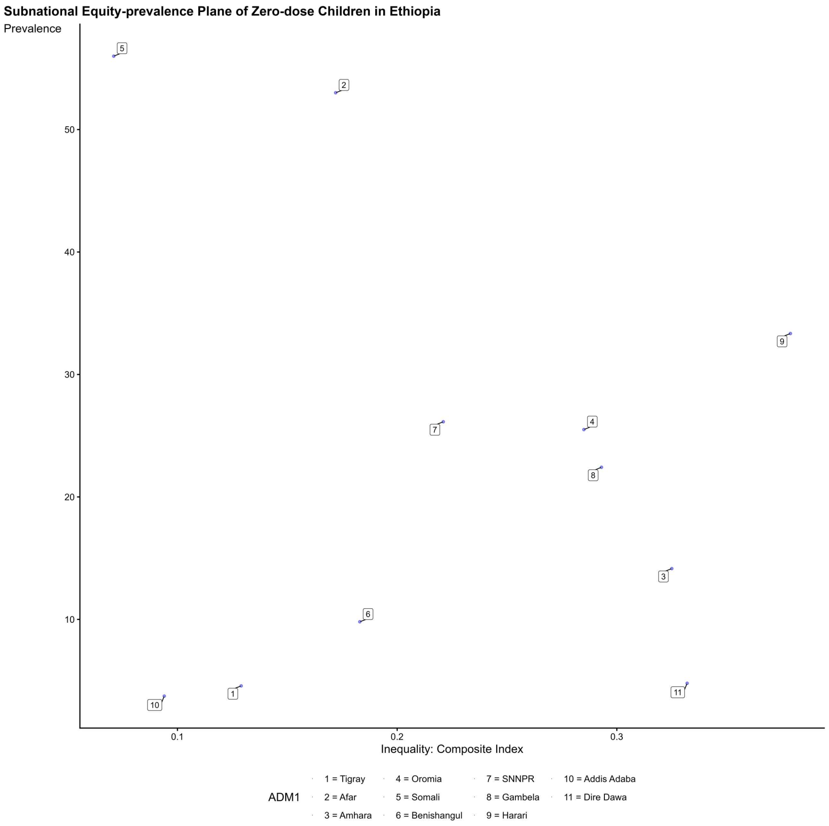
Subnational Equity-prevalence Plane of Zero-dose Children in Ethiopia
Prevalence
Inequality: Composite Index
0.1
0.2
0.3
10
20
30
40
50
ADM1
1 = Tigray
2 = Afar
3 = Amhara
4 = Oromia
5 = Somali
6 = Benishangul
7 = SNNPR
8 = Gambela
9 = Harari
10 = Addis Adaba
11 = Dire Dawa

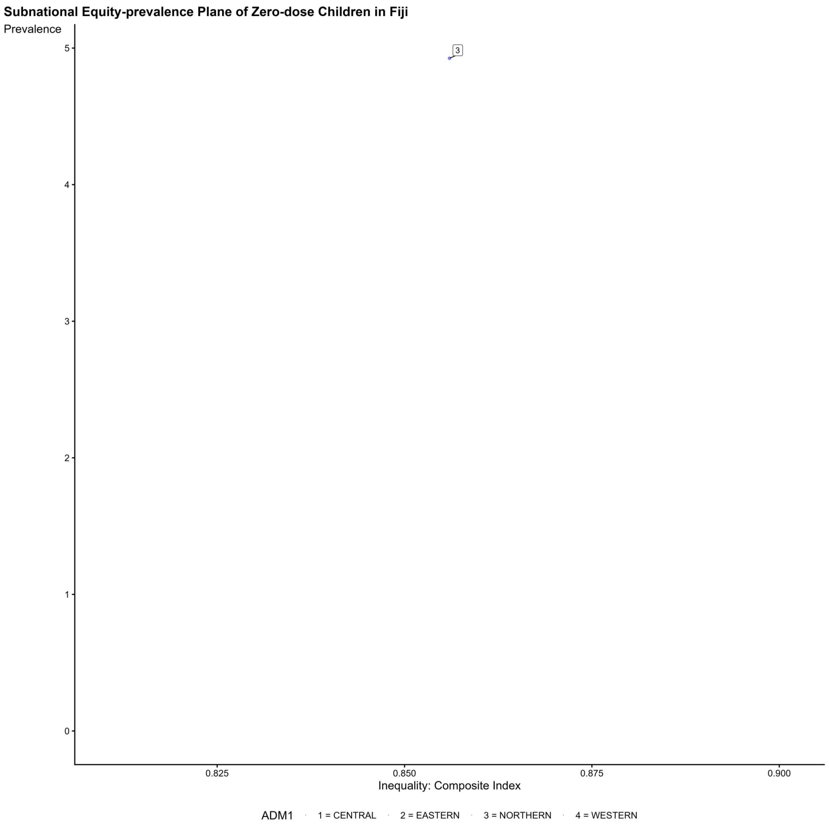
Subnational Equity-prevalence Plane of Zero-dose Children in Fiji
Prevalence
5
4
3
2
1
0
3
0.825
0.850
0.875
0.900
Inequality: Composite Index
ADM1
1 = CENTRAL
2 = EASTERN
3 = NORTHERN
4 = WESTERN

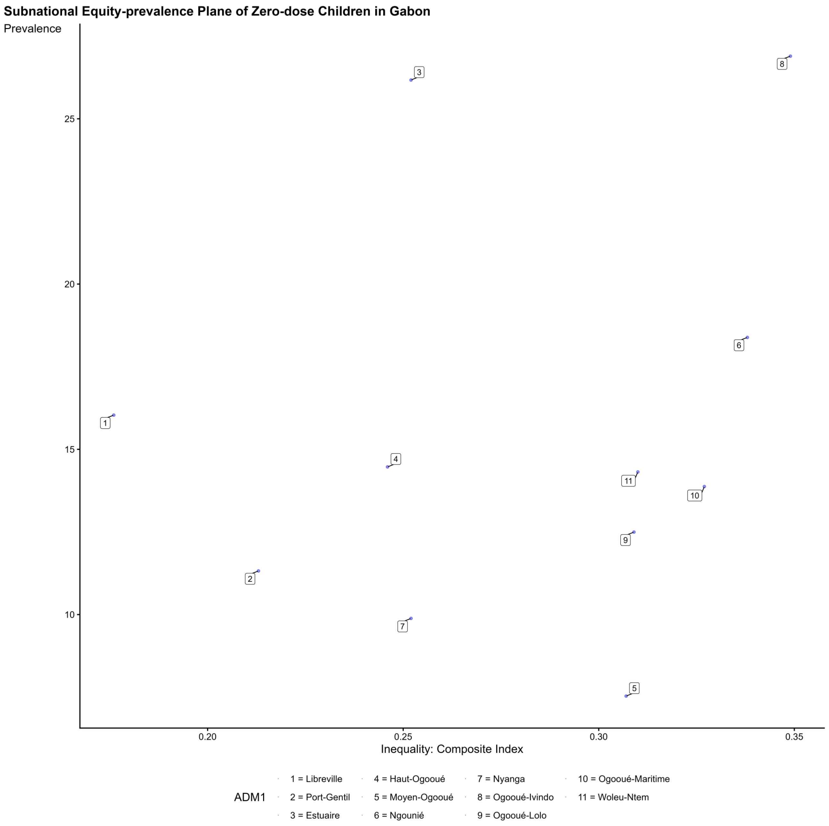
Subnational Equity-prevalence Plane of Zero-dose Children in Gabon
Prevalence
25
20
15
10
0.20
0.25
0.30
0.35
Inequality: Composite Index
ADM1
1 = Libreville
2 = Port-Gentil
3 = Estuaire
4 = Haut-Ogooué
5 = Moyen-Ogooué
6 = Ngounié
7 = Nyanga
8 = Ogooué-Ivindo
9 = Ogooué-Lolo
10 = Ogooué-Maritime
11 = Woleu-Ntem

**Subnational Equity-prevalence Plane of Zero-dose Children in Ghana**

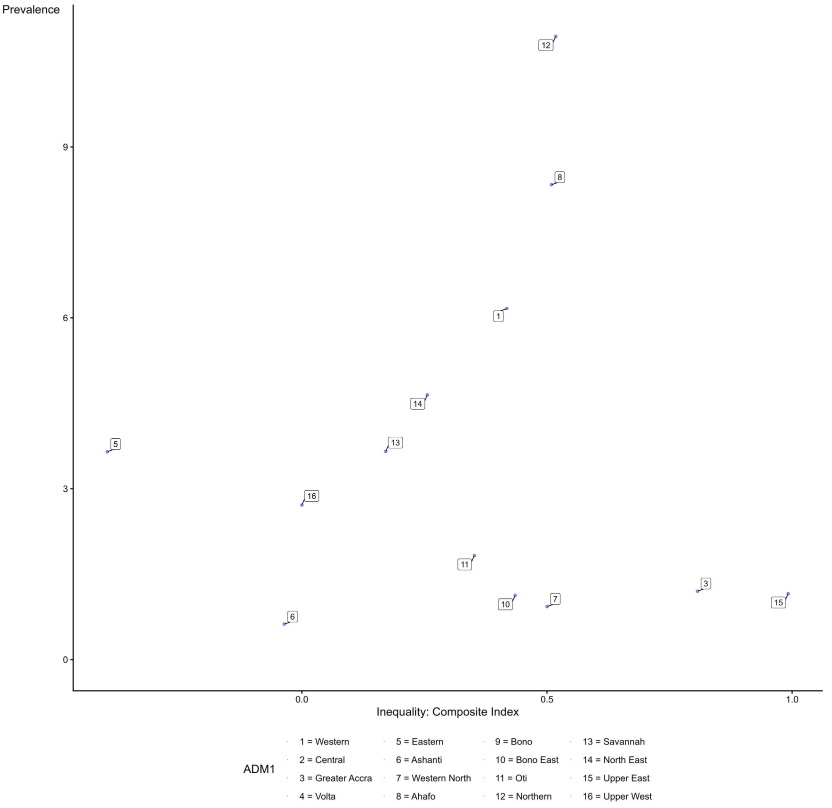

**Subnational Equity-prevalence Plane of Zero-dose Children in Guinea**

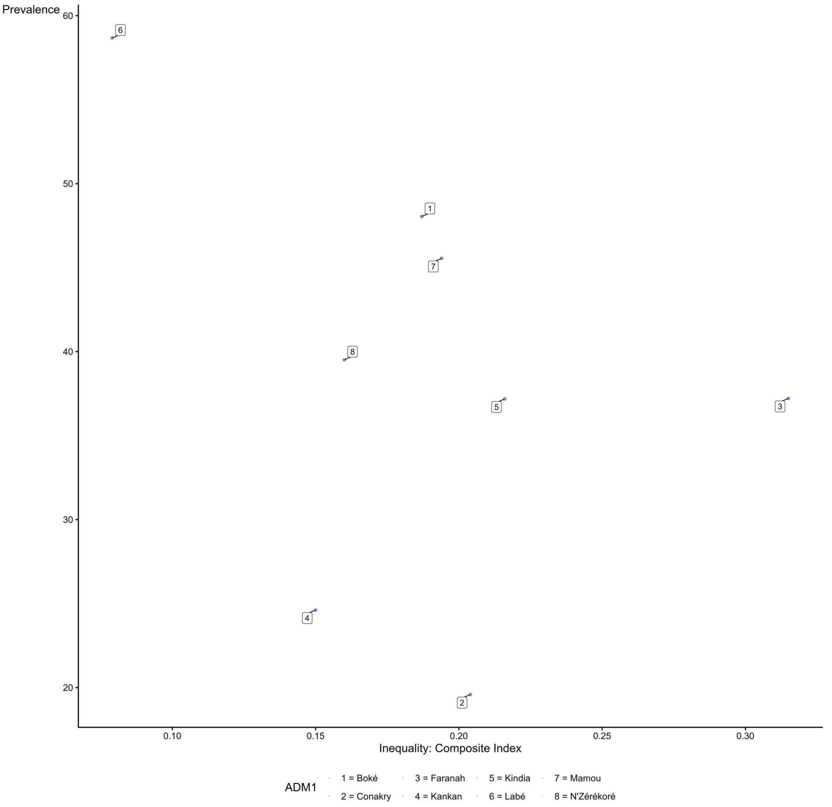

**Subnational Equity-prevalence Plane of Zero-dose Children in Gambia**

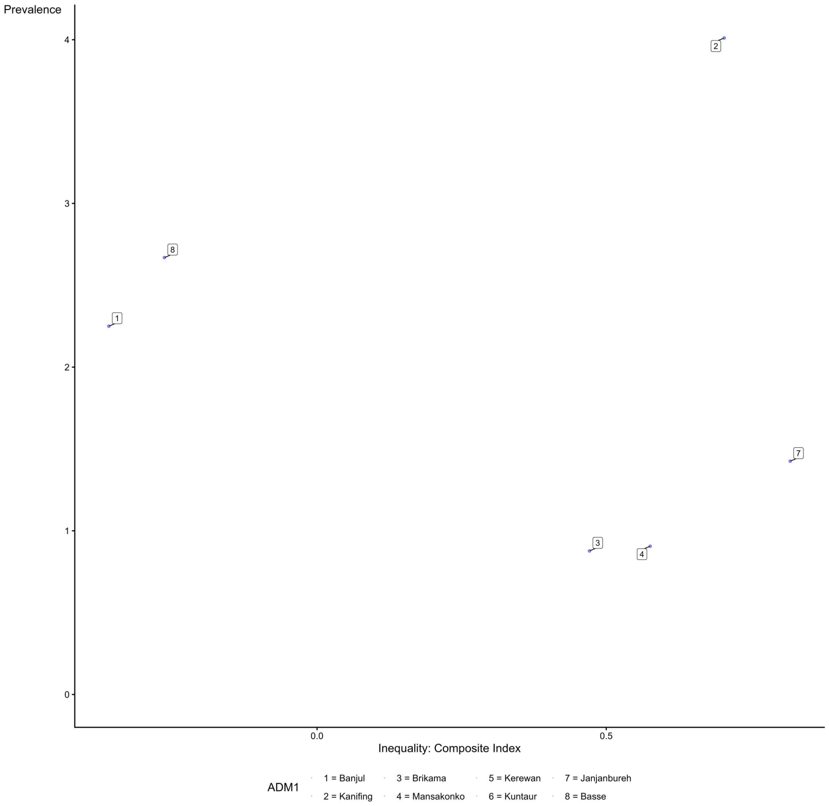

**Subnational Equity-prevalence Plane of Zero-dose Children in Guinea-Bissau**

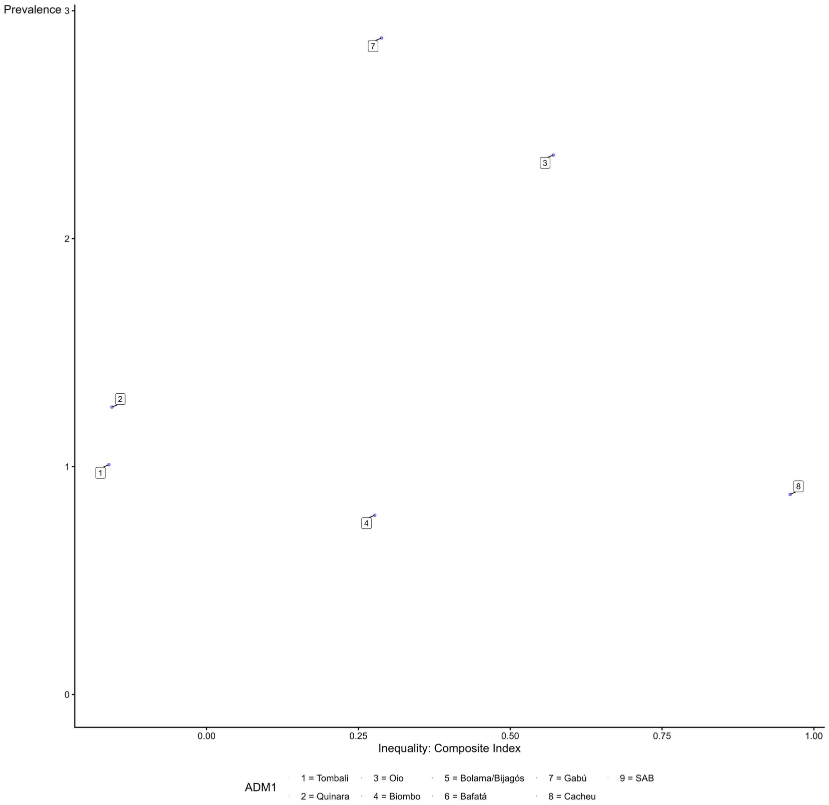

**Subnational Equity-prevalence Plane of Zero-dose Children in Guatemala**

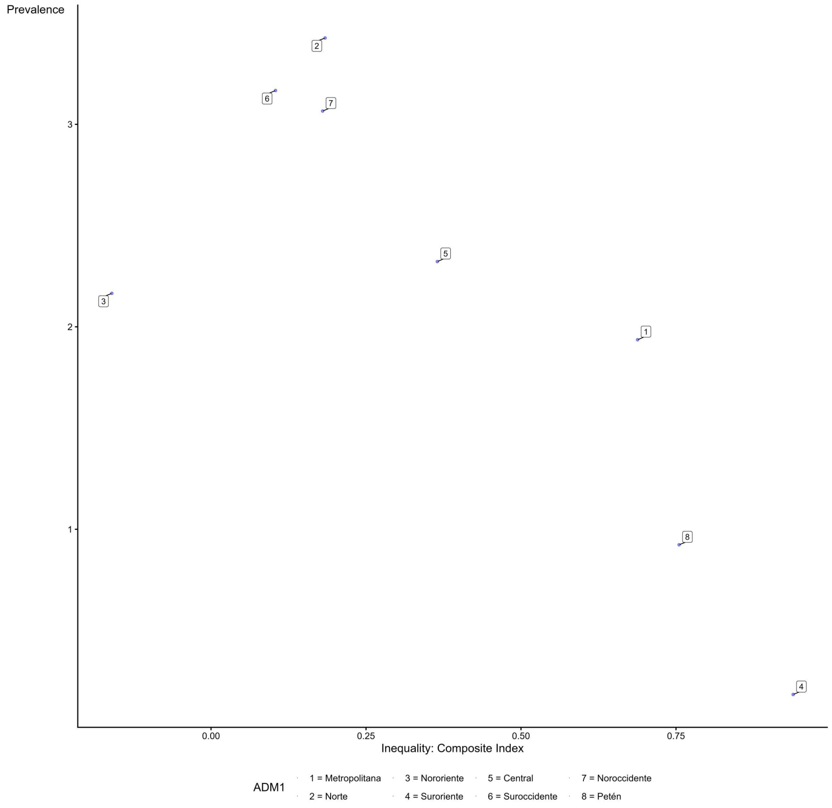

**Subnational Equity-prevalence Plane of Zero-dose Children in Guyana**

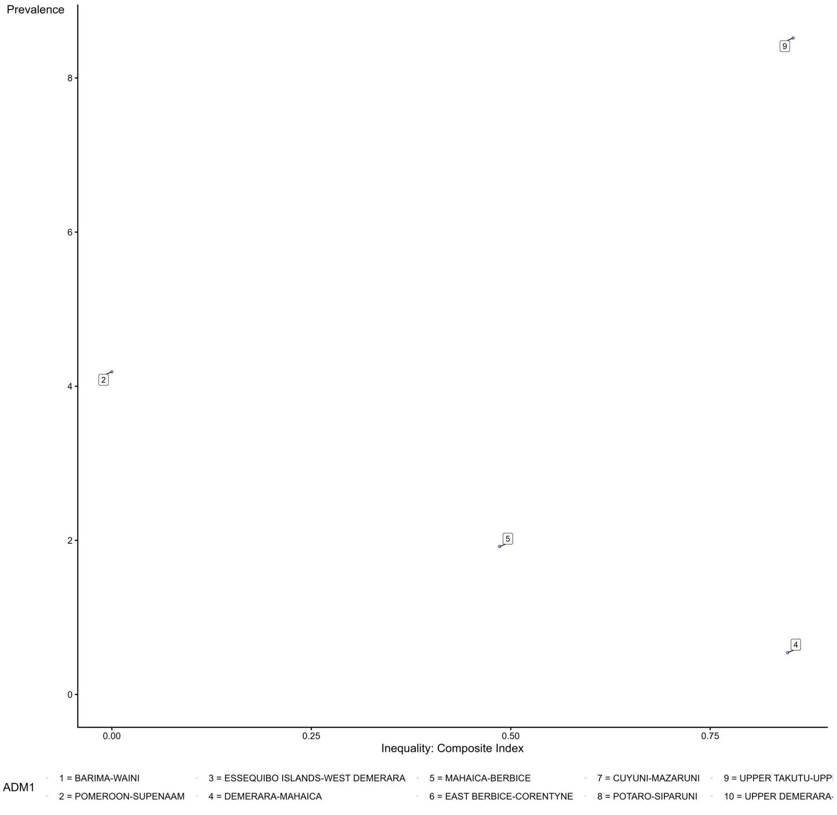

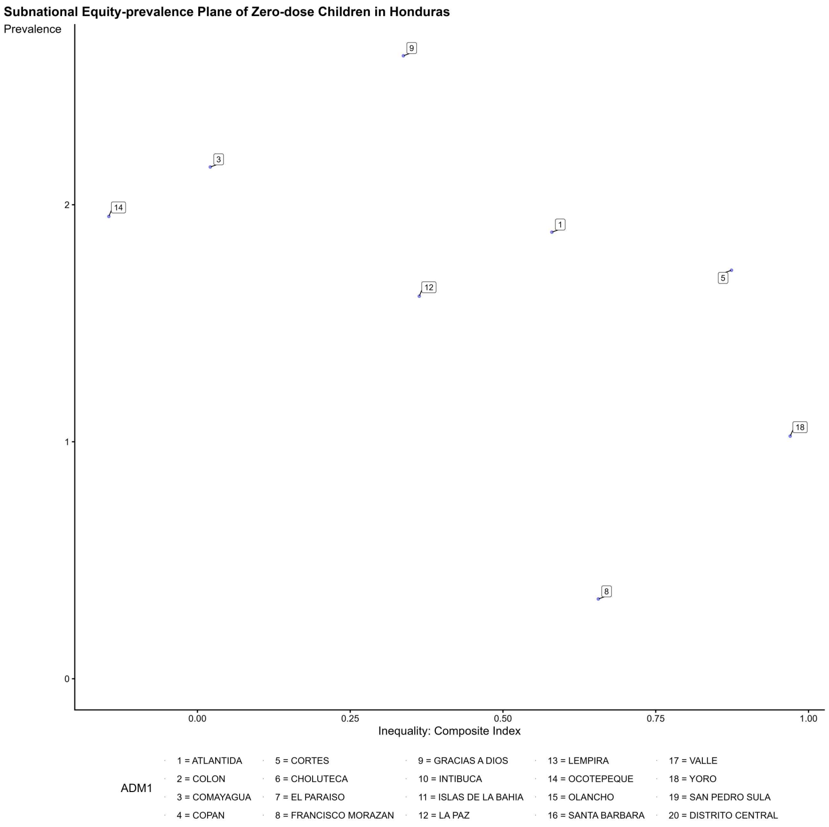
Subnational Equity-prevalence Plane of Zero-dose Children in Honduras
Prevalence
Inequality: Composite Index
ADM1
1 = ATLANTIDA
2 = COLON
3 = COMAYAGUA
4 = COPAN
5 = CORTES
6 = CHOLUTECA
7 = EL PARAISO
8 = FRANCISCO MORAZAN
9 = GRACIAS A DIOS
10 = INTIBUCA
11 = ISLAS DE LA BAHIA
12 = LA PAZ
13 = LEMPIRA
14 = OCOTEPEQUE
15 = OLANCHO
16 = SANTA BARBARA
17 = VALLE
18 = YORO
19 = SAN PEDRO SULA
20 = DISTRITO CENTRAL

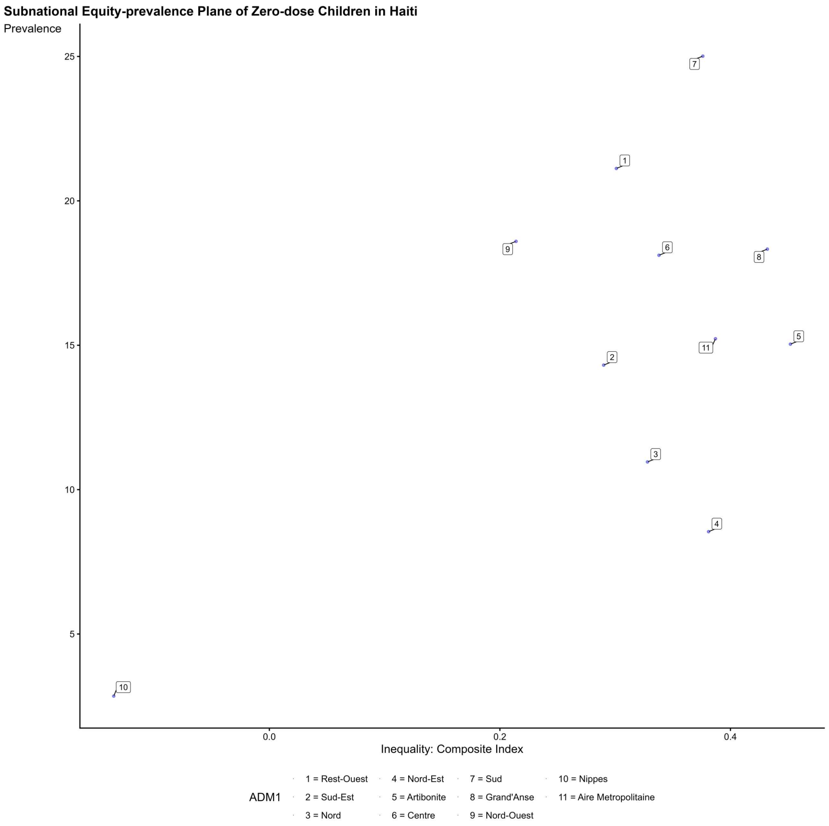
Subnational Equity-prevalence Plane of Zero-dose Children in Haiti
Prevalence
Inequality: Composite Index
ADM1
1 = Rest-Ouest
2 = Sud-Est
3 = Nord
4 = Nord-Est
5 = Artibonite
6 = Centre
7 = Sud
8 = Grand'Anse
9 = Nord-Ouest
10 = Nippes
11 = Aire Metropolitaine

**Subnational Equity-prevalence Plane of Zero-dose Children in Indonesia**

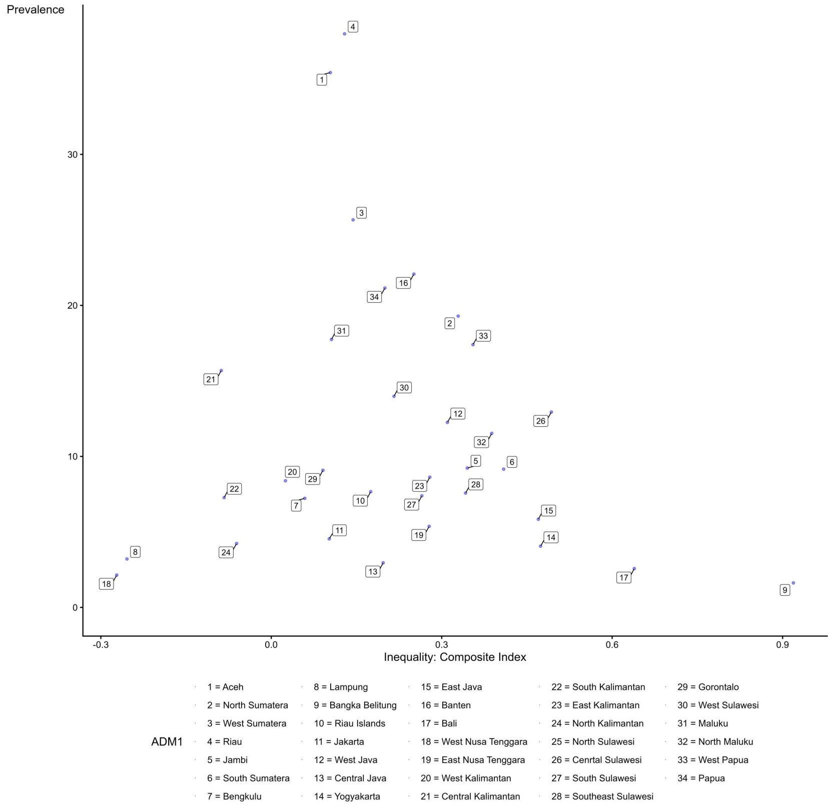

**Subnational Equity-prevalence Plane of Zero-dose Children in India**

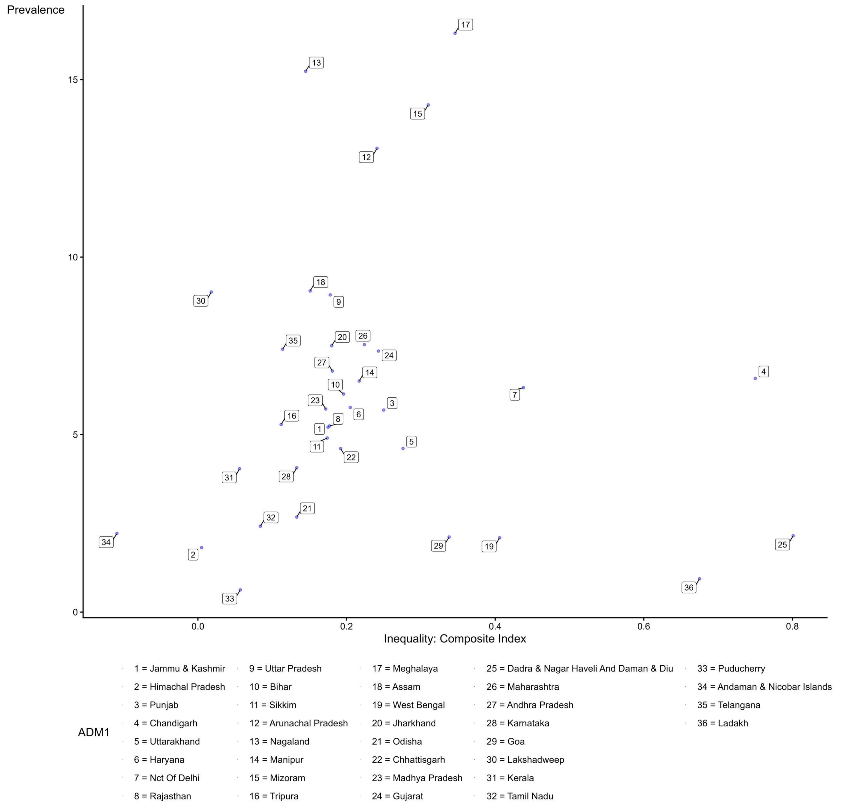

**Subnational Equity-prevalence Plane of Zero-dose Children in Iraq**

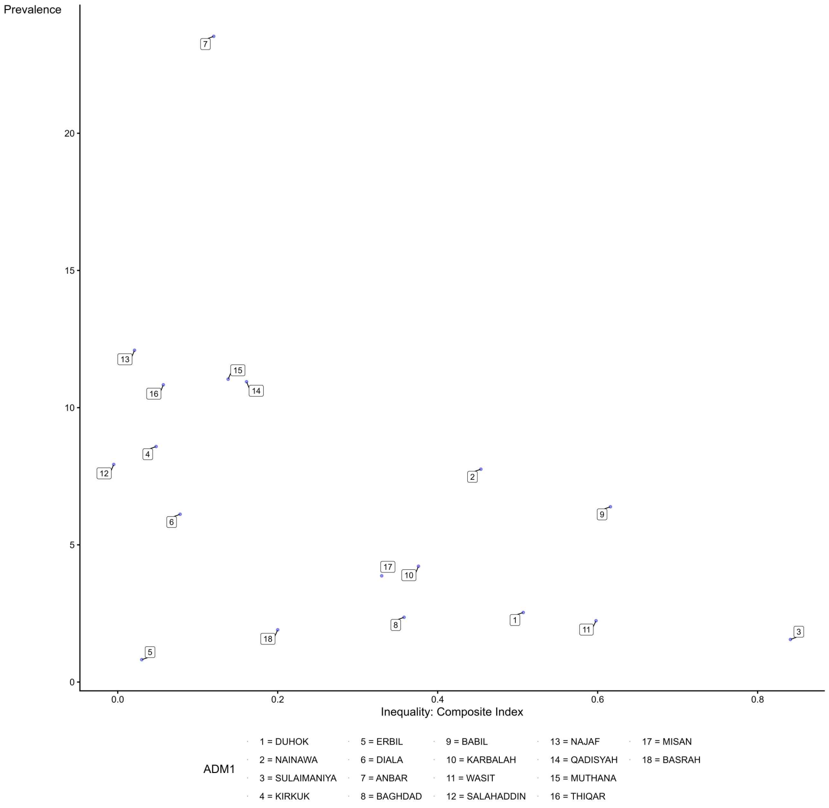

**Subnational Equity-prevalence Plane of Zero-dose Children in Jordan**

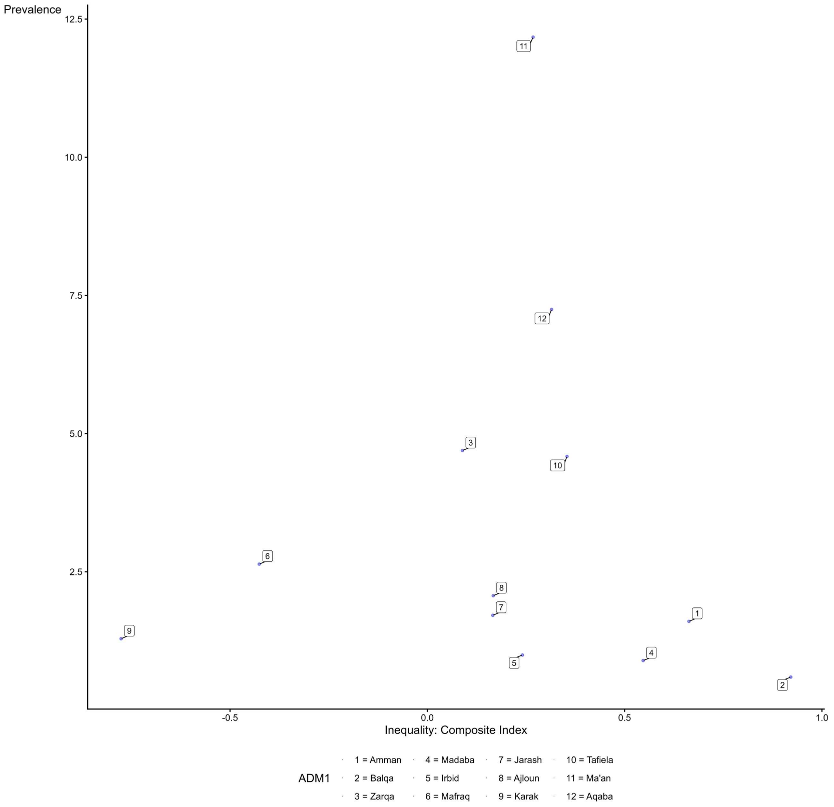

**Subnational Equity-prevalence Plane of Zero-dose Children in Kenya**

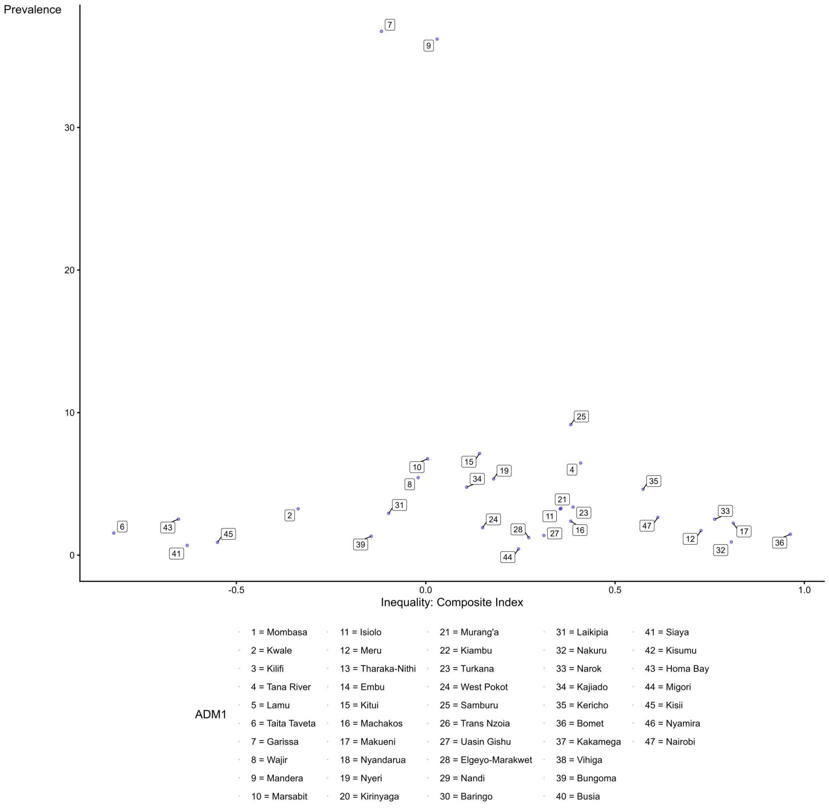

**Subnational Equity-prevalence Plane of Zero-dose Children in Kyrgyzstan**

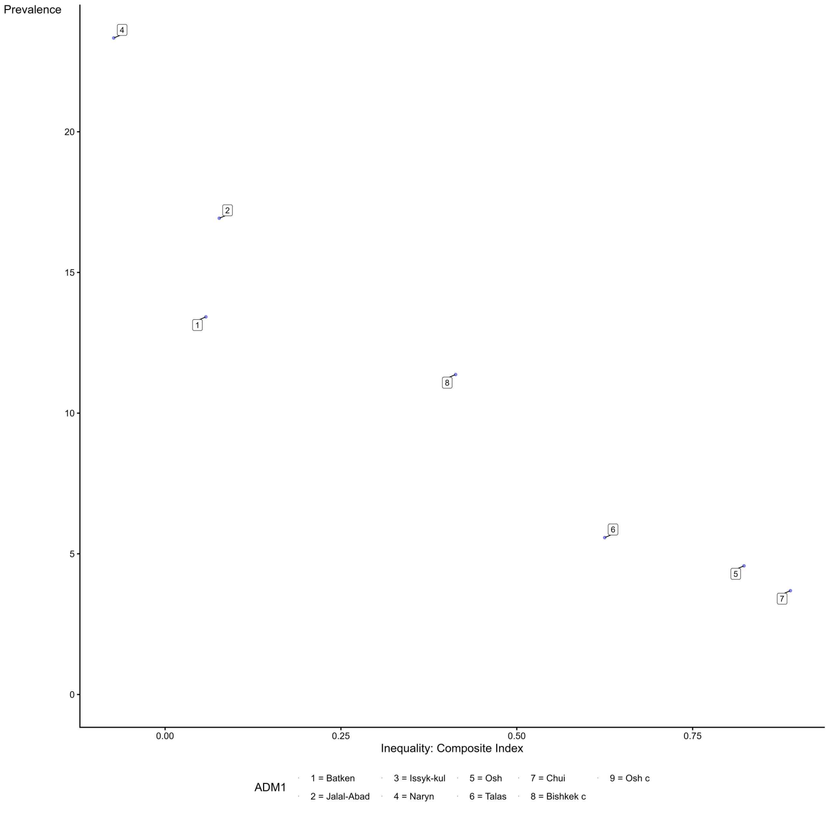

**Subnational Equity-prevalence Plane of Zero-dose Children in Cambodia**

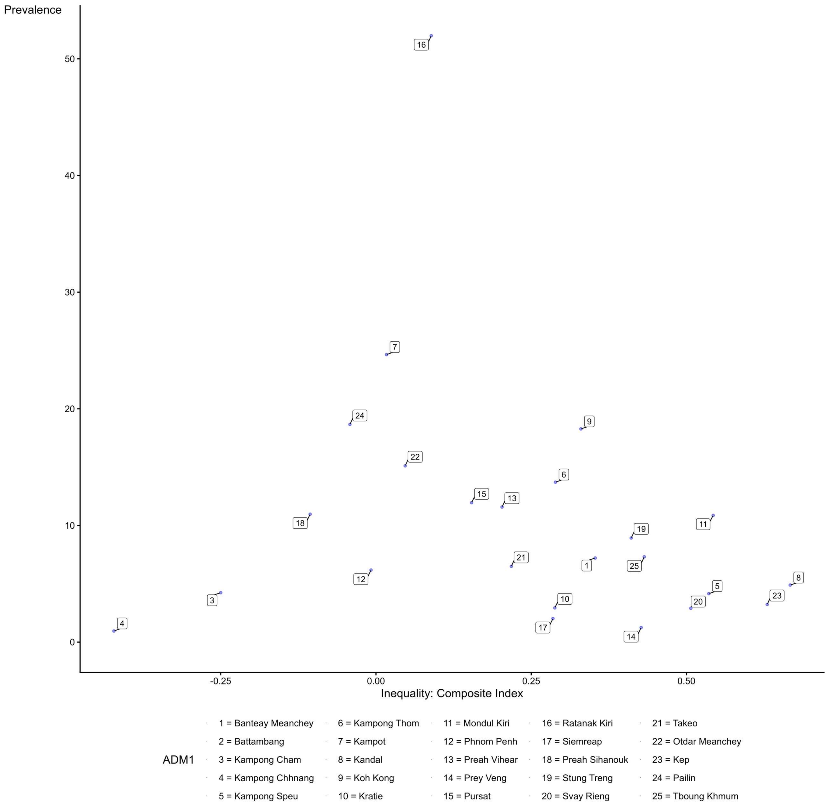

**Subnational Equity-prevalence Plane of Zero-dose Children in Laos**

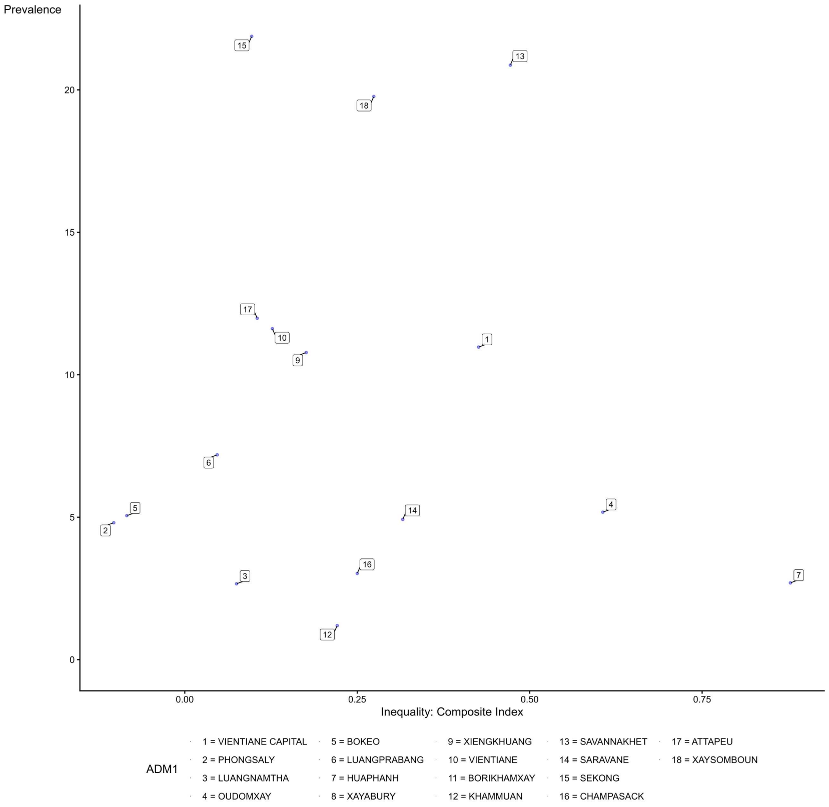

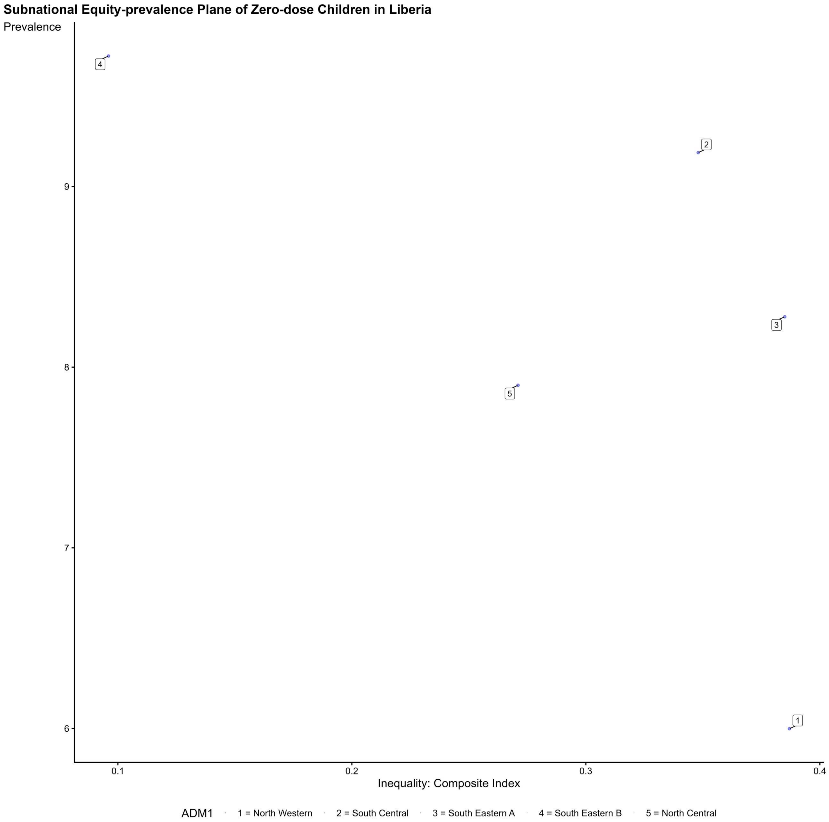

Subnational Equity-prevalence Plane of Zero-dose Children in Liberia
Prevalence
9
8
7
6
0.1
0.2
0.3
0.4
Inequality: Composite Index
ADM1
1 = North Western
2 = South Central
3 = South Eastern A
4 = South Eastern B
5 = North Central

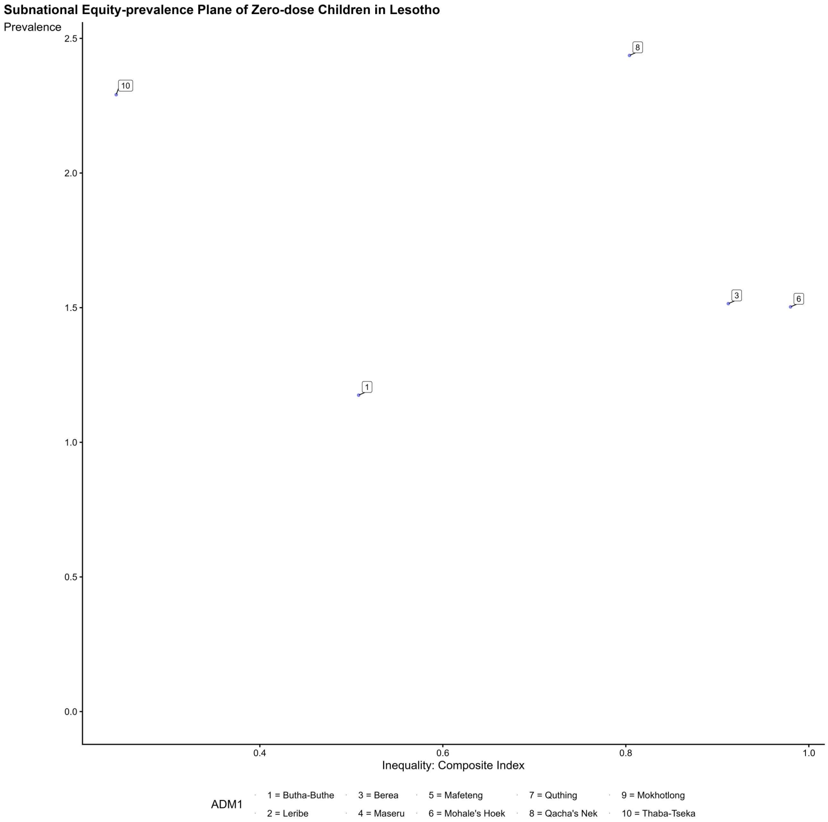

Subnational Equity-prevalence Plane of Zero-dose Children in Lesotho
Prevalence
2.5
2.0
1.5
1.0
0.5
0.0
0.4
0.6
0.8
1.0
Inequality: Composite Index
ADM1
1 = Butha-Buthe
2 = Leribe
3 = Berea
4 = Maseru
5 = Mafeteng
6 = Mohale's Hoek
7 = Quthing
8 = Qacha's Nek
9 = Mokhotlong
10 = Thaba-Tseka

##### Subnational Equity-prevalence Plane of Zero-dose Children in Madagascar

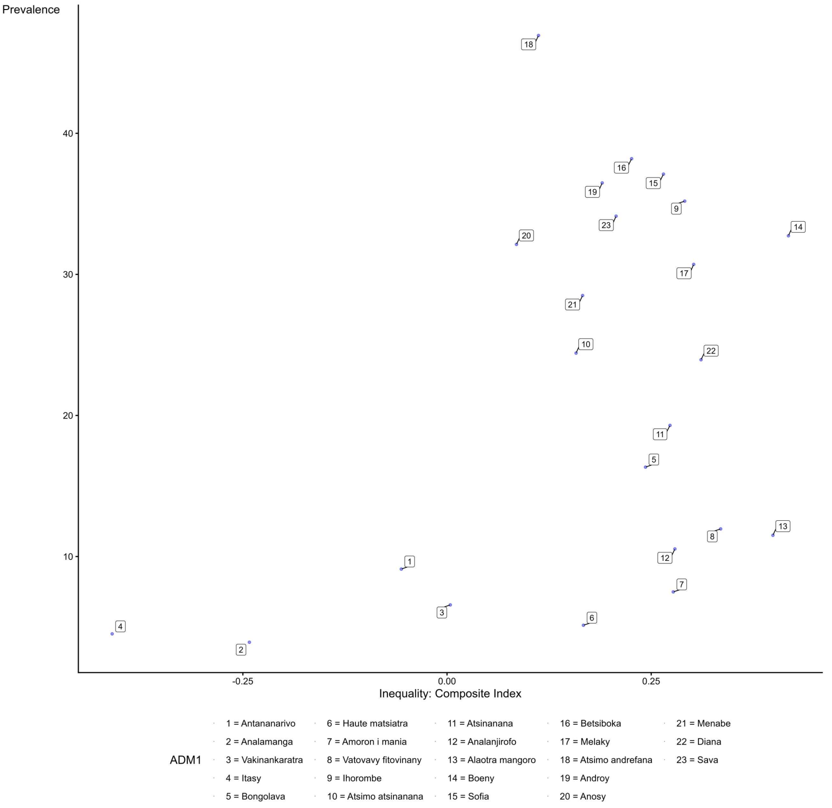

##### Subnational Equity-prevalence Plane of Zero-dose Children in North Macedonia

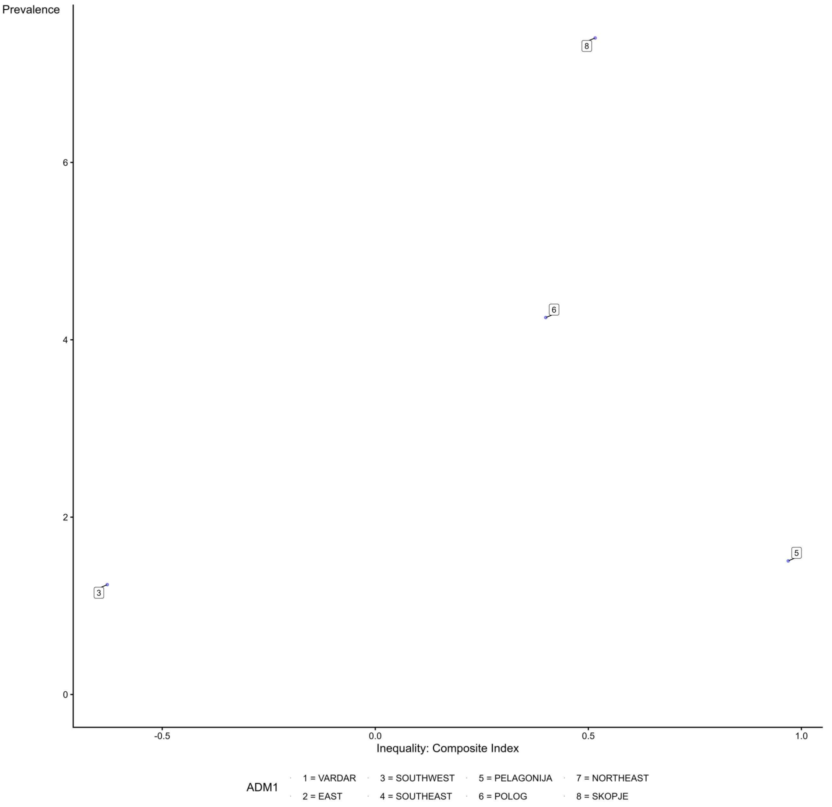

**Subnational Equity-prevalence Plane of Zero-dose Children in Mali**

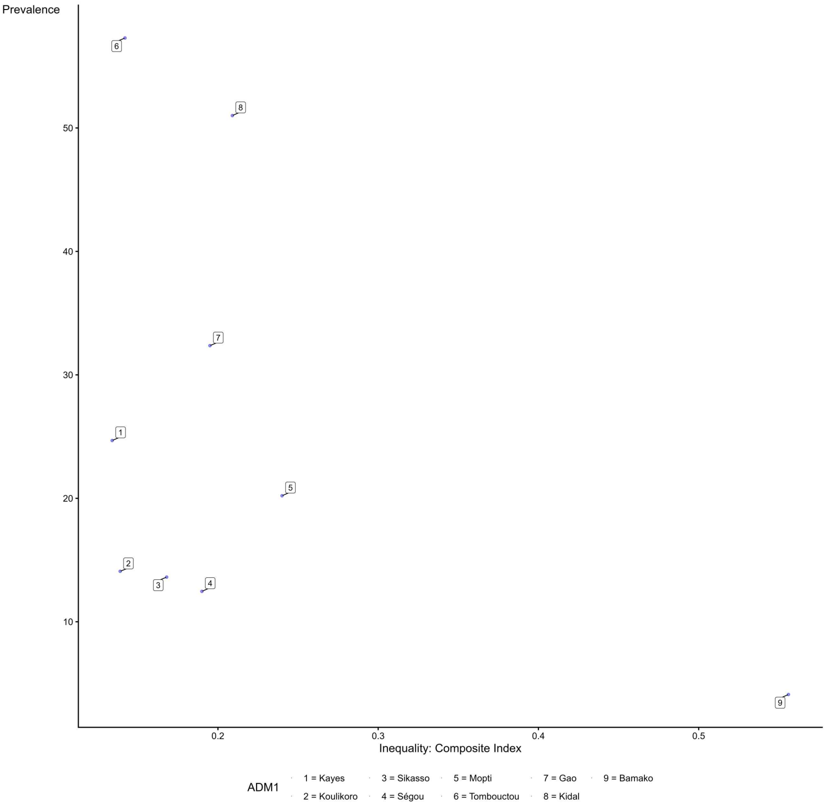

**Subnational Equity-prevalence Plane of Zero-dose Children in Myanmar**

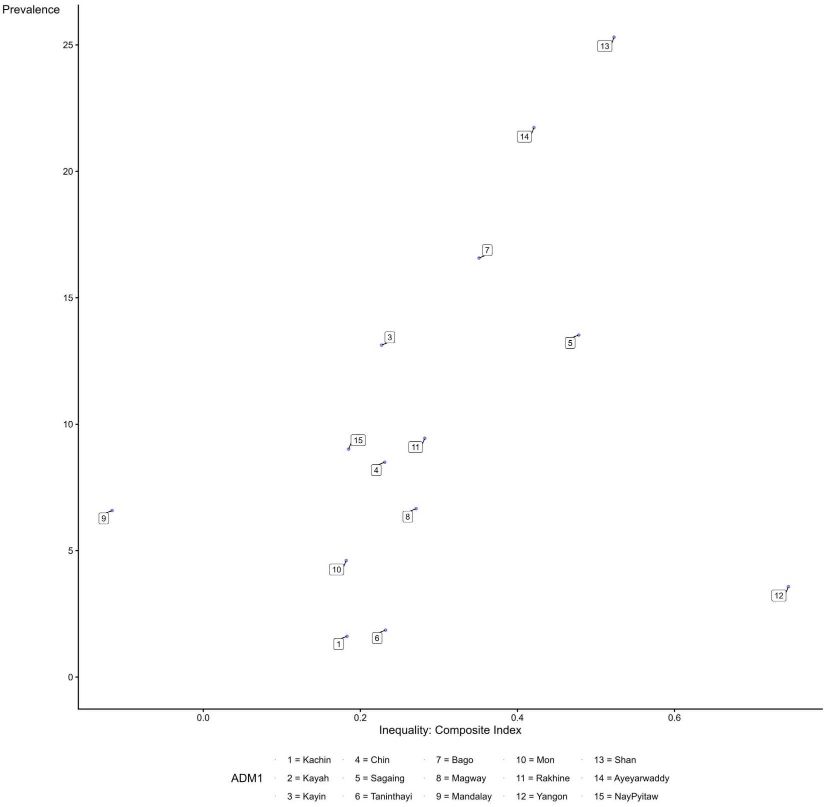

**Subnational Equity-prevalence Plane of Zero-dose Children in Mongolia**

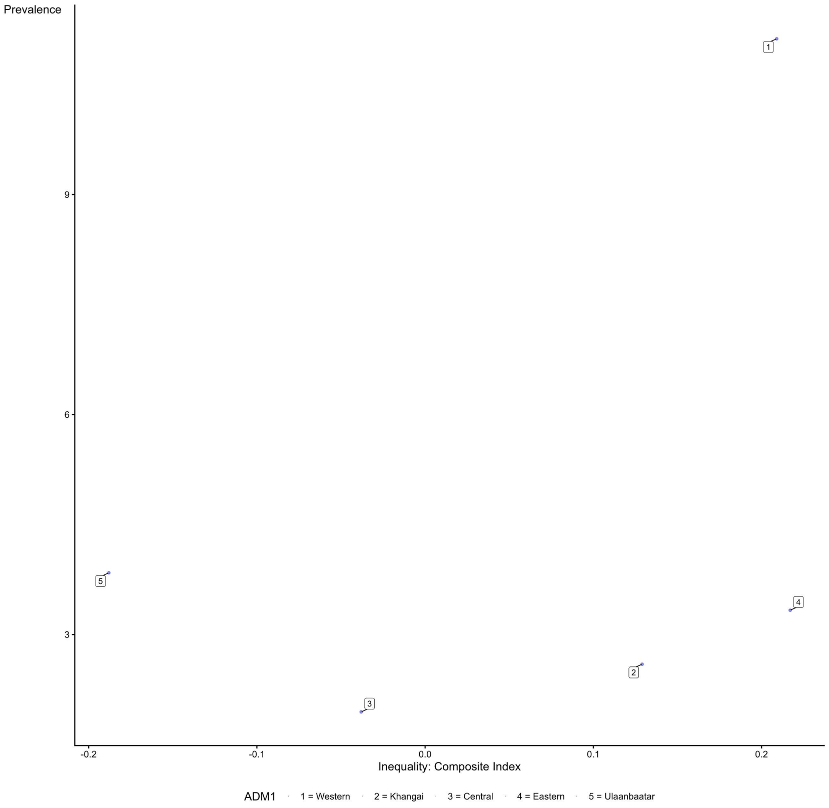

**Subnational Equity-prevalence Plane of Zero-dose Children in Mozambique**

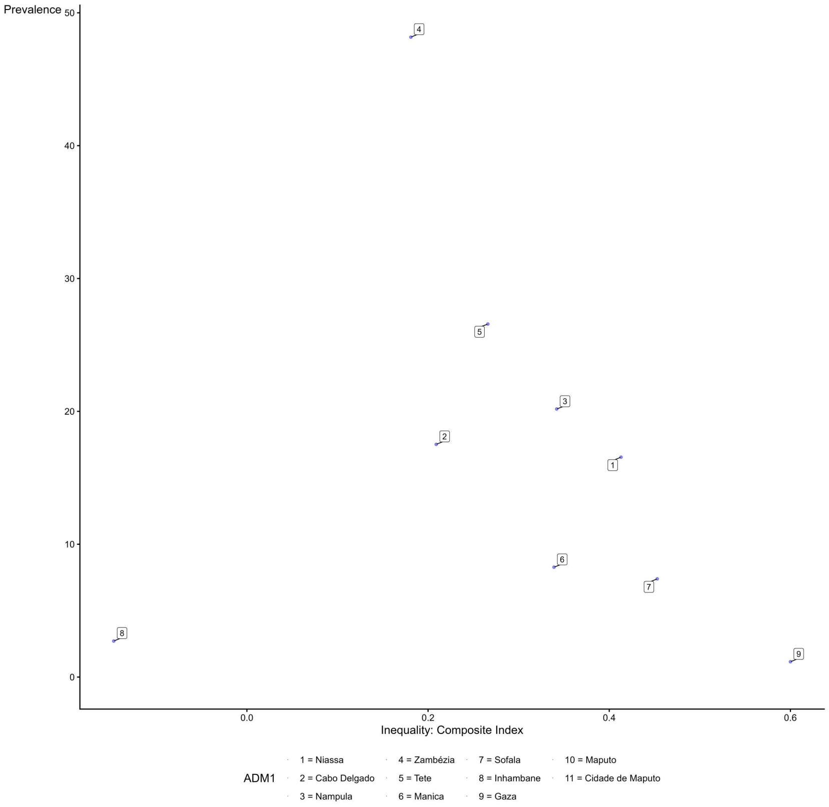

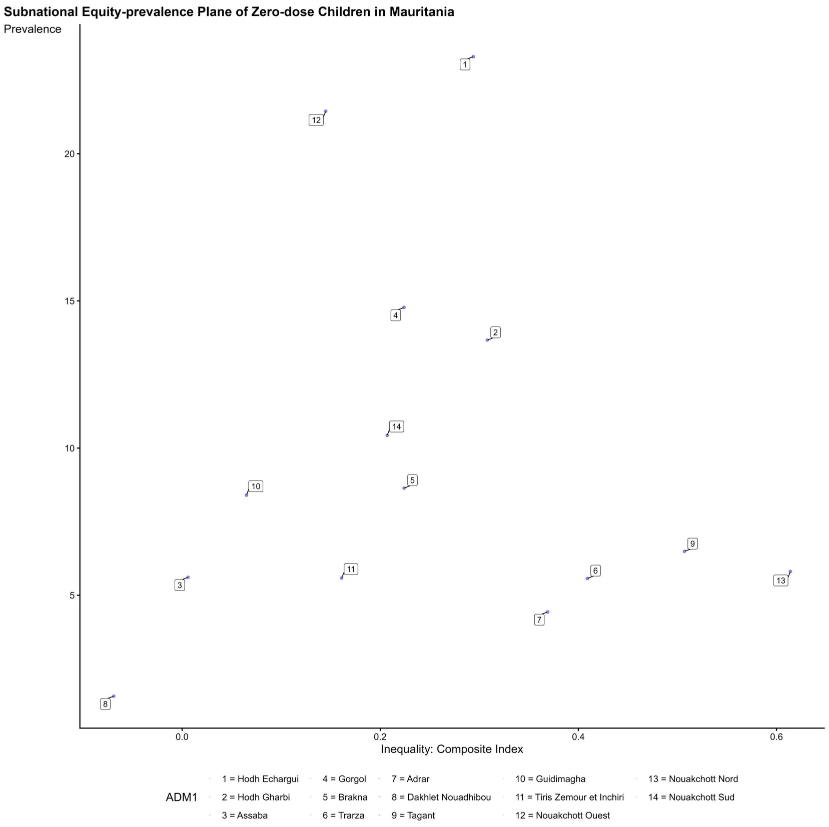
Subnational Equity-prevalence Plane of Zero-dose Children in Mauritania
Prevalence
20
15
10
5
0.0
0.2
0.4
0.6
Inequality: Composite Index
ADM1
1 = Hodh Echargui
2 = Hodh Gharbi
3 = Assaba
4 = Gorgol
5 = Brakna
6 = Trarza
7 = Adrar
8 = Dakhlet Nouadhibou
9 = Tagant
10 = Guidimagha
11 = Tiris Zemour et Inchiri
12 = Nouakchott Ouest
13 = Nouakchott Nord
14 = Nouakchott Sud

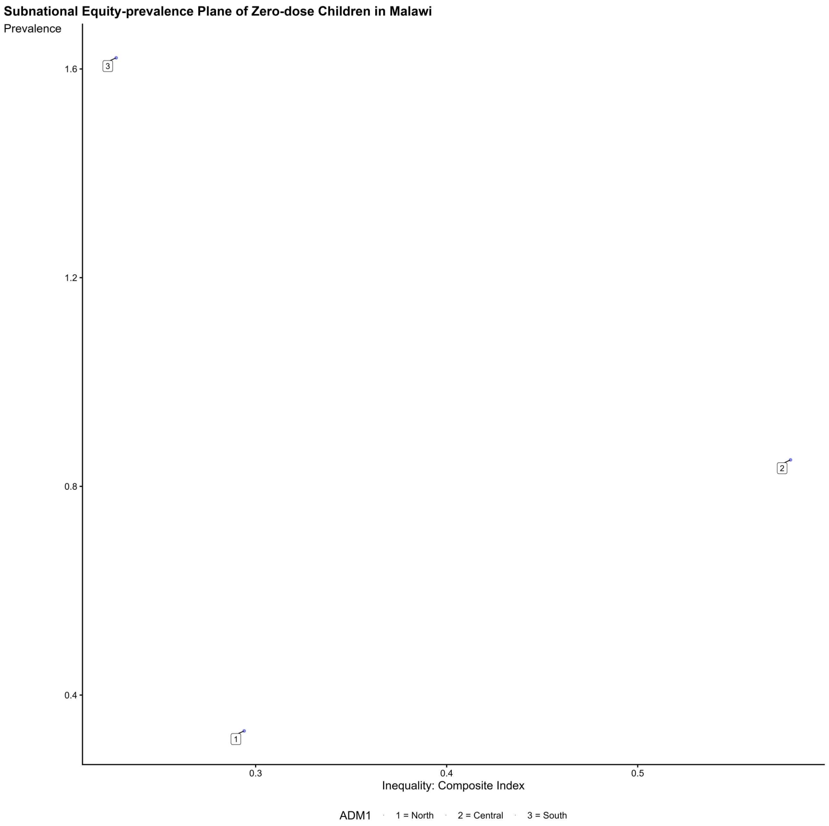
Subnational Equity-prevalence Plane of Zero-dose Children in Malawi
Prevalence
1.6
1.2
0.8
0.4
0.3
0.4
0.5
Inequality: Composite Index
ADM1
1 = North
2 = Central
3 = South

**Subnational Equity-prevalence Plane of Zero-dose Children in Nigeria**

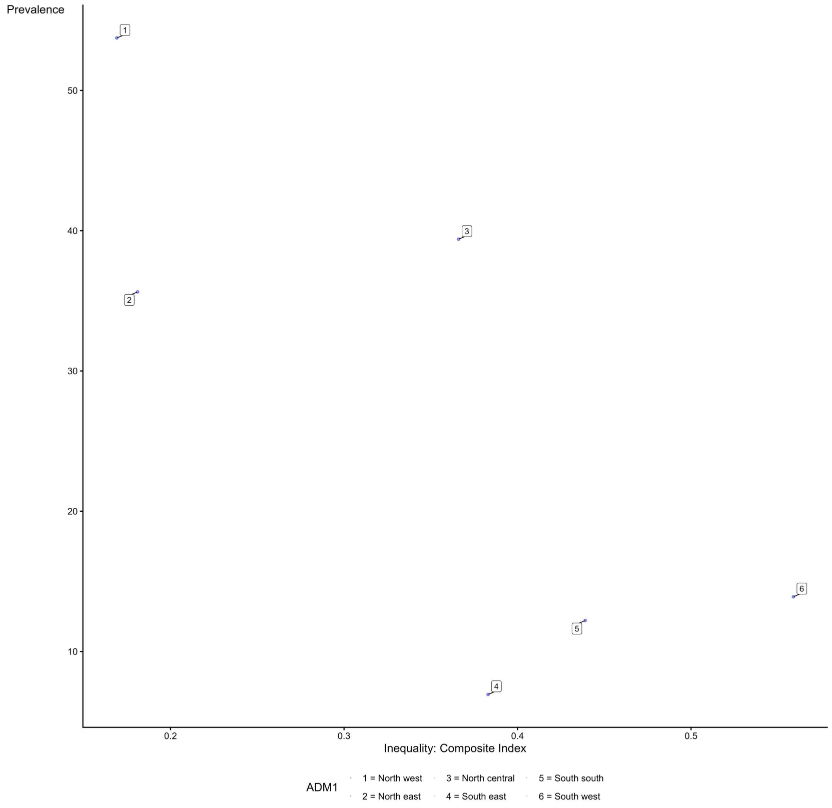

**Subnational Equity-prevalence Plane of Zero-dose Children in Nepal**

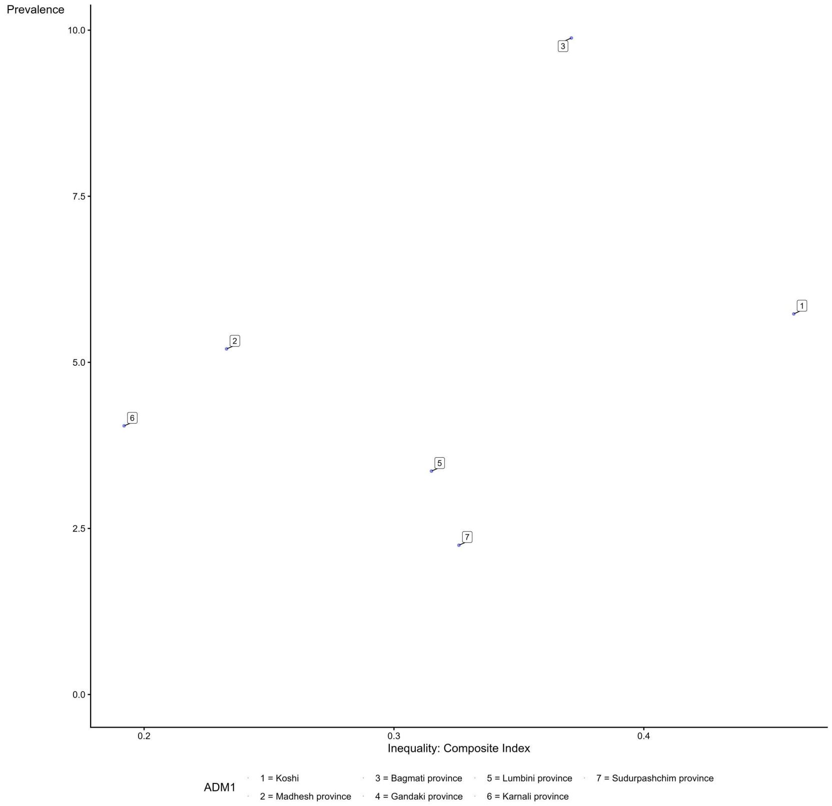

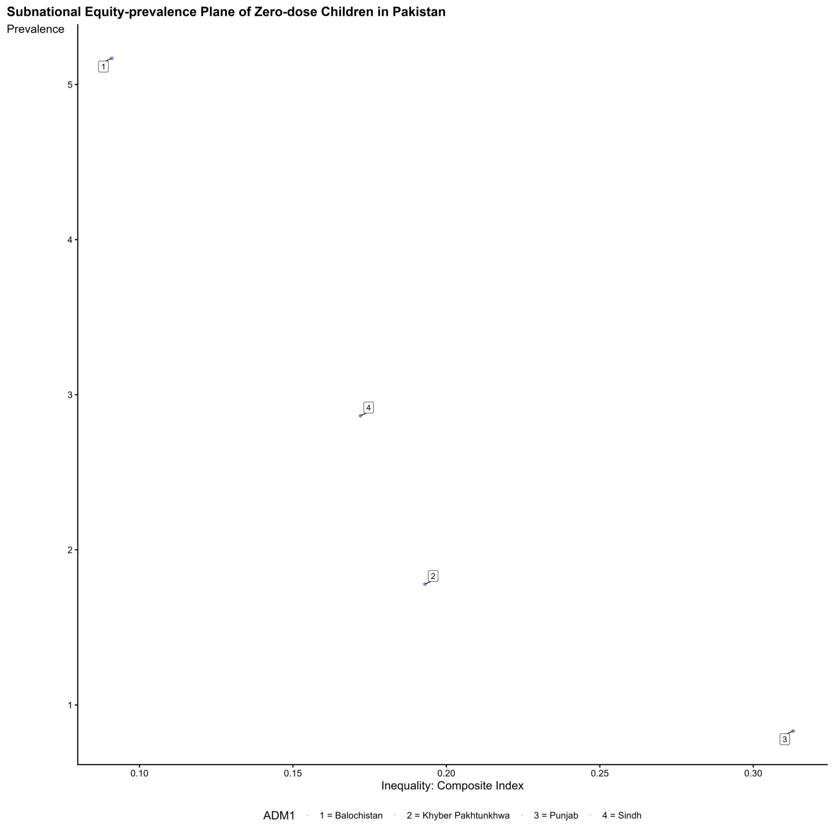
Subnational Equity-prevalence Plane of Zero-dose Children in Pakistan
Prevalence
Inequality: Composite Index
ADM1
1 = Balochistan
2 = Khyber Pakhtunkhwa
3 = Punjab
4 = Sindh

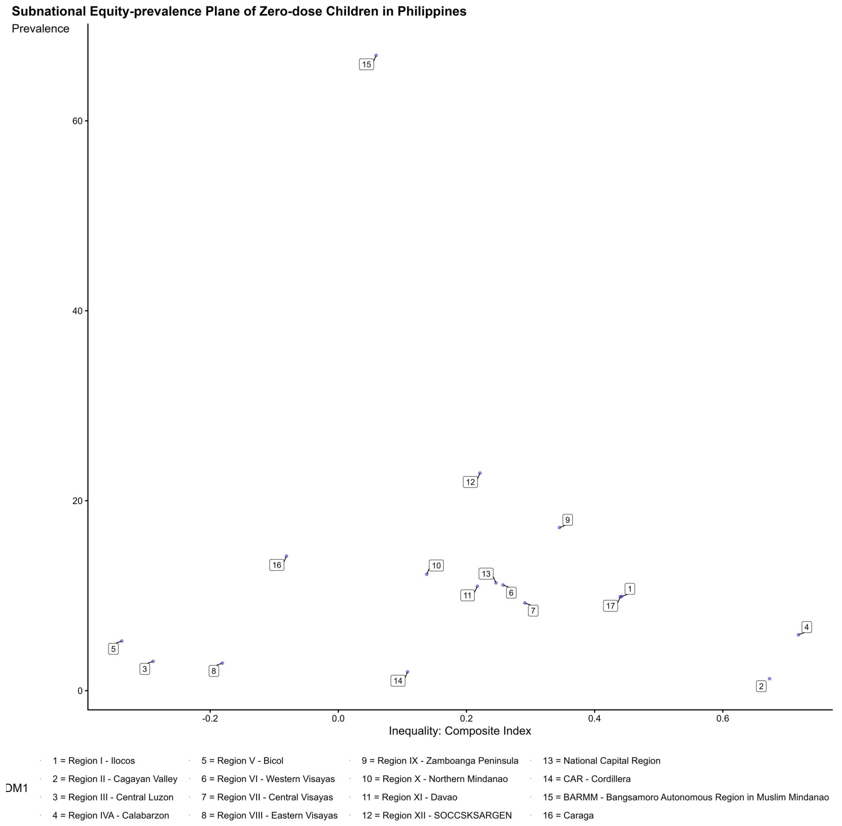
Subnational Equity-prevalence Plane of Zero-dose Children in Philippines
Prevalence
Inequality: Composite Index
ADM1
1 = Region I - Ilocos
2 = Region II - Cagayan Valley
3 = Region III - Central Luzon
4 = Region IVA - Calabarzon
5 = Region V - Bicol
6 = Region VI - Western Visayas
7 = Region VII - Central Visayas
8 = Region VIII - Eastern Visayas
9 = Region IX - Zamboanga Peninsula
10 = Region X - Northern Mindanao
11 = Region XI - Davao
12 = Region XII - SOCCSKSARGEN
13 = National Capital Region
14 = CAR - Cordillera
15 = BARMM - Bangsamoro Autonomous Region in Muslim Mindanao
16 = Caraga

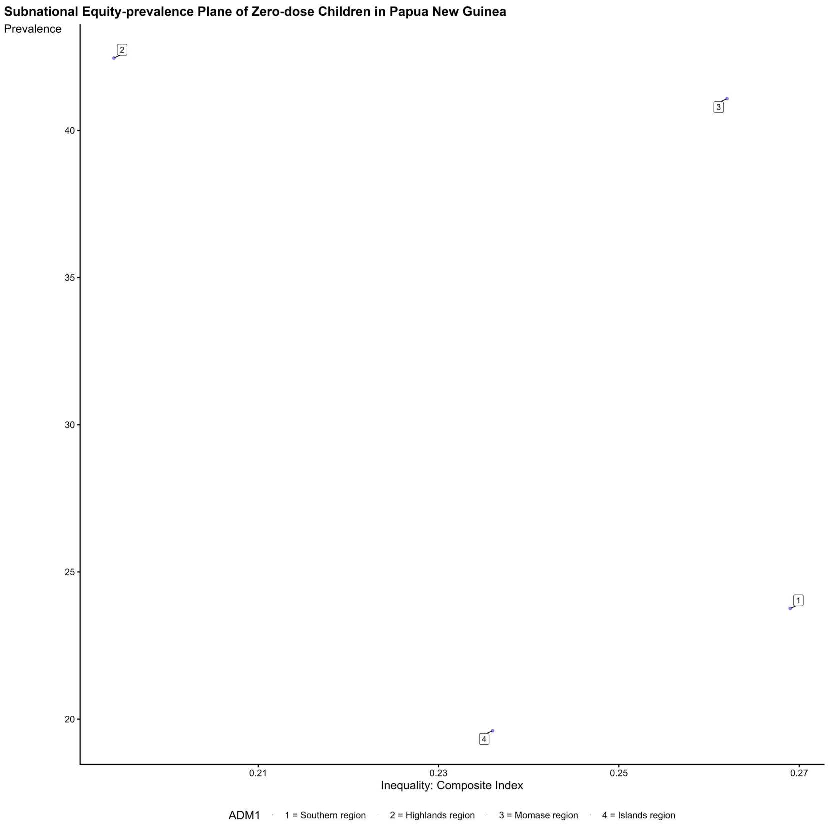
Subnational Equity-prevalence Plane of Zero-dose Children in Papua New Guinea
Prevalence
Inequality: Composite Index
ADM1
1 = Southern region
2 = Highlands region
3 = Momase region
4 = Islands region

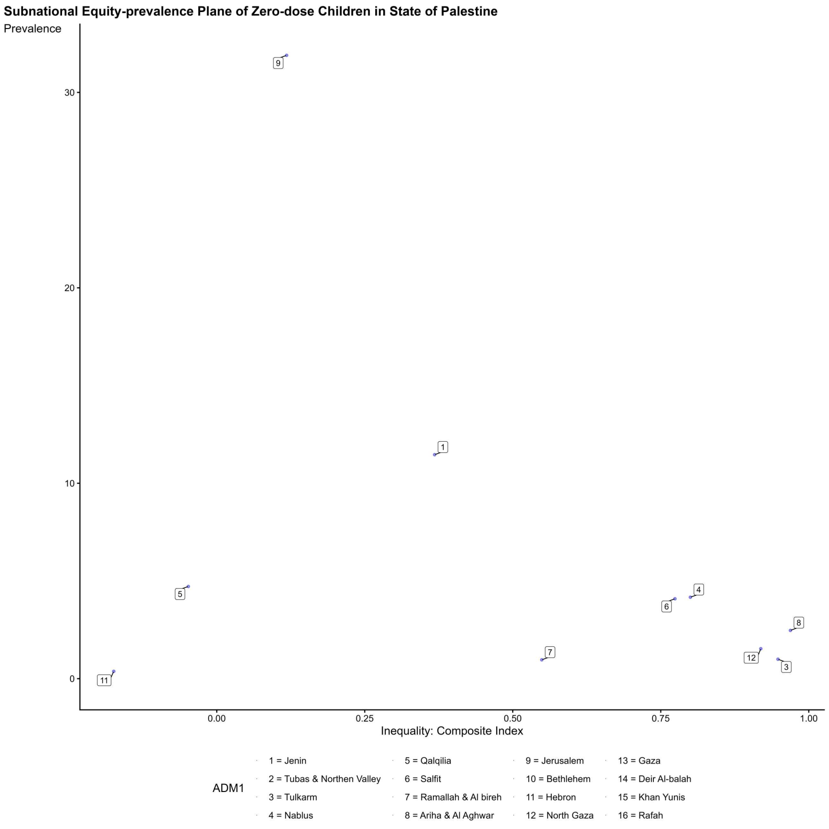
Subnational Equity-prevalence Plane of Zero-dose Children in State of Palestine
Prevalence
Inequality: Composite Index
ADM1
1 = Jenin
2 = Tubas & Northen Valley
3 = Tulkarm
4 = Nablus
5 = Qalqilia
6 = Salfit
7 = Ramallah & Al bireh
8 = Ariha & Al Aghwar
9 = Jerusalem
10 = Bethlehem
11 = Hebron
12 = North Gaza
13 = Gaza
14 = Deir Al-balah
15 = Khan Yunis
16 = Rafah

Subnational Equity-prevalence Plane of Zero-dose Children in Rwanda

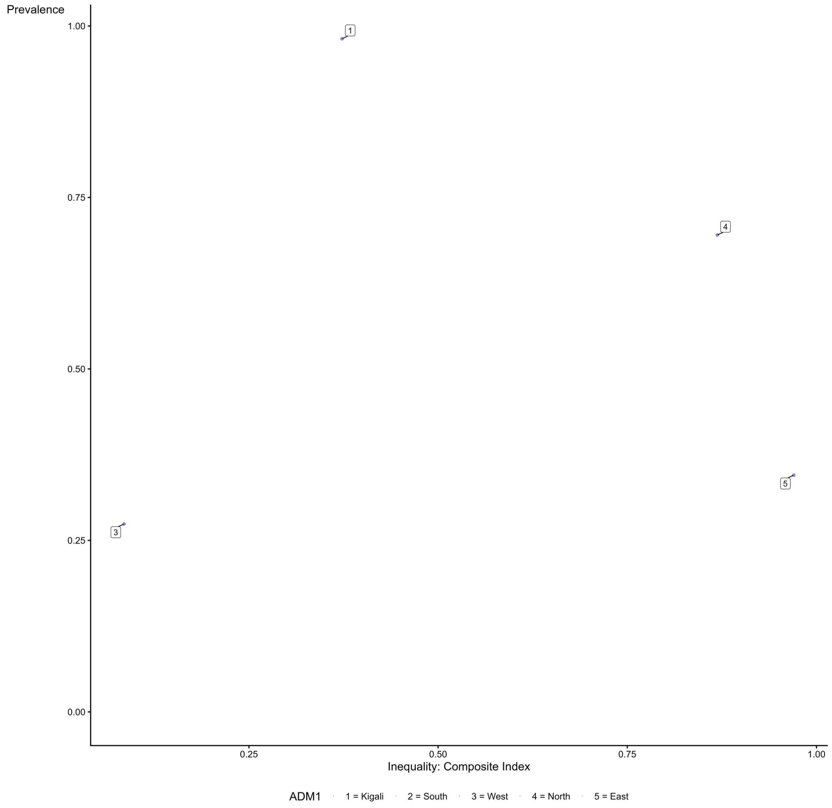

Subnational Equity-prevalence Plane of Zero-dose Children in Senegal

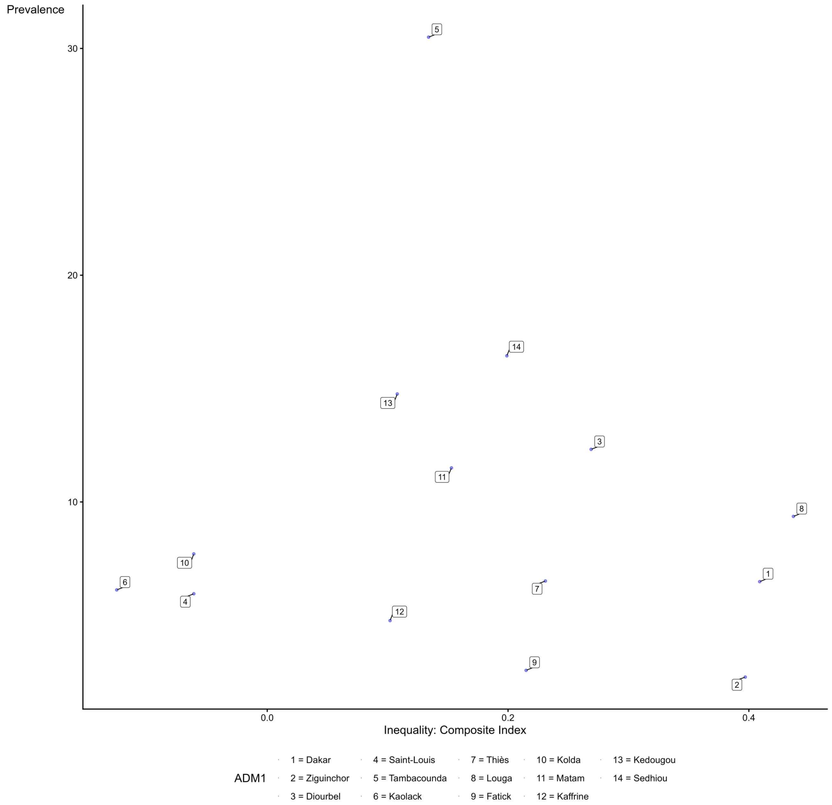

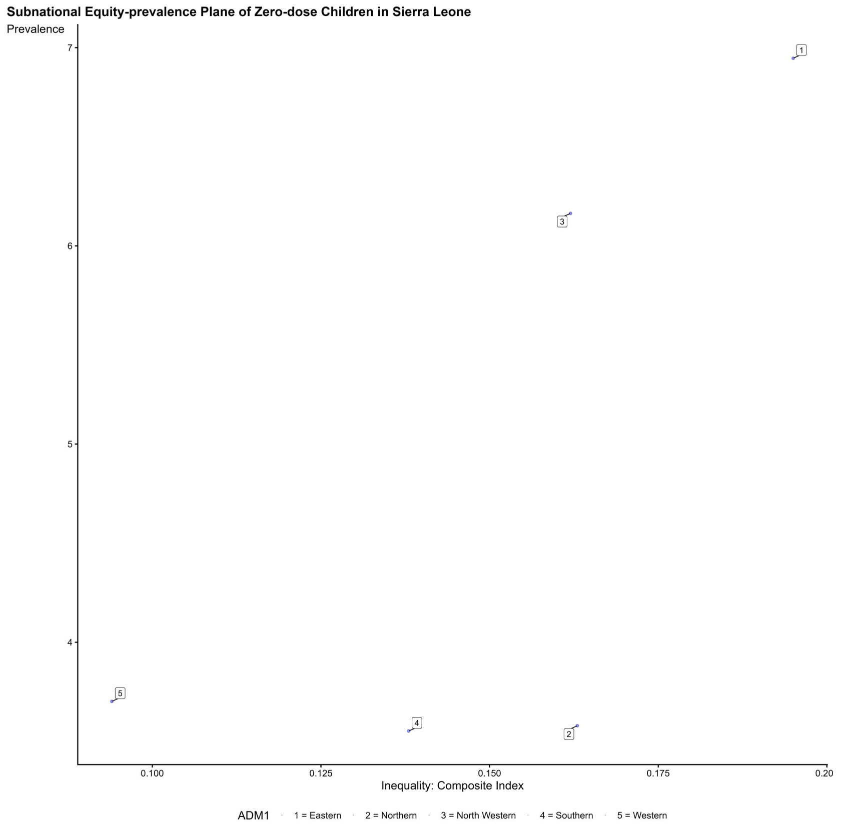
Subnational Equity-prevalence Plane of Zero-dose Children in Sierra Leone
Prevalence
7
6
5
4
0.100
0.125
0.150
0.175
0.20
Inequality: Composite Index
1
2
3
4
5
ADM1
1 = Eastern
2 = Northern
3 = North Western
4 = Southern
5 = Western

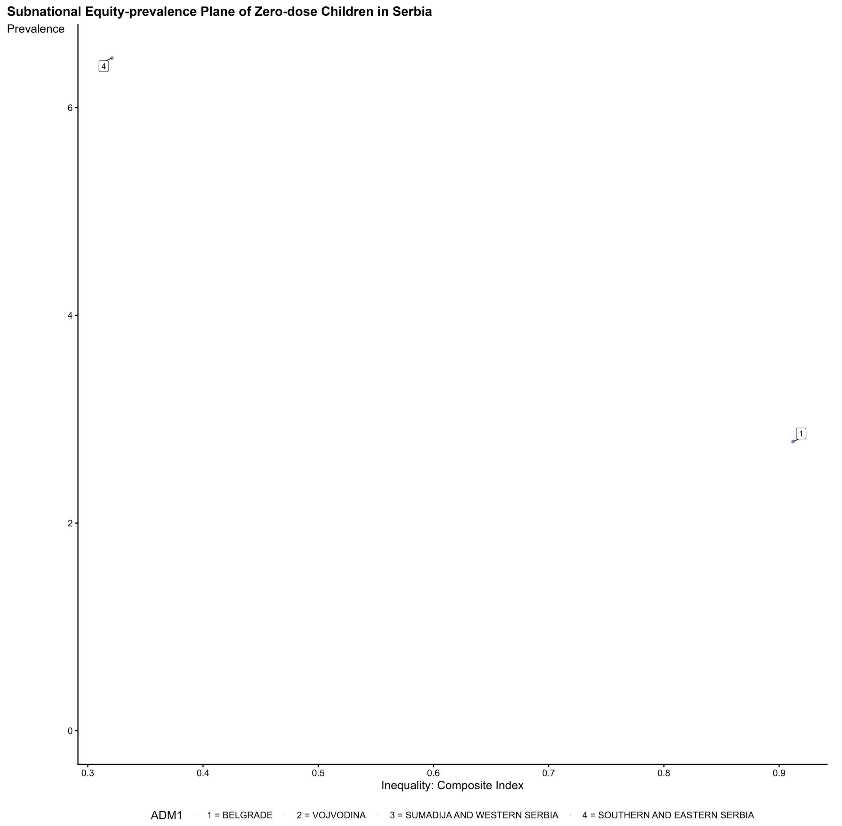
Subnational Equity-prevalence Plane of Zero-dose Children in Serbia
Prevalence
6
4
2
0
0.3
0.4
0.5
0.6
0.7
0.8
0.9
Inequality: Composite Index
4
1
ADM1
1 = BELGRADE
2 = VOJVODINA
3 = SUMADIJA AND WESTERN SERBIA
4 = SOUTHERN AND EASTERN SERBIA

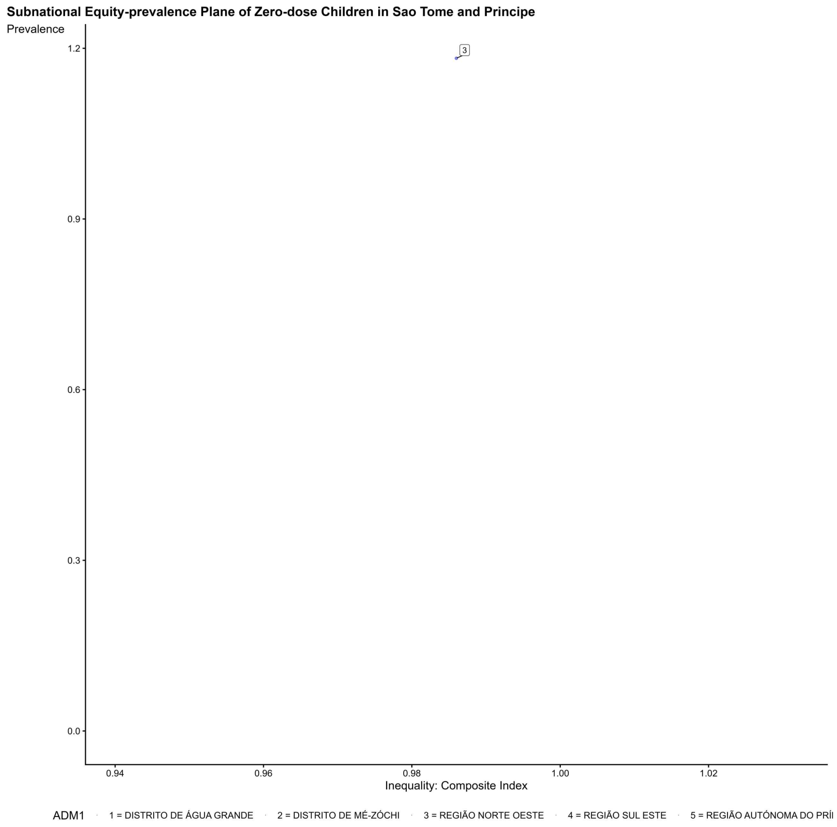
Subnational Equity-prevalence Plane of Zero-dose Children in Sao Tome and Principe
Prevalence
1.2
0.9
0.6
0.3
0.0
3
0.94
0.96
0.98
1.00
1.02
Inequality: Composite Index
ADM1
1 = DISTRITO DE ÁGUA GRANDE
2 = DISTRITO DE MÉ-ZÓCHI
3 = REGIÃO NORTE OESTE
4 = REGIÃO SUL ESTE
5 = REGIÃO AUTÓNOMA DO PRÍ

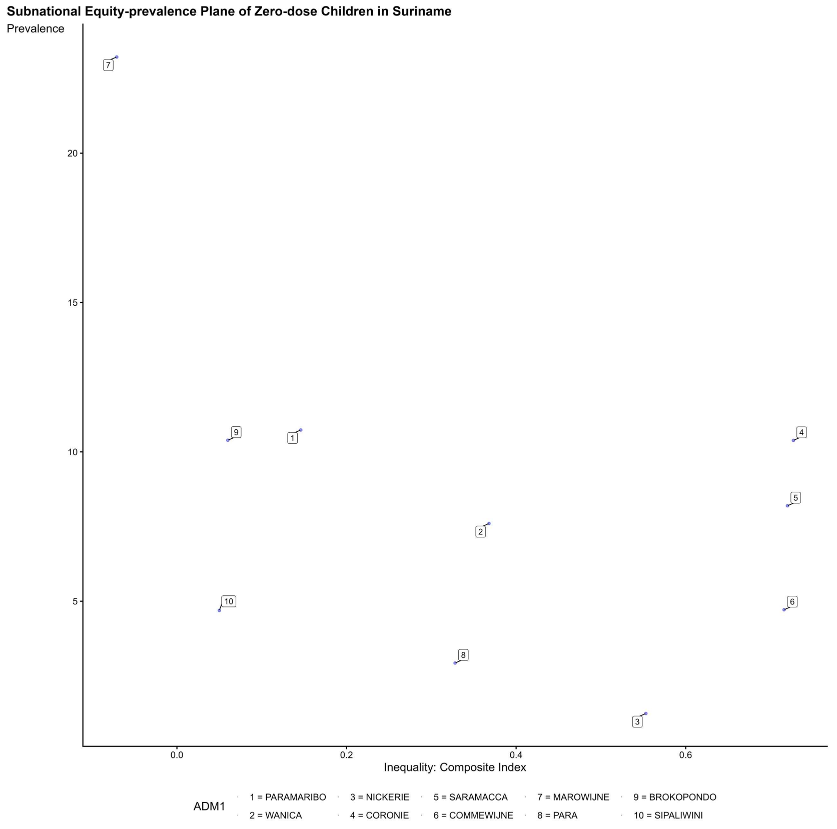
Subnational Equity-prevalence Plane of Zero-dose Children in Suriname
Prevalence
20
15
10
5
7
9
1
4
5
2
10
6
8
3
0.0
0.2
0.4
0.6
Inequality: Composite Index
ADM1
1 = PARAMARIBO
3 = NICKERIE
5 = SARAMACCA
7 = MAROWIJNE
9 = BROKOPONDO
2 = WANICA
4 = CORONIE
6 = COMMEWIJNE
8 = PARA
10 = SIPALIWINI

**Subnational Equity-prevalence Plane of Zero-dose Children in Eswatini**

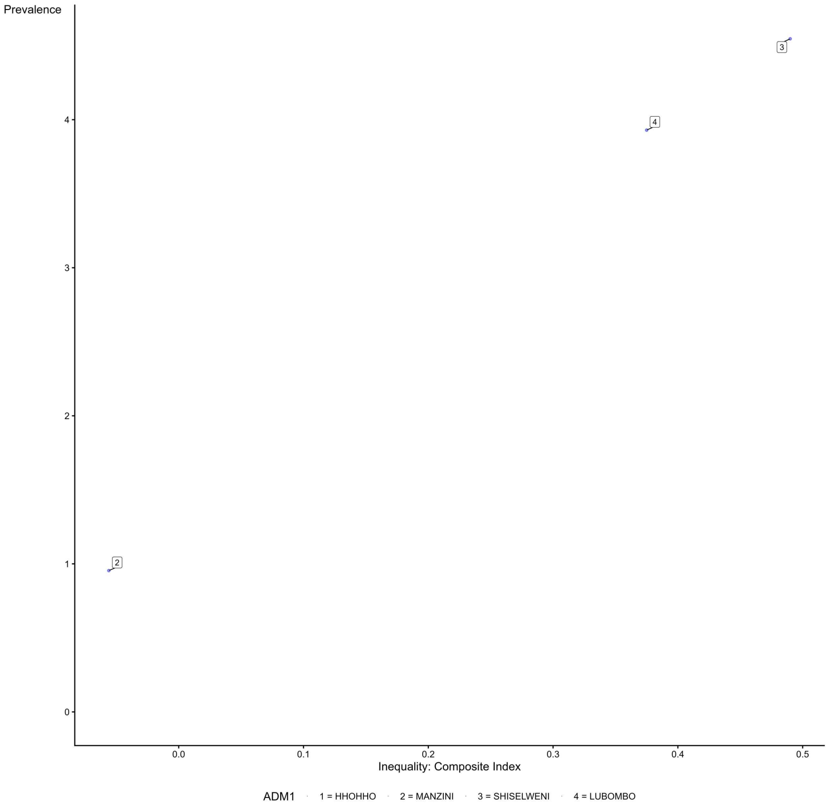

**Subnational Equity-prevalence Plane of Zero-dose Children in Chad**

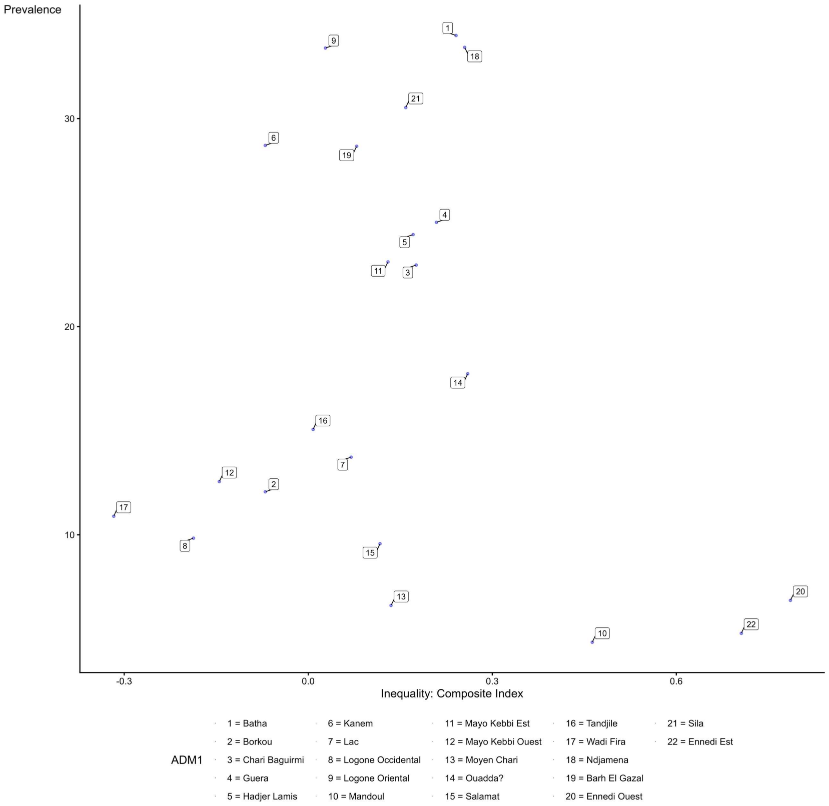

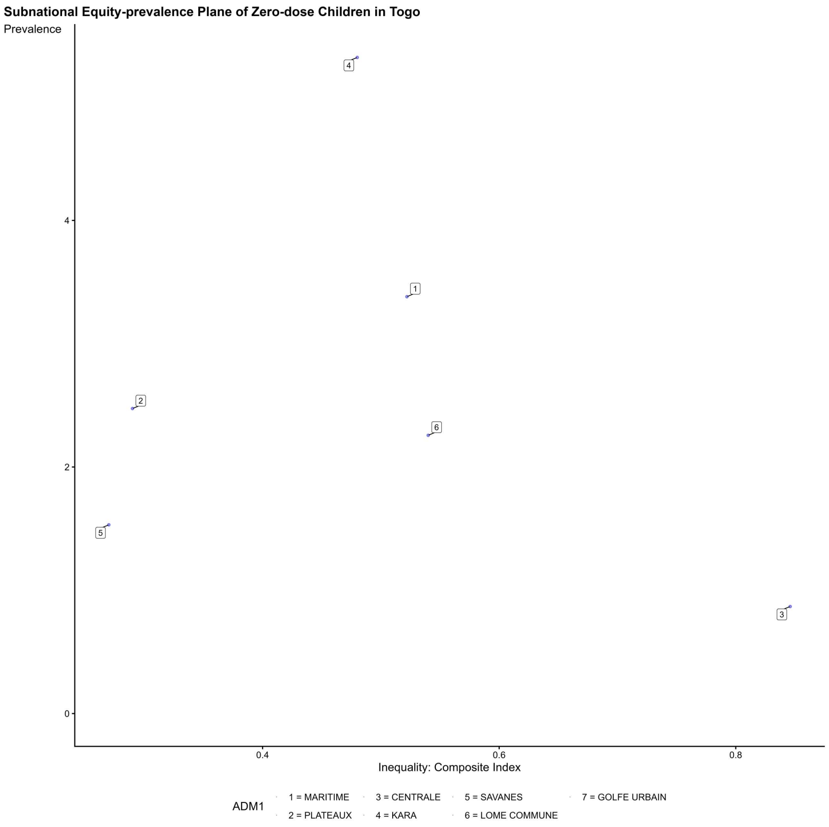
Subnational Equity-prevalence Plane of Zero-dose Children in Togo
Prevalence
0
2
4
0.4
0.6
0.8
Inequality: Composite Index
1
2
3
4
5
6
ADM1
1 = MARITIME
2 = PLATEAUX
3 = CENTRALE
4 = KARA
5 = SAVANES
6 = LOME COMMUNE
7 = GOLFE URBAIN

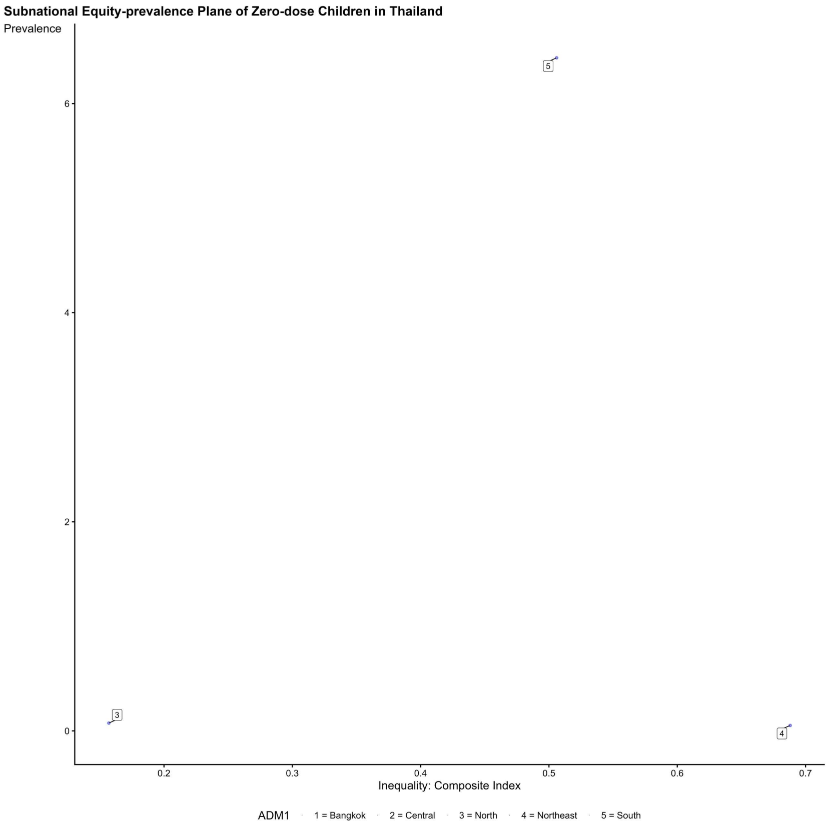
Subnational Equity-prevalence Plane of Zero-dose Children in Thailand
Prevalence
0
2
4
6
0.2
0.3
0.4
0.5
0.6
0.7
Inequality: Composite Index
3
4
5
ADM1
1 = Bangkok
2 = Central
3 = North
4 = Northeast
5 = South

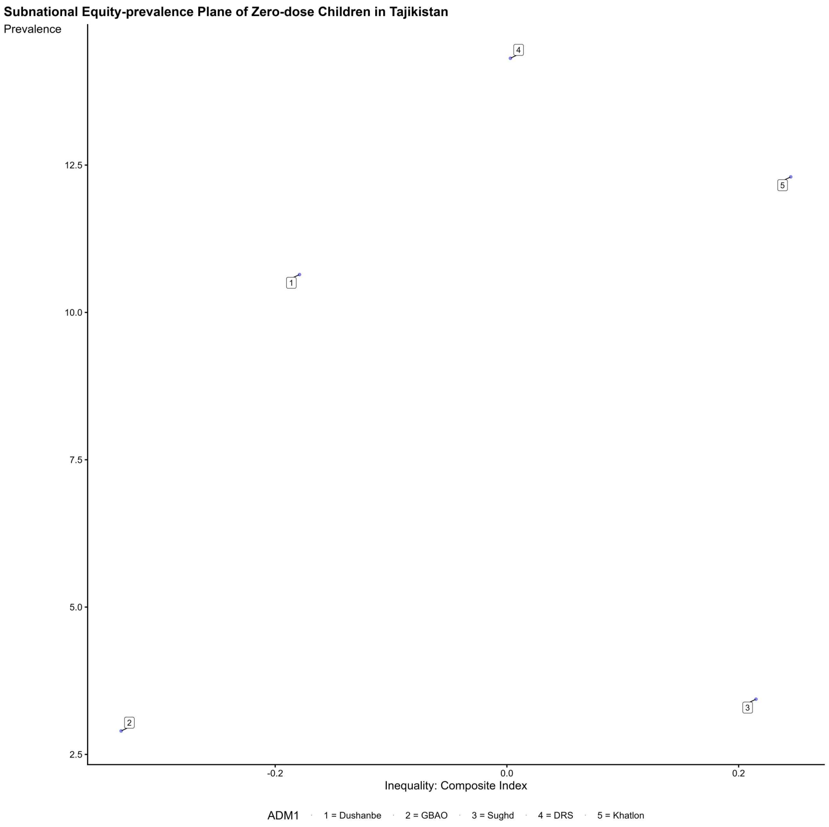
Subnational Equity-prevalence Plane of Zero-dose Children in Tajikistan
Prevalence
Inequality: Composite Index
ADM1
1 = Dushanbe
2 = GBAO
3 = Sughd
4 = DRS
5 = Khatlon

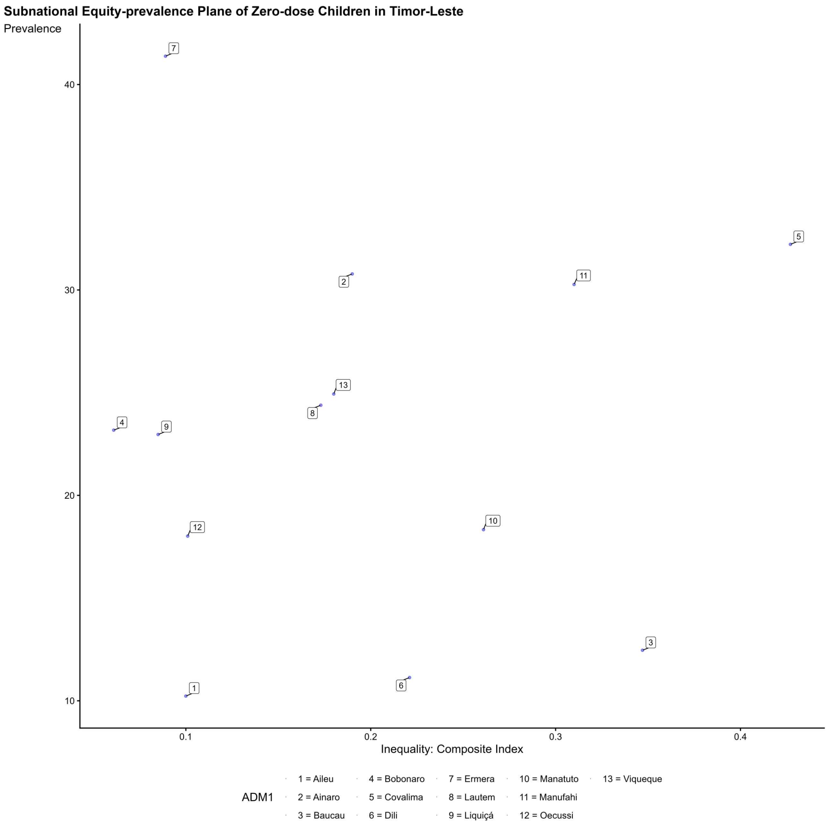
Subnational Equity-prevalence Plane of Zero-dose Children in Timor-Leste
Prevalence
Inequality: Composite Index
ADM1
1 = Aileu
2 = Ainaro
3 = Baucau
4 = Bobonaro
5 = Covalima
6 = Dili
7 = Ermera
8 = Lautem
9 = Liquiçá
10 = Manatuto
11 = Manufahi
12 = Oecussi
13 = Viqueque

**Subnational Equity-prevalence Plane of Zero-dose Children in Tonga**

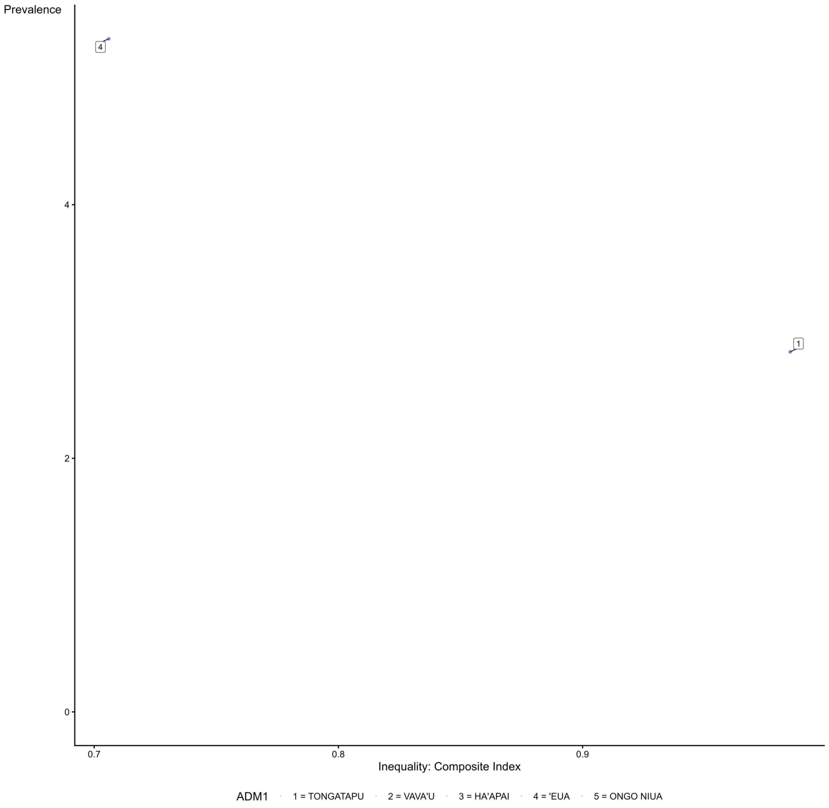

**Subnational Equity-prevalence Plane of Zero-dose Children in Trinidad and Tobago**

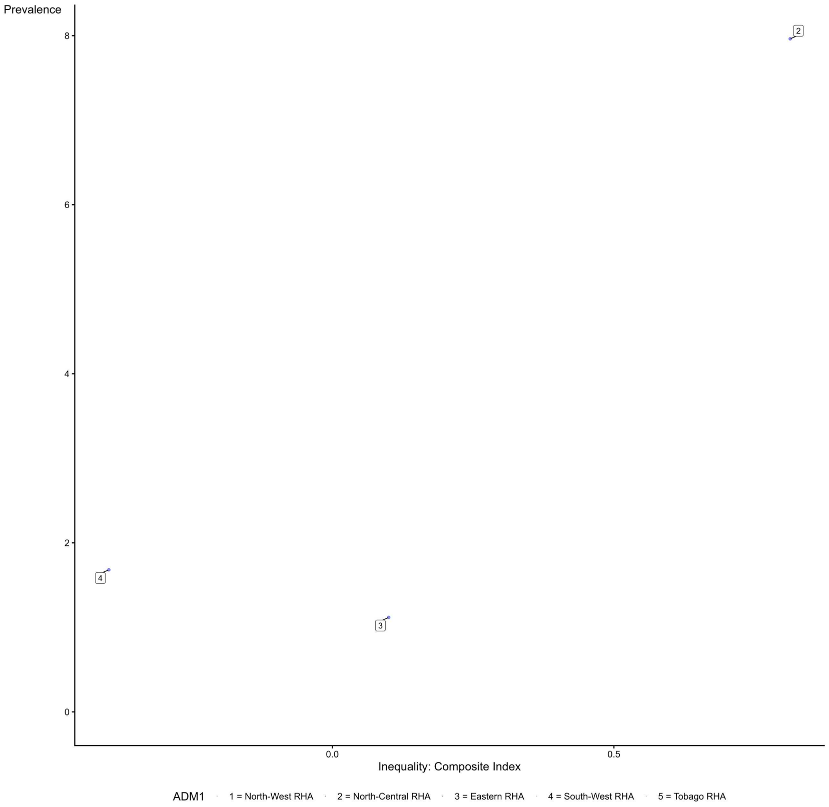

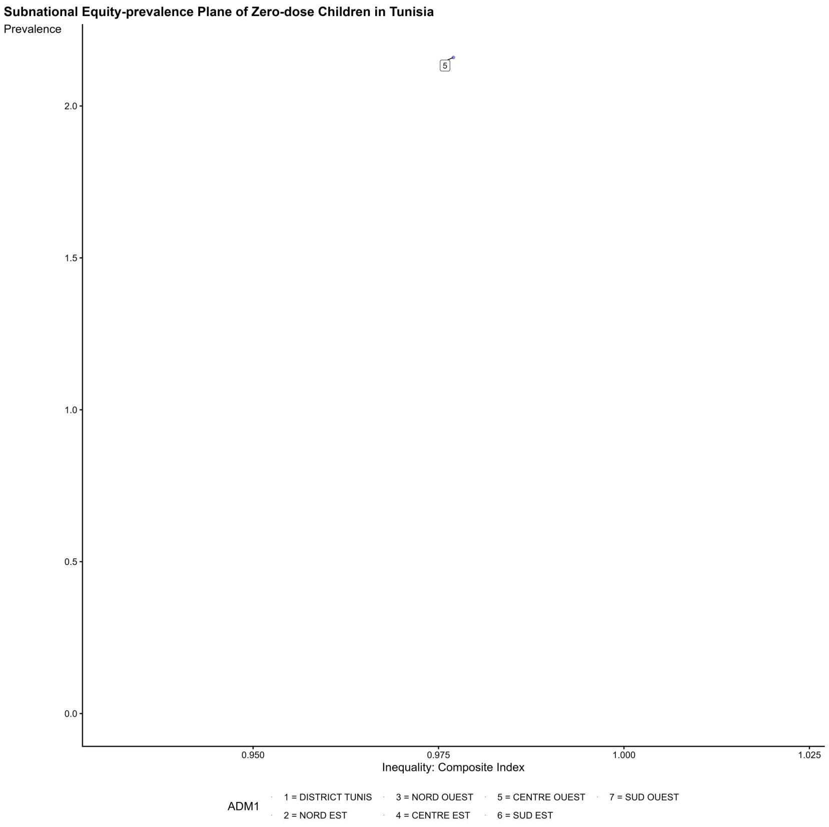
Subnational Equity-prevalence Plane of Zero-dose Children in Tunisia
Prevalence
2.0
1.5
1.0
0.5
0.0
5
0.950
0.975
1.000
1.025
Inequality: Composite Index
ADM1
1 = DISTRICT TUNIS
2 = NORD EST
3 = NORD OUEST
4 = CENTRE EST
5 = CENTRE OUEST
6 = SUD EST
7 = SUD OUEST

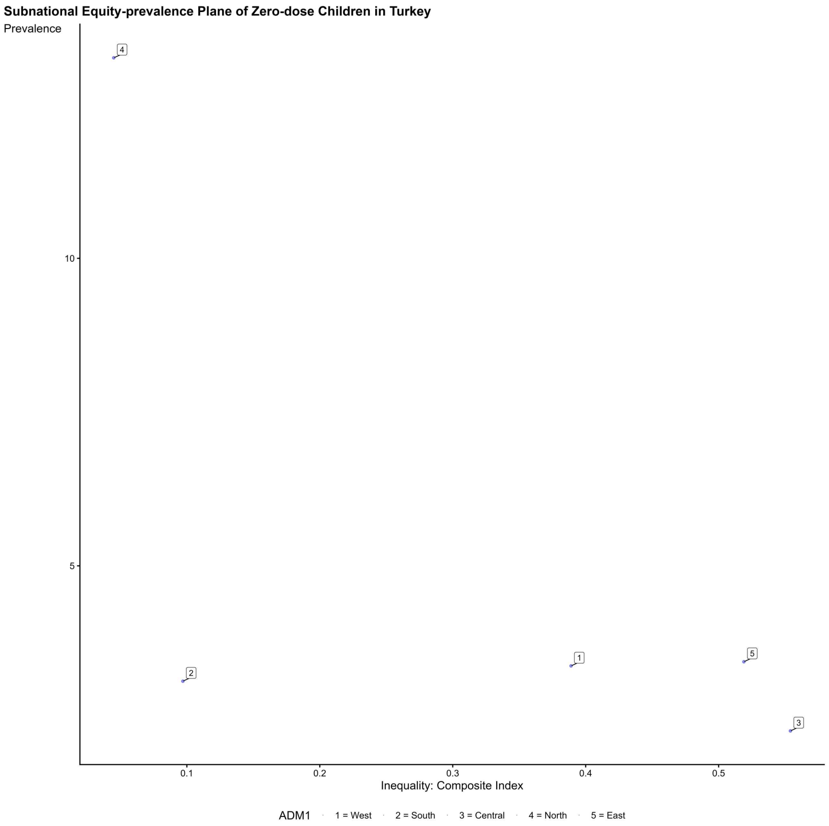
Subnational Equity-prevalence Plane of Zero-dose Children in Turkey
Prevalence
10
5
4
2
1
5
3
0.1
0.2
0.3
0.4
0.5
Inequality: Composite Index
ADM1
1 = West
2 = South
3 = Central
4 = North
5 = East

**Subnational Equity-prevalence Plane of Zero-dose Children in Tanzania**

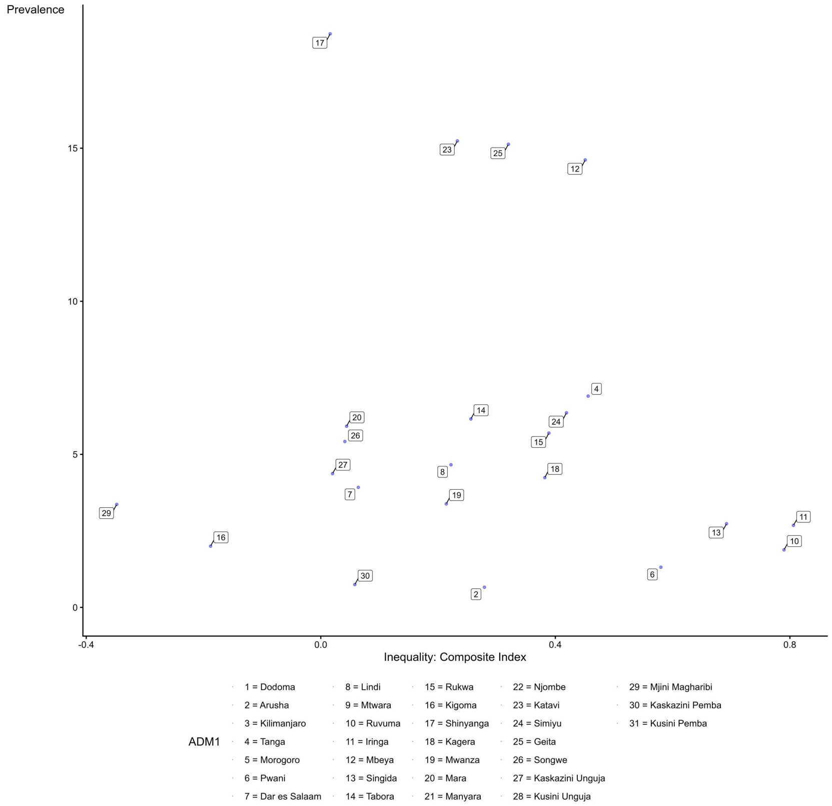

**Subnational Equity-prevalence Plane of Zero-dose Children in Uganda**

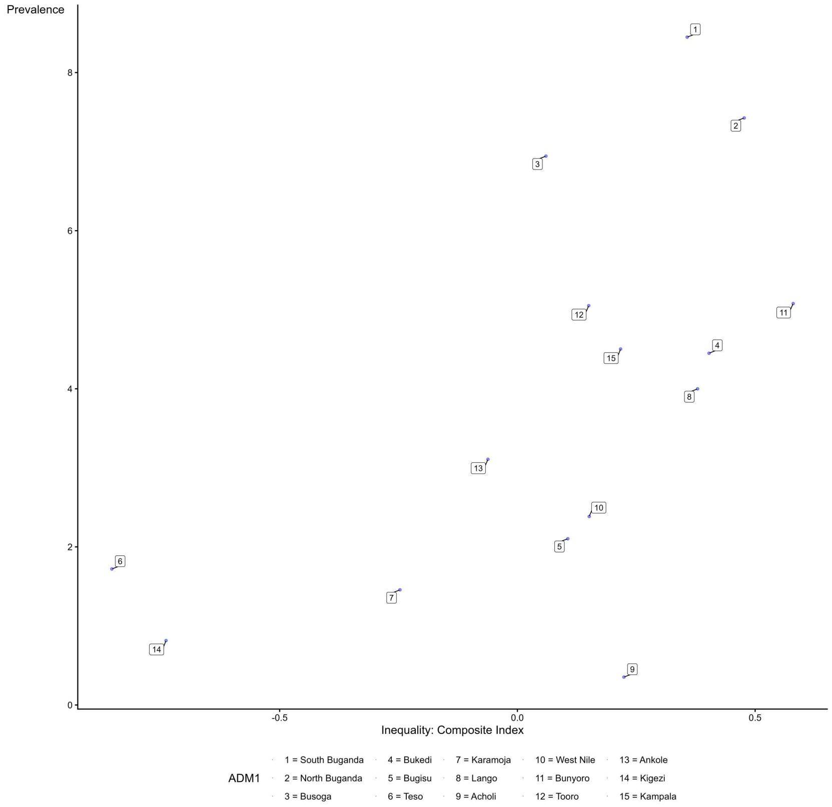

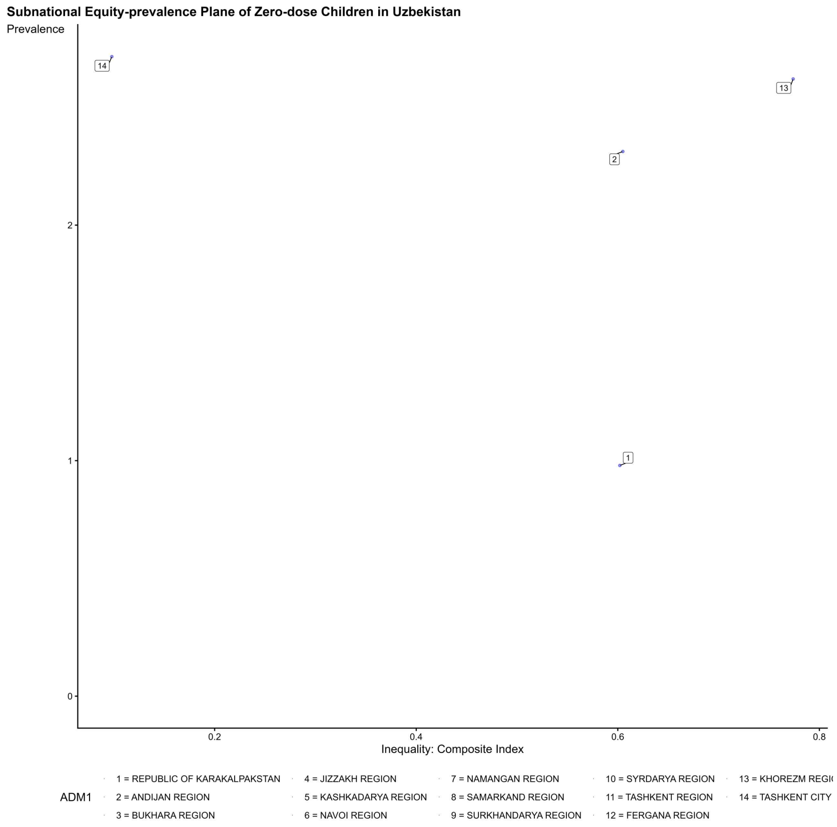
Subnational Equity-prevalence Plane of Zero-dose Children in Uzbekistan
Prevalence
Inequality: Composite Index
14
13
2
1
ADM1
1 = REPUBLIC OF KARAKALPAKSTAN
2 = ANDIJAN REGION
3 = BUKHARA REGION
4 = JIZZAKH REGION
5 = KASHKADARYA REGION
6 = NAVOI REGION
7 = NAMANGAN REGION
8 = SAMARKAND REGION
9 = SURKHANDARYA REGION
10 = SYRDARYA REGION
11 = TASHKENT REGION
12 = FERGANA REGION
14 = TASHKENT CITY

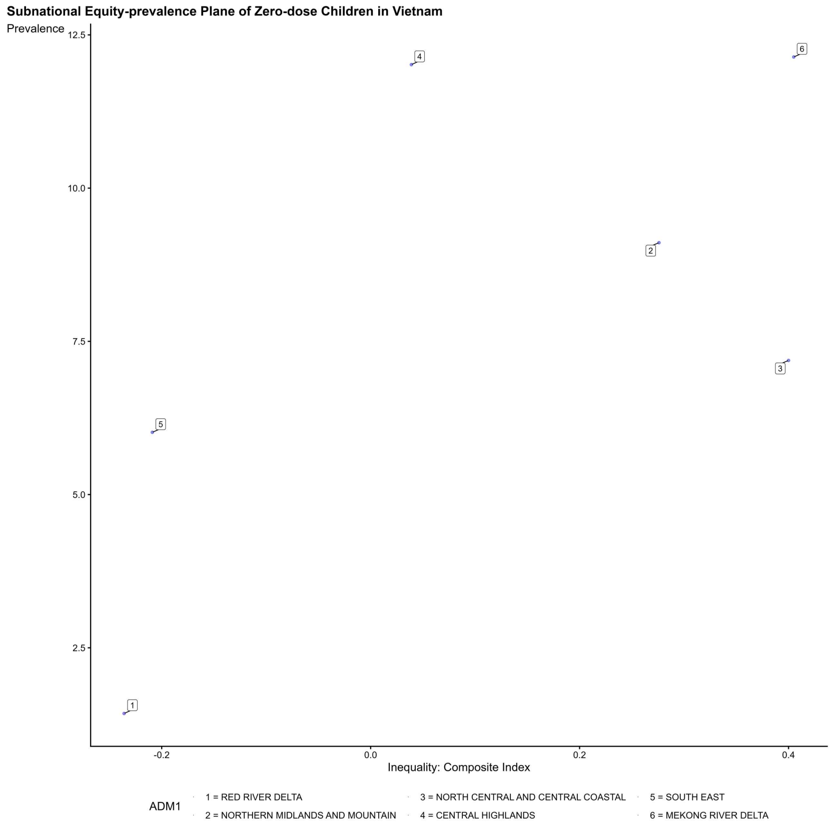
Subnational Equity-prevalence Plane of Zero-dose Children in Vietnam
Prevalence
Inequality: Composite Index
4
6
2
3
5
1
ADM1
1 = RED RIVER DELTA
2 = NORTHERN MIDLANDS AND MOUNTAIN
3 = NORTH CENTRAL AND CENTRAL COASTAL
4 = CENTRAL HIGHLANDS
5 = SOUTH EAST
6 = MEKONG RIVER DELTA

**Subnational Equity-prevalence Plane of Zero-dose Children in Vanuatu**

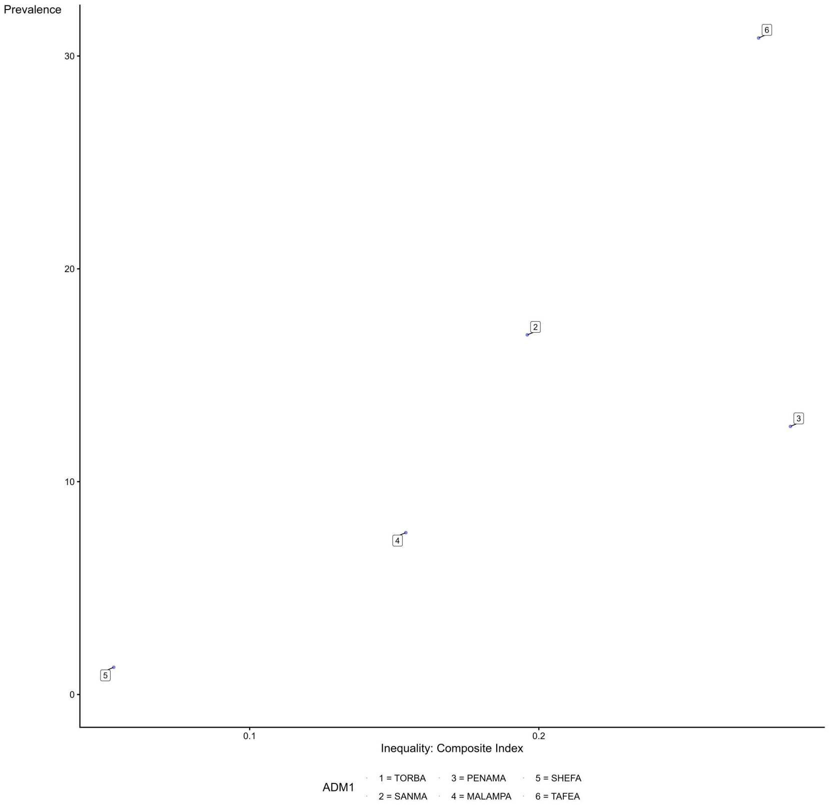

**Subnational Equity-prevalence Plane of Zero-dose Children in Yemen**

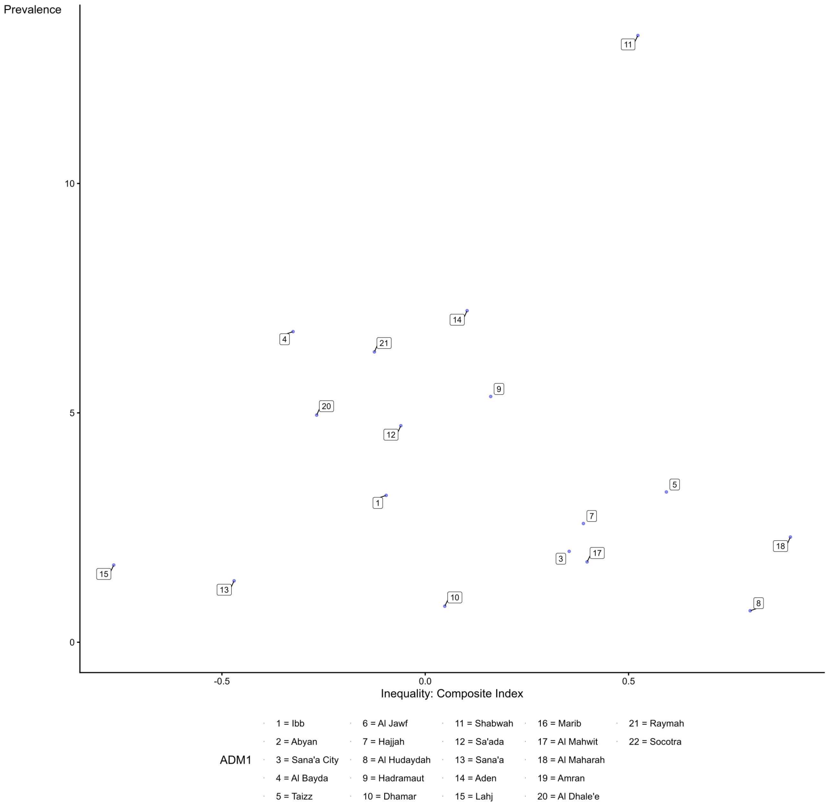

**Subnational Equity-prevalence Plane of Zero-dose Children in South Africa**

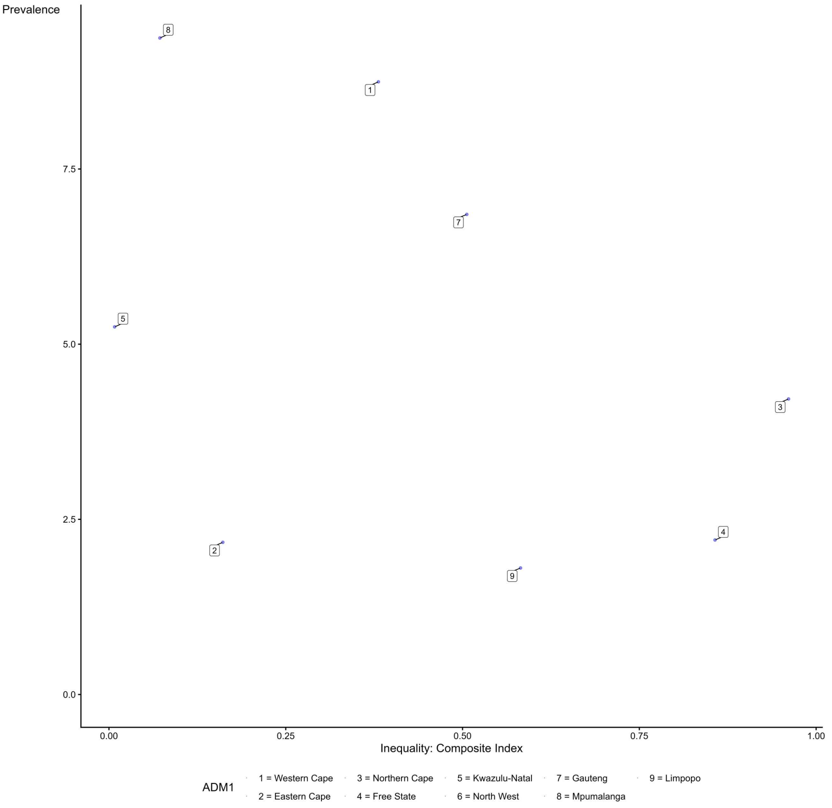

**Subnational Equity-prevalence Plane of Zero-dose Children in Zambia**

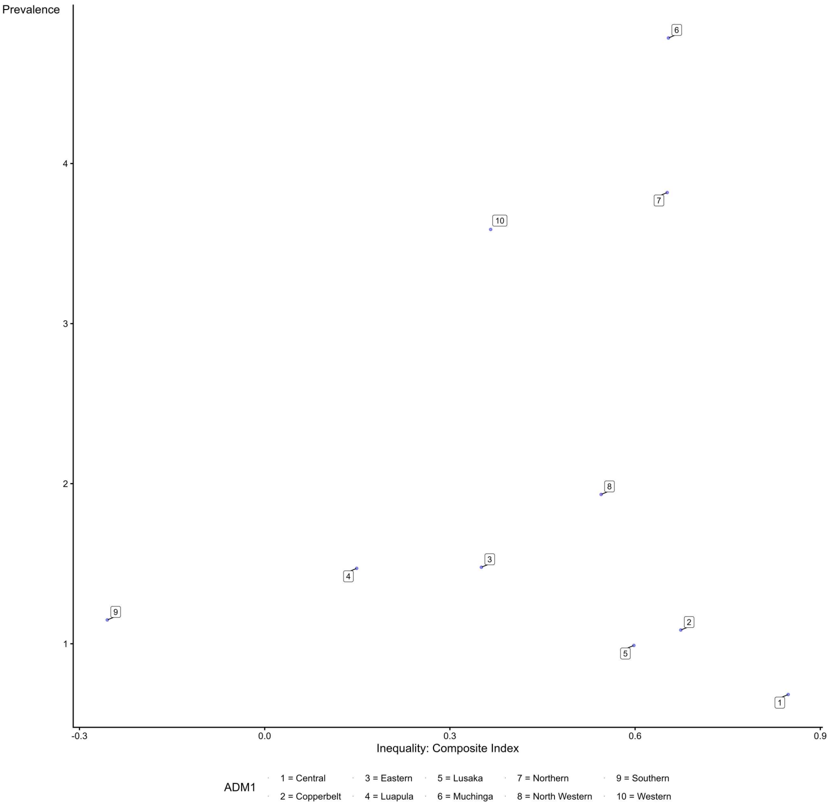

**Subnational Equity-prevalence Plane of Zero-dose Children in Zimbabwe**

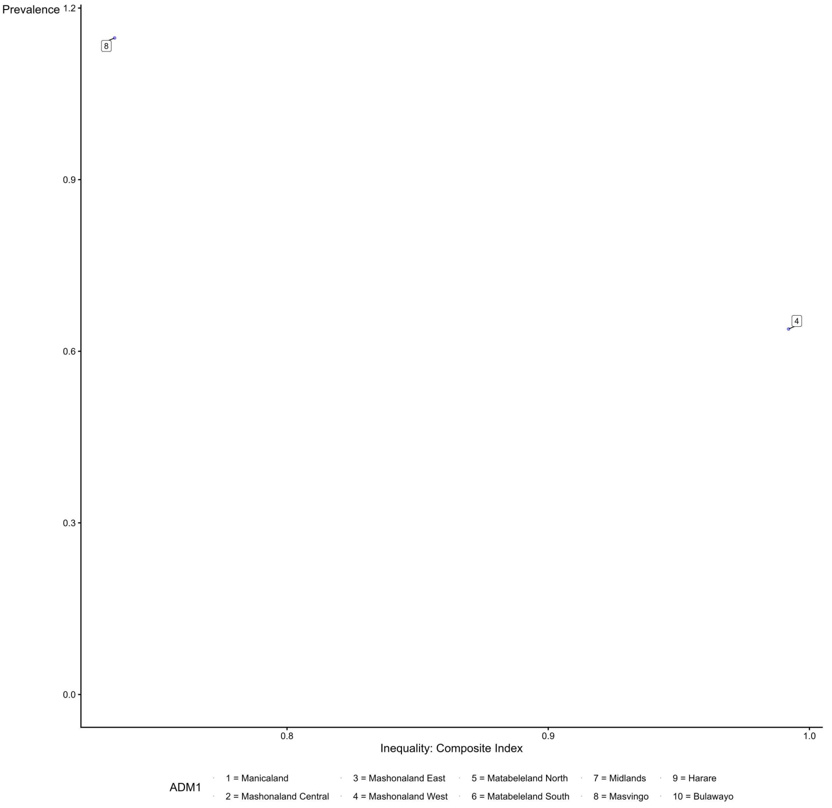

**Supplementary Fig.5 Decomposition of inequity in the prevalence of zero-dose children across DHS and MICS surveys**

Decomposition of Zero-dose Equity in Afghanistan

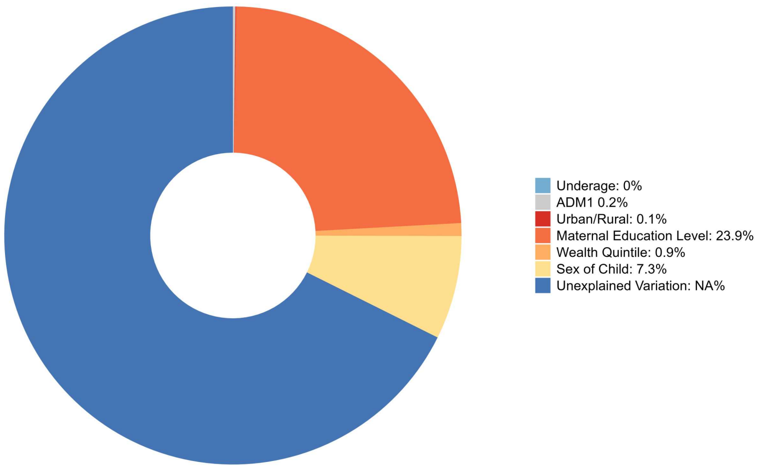

Decomposition of Zero-dose Equity in Angola

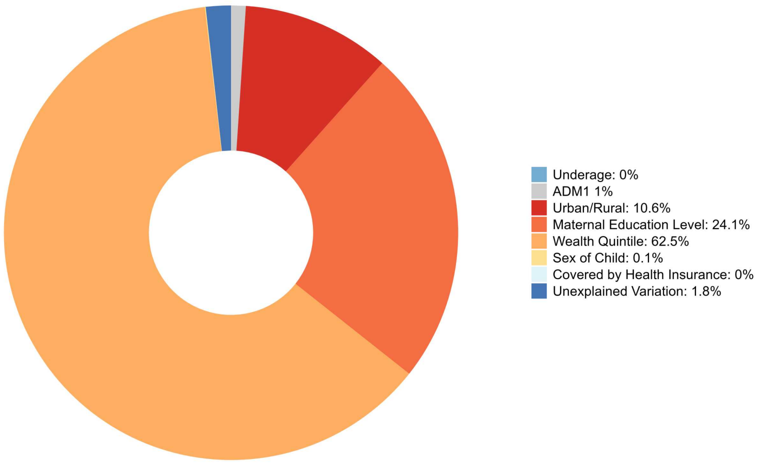

**Decomposition of Zero-dose Equity in Armenia**

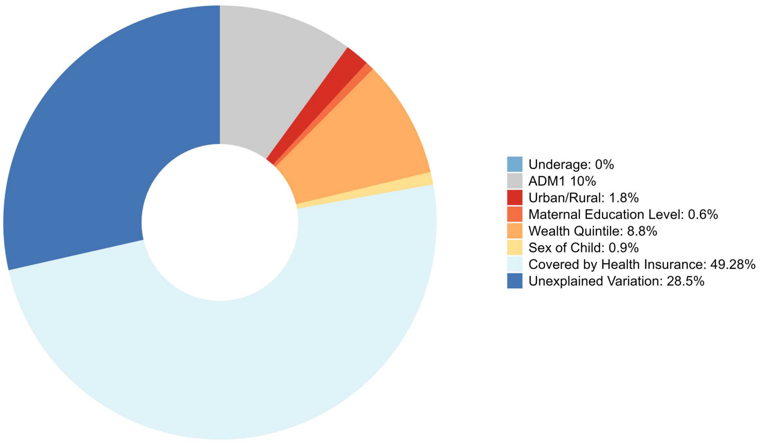

**Decomposition of Zero-dose Equity in Azerbaijan**

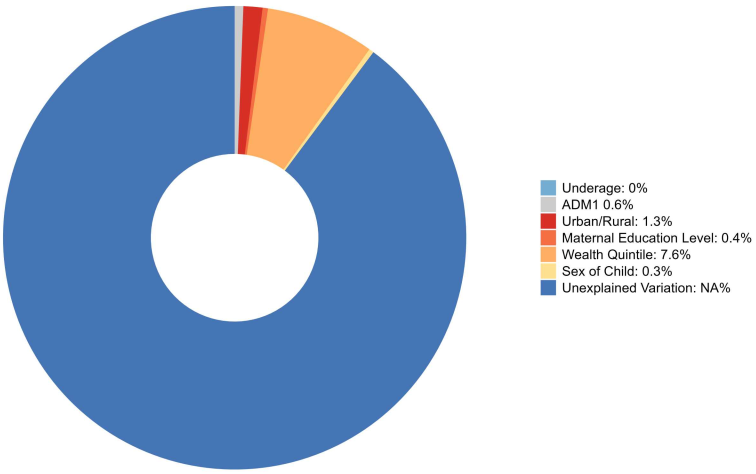

**Decomposition of Zero-dose Equity in Burundi**

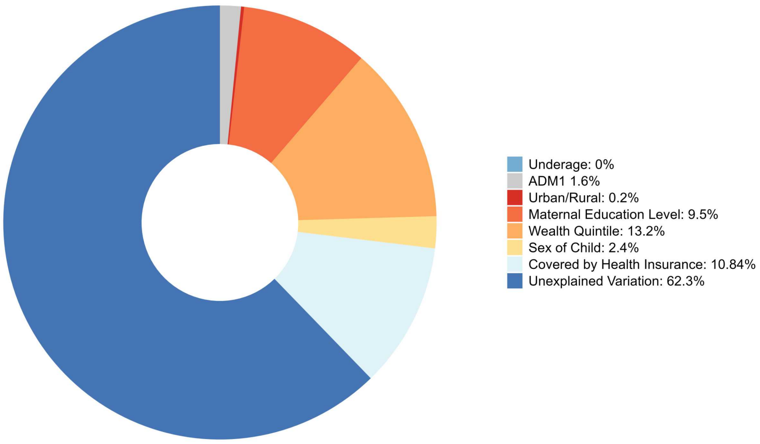

**Decomposition of Zero-dose Equity in Benin**

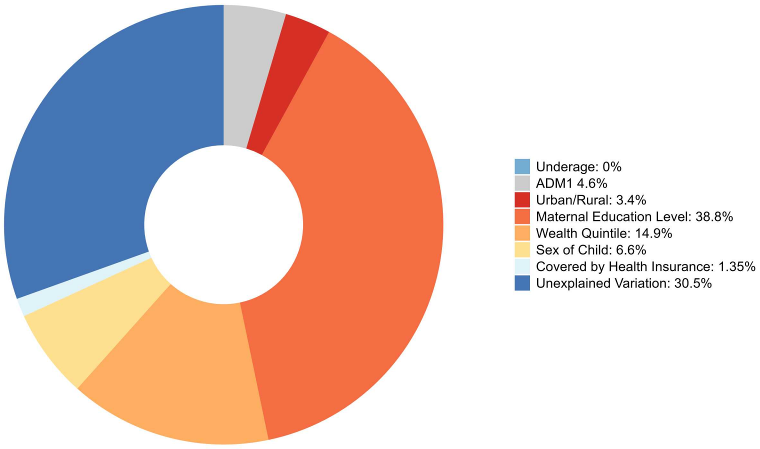

**Decomposition of Zero-dose Equity in Burkina Faso**

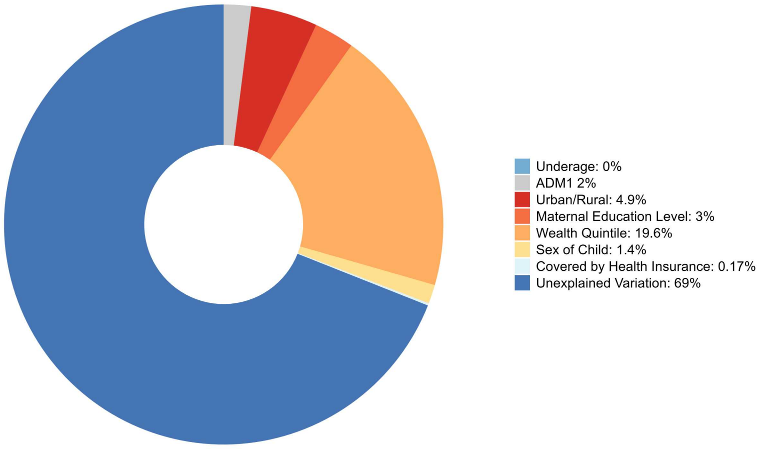

**Decomposition of Zero-dose Equity in Bangladesh**

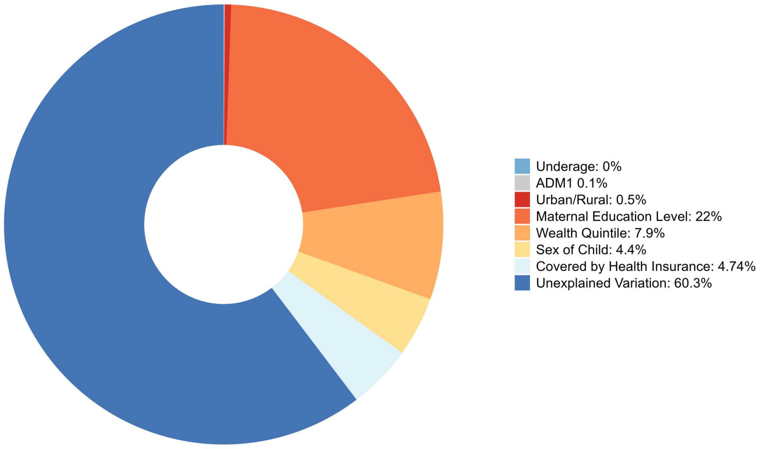

**Decomposition of Zero-dose Equity in Central African Republic**

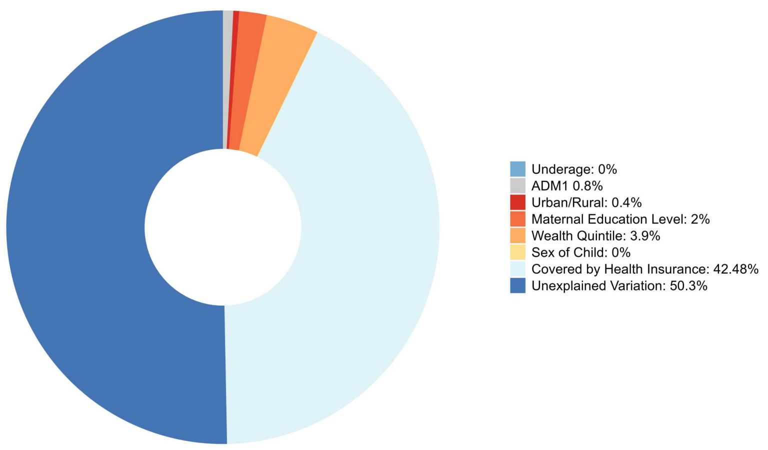

**Decomposition of Zero-dose Equity in Côte d'Ivoire**

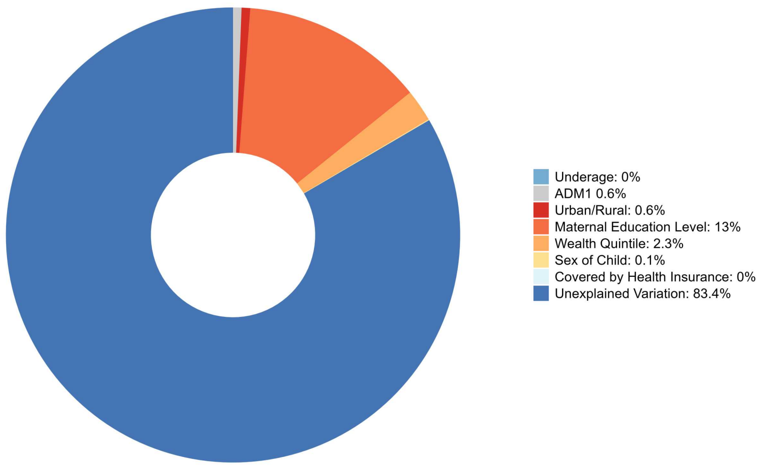

**Decomposition of Zero-dose Equity in Cameroon**

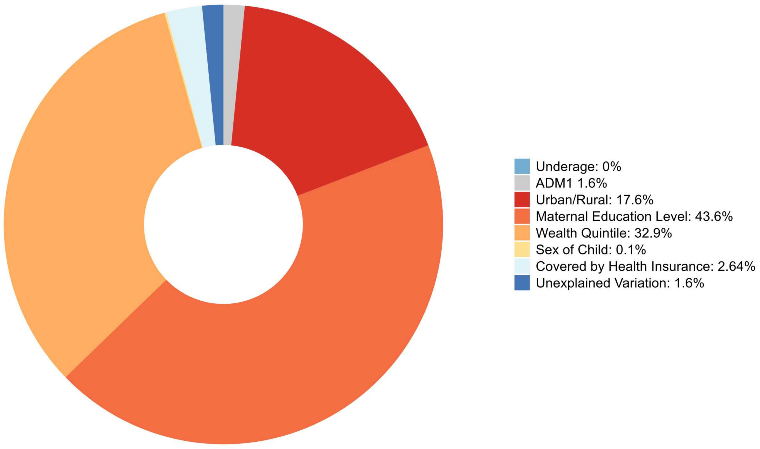

**Decomposition of Zero-dose Equity in Democratic Republic of the Congo**

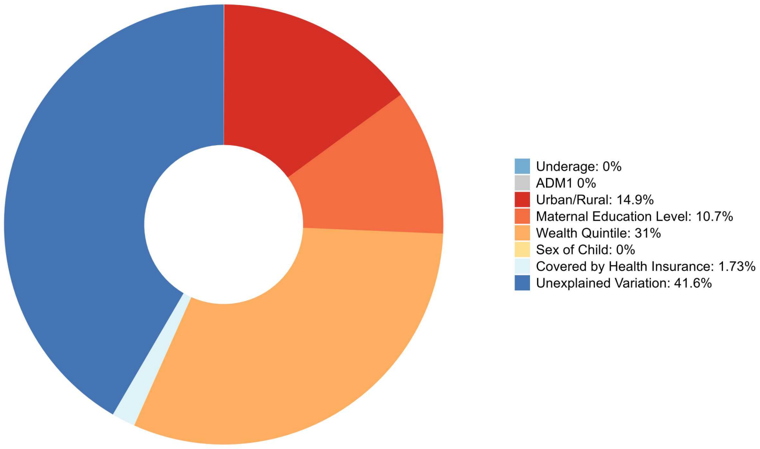

**Decomposition of Zero-dose Equity in Comoros**

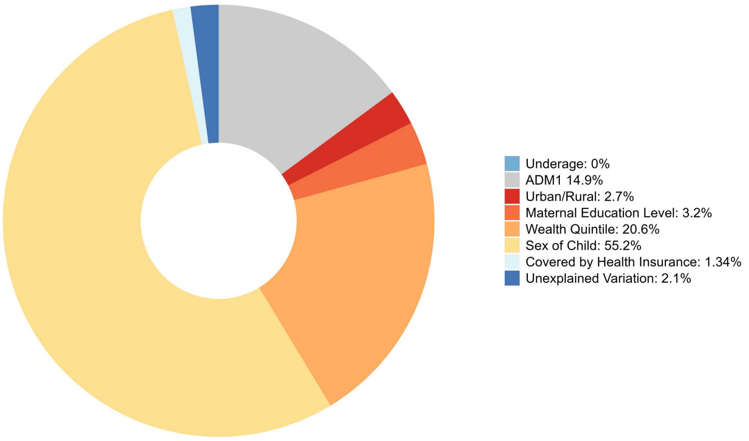

**Decomposition of Zero-dose Equity in Costa Rica**

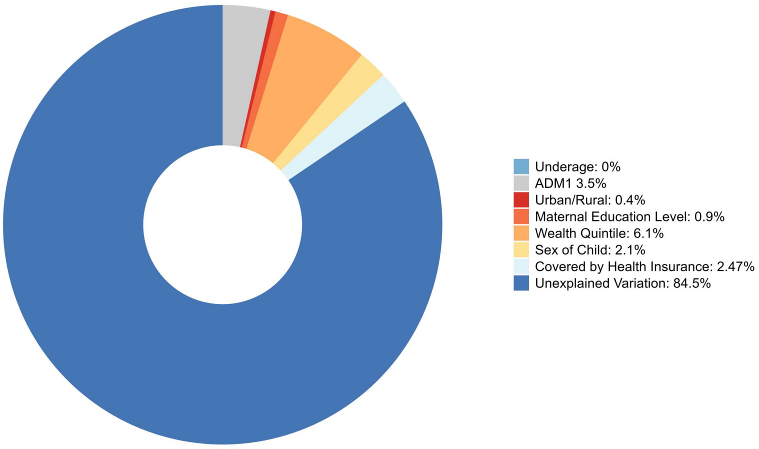

**Decomposition of Zero-dose Equity in Cuba**

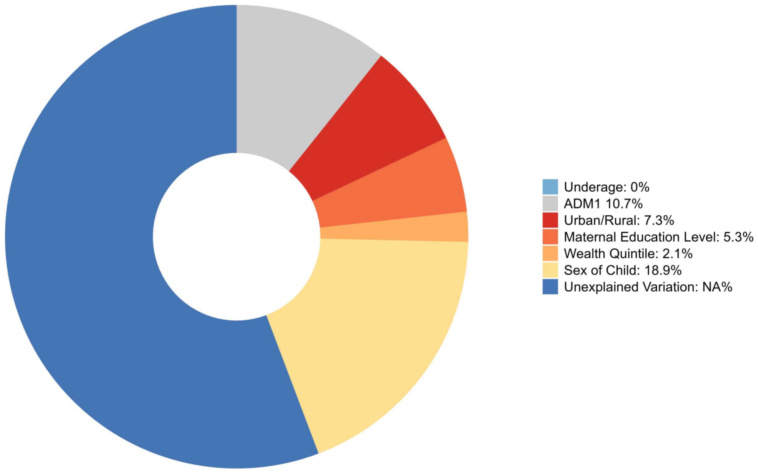

**Decomposition of Zero-dose Equity in Dominican Republic**

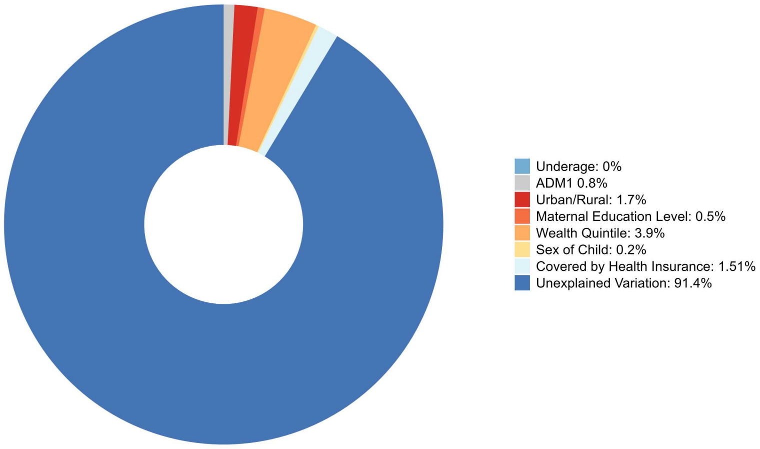

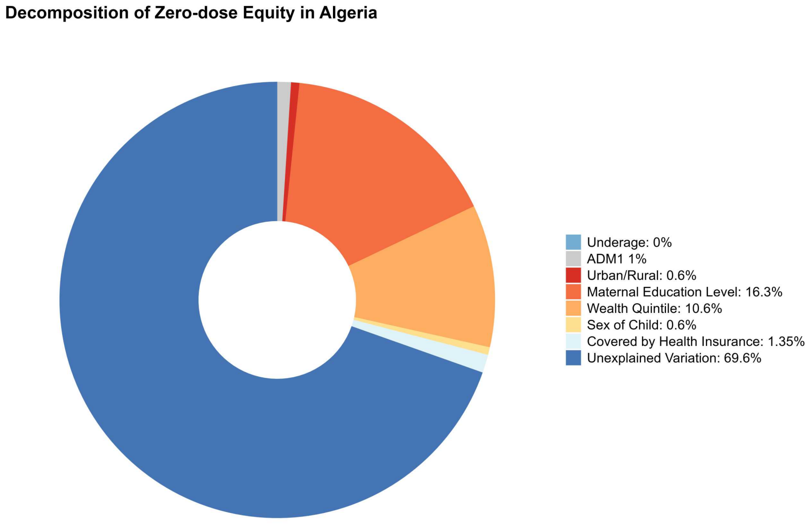
Decomposition of Zero-dose Equity in Algeria
Underage: 0%
ADM1 1%
Urban/Rural: 0.6%
Maternal Education Level: 16.3%
Wealth Quintile: 10.6%
Sex of Child: 0.6%
Covered by Health Insurance: 1.35%
Unexplained Variation: 69.6%

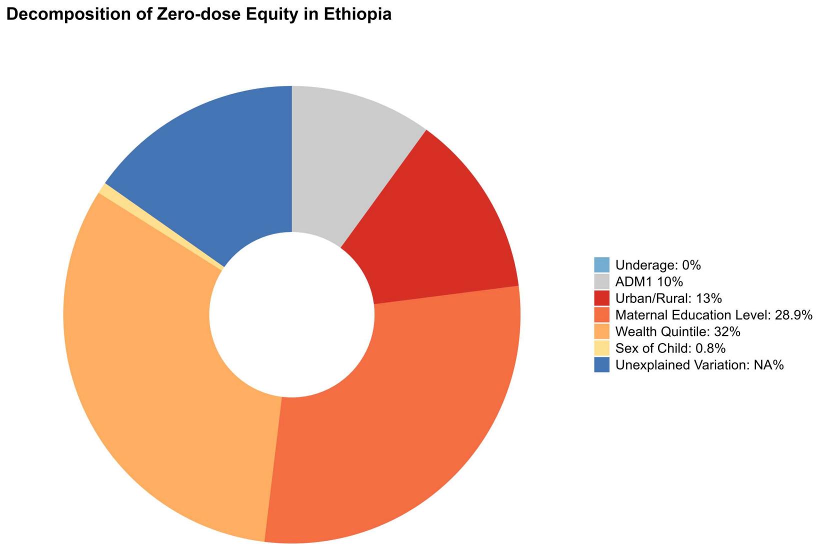
Decomposition of Zero-dose Equity in Ethiopia
Underage: 0%
ADM1 10%
Urban/Rural: 13%
Maternal Education Level: 28.9%
Wealth Quintile: 32%
Sex of Child: 0.8%
Unexplained Variation: NA%

Decomposition of Zero-dose Equity in Fiji

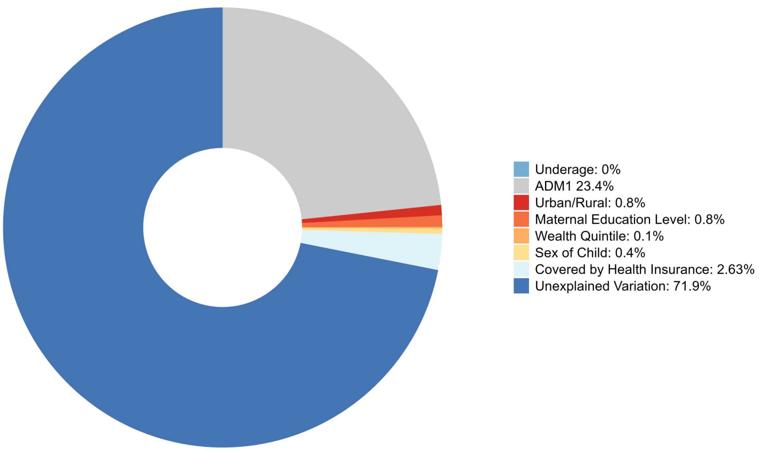

Decomposition of Zero-dose Equity in Gabon

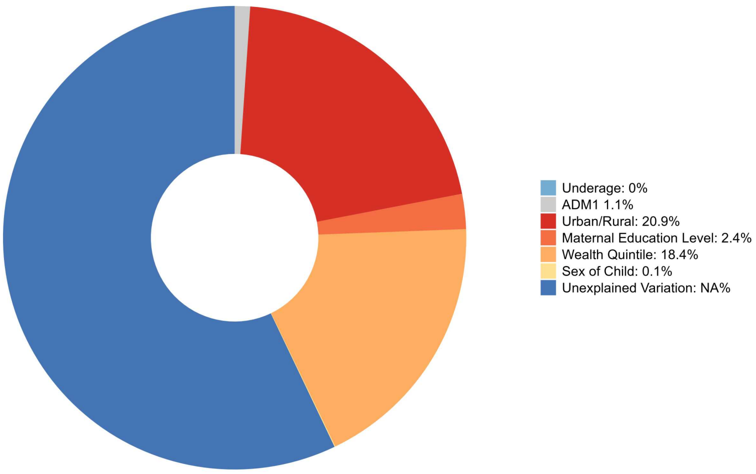

**Decomposition of Zero-dose Equity in Ghana**

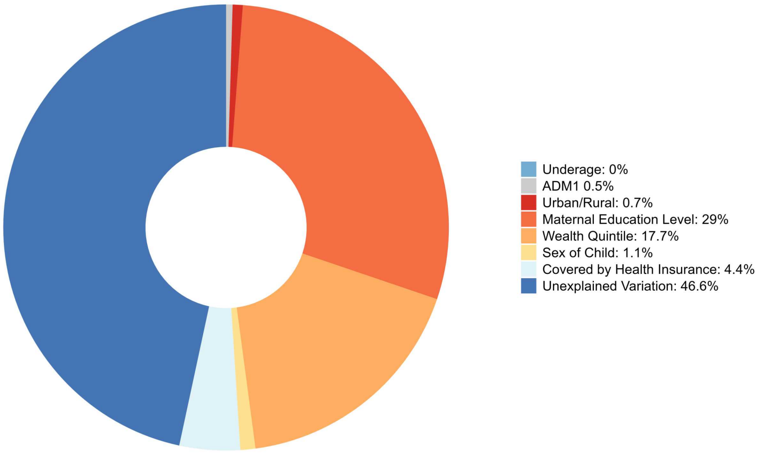

**Decomposition of Zero-dose Equity in Guinea**

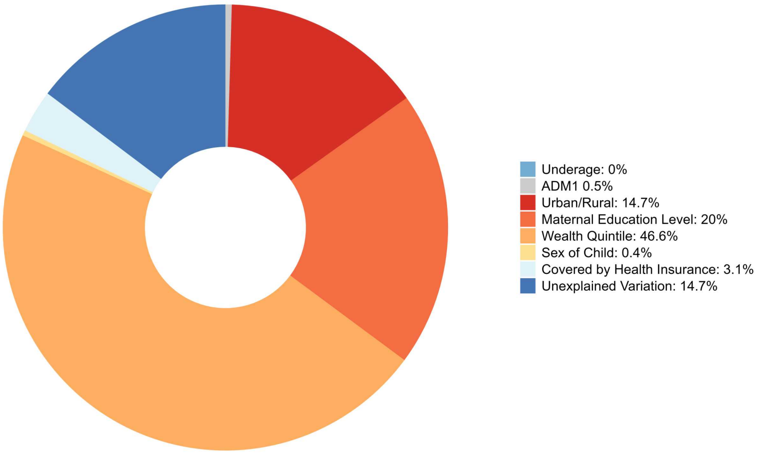

**Decomposition of Zero-dose Equity in Gambia**

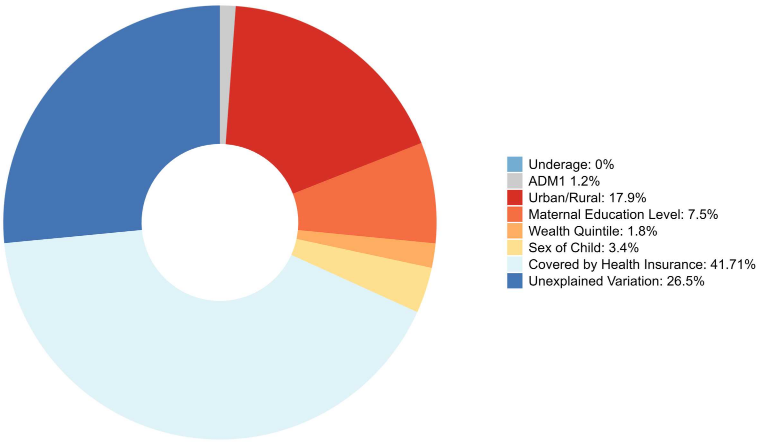

**Decomposition of Zero-dose Equity in Guinea-Bissau**

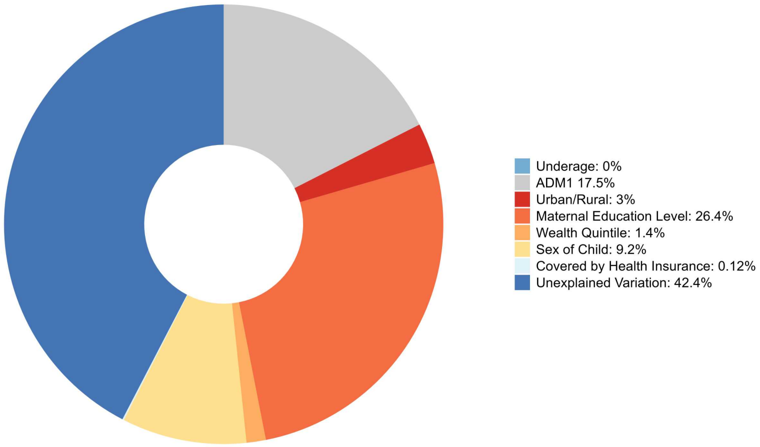

**Decomposition of Zero-dose Equity in Guatemala**

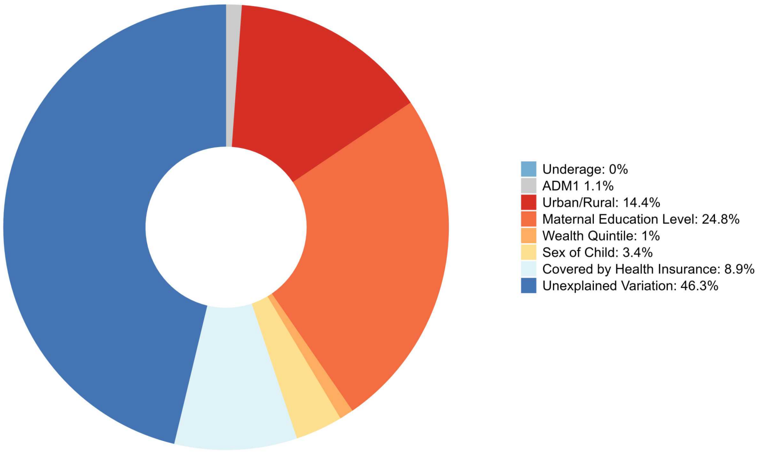

**Decomposition of Zero-dose Equity in Guyana**

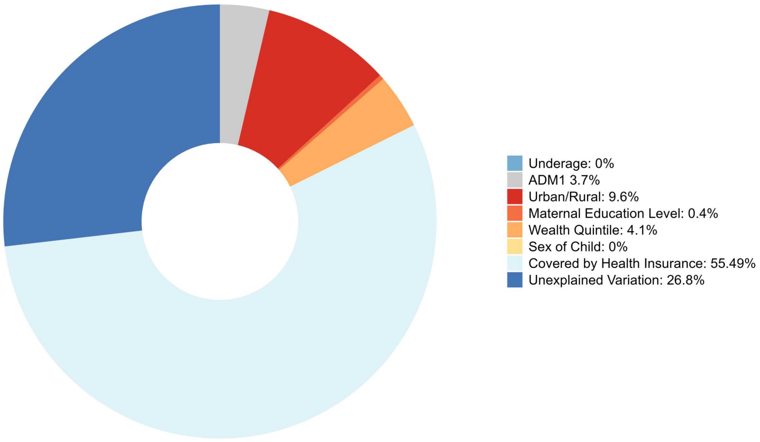

**Decomposition of Zero-dose Equity in Honduras**

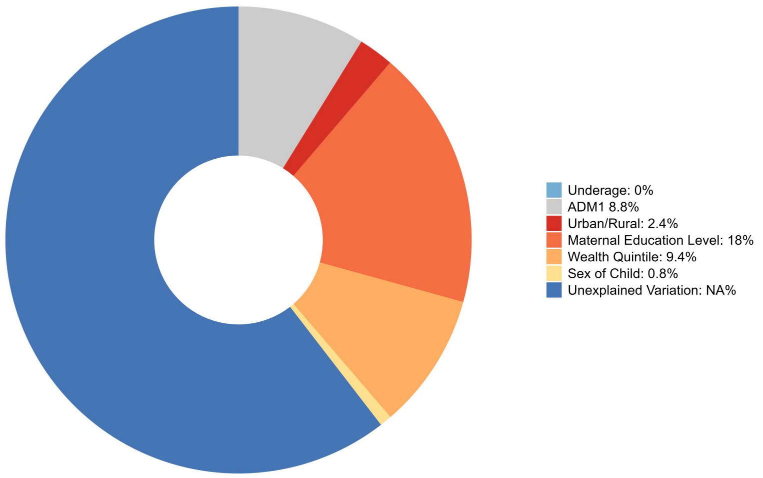

**Decomposition of Zero-dose Equity in Haiti**

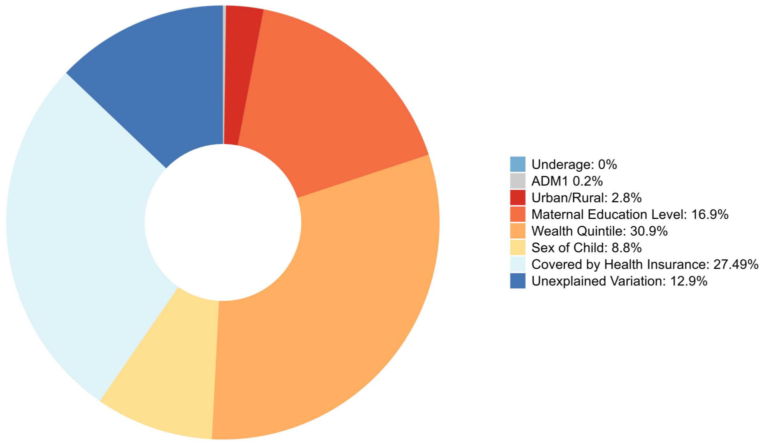

**Decomposition of Zero-dose Equity in Indonesia**

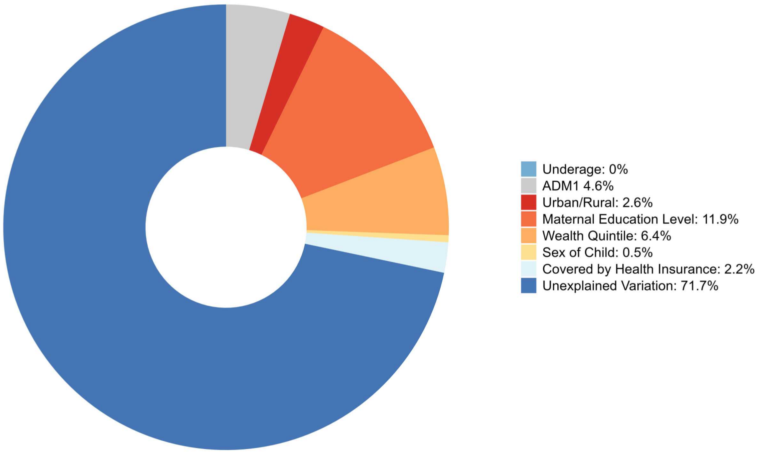

**Decomposition of Zero-dose Equity in India**

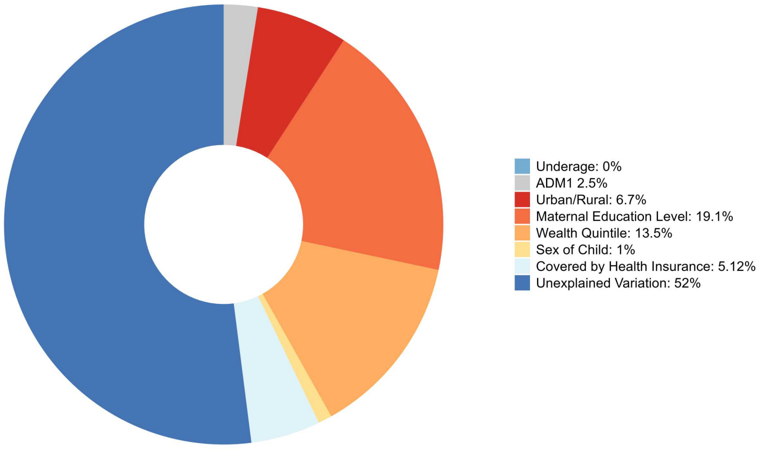

**Decomposition of Zero-dose Equity in Iraq**

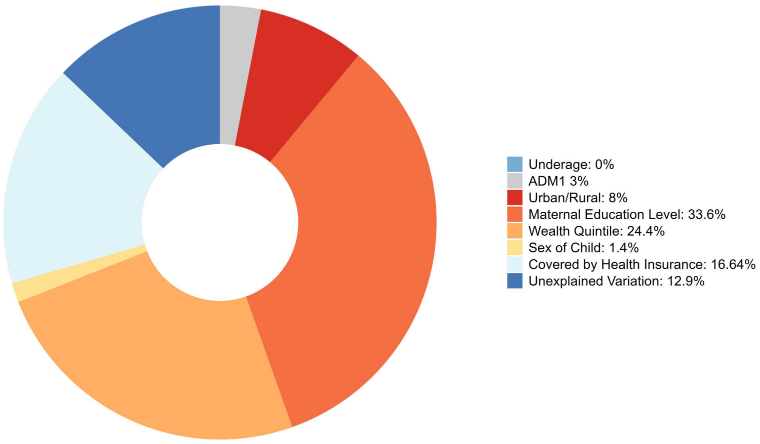

**Decomposition of Zero-dose Equity in Jordan**

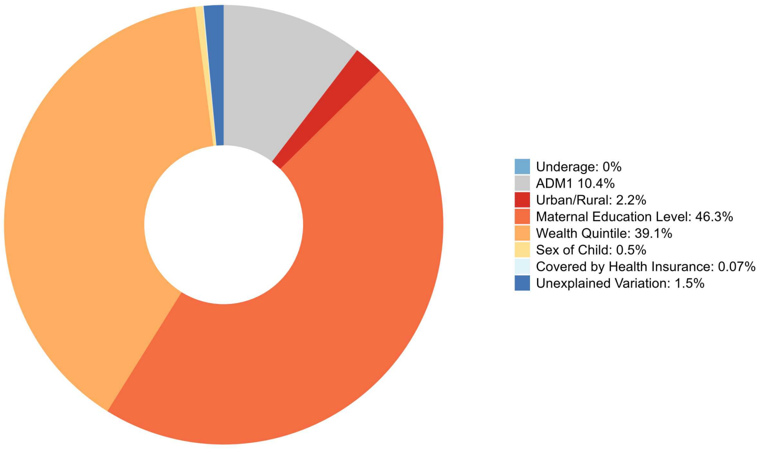

**Decomposition of Zero-dose Equity in Kenya**

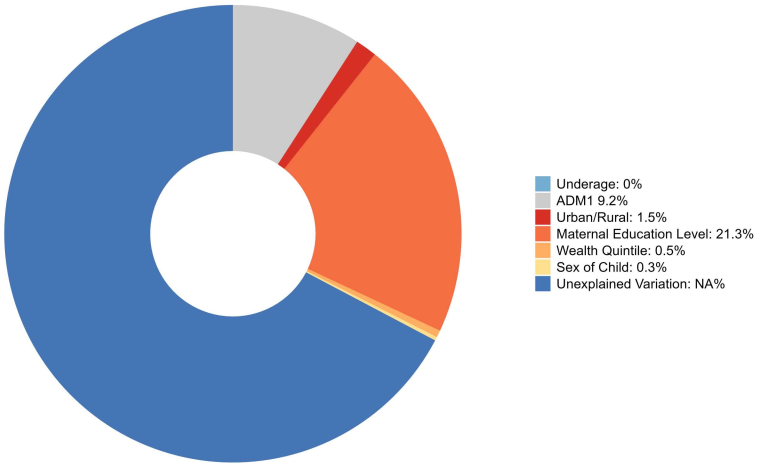

**Decomposition of Zero-dose Equity in Kyrgyzstan**

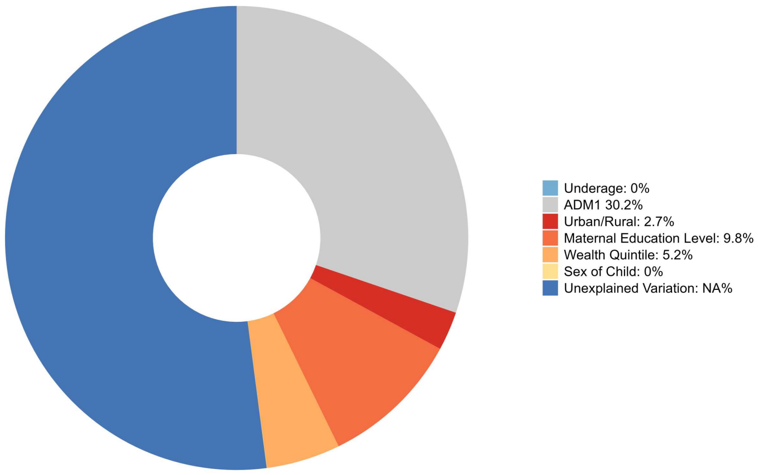

#### Decomposition of Zero-dose Equity in Cambodia

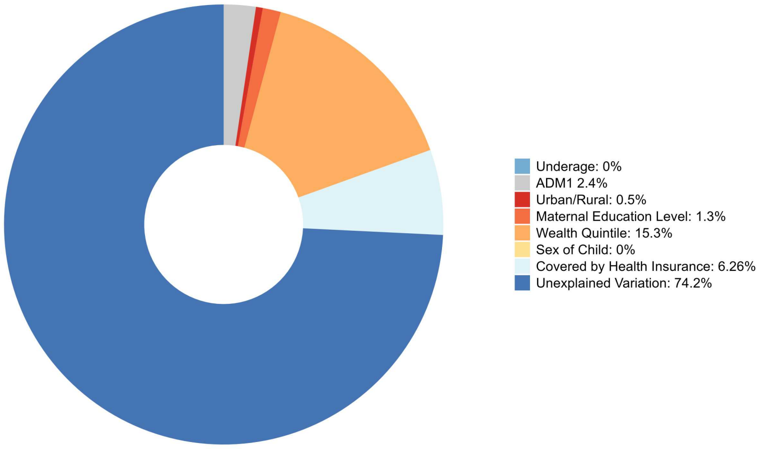

#### Decomposition of Zero-dose Equity in Laos

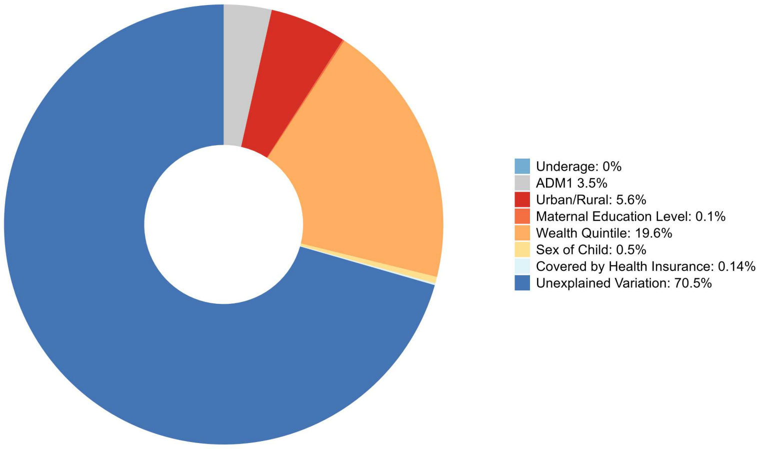

**Decomposition of Zero-dose Equity in Liberia**

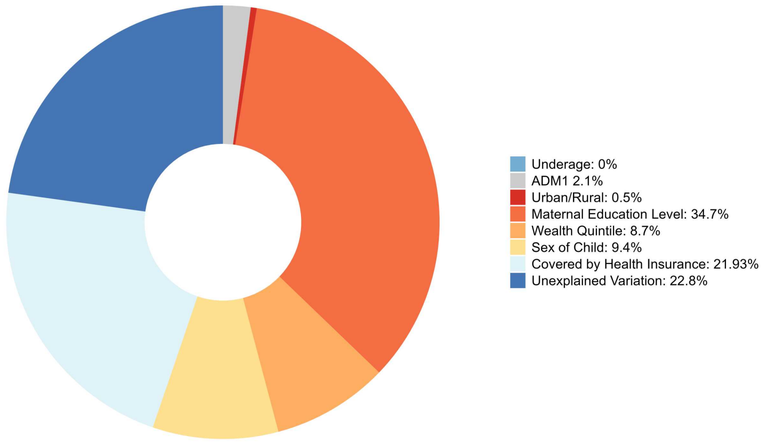

**Decomposition of Zero-dose Equity in Lesotho**

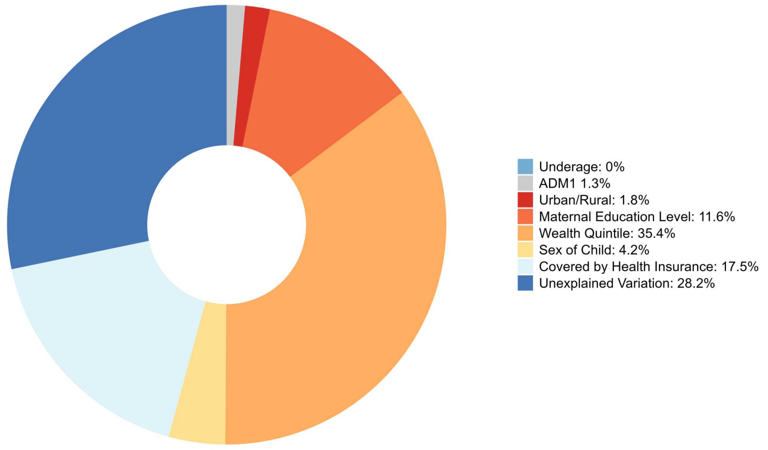

Decomposition of Zero-dose Equity in Madagascar

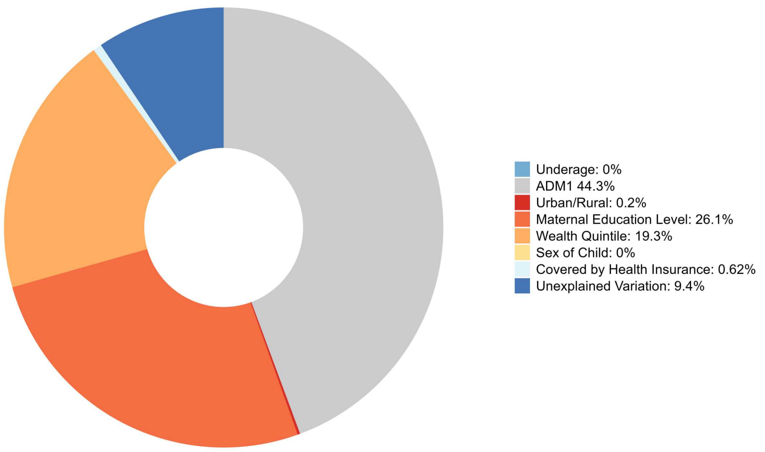

Decomposition of Zero-dose Equity in North Macedonia

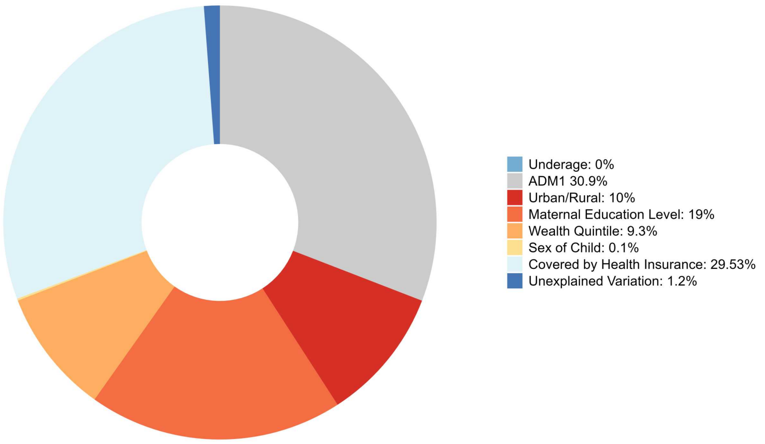

Decomposition of Zero-dose Equity in Mali

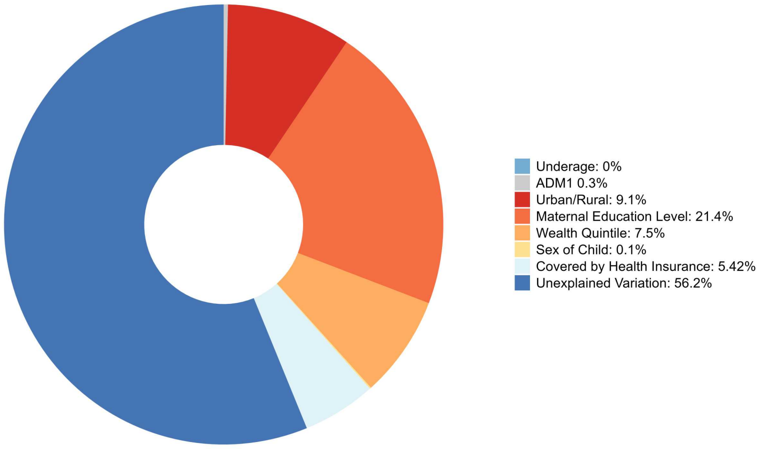

Decomposition of Zero-dose Equity in Myanmar

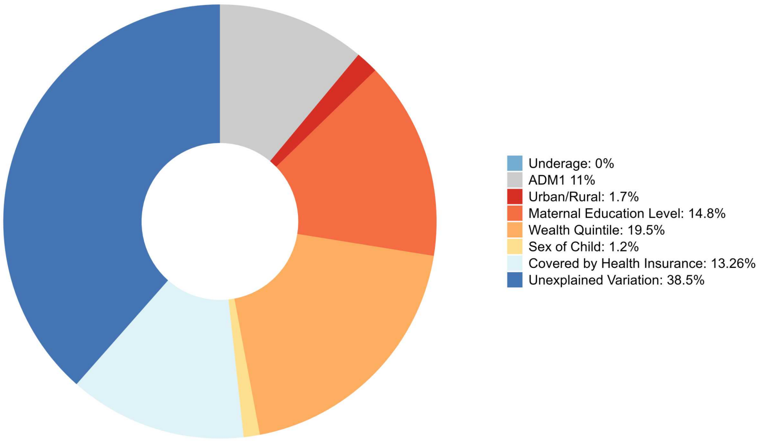

Decomposition of Zero-dose Equity in Mongolia

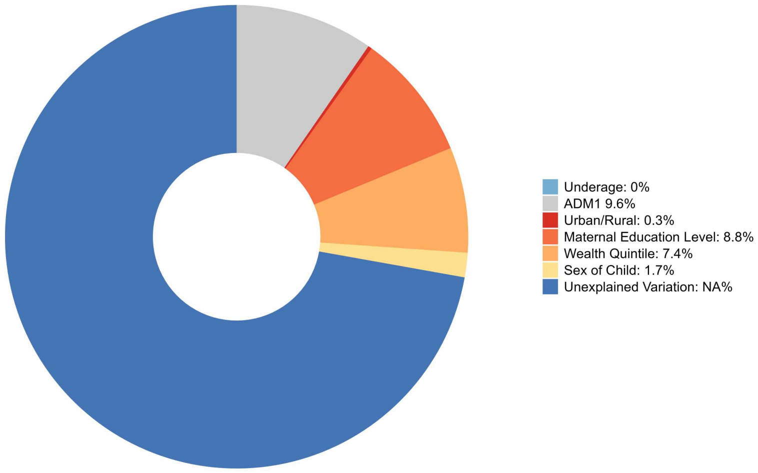

Decomposition of Zero-dose Equity in Mozambique

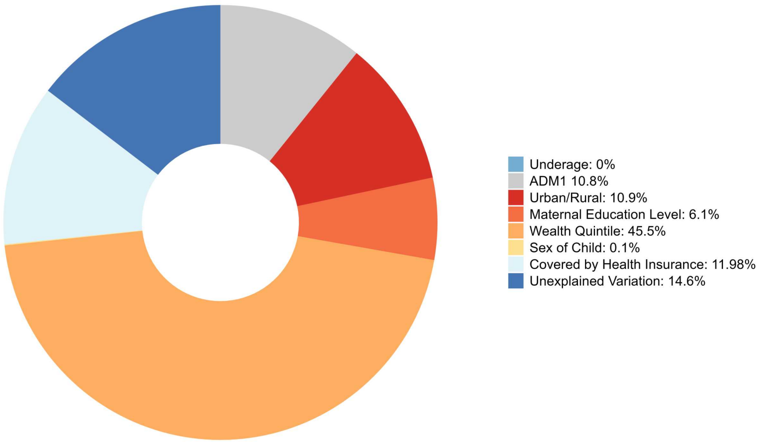

**Decomposition of Zero-dose Equity in Mauritania**

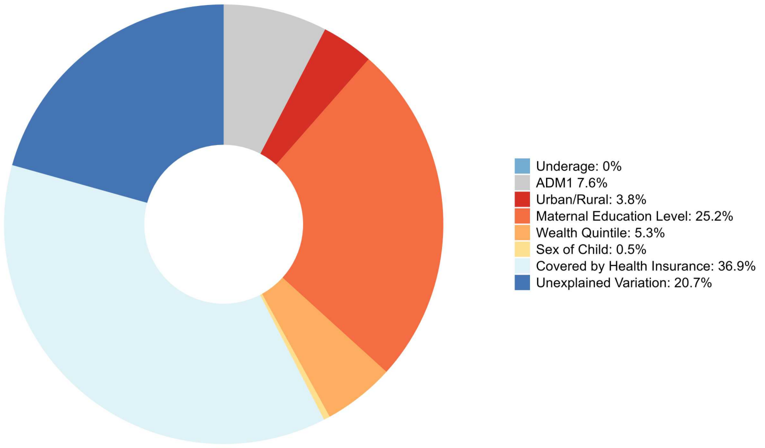

**Decomposition of Zero-dose Equity in Malawi**

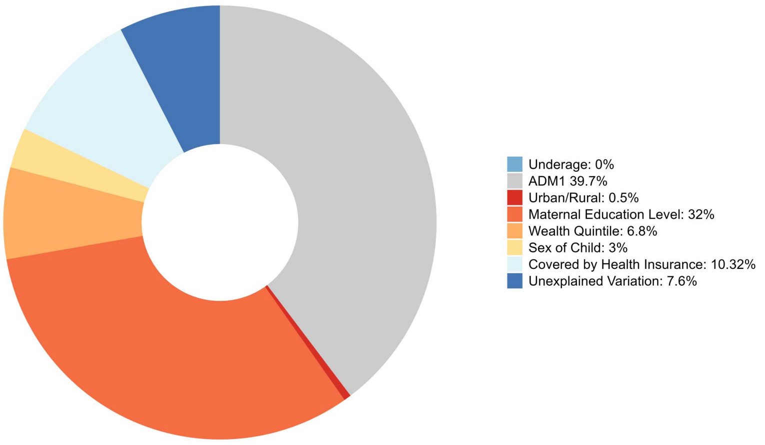

**Decomposition of Zero-dose Equity in Nigeria**

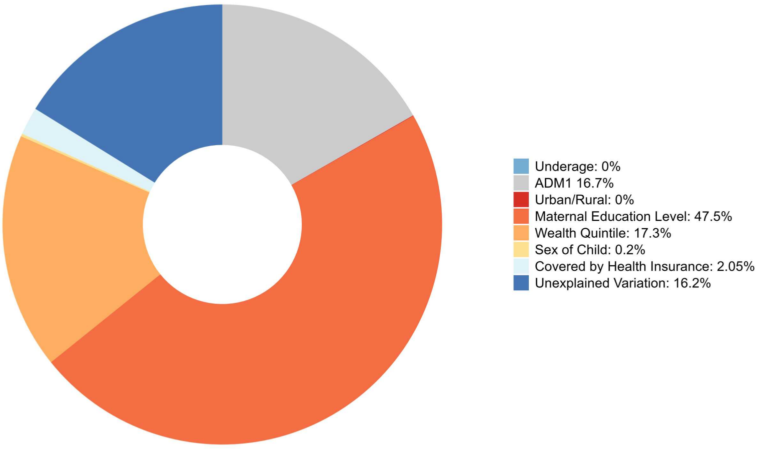

**Decomposition of Zero-dose Equity in Nepal**

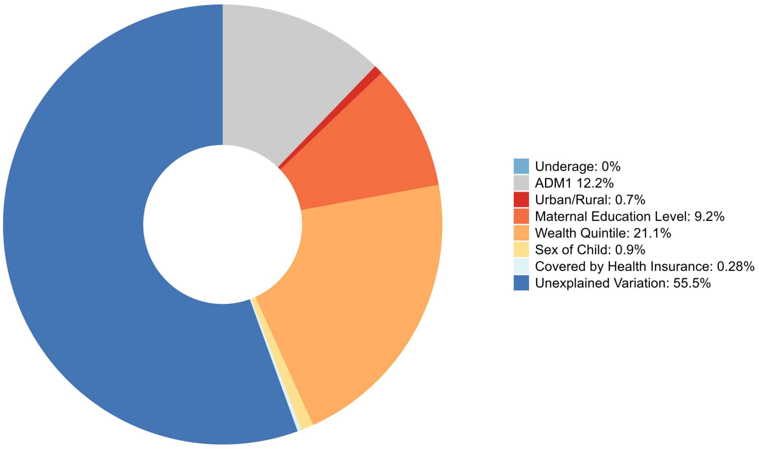

Decomposition of Zero-dose Equity in Pakistan

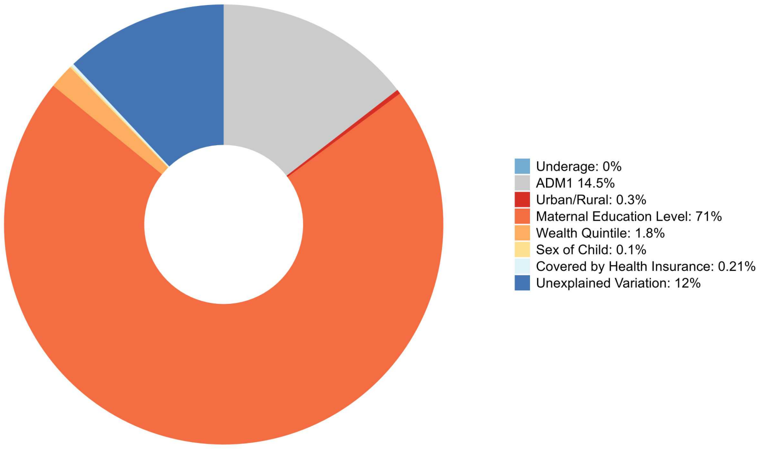

Decomposition of Zero-dose Equity in Philippines

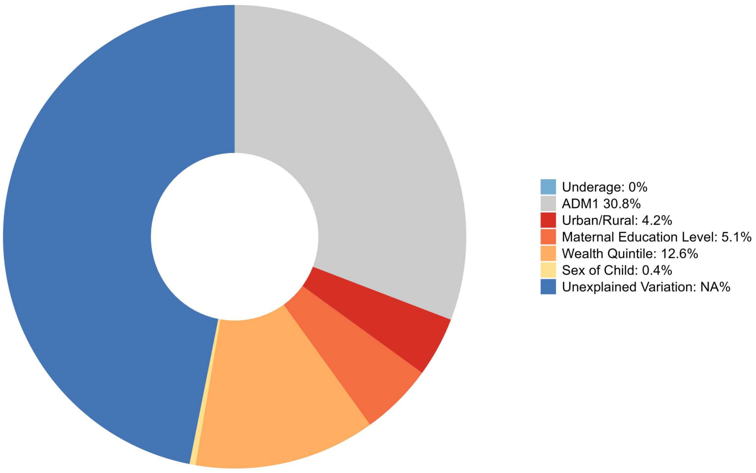

**Decomposition of Zero-dose Equity in Papua New Guinea**

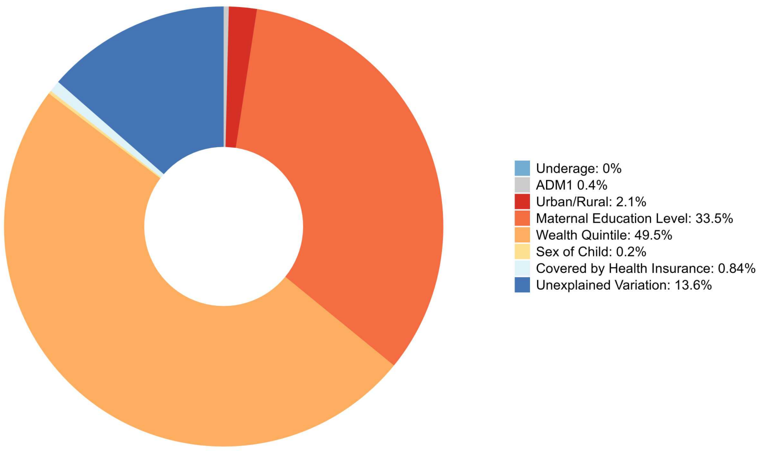

**Decomposition of Zero-dose Equity in State of Palestine**

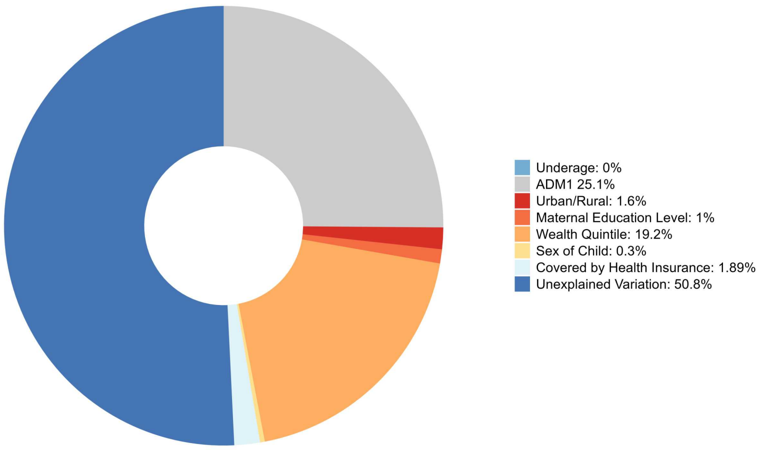

**Decomposition of Zero-dose Equity in Rwanda**

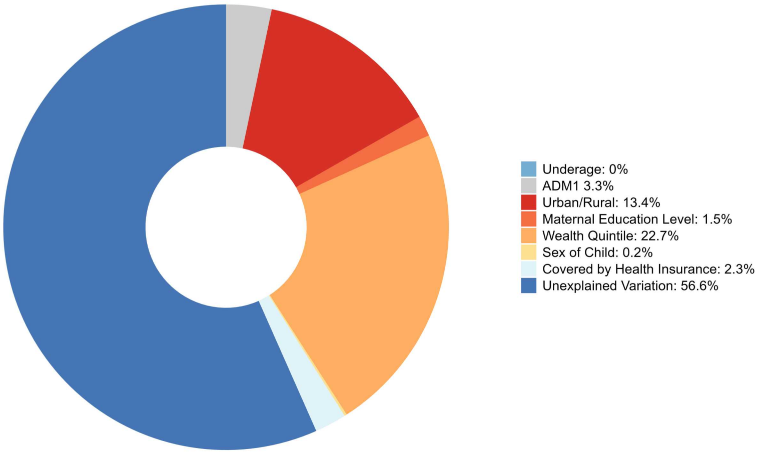

**Decomposition of Zero-dose Equity in Senegal**

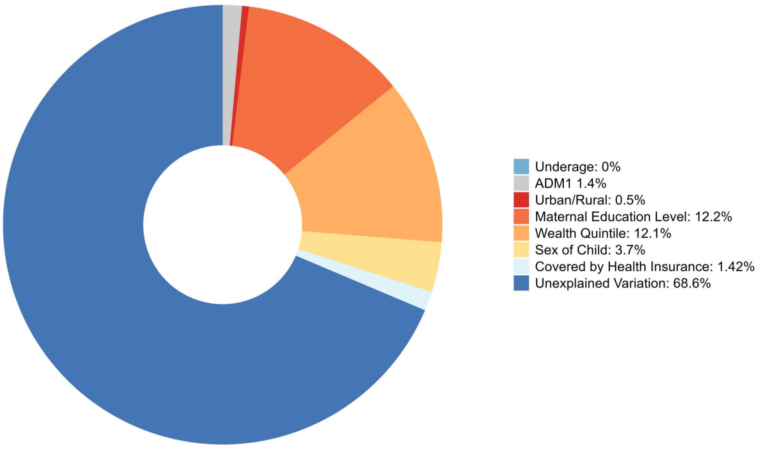

**Decomposition of Zero-dose Equity in Sierra Leone**

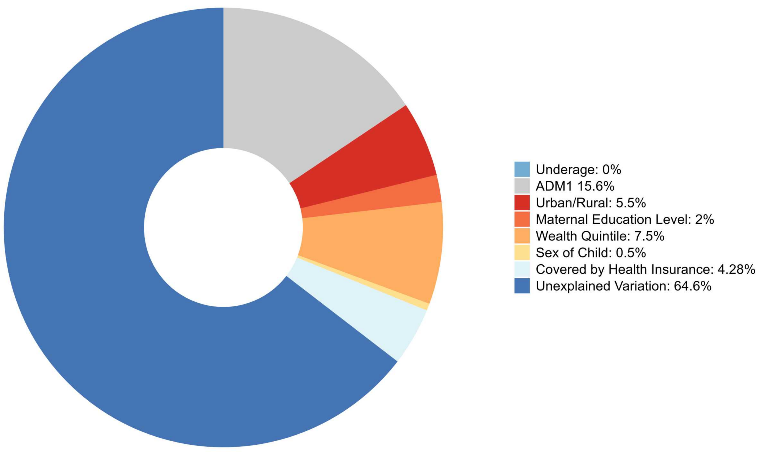

**Decomposition of Zero-dose Equity in Serbia**

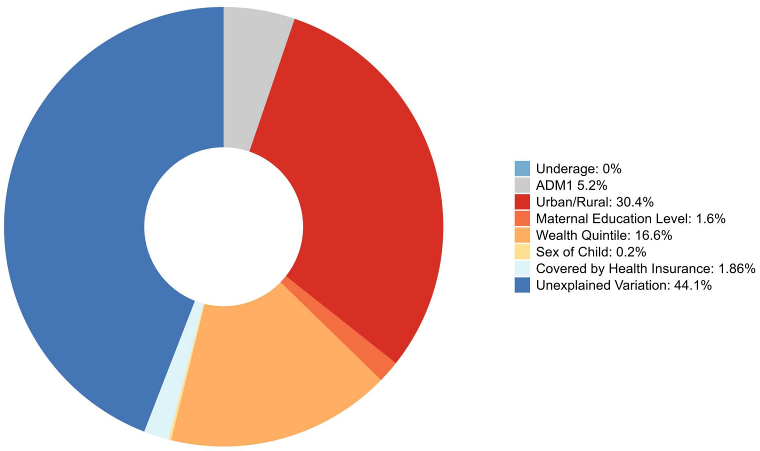

**Decomposition of Zero-dose Equity in Sao Tome and Principe**

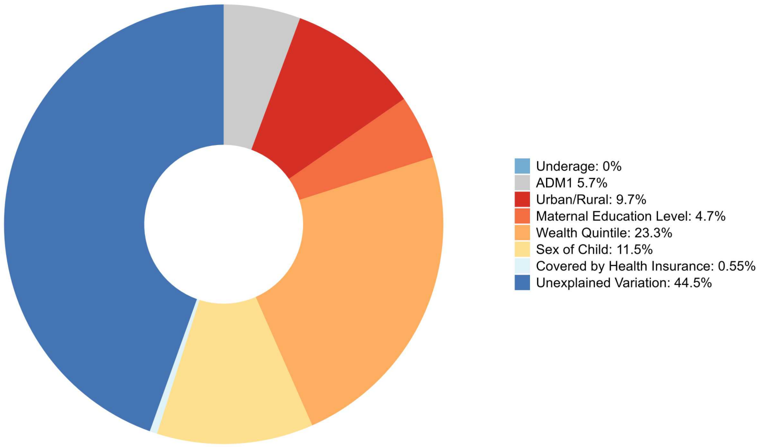

**Decomposition of Zero-dose Equity in Suriname**

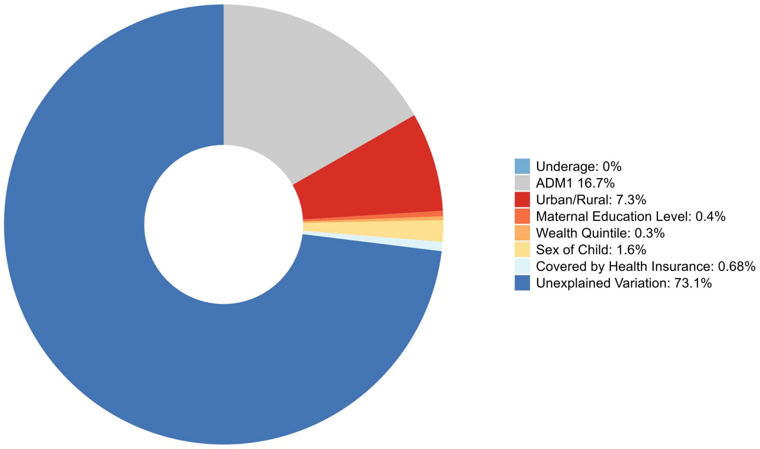

**Decomposition of Zero-dose Equity in Eswatini**

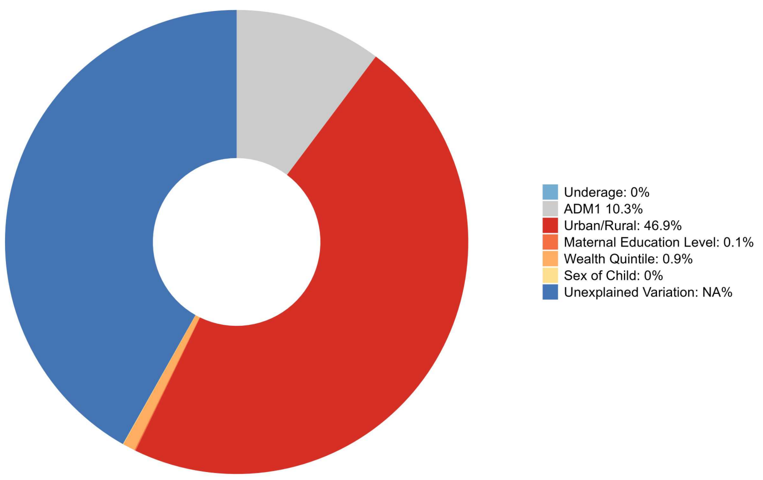

**Decomposition of Zero-dose Equity in Chad**

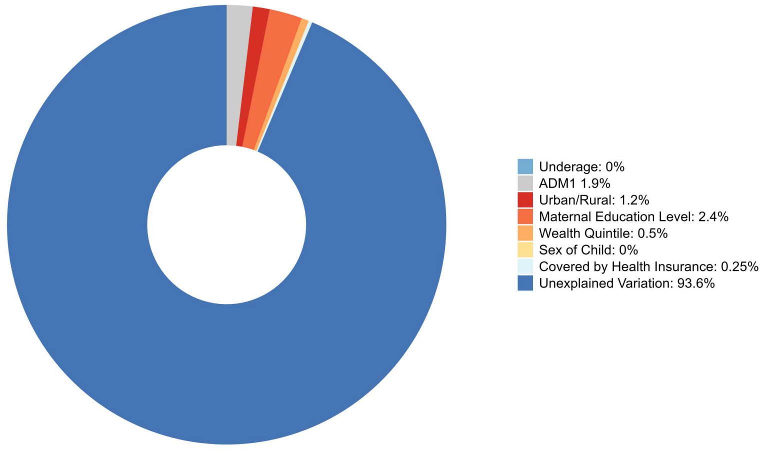

**Decomposition of Zero-dose Equity in Togo**

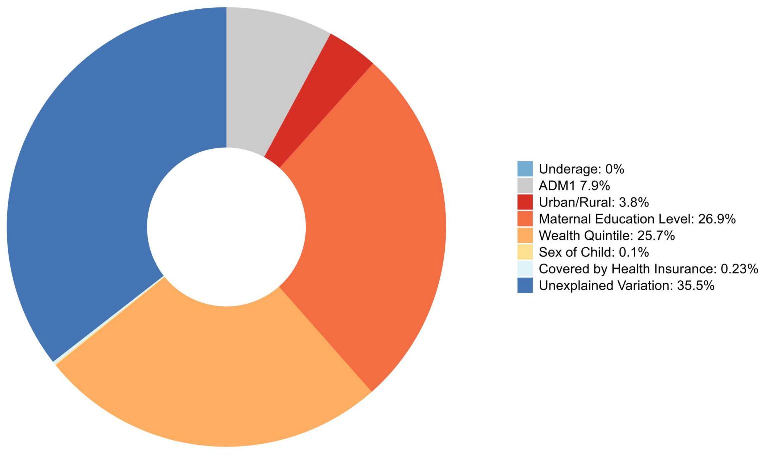

**Decomposition of Zero-dose Equity in Thailand**

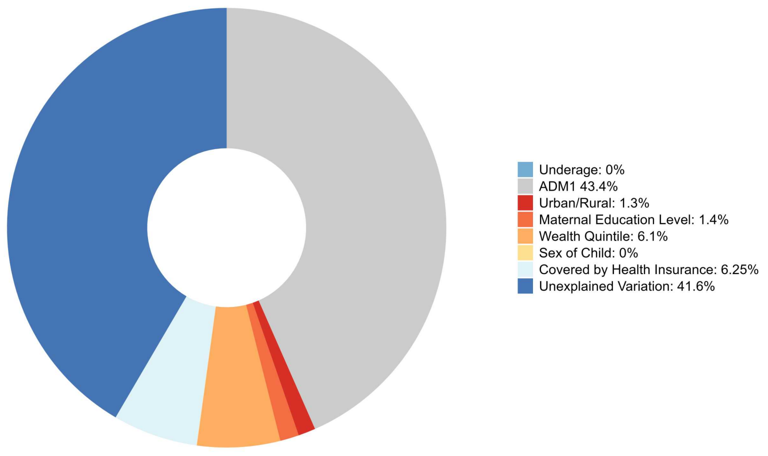

**Decomposition of Zero-dose Equity in Tajikistan**

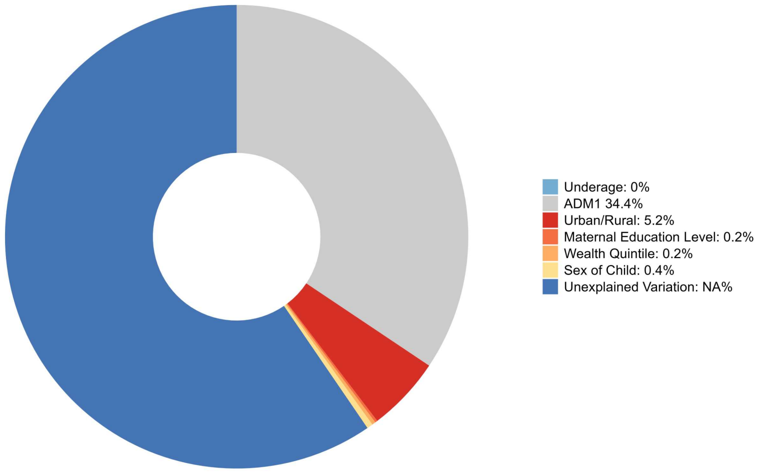

**Decomposition of Zero-dose Equity in Timor-Leste**

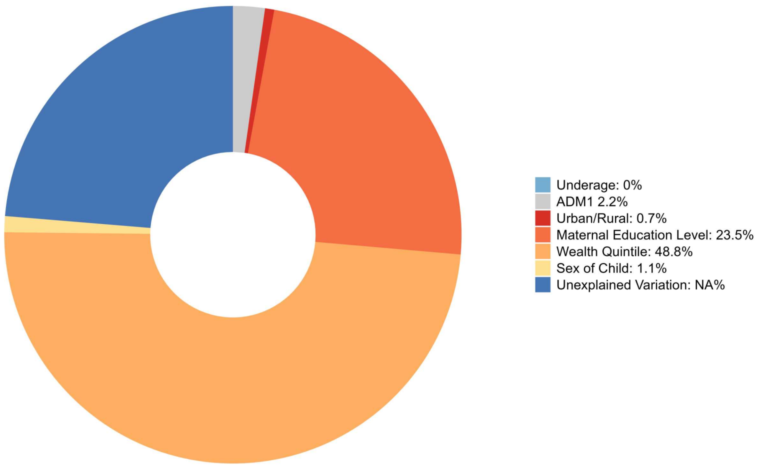

**Decomposition of Zero-dose Equity in Tonga**

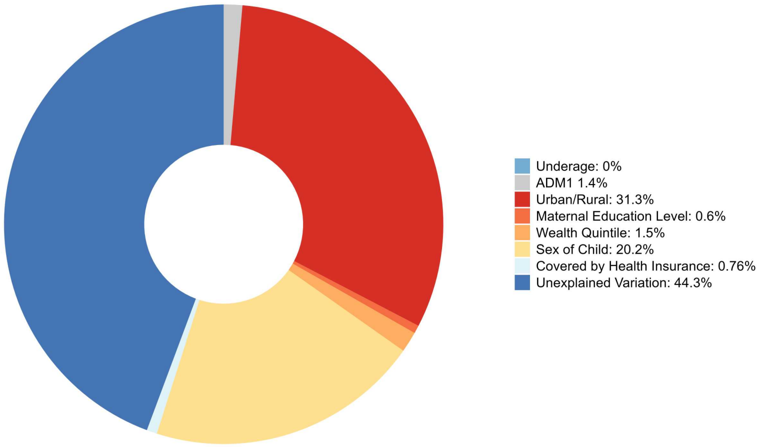

**Decomposition of Zero-dose Equity in Trinidad and Tobago**

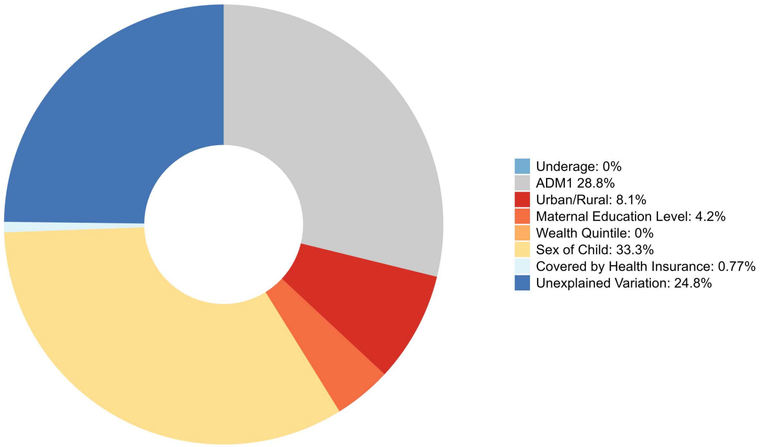

**Decomposition of Zero-dose Equity in Tunisia**

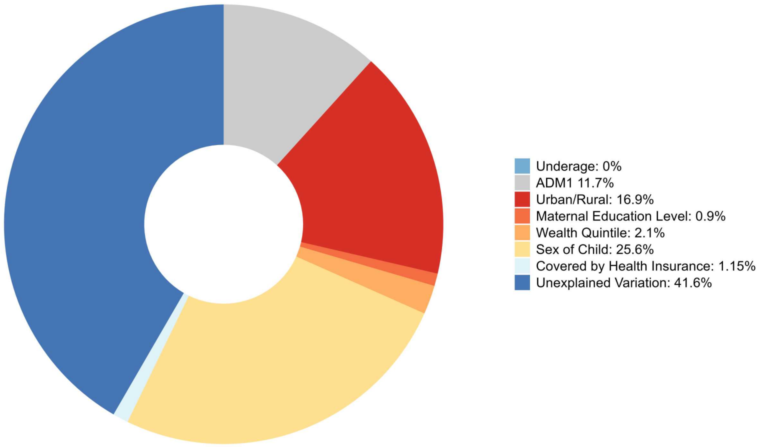

**Decomposition of Zero-dose Equity in Turkey**

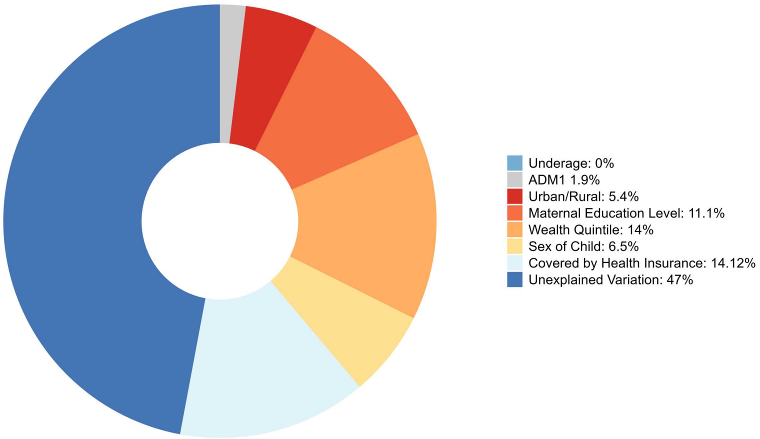

**Decomposition of Zero-dose Equity in Tanzania**

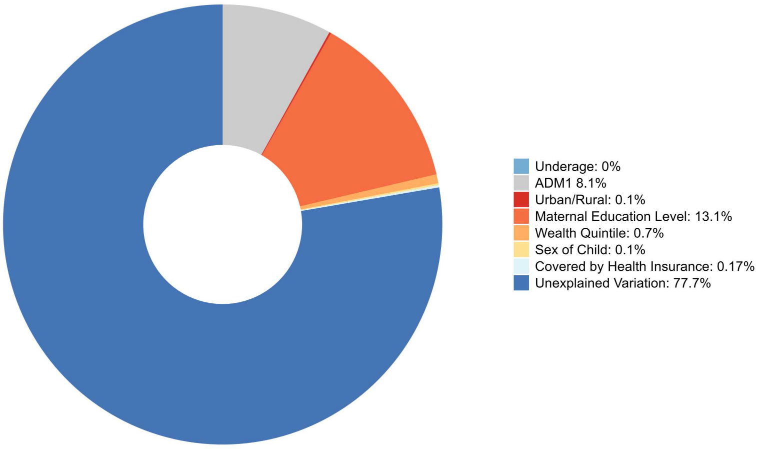

**Decomposition of Zero-dose Equity in Uganda**

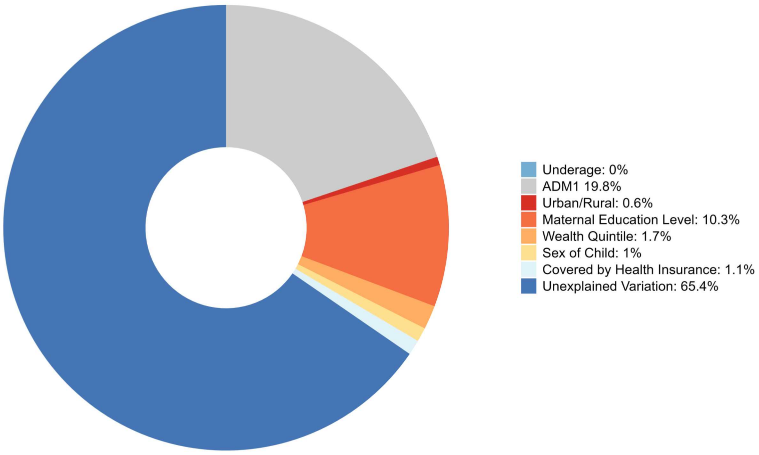

**Decomposition of Zero-dose Equity in Vietnam**

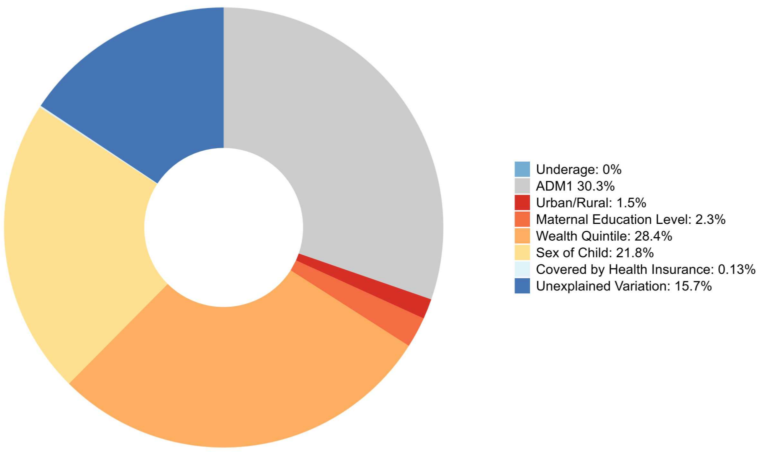

**Decomposition of Zero-dose Equity in Uzbekistan**

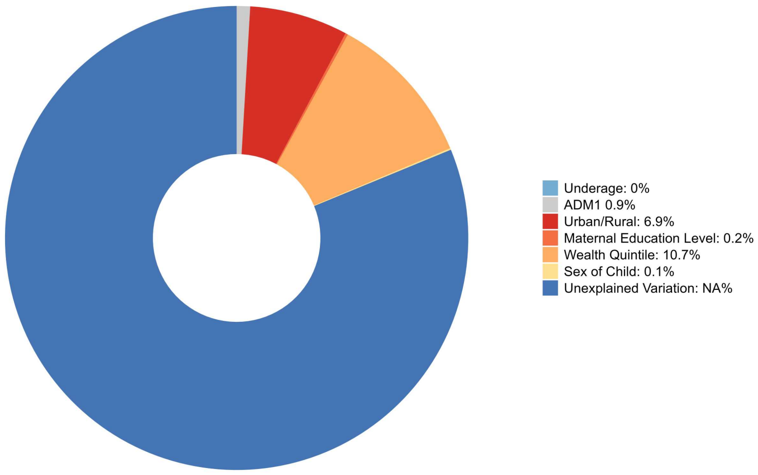

Decomposition of Zero-dose Equity in Vanuatu

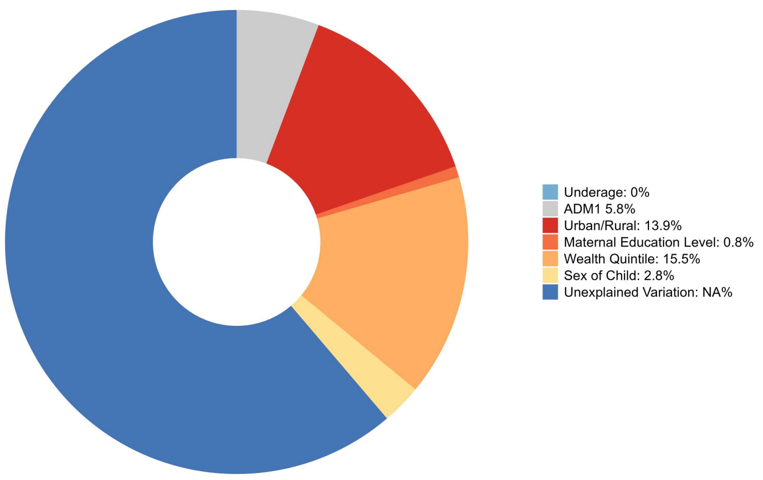

Decomposition of Zero-dose Equity in Yemen

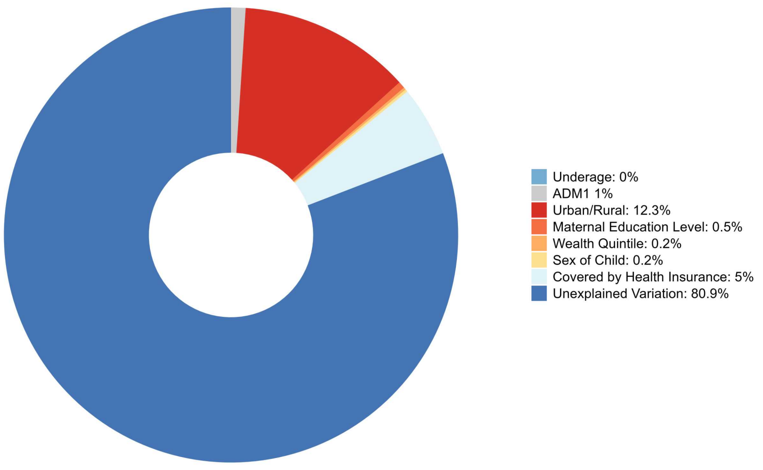

#### Decomposition of Zero-dose Equity in South Africa

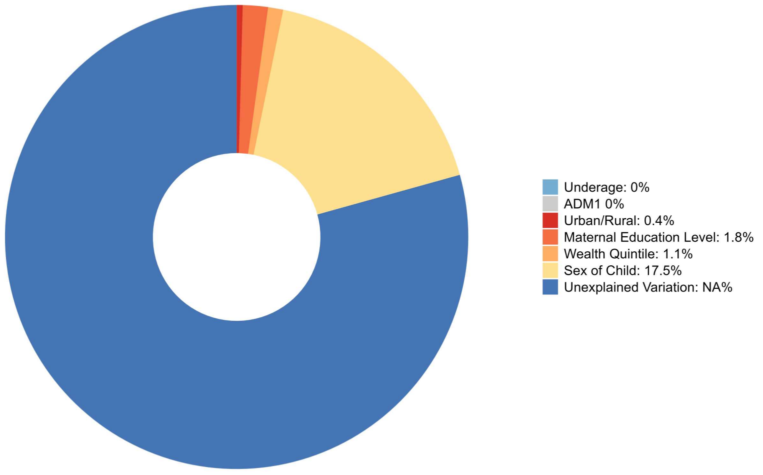

#### Decomposition of Zero-dose Equity in Zambia

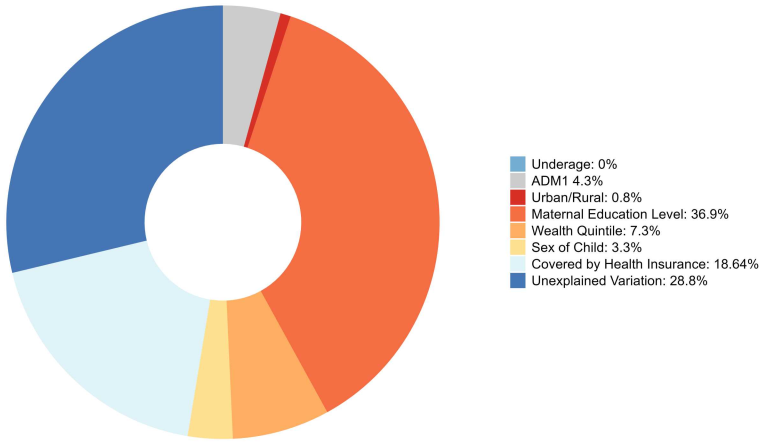

**Decomposition of Zero-dose Equity in Zimbabwe**

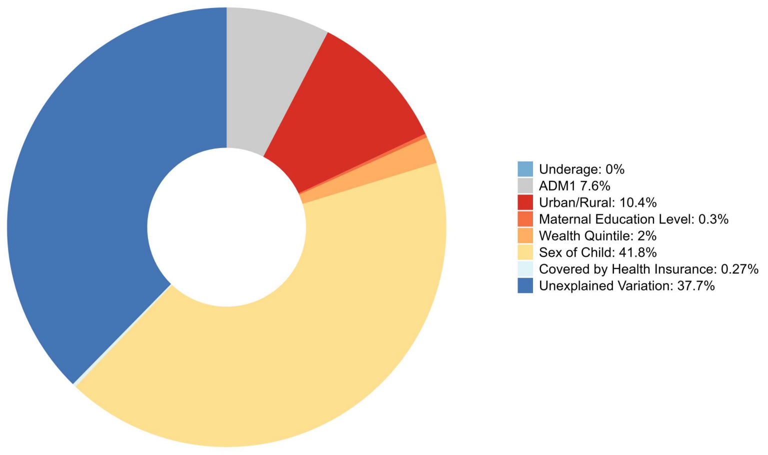

#### D: STROBE statement

This study was reported according to the Strengthening the Reporting of Observational Studies in Epidemiology (STROBE) statement for cross-sectional studies. The checklist below describes where each recommended reporting item is addressed in the manuscript.

**Supplementary Table 6. STROBE statement for cross-sectional studies**

| | Item No. | Recommendation | Location |
|---|---|---|---|
| **Title and abstract** | 1 | (*a*) Indicate the study's design with a commonly used term in the title or the abstract | Page 2 |
| | | (*b*) Provide in the abstract an informative and balanced summary of what was done and what was found | Page 2 |
| **Introduction** | | | |
| Background/rationale | 2 | Explain the scientific background and rationale for the investigation being reported | Page 3 |
| Objectives | 3 | State specific objectives, including any prespecified hypotheses | Page 4 |
| **Methods** | | | |
| Study design | 4 | Present key elements of study design early in the paper | Page 8 |
| Setting | 5 | Describe the setting, locations, and relevant dates, including periods of recruitment, exposure, follow-up, and data collection | Page 8; Supplementary Table 1 |
| Participants | 6 | (*a*) Give the eligibility criteria, and the sources and methods of selection of participants. | Pages 8-9; Supplementary Fig. 1 |
| | | (*b*) For matched studies, give matching criteria and number of exposed and unexposed | Not applicable |
| Variables | 7 | Clearly define all outcomes, exposures, predictors, potential confounders, and effect modifiers. Give diagnostic criteria, if applicable | Page 9-11 |
| Data sources and measurement | 8 | For each variable of interest, give sources of data and details of methods of assessment (measurement). Describe comparability of assessment methods if there is more than one group | Page 9-11 |
| Bias | 9 | Describe any efforts to address potential sources of bias | Page 8-9 |
| Study size | 10 | Explain how the study size was arrived at | Page 9; Supplementary Fig. 1 |
| Quantitative variables | 11 | Explain how quantitative variables were handled in the analyses. If applicable, describe which groupings were chosen and why | Pages 9-11 |

| | Item No. | Recommendation | Location |
|---|---|---|---|
| Statistical methods | 12 | (*a*) Describe all statistical methods, including those used to control for confounding | Page 9-12 |
| | | (*b*) Describe any methods used to examine subgroups and interactions | Page 10-11 |
| | | (*c*) Explain how missing data were addressed | Page 9-10 |
| **Results** | | | |
| Participants | 13 | (a) Report numbers of individuals at each stage of study—e.g numbers potentially eligible, examined for eligibility, confirmed eligible, included in the study, completing follow-up, and analysed | Page 9; Supplementary Fig. 1 |
| | | (b) Give reasons for non-participation at each stage | Page 9; Supplementary Fig. 1 |
| Descriptive data | 14 | (a) Give characteristics of study participants (eg demographic, clinical, social) and information on exposures and potential confounders | Page 2; Table 1 |
| | | (b) Indicate number of participants with missing data for each variable of interest | Page 9-10; Table 1 |
| Outcome data | 15 | Report numbers of outcome events or summary measures over time | Page 2-4 |
| Main results | 16 | (*a*) Give unadjusted estimates and, if applicable, confounder-adjusted estimates and their precision (e.g 95% confidence interval). Make clear which confounders were adjusted for and why they were included | Page 2-4; Tables 1 – 3 |
| | | (*b*) Report category boundaries when continuous variables were categorized | Not applicable |
| | | (*c*) If relevant, consider translating estimates of relative risk into absolute risk for a meaningful time period | Not applicable |
| Other analyses | 17 | Report other analyses done—e.g analyses of subgroups and interactions, and sensitivity analyses | Pages 3–4; Supplementary Figs. 2–5 |
| **Discussion** | | | |
| Key results | 18 | Summarise key results with reference to study objectives | Page 5 |
| Limitations | 19 | Discuss limitations of the study, taking into account sources of potential bias or imprecision. Discuss both direction and magnitude of any potential bias | Page 7-8 |
| Interpretation | 20 | Give a cautious overall interpretation of results considering objectives, limitations, multiplicity of analyses, results from similar studies, and other relevant evidence | Page 5-8 |

| | Item No. | Recommendation | Location |
|---|---|---|---|
| Generalisability | 21 | Discuss the generalisability (external validity) of the study results | Page 5-8 |
| **Other information** | | | |
| Funding | 22 | Give the source of funding and the role of the funders for the present study and, if applicable, for the original study on which the present article is based | Page 15 |
